# Identification of Key Genes and Signaling Pathways Associated with the Progression of Glioblastoma multiform

**DOI:** 10.1101/2020.12.21.20248616

**Authors:** Basavaraj Vastrad, Chanabasayya Vastrad, Iranna Kotturshetti

## Abstract

Genomic features have been gradually regarded as part of the basics to the clinical diagnosis, prognosis and treatment for glioblastoma multform (GBM). However, the molecular modifications taking place during the advancement of GBM remain unclear. Therefore, recognition of potential important genes and pathways in the gastric cancer progression is important to clinical practices. In the present study, gene expression dataset (GSE116520) of GBM were selected from the Gene Expression Omnibus (GEO) database and were further used to identify differentially expressed genes (DEGs). Then, pathway and Gene Ontology (GO) enrichment analyses were conducted, and a protein-protein interaction (PPI) network was constructed to explore the potential mechanism of GBM carcinogenesis. Significant modules were discovered using the PEWCC1 plugin for Cytoscape. In addition, a target gene - miRNA regulatory network and target gene - TF regulatory network in GBM were constructed using common deregulated miRNAs, TFs and DEGs. Finally, we carried on validation of hub genes by UALCAN, cBioporta, human protein atlas, ROC (Receiver operating characteristic) curve analysis, RT-PCR and immune infiltration analysis. The results indicated that a total of 947 differential expressed genes (DEGs) (477 up regulated and 470 down regulated) was identified in microarray profiles. Pathway enrichment analysis revealed that DEGs (up and down regulated) were mainly associated in reactive oxygen species degradation, ribosome, homocarnosine biosynthesis and GABAergic synapse, whereas GO enrichment analyses revealed that DEGs (up and down regulated) were mainly associated in macromolecule catabolic process, cytosolic part, synaptic signaling and synapse part as the main pathways associated in these processes. Finally, we filtered out hub genes, including MYC, ARRB1, RPL7A, SNAP25, SOD2, SVOP, ABCC3 and ABCA2, from the all networks. Validation of hub genes suggested the robustness of the above results. In conclusion, these results provided novel and reliable biomarkers for GBM, which will be useful for further clinical applications in GBM diagnosis, prognosis and targeted therapy.

## Introduction

Glioblastoma multform (GBM) is one of the most malignant glial tumors with the 5-year survival rate 9.8% [1]. In current years, although novel advances have been made in multimodal treatment of cancers, indigent prognosis and high mortality of GBM has remained consistent. About 296,851 individuals in the global were diagnosed with GBM in 2018, of which 241,037 people died, resulting in roughly equal morbidity and mortality [2]. Present situation, radiotherapy [3], chemotherapy [4] and surgical resection [5] are still the most effective way of improving the survival rate of GBM patients. However, GBM is difficult to diagnose in the early stages due to its concealed location and uncommon clinical symptoms. In most cases, majority of the patients tend to be in the final stage when they are clinically diagnosed and lose the chance of radiotherapy, chemotherapy and surgical resection. Therefore, the genes associated in the occurrence and advancement of GBM needs to be explored, which will contribute to the finding of diagnostics markers, prognostic markers and therapeutic targets of GBM.

The underlying molecular pathogenesis of GBM remains inadequately unexplored. Therefore, it is encourage the need to advance a further diagnose the etiological factors, molecular mechanisms, and pathways of GBM to discover novel diagnostic and treatment strategies for GBM. Fortunately, with the development of highthroughput DNA microarray analyses, various genes and pathways have been demonstrated to be correlated with the genesis and progression of GBM [6]. Genes such as NDRG2 [7], PARK2 [8], WT1 [9], RB1 [10] and HDAC (histone deacetylase) [11] were linked with pathogenesis of GBM. Pathways such as Akt pathway [12], EGFR–MEK–ERK signaling pathway [13], AMPK-TSC-mTOR signaling pathway [14], NFκB pathway [15] and MAP kinase pathway [16] were involved in progression of GBM. Therefore, finding differentially expressed genes (DEGs) and pathways, illuminate the interactions network among them, are important for GBM.

In this study, we downloaded the original data (GSE116520) from Gene Expression Omnibus (GEO, http://www.ncbi.nlm.nih.gov/geo/). The differentially expressed genes (DEGs) of normal control from GBM were screened using limma R bioconductor tool. Subsequently, the pathway and gene ontology (GO) enrichment analysis for DEGs were analyzed. Additionally, we established protein-protein interaction (PPI) network, target gene - miRNA regulatory network and target gene - TF regulatory network of the DEGs. Expression levels of these candidate genes were finally verified by survival analysis, expression analysis (based on sample type and patients age), mutation analysis and immune histochemical (IHC) analysis, ROC (Receiver operating characteristic) curve analysis, RT-PCR and immune infiltration analysis. Overall, our systematic analysis will gain insights into GBM pathogenesis at molecular level and help to identify the potential candidate biomarkers for diagnosis, prognosis, and drug targets for GBM.

## Materials and methods

### Selection of GEO data set

Firstly, GBM-related chips GSE116520 [17] were retrieved and downloaded from the Gene Expression Omnibus (GEO) database (https://www.ncbi.nlm.nih.gov/geo/) with “Glioblastoma multform” serving as the retrieval key word. GSE116520 included eight normal control samples (brain) and seventeen GBM samples. The microarray platform was GPL10558 Illumina HumanHT-12 V4.0 expression beadchip Array. Flow chart of complete studies is shown in Fig. 1.

**Fig. 1.**
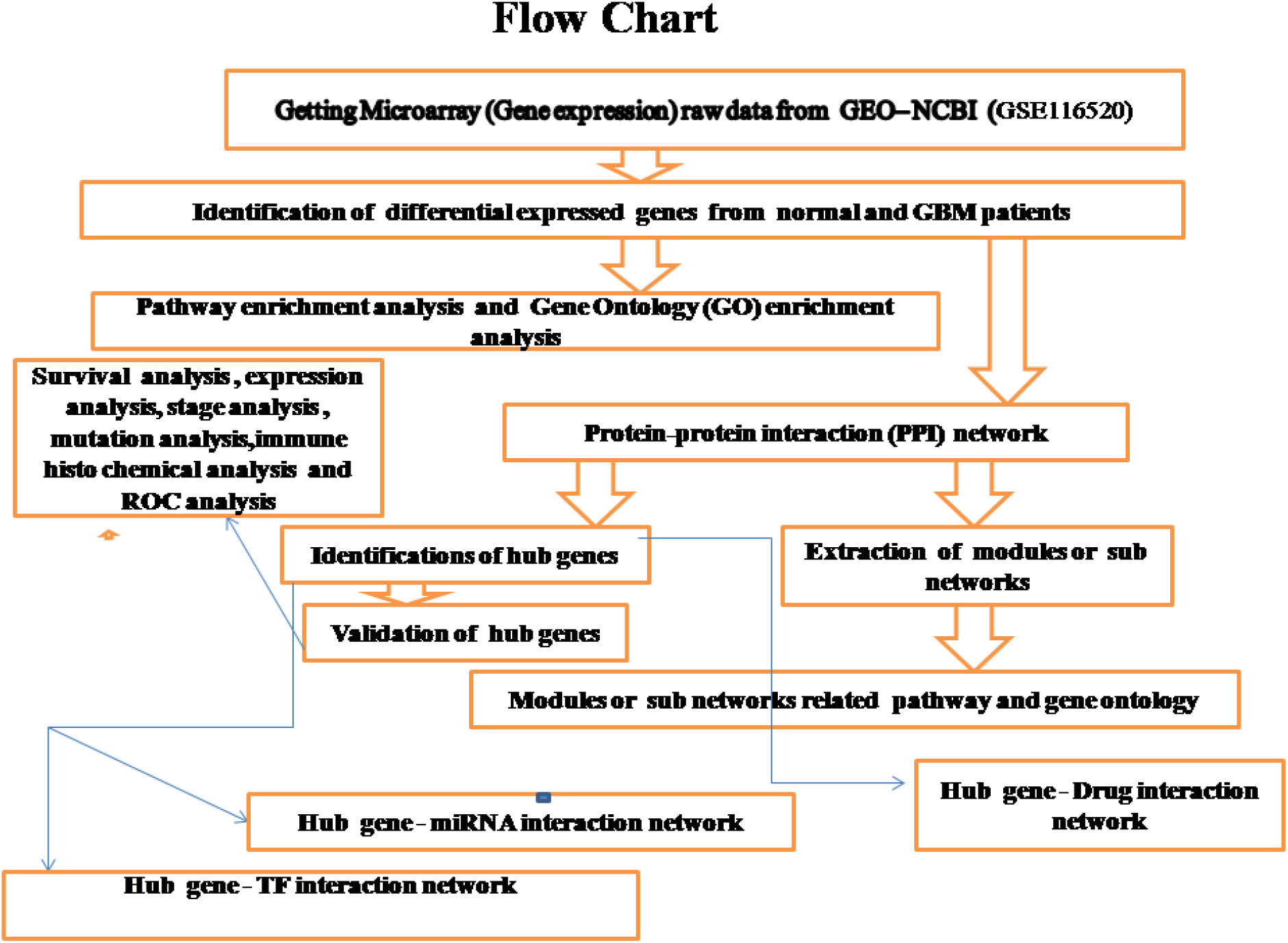
Study design (flow diagram of study)

### Data pre-processing

The downloaded probe-level raw data in TXT files were preprocessed using beadarray package [18] in R (version 3.3.2), including log-transformation, imputation of missing values, background correction, and quantile normalization. While several probes mapped to one gene, equate value of this probes was determined and used as the final expression value.

### Differential expression analysis

The DEGs between GBM tissues and normal control tissues in each individual experiment were identified using Bayes moderated t-test method based on limma package [19], with the threshold criteria of FDR (false discovery rate) < 0.05, |log2FC (fold change)| > 1.88 for up regulated genes and |log2FC (fold change)| <- 2.25 for down regulated genes. The relationships between samples and DEGs were shown by hierarchical clustering heatmaps and volcano plots.

### Pathway enrichment analysis of DEGs

The pathway enrichment analyses were performed by ToppGene (ToppFun) (https://toppgene.cchmc.org/enrichment.jsp) [20]. BIOCYC (https://biocyc.org/) [21], Kyoto Encyclopedia of Genes and Genomes (KEGG; http://www.genome.jp/kegg/) [22], Pathway Interaction Database (PID, http://pid.nci.nih.gov/) [23], Reactome (https://reactome.org/PathwayBrowser/) [24], Molecular signatures database (MSigDB, http://software.broadinstitute.org/gsea/msigdb/) [25], GenMAPP (http://www.genmapp.org/) [26], Pathway Ontology (https://bioportal.bioontology.org/ontologies/PW) [27], PantherDB (http://www.pantherdb.org/) [28] and Small Molecule Pathway Database (SMPDB) (http://smpdb.ca/) [29] pathway enrichment analysis were carried out for the DEGs, with a P < 0.05 considered to indicate statistical significance.

### Gene ontology enrichment analysis of DEGs

To explore the biological functional roles of the above DEGs, a GO (http://www.geneontology.org/) [30] enrichment analysis was performed on ToppGene (ToppFun) (https://toppgene.cchmc.org/enrichment.jsp) [20]. Significant results of biological process (BP), cellular component (CC) and molecular function (MF) with a cut-off of false discovery rate <0.05 were selected.

### PPI network construction and module analysis

To further investigate the molecular mechanism of GBM, all DEGs were used to construct the PPI network using the biological online database tool (Integrated Interactions Database, IID, http://iid.ophid.utoronto.ca/) [31] to determine and predict the interaction among them. This database integrates various PPI data bases such as Biological General Repository for Interaction Datasets (BioGRID, https://thebiogrid.org/) [32], IntAct (https://www.ebi.ac.uk/intact/) [33], I2D (http://ophid.utoronto.ca/ophidv2.204/) [34], Molecular INTeraction database (MINT, https://mint.bio.uniroma2.it/) [35], InnateDB [https://www.innatedb.com/] [36], Database of Interacting Proteins (DIP, https://dip.doe-mbi.ucla.edu/dip/Main.cgi) [37], Human Protein Reference Database (HPRD, http://www.hprd.org/) [38] and the Biomolecular Interaction Network Database (BIND, http://bind.ca) [39]. A combined score > 0.7 (high confidence score) was considered significant, and then the PPI network was visualized using Cytoscape software (http://www.cytoscape.org/) (Version 3.7.2) [40]. To evaluate the importance of nodes in the PPI network, the degree centrality, betweenness centrality, stress centrality, closeness centrality and clustering coefficient of nodes were calculated and utilized in the present study [41–45] using the network analyzer plugin in Cytoscape software. The hub genes, a small number of important nodes for the protein interactions in the PPI network, were chosen with a degree centrality > 50, betweenness centrality > 0.02, stress centrality > 2100000, closeness centrality > 0.26 and clustering coefficient = 0. Because a higher k-core score means a more topological central location, modules in the PPI network were explored by k-core scoring using the PEWCC1 plugin in Cytoscape software [46], and significant modules with a k-core > 6 were considered potential core regulatory networks.

### Construction of target gene - miRNA regulatory network

To identify regulatory miRNAs that influence target gene (i.e., up and down regulated genes) at the posttranscriptional level, target gene - miRNA interactions were obtained from DIANA-TarBase (http://diana.imis.athena-innovation.gr/DianaTools/index.php?r=tarbase/index) [47] and miRTarBase (http://mirtarbase.mbc.nctu.edu.tw/php/download.php) [48] both of which include experimentally supported target gene - miRNA interactions and topological parameter (degree) were analyzed using NetworkAnalyst (https://www.networkanalyst.ca/) [49].

### Construction of target gene - TF regulatory network

To identify regulatory TFs that control the i.e., up and down regulated genes) at a transcriptional level, TF-target gene interactions were obtained using the ChEA database (http://amp.pharm.mssm.edu/lib/chea.jsp) [50] and were identified topological parameter (degree centrality) using (https://www.networkanalyst.ca/) [49].

### Validation of hub genes and clinical significance

The UALCAN (https://ualcan.path.uab.edu/index.html) [51] online database was used for survival analysis, expression analysis and age related expression analysis of the hub genes, which analyzed RNA sequencing expression data from TCGA projects. The mutation frequency of hub genes was inquired in cBioportal online database (http://www.cbioportal.org/) [52]. The hub gene expressions in GBM tissues were determined from the human protein atlas (www.proteinatlas.org) [53]. To explore diagnostic biomarkers of GBM, we used the above hub genes as candidates to find their diagnostic value based on generalized linear models (GLM). The pROC package [54] in R was used for GLM analysis. In brief, half of the samples (GBM = 17, controls = 8) were randomly distributed as the training set, which was used to build a model. An ROC (Receiver operating characteristic) curve analysis was practiced to calculate the specificity and sensitivity of the GLM prediction model. The AUC was computed to evaluate the diagnostic efficiency of the classifier. All cell culture samples of normal (HCN-1A) and GBM (U-118 MG) were lysed using TRIzol® (Invitrogen; Thermo Fisher Scientific, Inc.), and total RNAs were extracted and reverse transcribed into cDNA templates using PrimeScript® RT Reagent kit (Takara Biotechnology Co., Ltd.) according to the manufacturer’s instructions. PCR was performed using an 7900HT real-time PCR instrument with an initial denaturation at 95 °C for 30 s, followed by 40 cycles at 95 °C for 15 s and 60 °C for34 s, and a fnal dissociation curve analysis of one cycle at 95 °C for 15 s, 60 °C for 1 min, and 95 °C for 15 s. Each cDNA sample was assayed three times and relative expression was resolved using the 2^−ΔΔCT^ method [55]. The specific PCR primers for the hub genes and β-actin as the internal control gene were designed with Primer Express version 2.0. TIMER (https://cistrome.shinyapps.io/timer/) [56] is a user friendly, interactive web resource for immune infiltration analysis from RNA-Seq expression profiling database (The Cancer Genome Atlas (TCGA)). Immune infiltration analysis was evaluated using immune infiltrates (B cells, CD4+ T cells, CD8+ T cells, neutrophils, macrophages, and dendritic cells) across GBM.

## Results

### Data preprocessing and screening of DEGs

The gene expression profile GSE116520 was downloaded from the GEO. The data before and after normalization are shown in Fig. 2A and Fig. 2B. The limma method was used to identify DEGs in GBM tissue compared with normal control tissues (brain). P value < 0.05, log FC > 1.88 for up regulated genes, and log FC <- 2.25 for down regulated genes were used as the cut-off criteria. After analyzing, total of 947 DEGs were selected between the GBM tissues and normal control tissues, including 477 up genes and 470 down regulated genes (Table 1). The result is displayed in the volcano plot (Fig. 3). The heatmap of the DEGs (up and down regulated genes) are shown in Fig. 4 and Fig. 5.

**Fig. 2.**
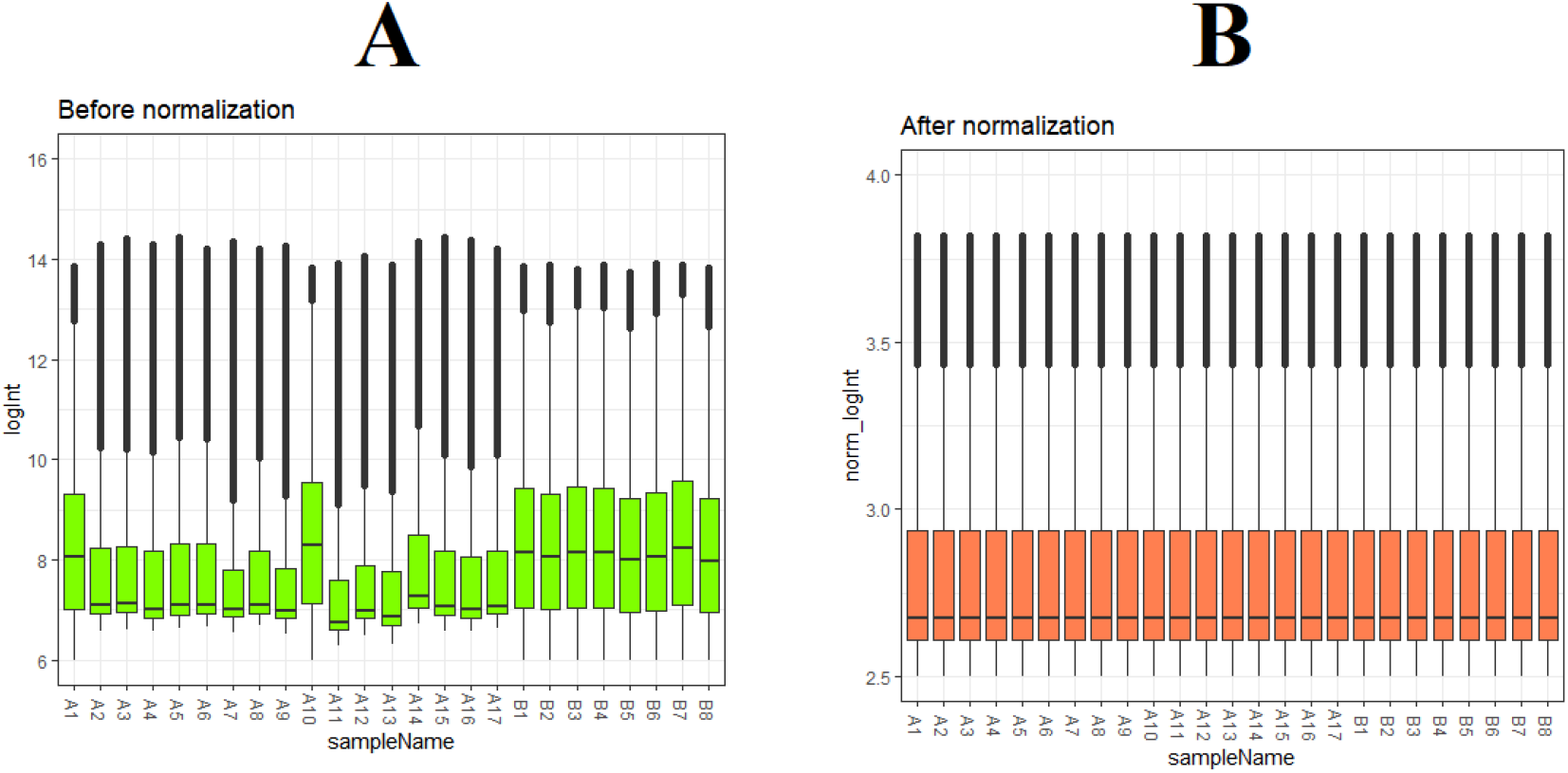
Box plots of the gene expression data before normalization (A) and after normalization (B). Horizontal axis represents the sample symbol and the vertical axis represents the gene expression values. The black line in the box plot represents the median value of gene expression. (A1 – A17 = GBM tissues samples; B1 – B8 = normal control samples)

**Fig. 3.**
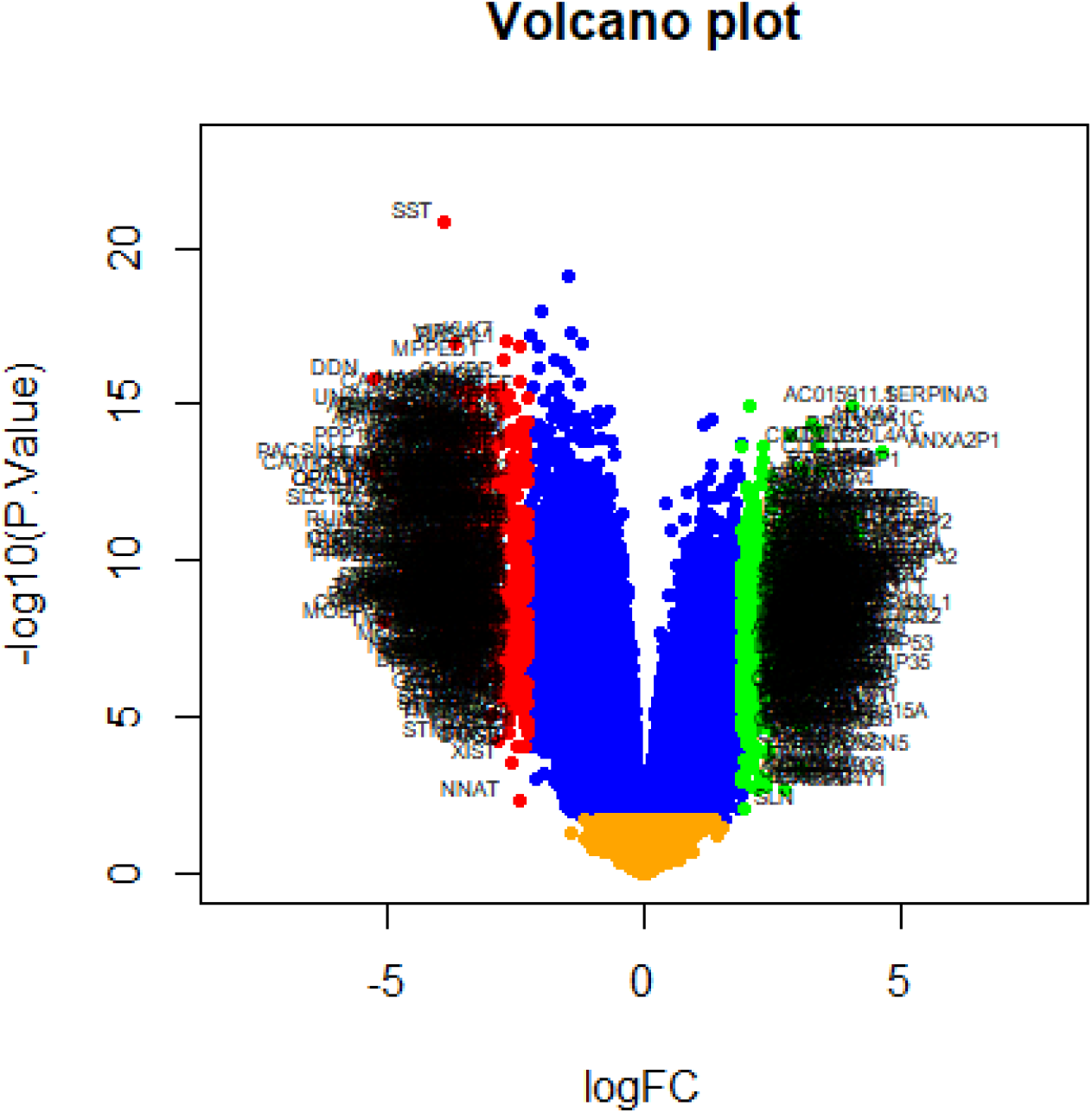
Volcano plot of differentially expressed genes. Genes with a significant change of more than two-fold were selected.

**Fig. 4.**
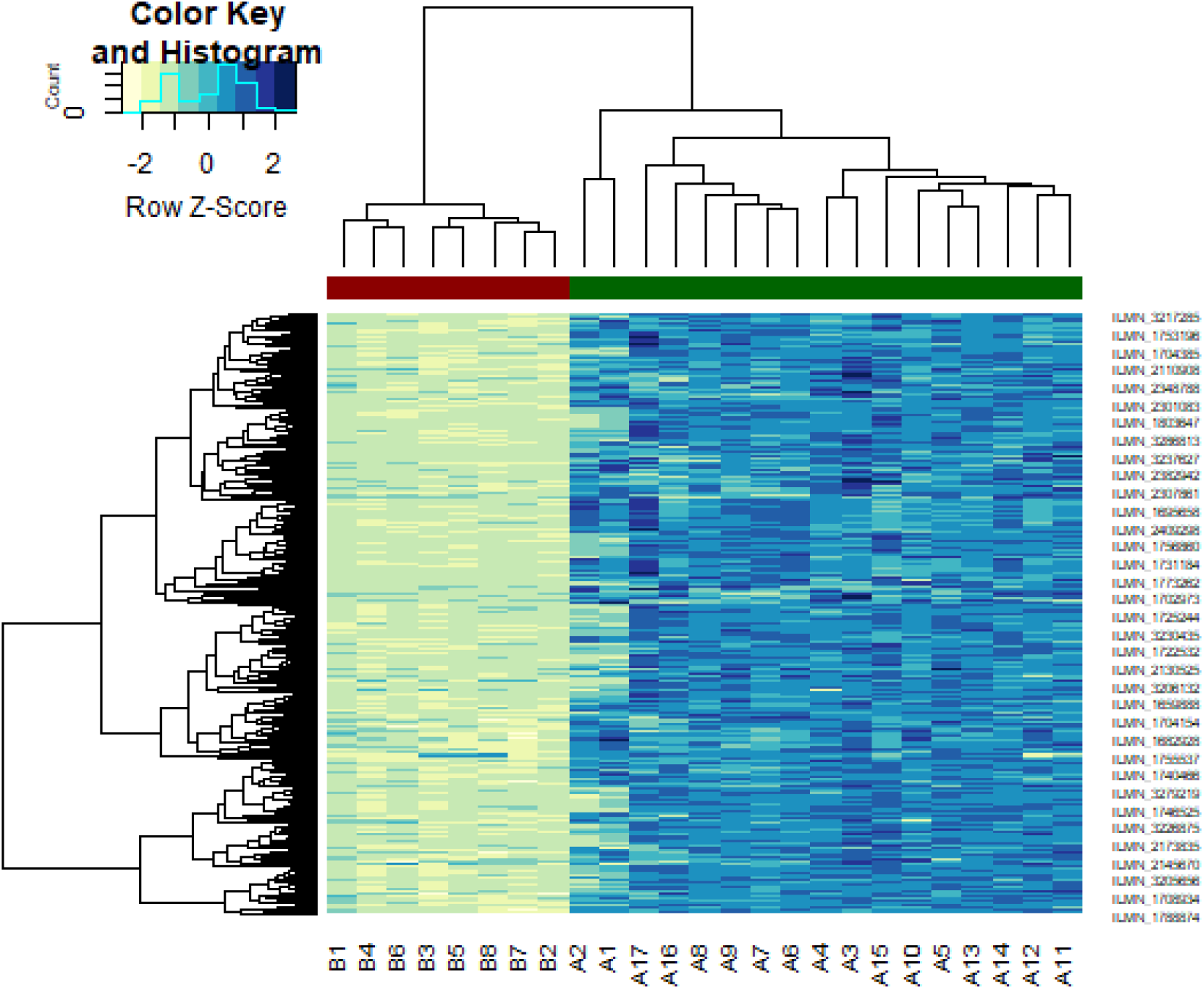
Heat map of up regulated differentially expressed genes. Legend on the top left indicate log fold change of genes. (A1 – A17 = GBM tissues samples; B1 – B8 = normal control samples)

**Fig. 5.**
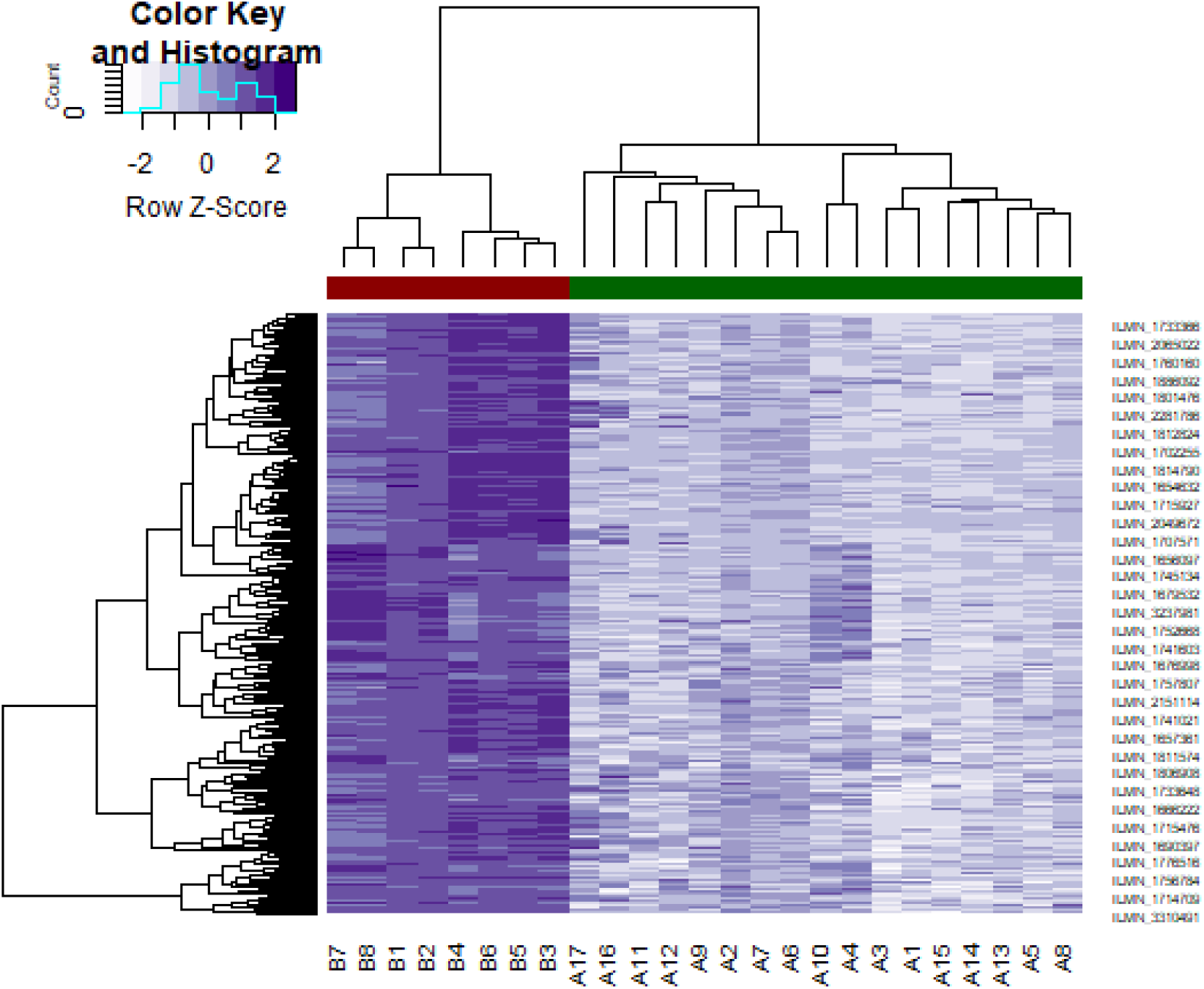
Heat map of down regulated differentially expressed genes. Legend on the top left indicate log fold change of genes. (A1 – A17 = GBM tissues samples; B1 – B8 = normal control samples)

**Table 1.**
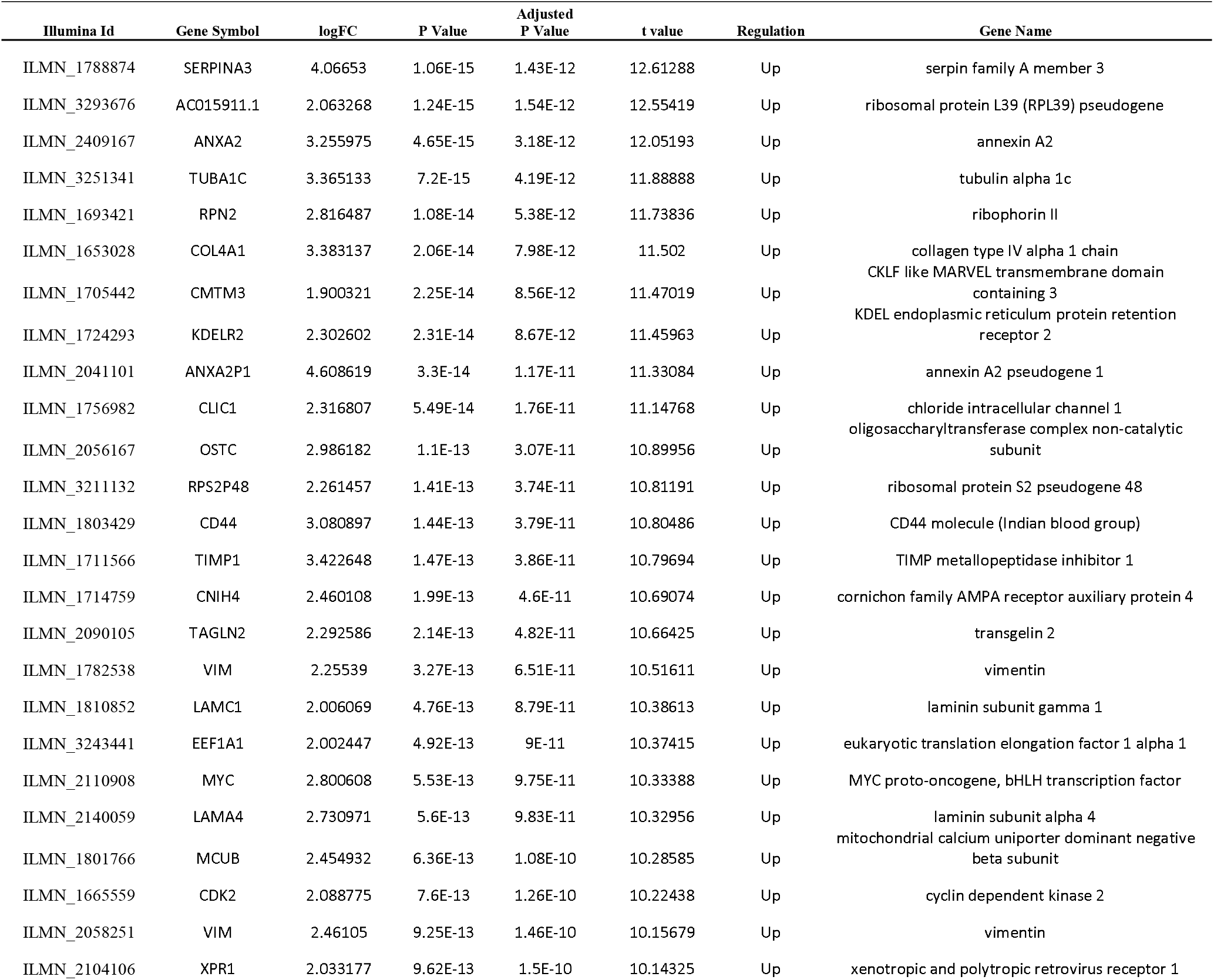
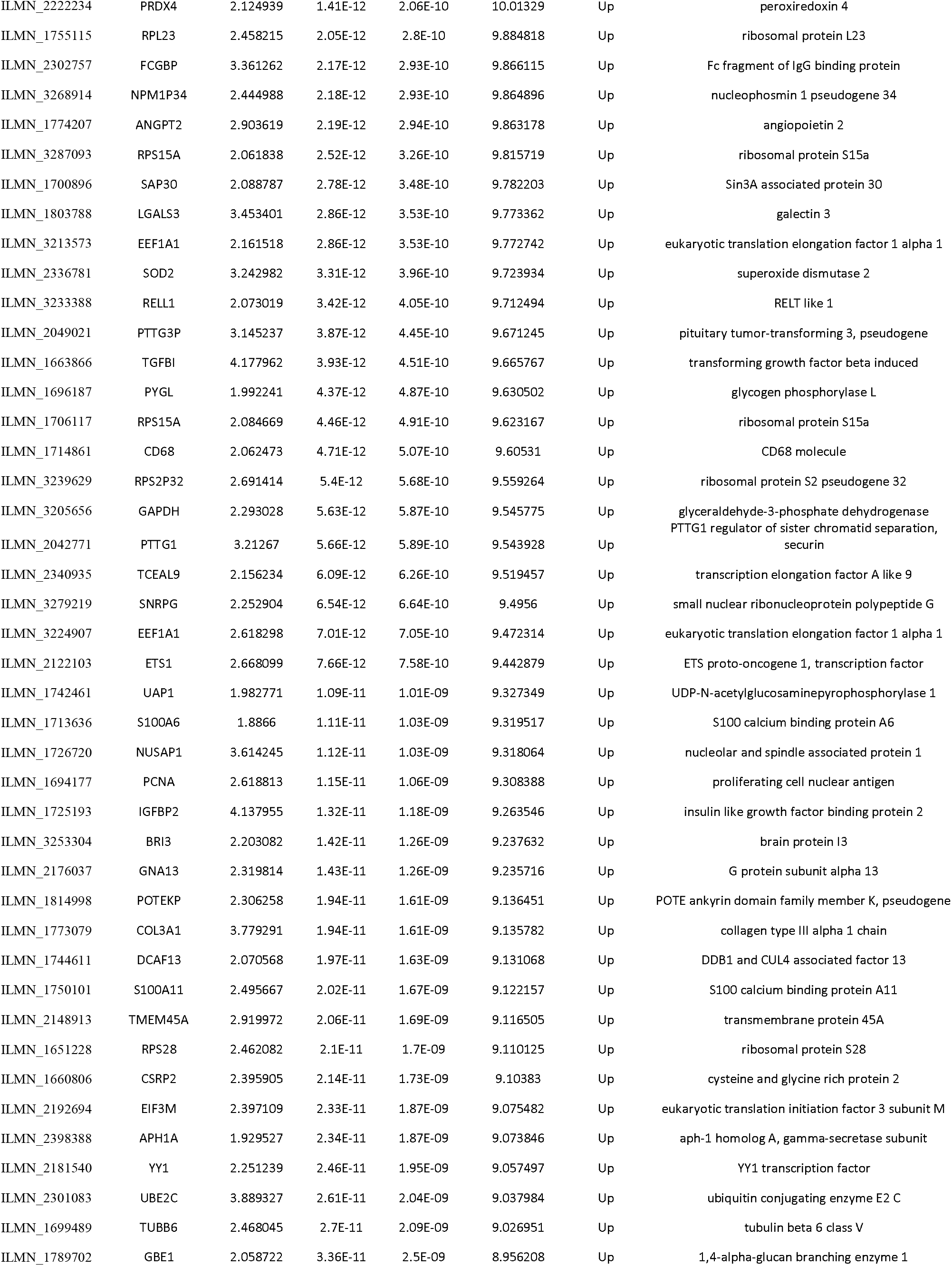
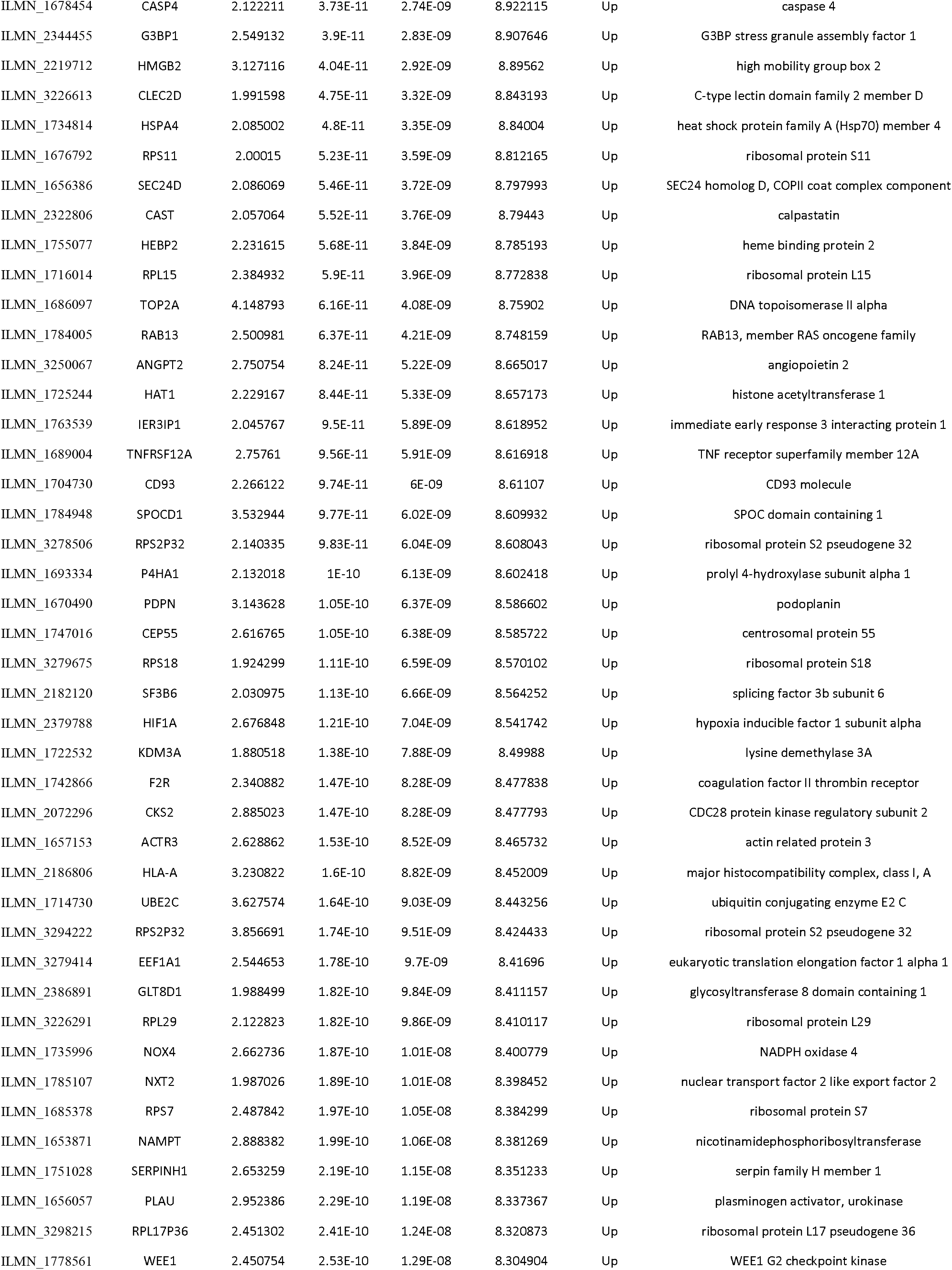
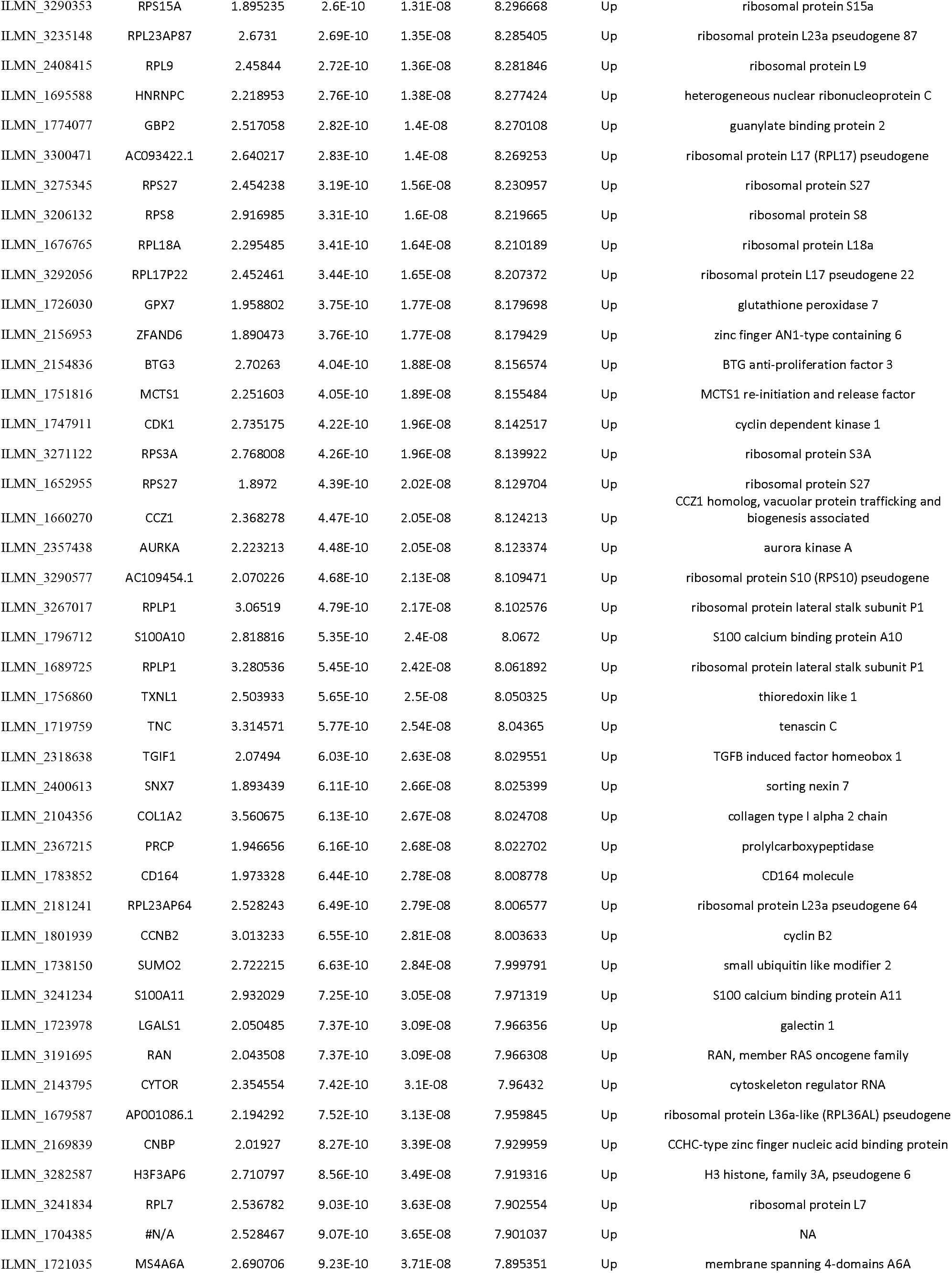
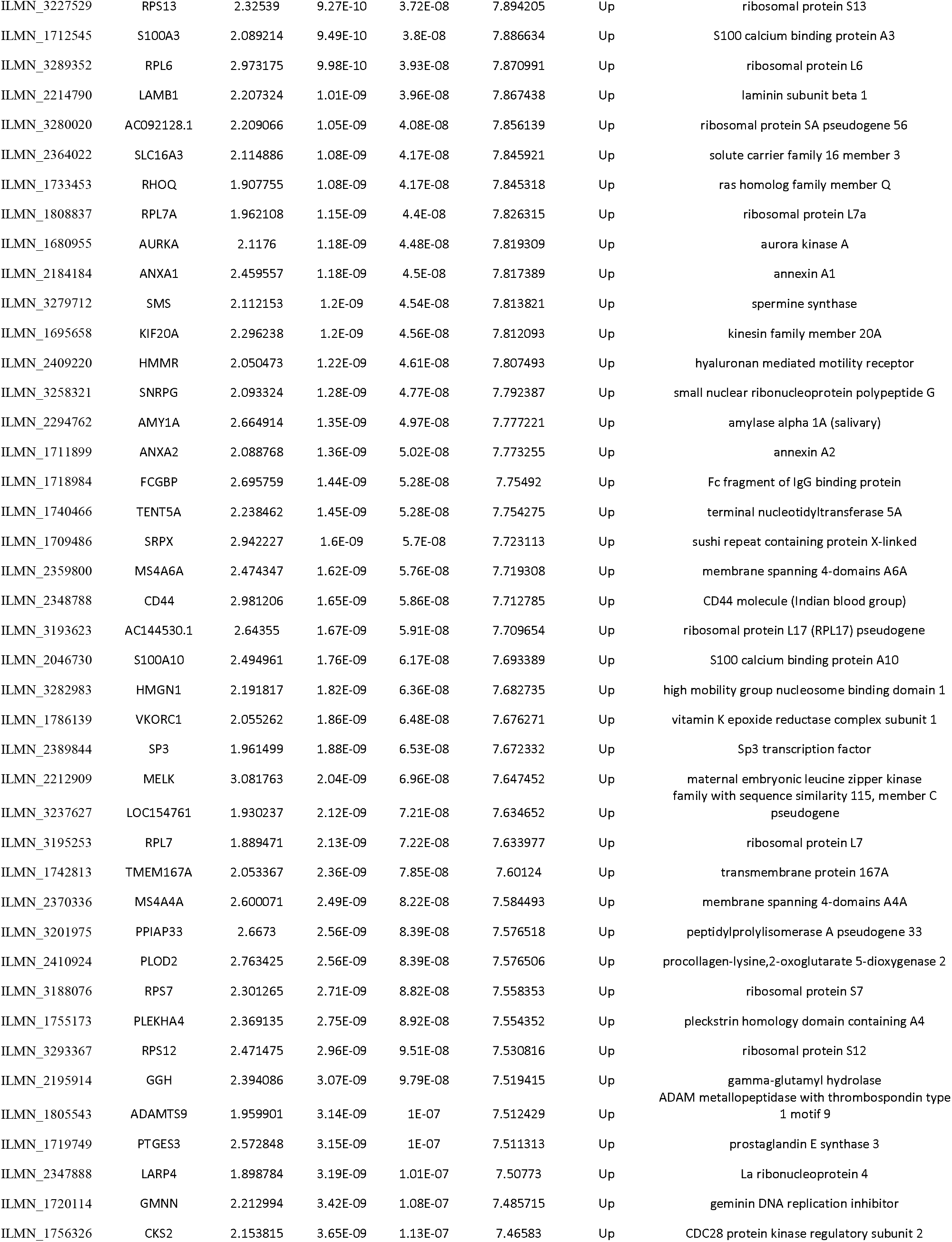
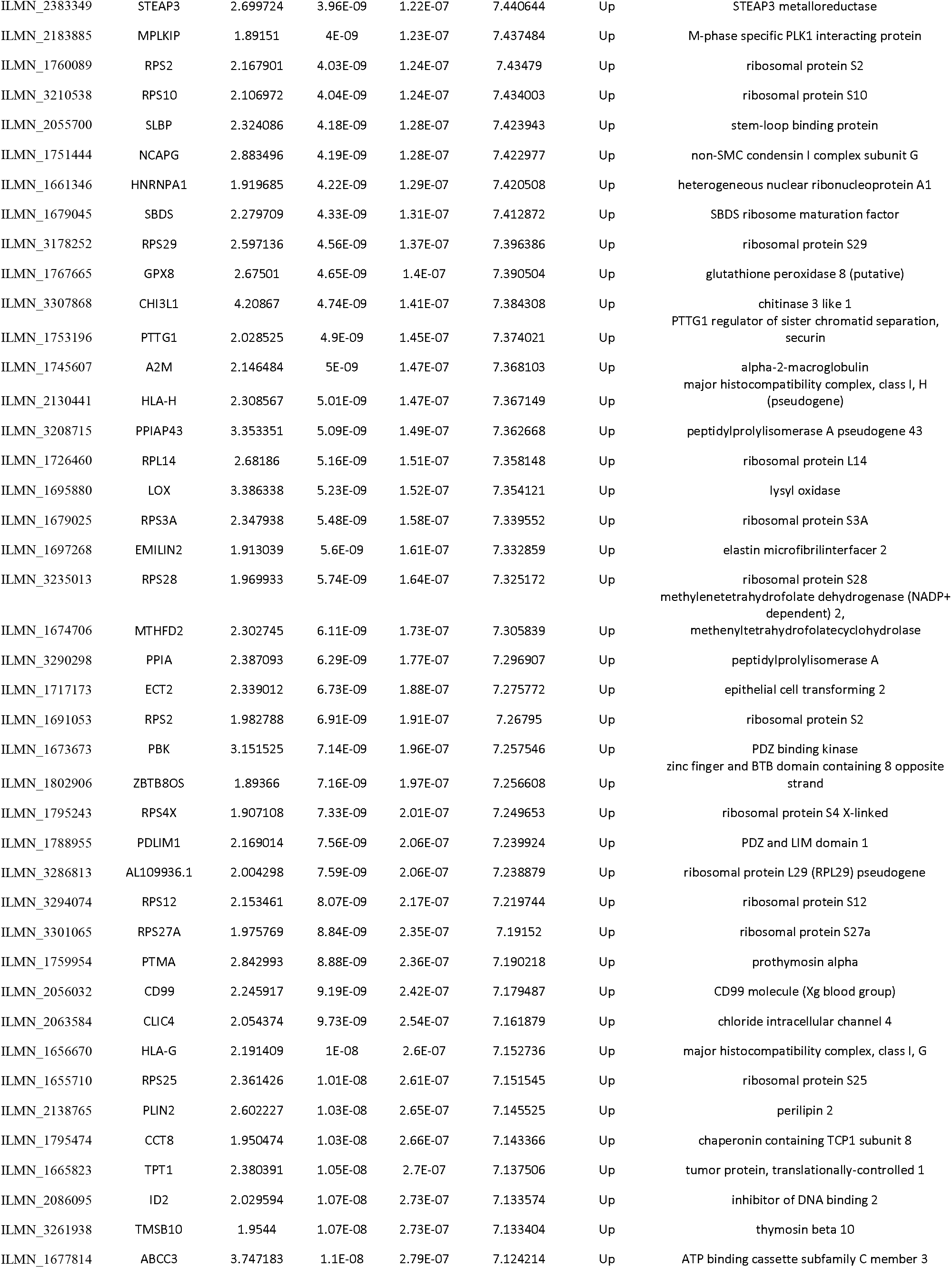
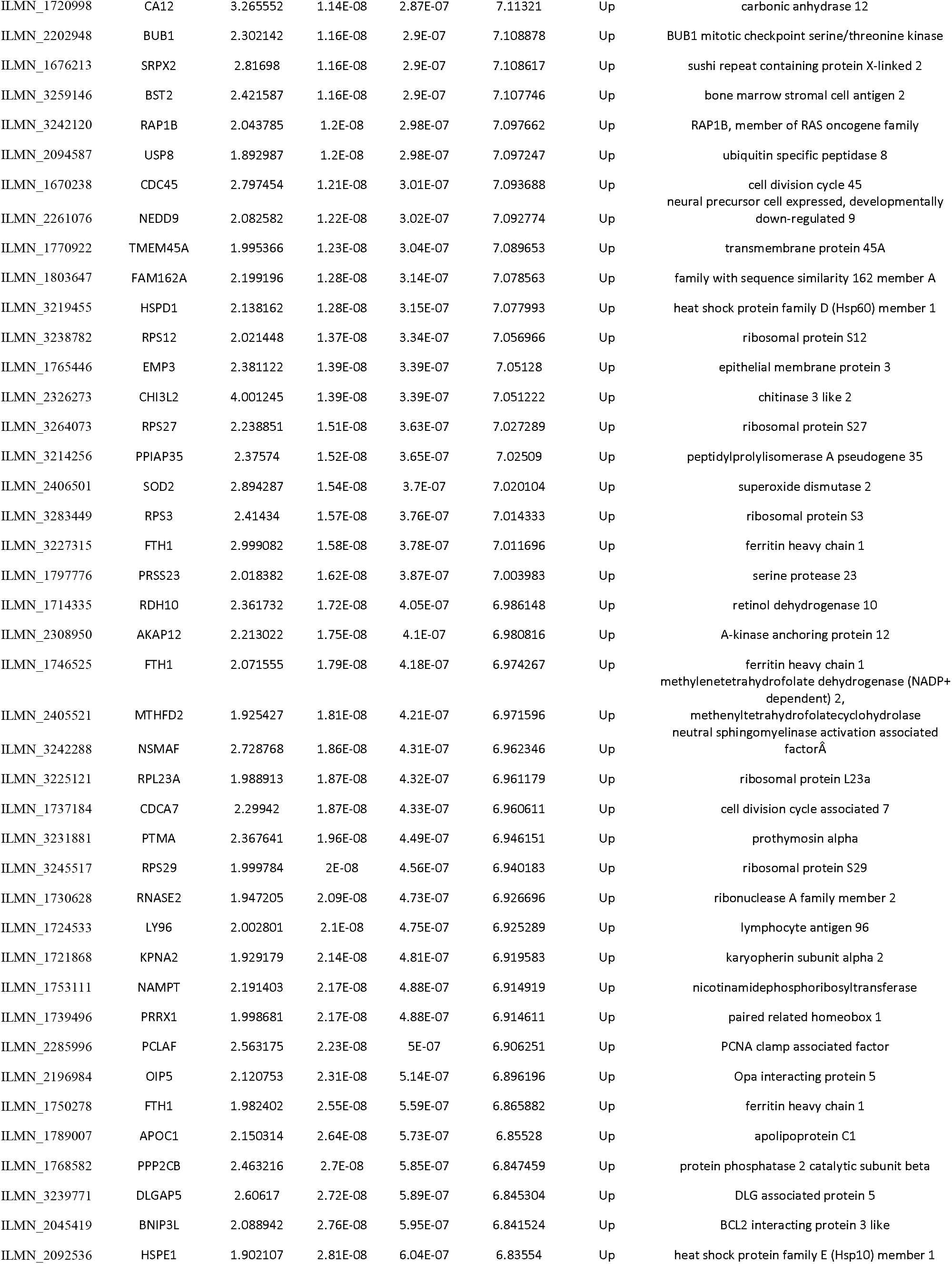
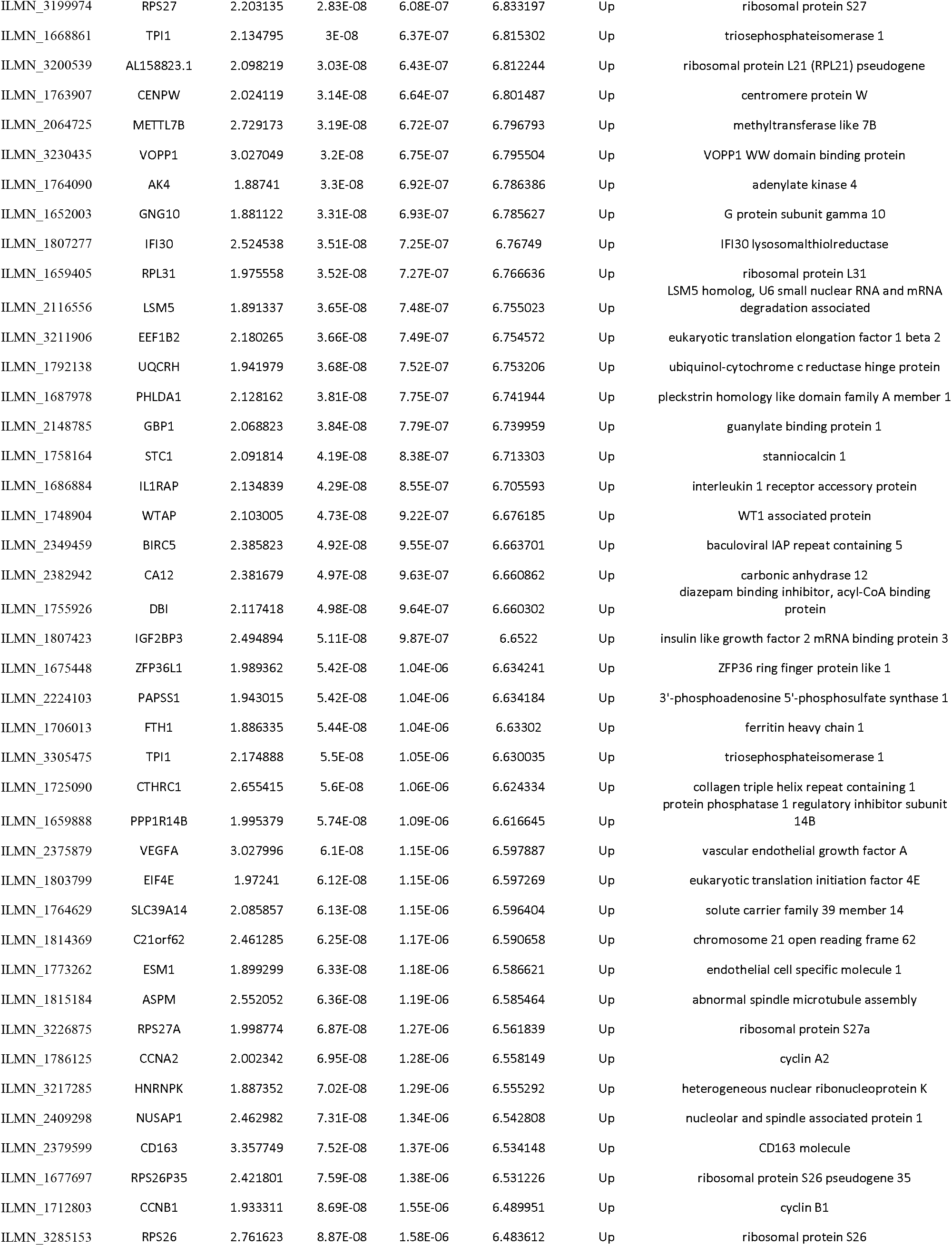
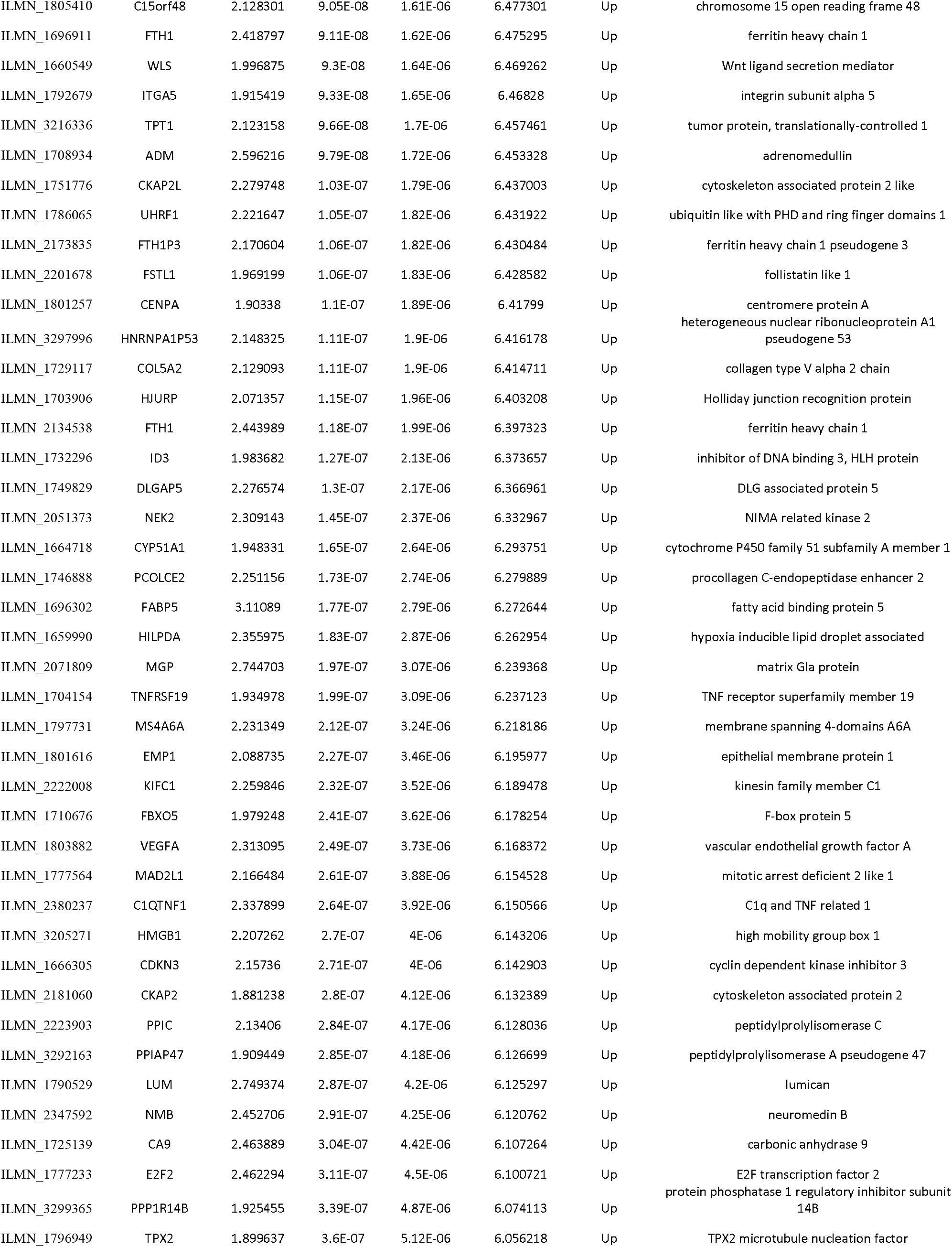
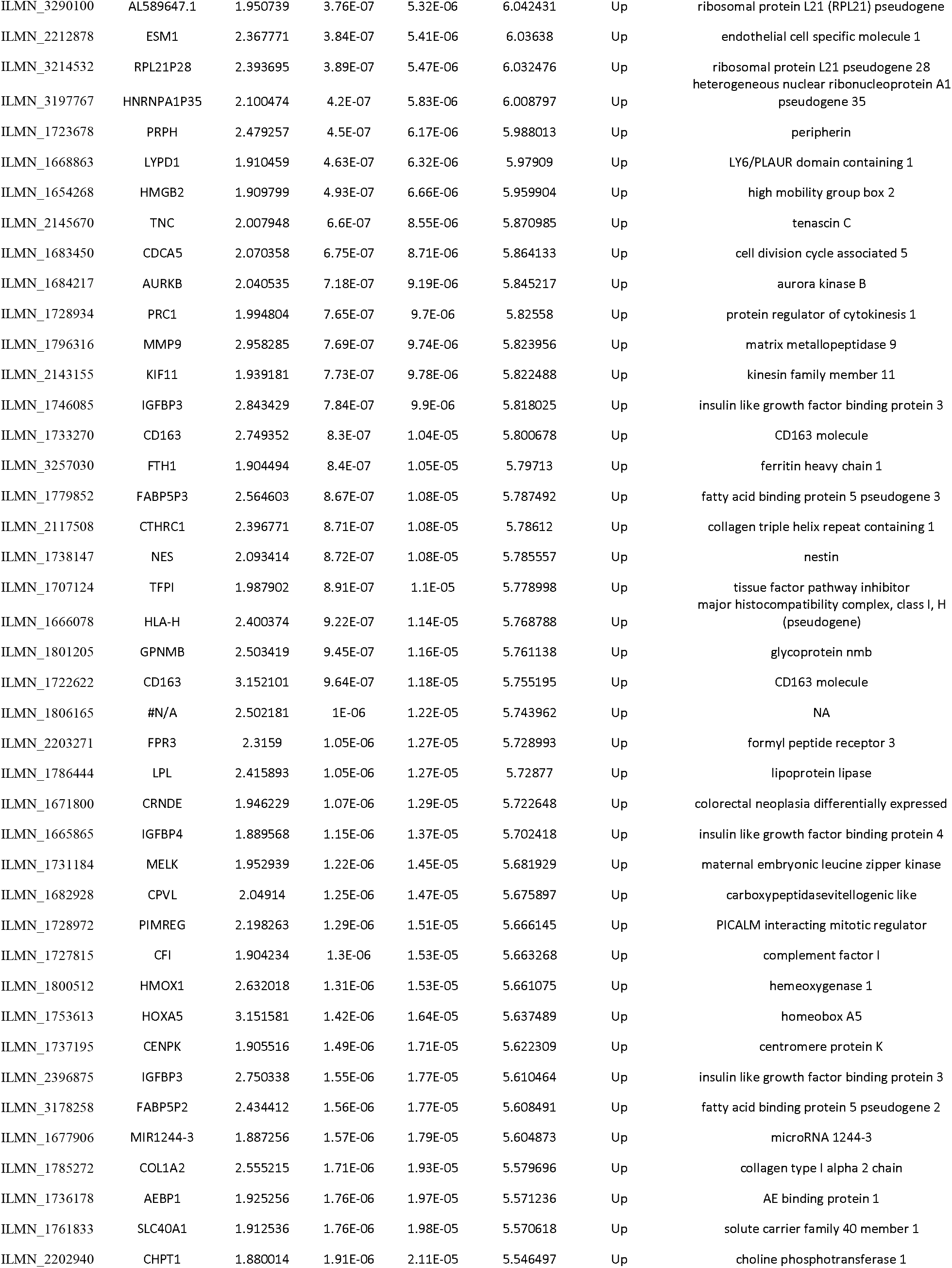
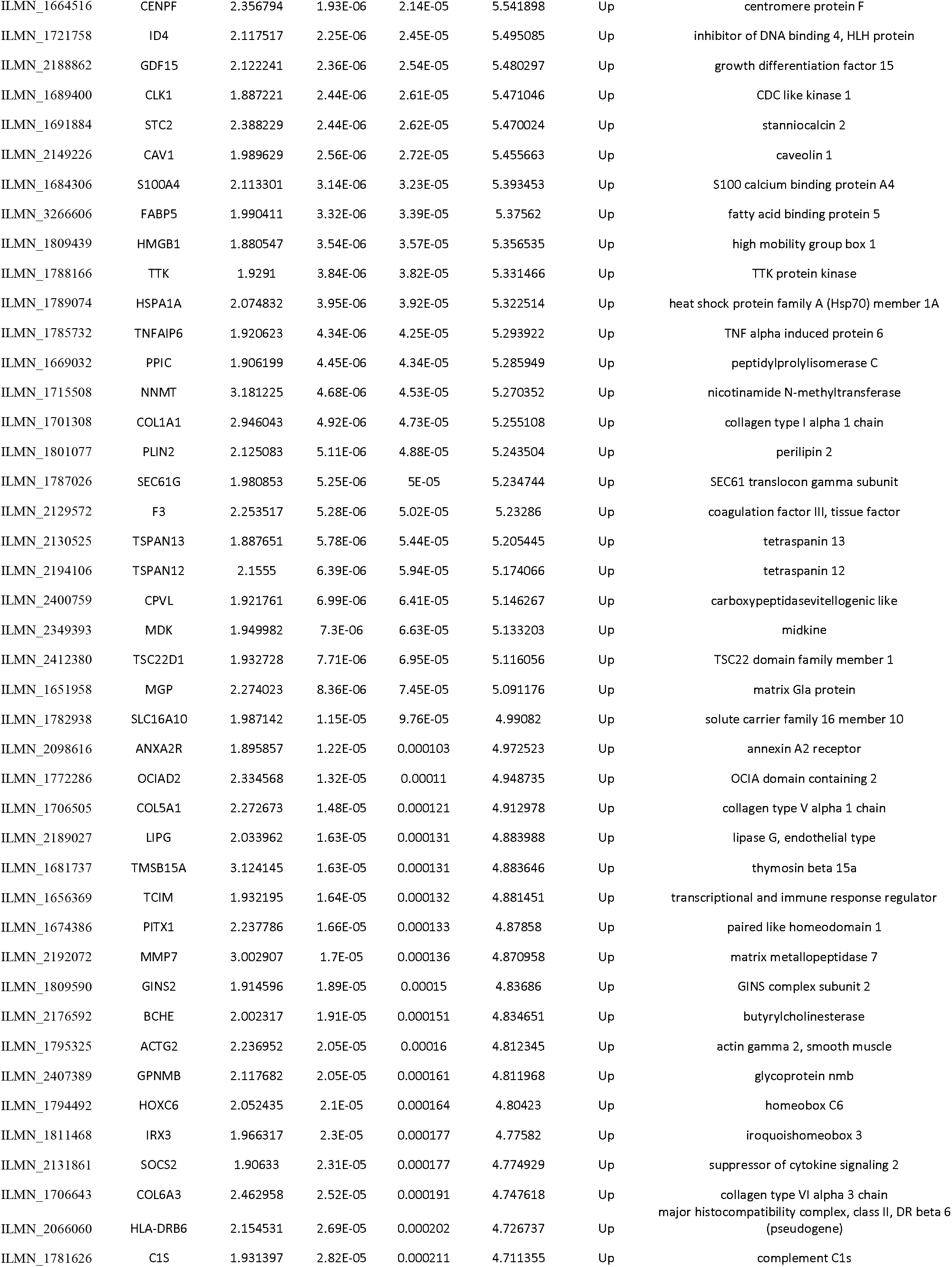
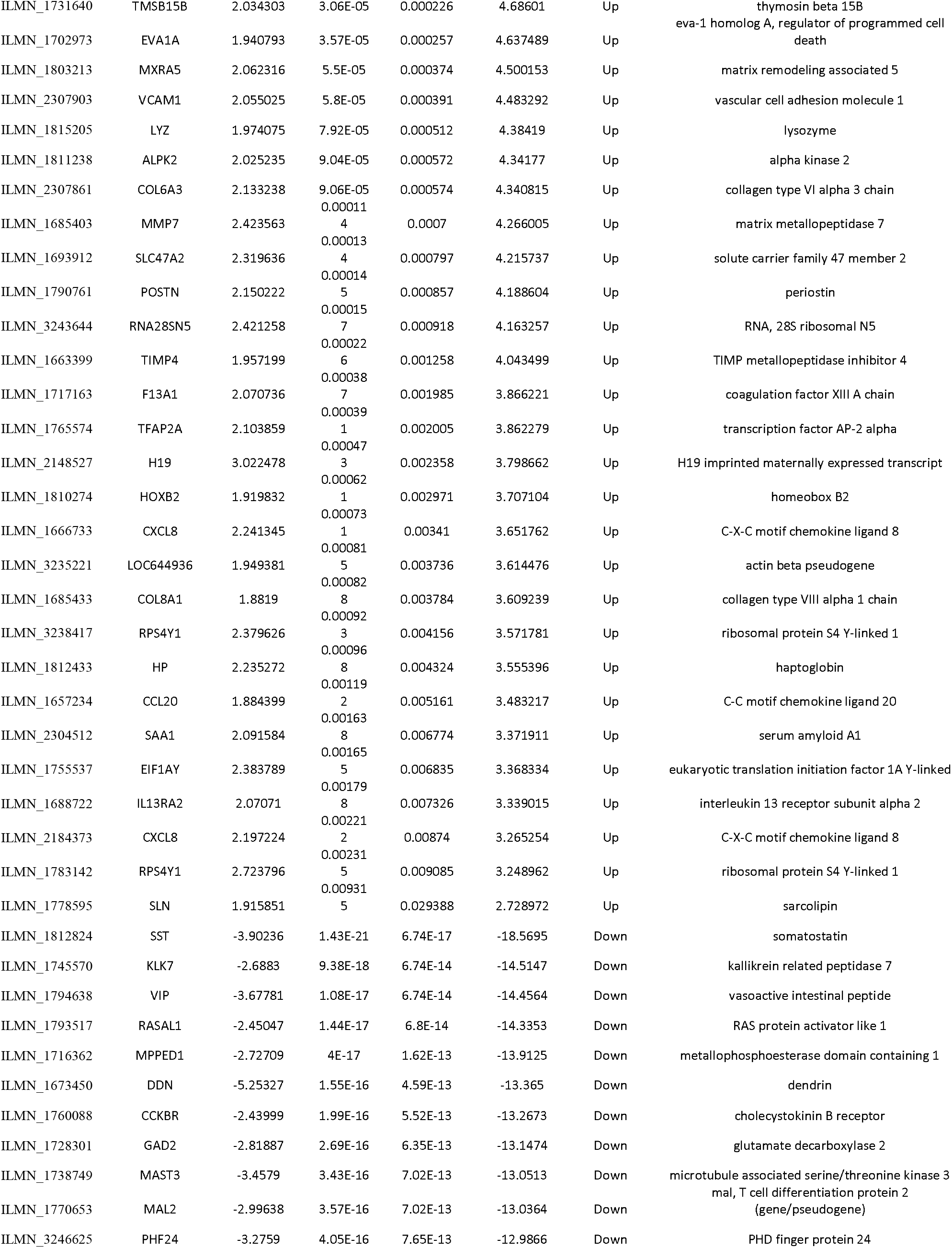
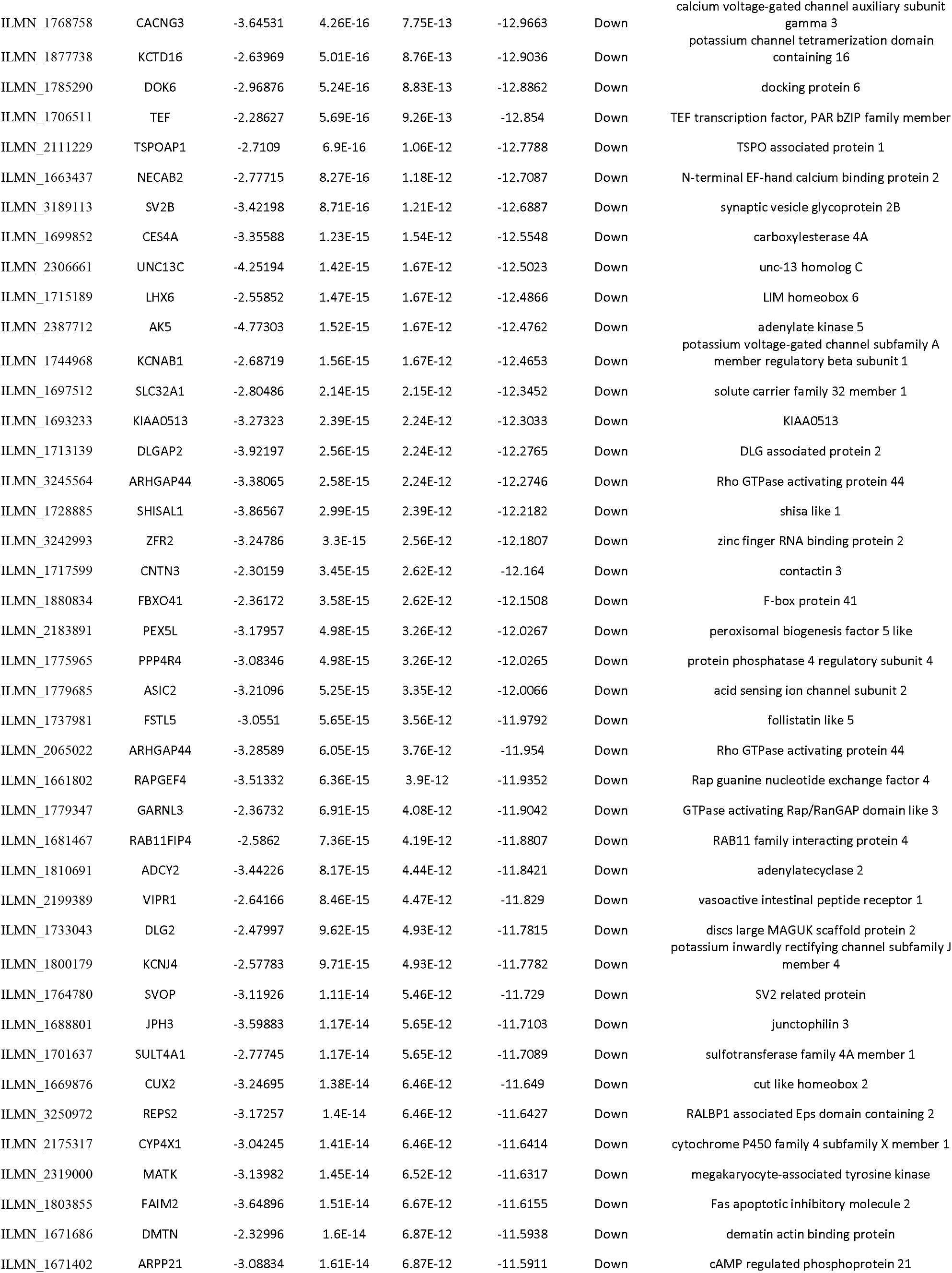
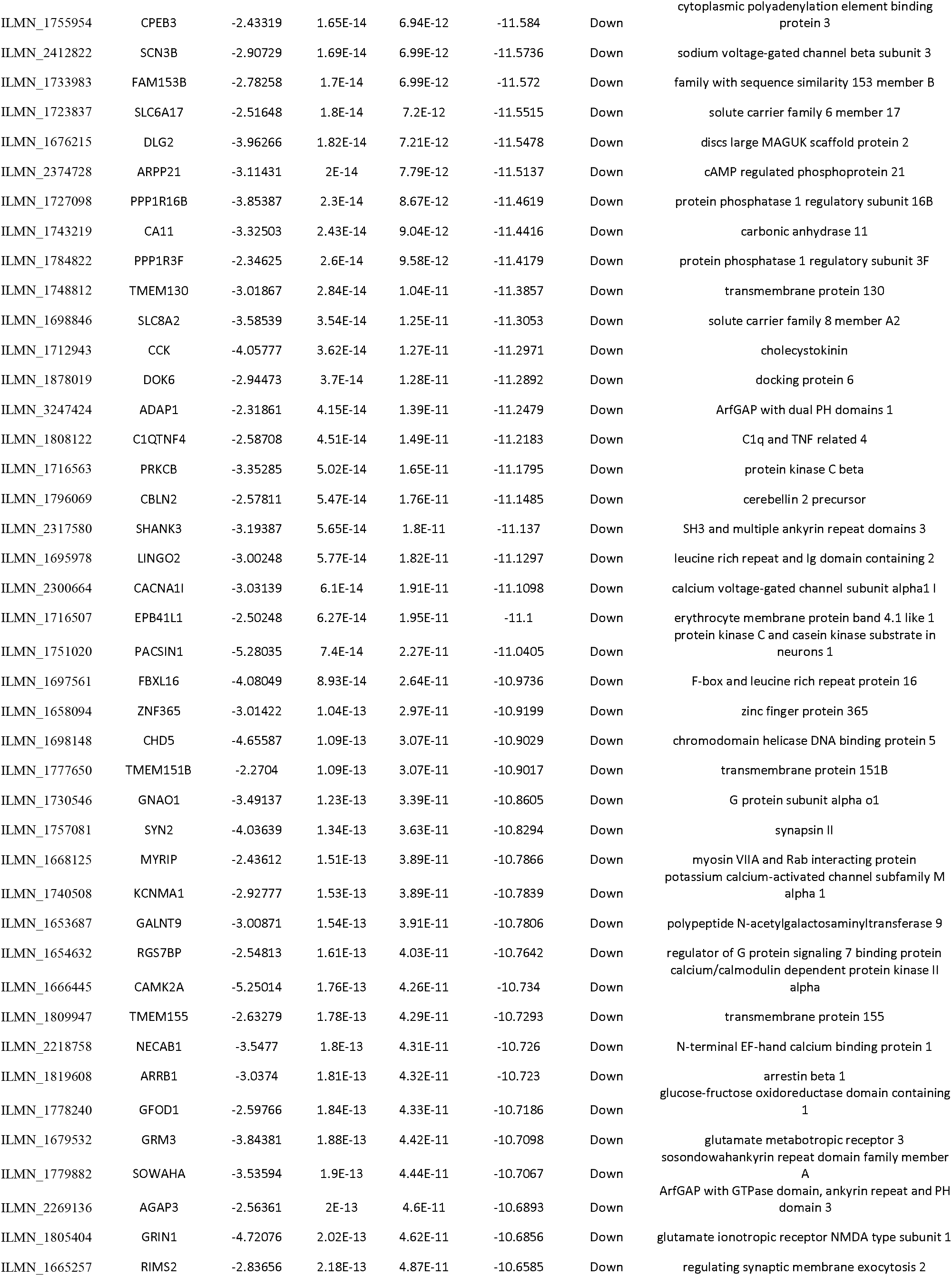
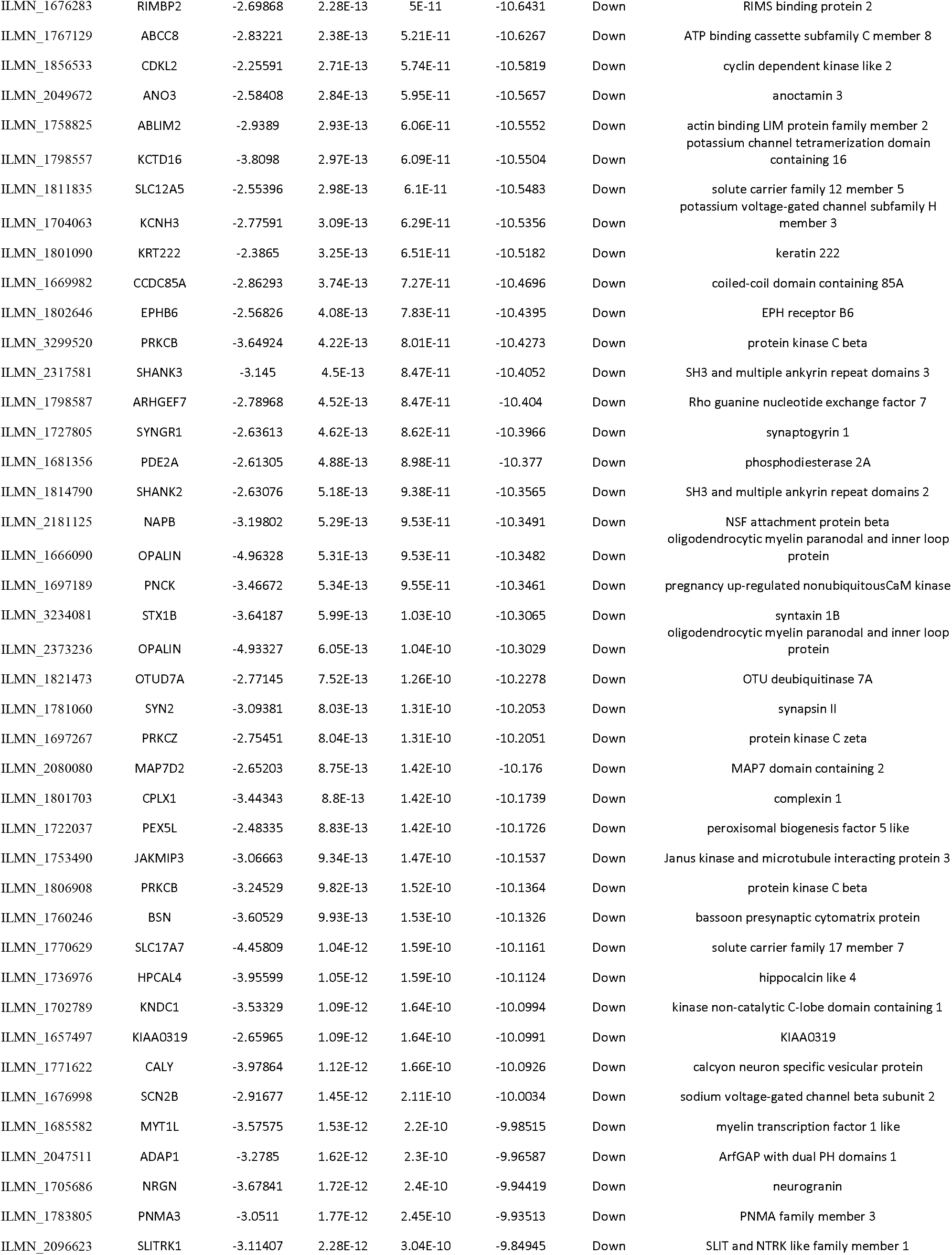
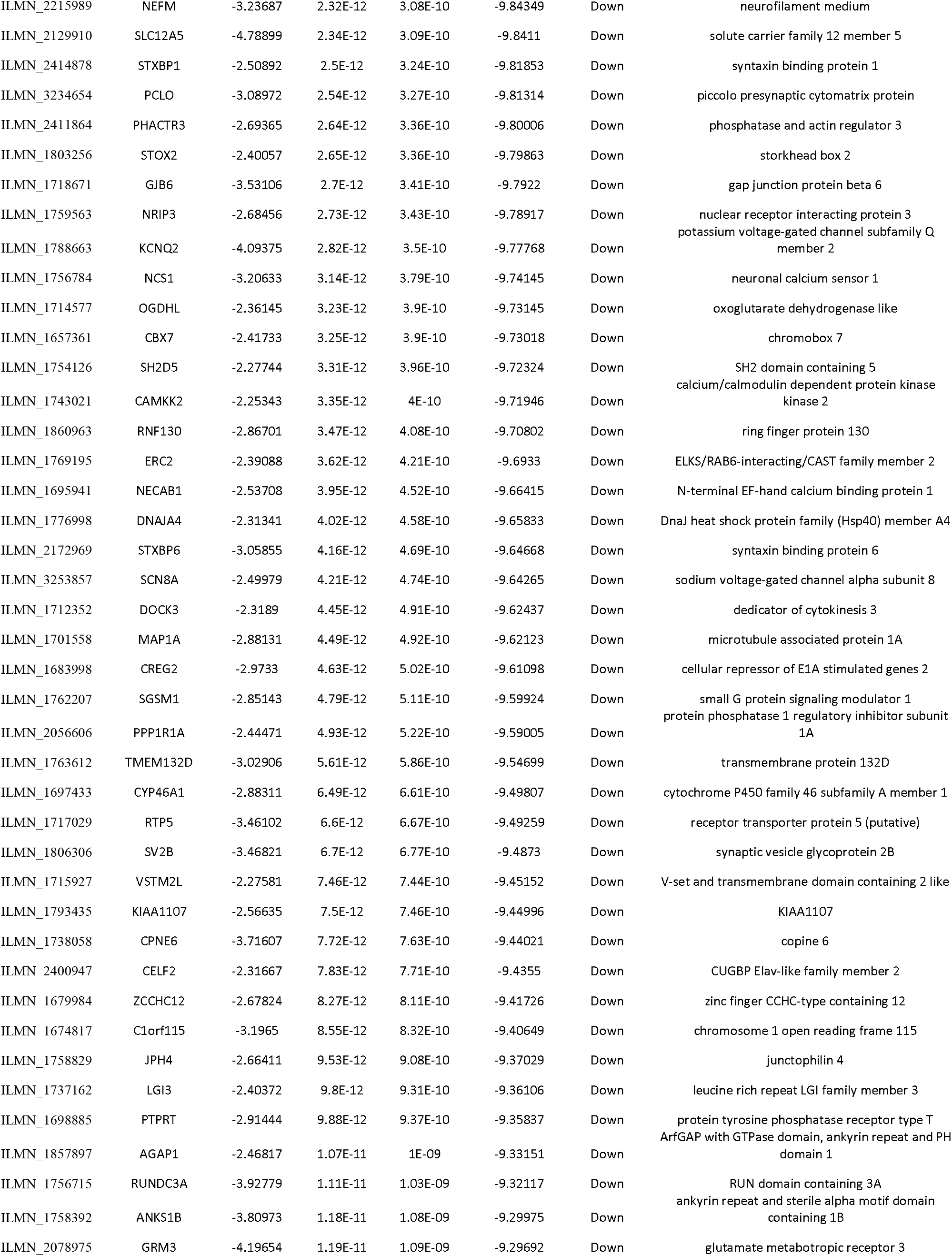
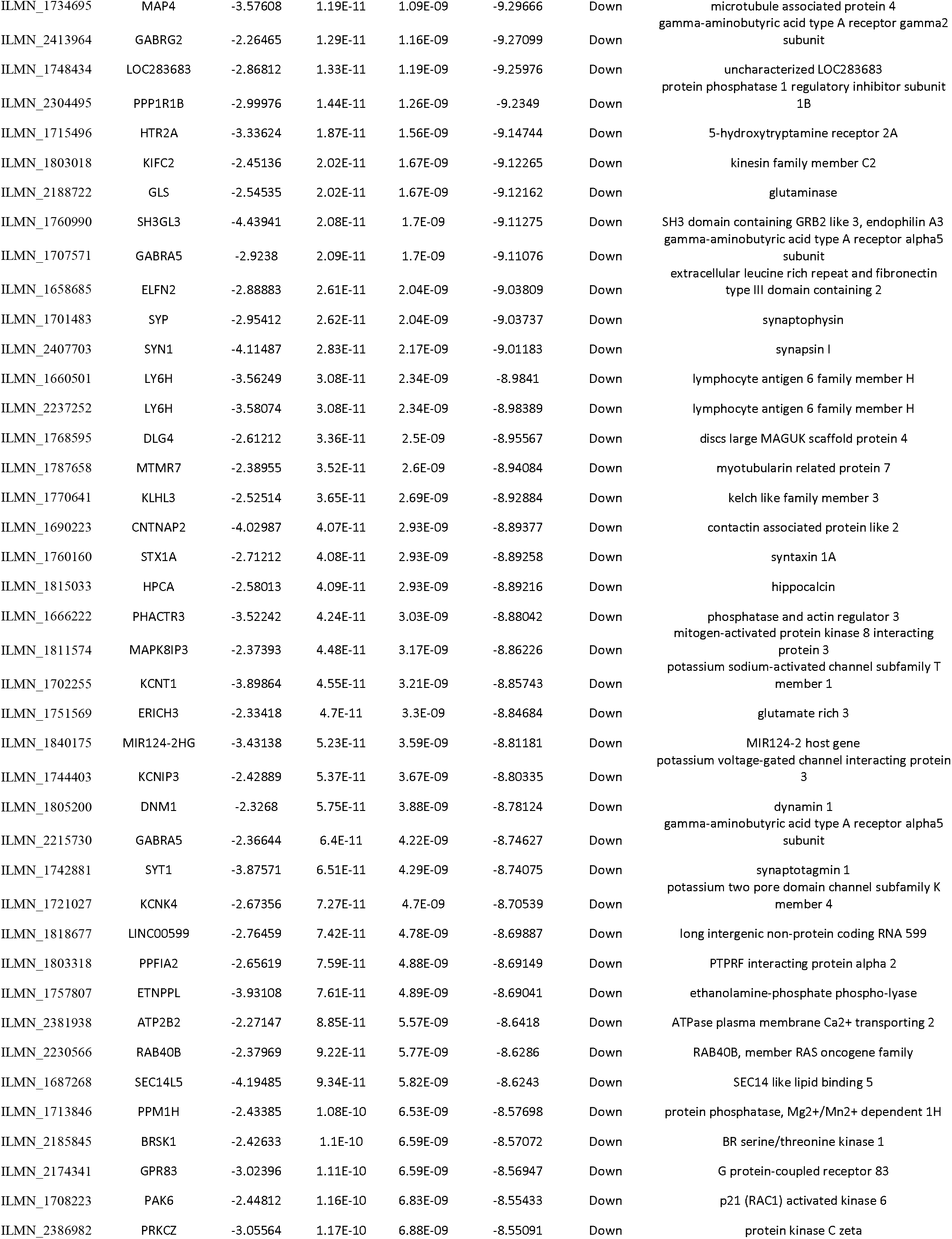
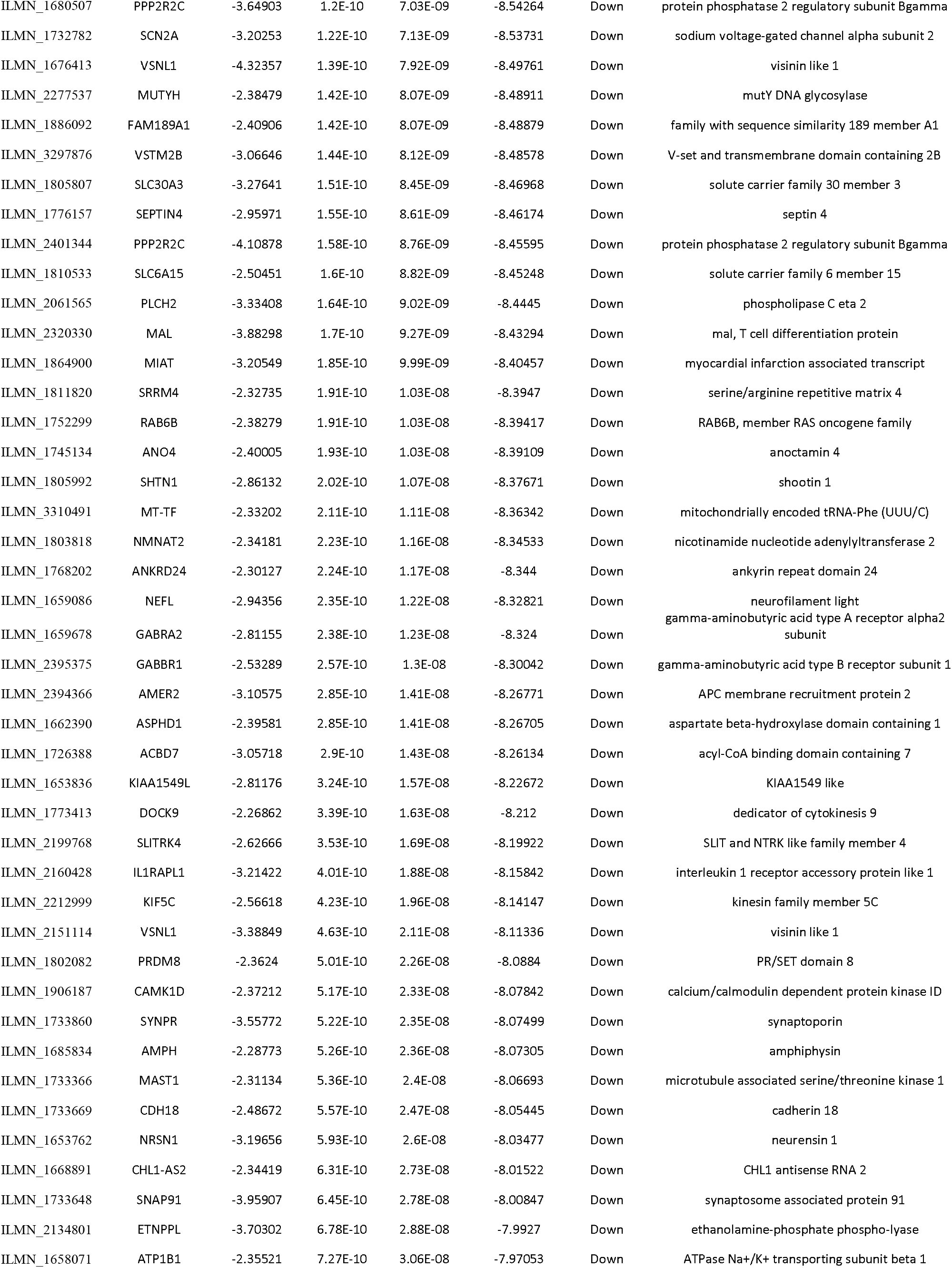
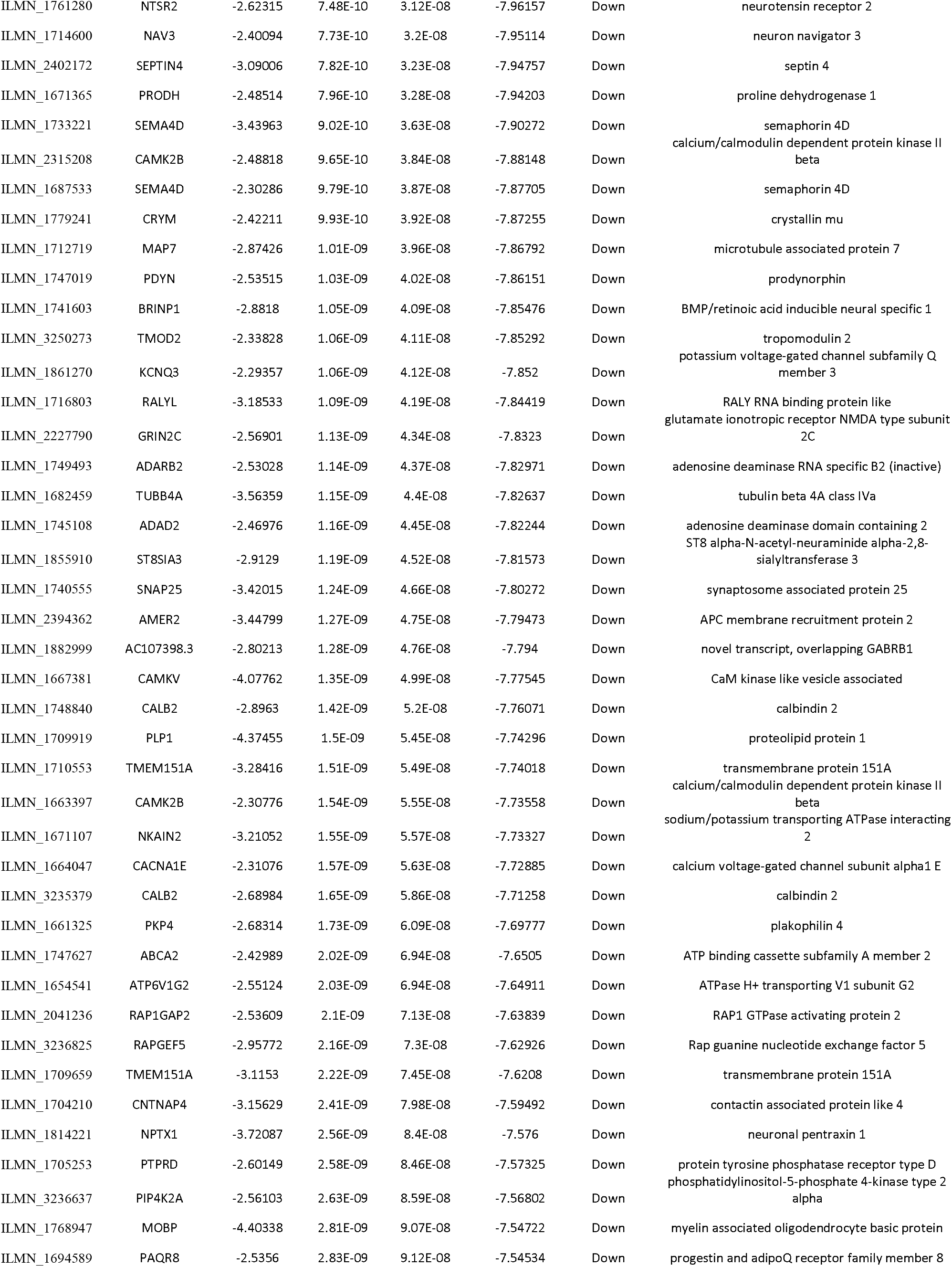
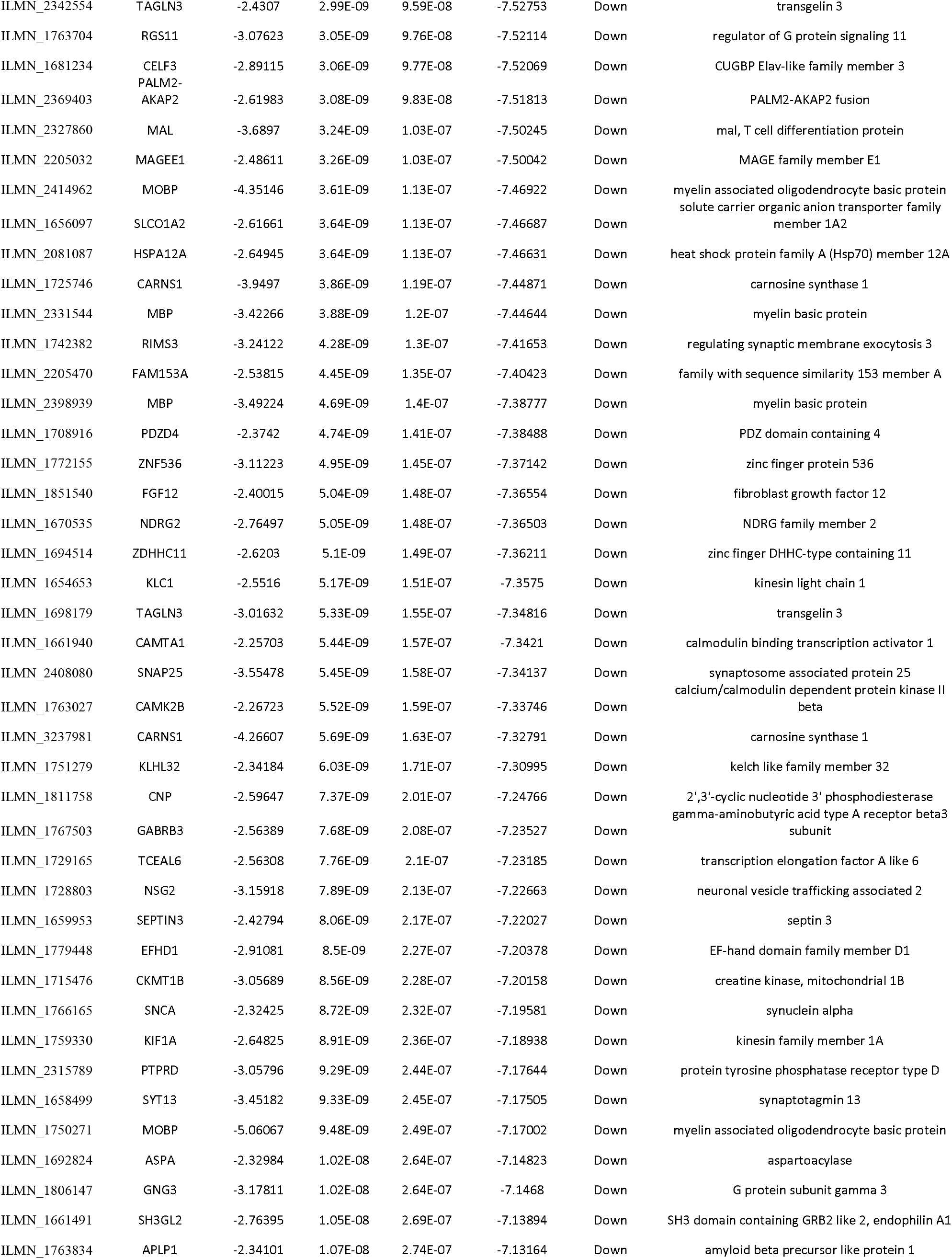
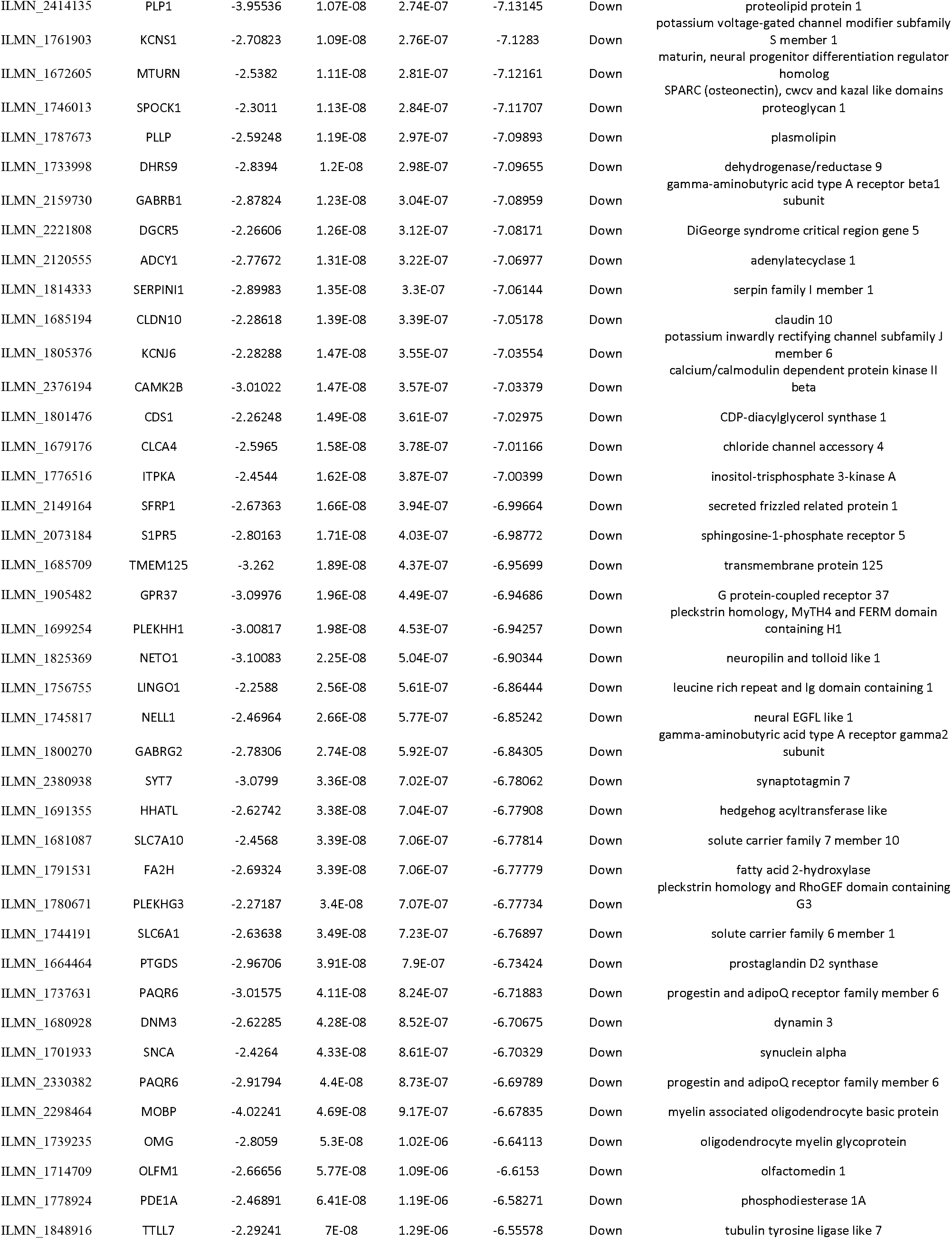
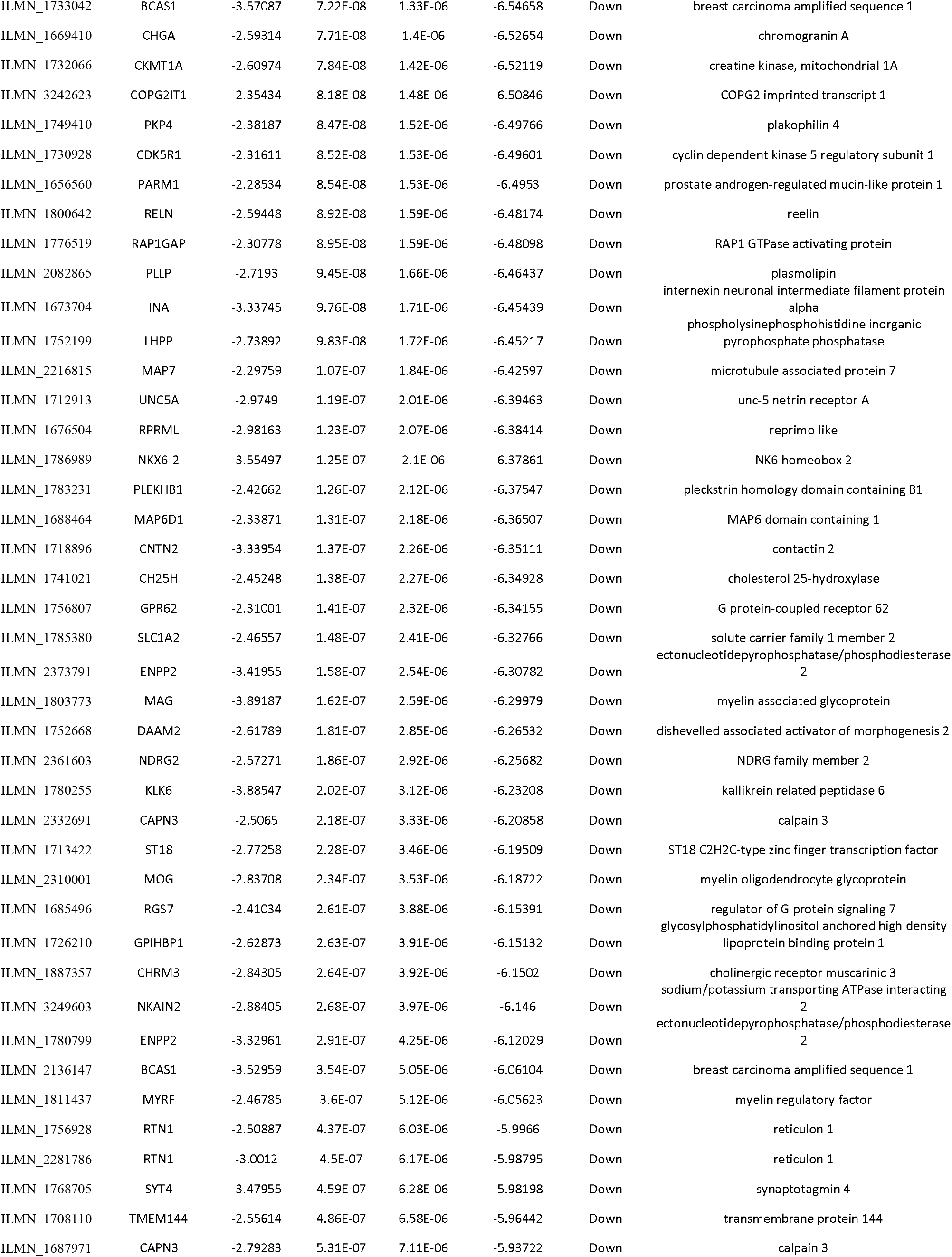
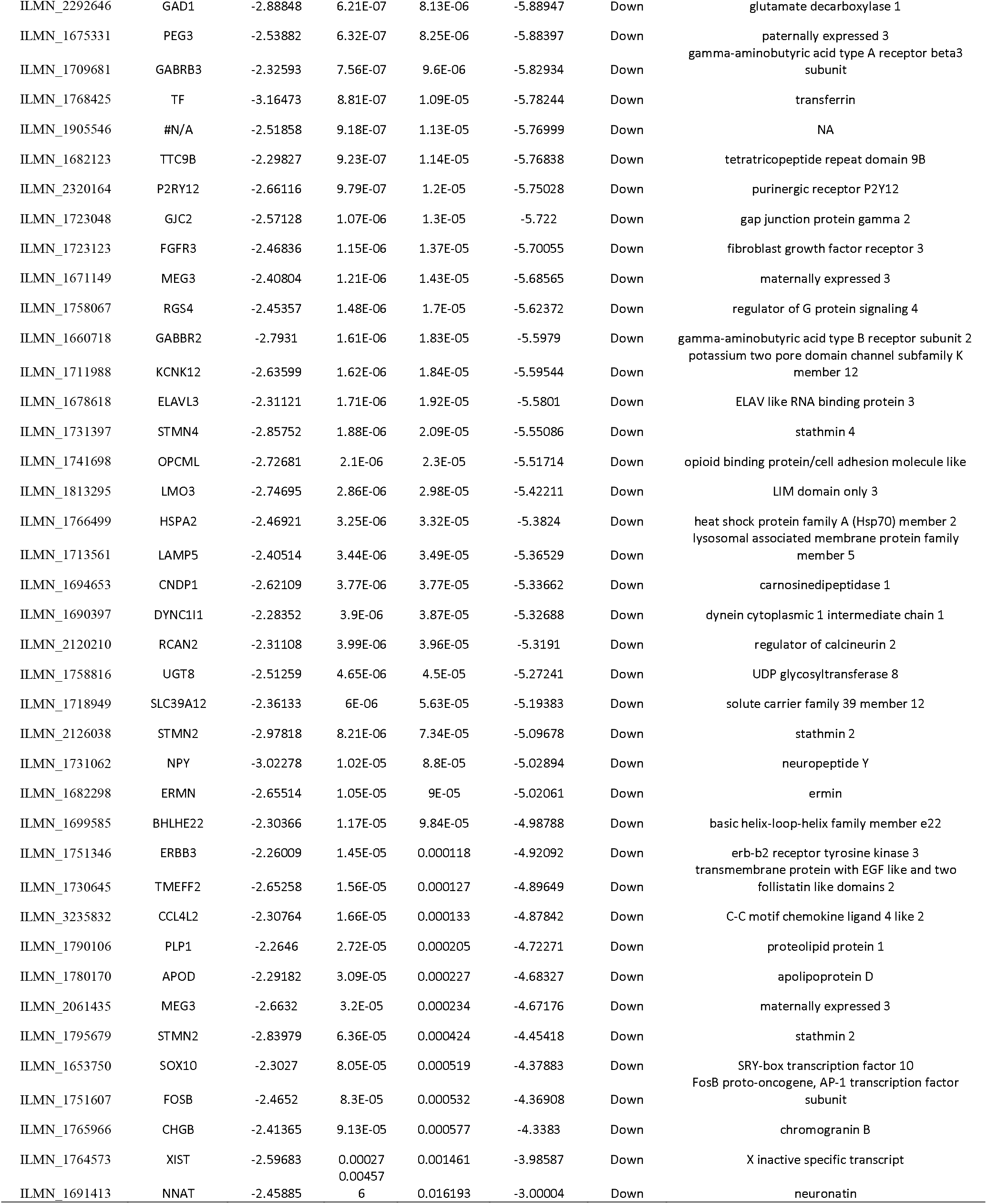
The statistical metrics for key differentially expressed genes (DEGs)

### Pathway enrichment analysis of DEGs

In order to investigate the biological functions of these DEGs (up and down regulated genes) in GBM. Pathway enrichment analysis was performed using ToppGene. Pathway enrichment analysis results indicated that DEGs (up and down regulated genes) were significantly enriched in reactive oxygen species degradation, glutamate removal from folates, ribosome, cell cycle, FOXM1 transcription factor network, PLK1 signaling events. translation, extracellular matrix organization, starch and sucrose_metabolism, nitrogen_metabolism, ensemble of genes encoding core extracellular matrix including ECM glycoproteins, collagens and proteoglycans, ensemble of genes encoding extracellular matrix and extracellular matrix-associated proteins, integrin signalling pathway, p53 pathway, hypertension, G2/M DNA replication checkpoint, and nicotinate and nicotinamide metabolism, homocarnosine biosynthesis, fatty acid alpha-oxidation III, GABAergic synapse, insulin secretion, effects of botulinumtoxin, internalization of ErbB1, neuronal system, transmission across chemical synapses, alanine and aspartate metabolism, glycans biosynthesis, Wnt/Ca2+/cyclic GMP signaling., fl-arrestins in GPCR desensitization, synaptic vesicle trafficking, muscarinic acetylcholine receptor 1 and 3 signaling pathway, insulin secretion pathway, glutamate metabolic, pirenzepine pathway and homocarnosinosis are listed in Table 2 and Table 3.

**Table 2.**
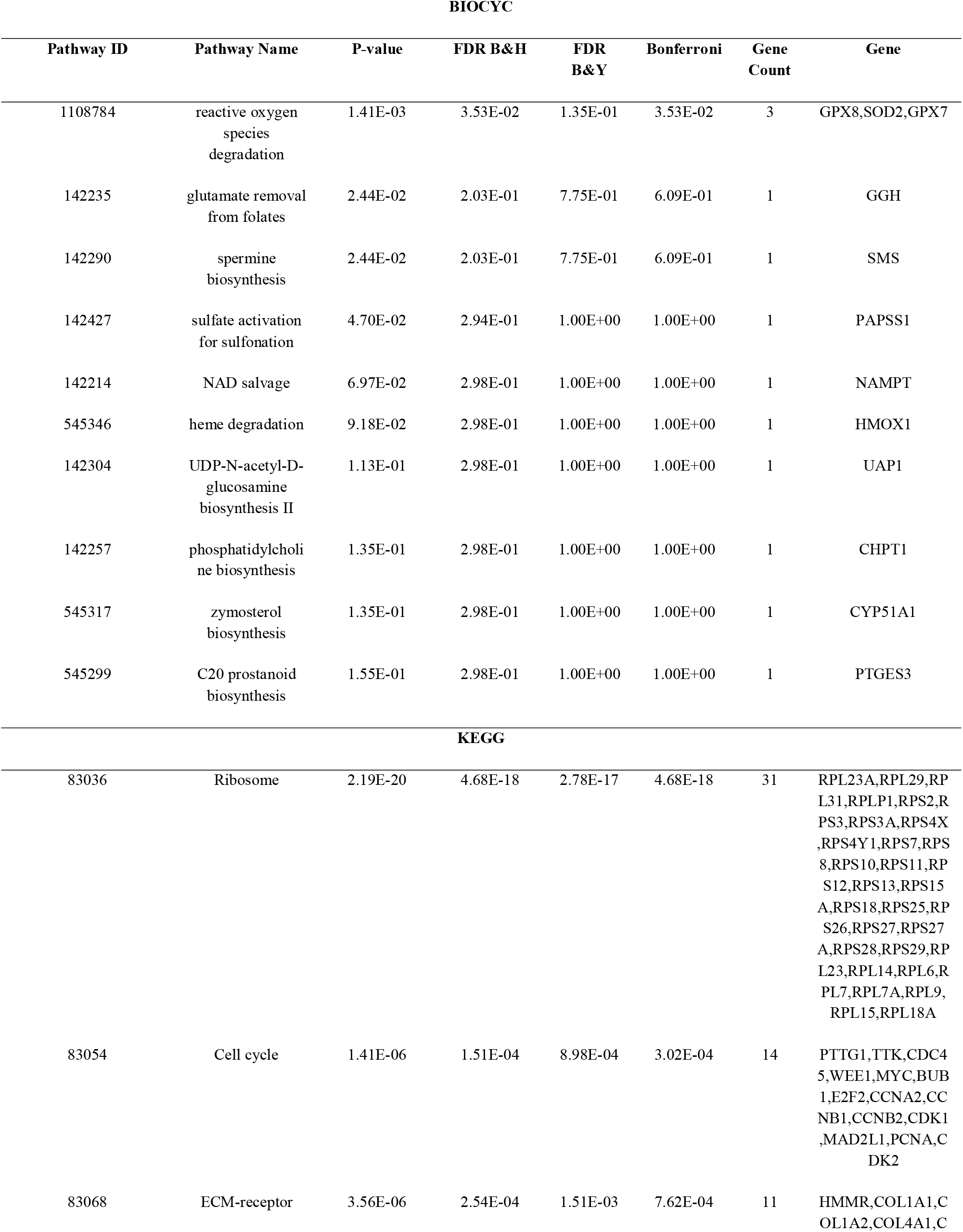
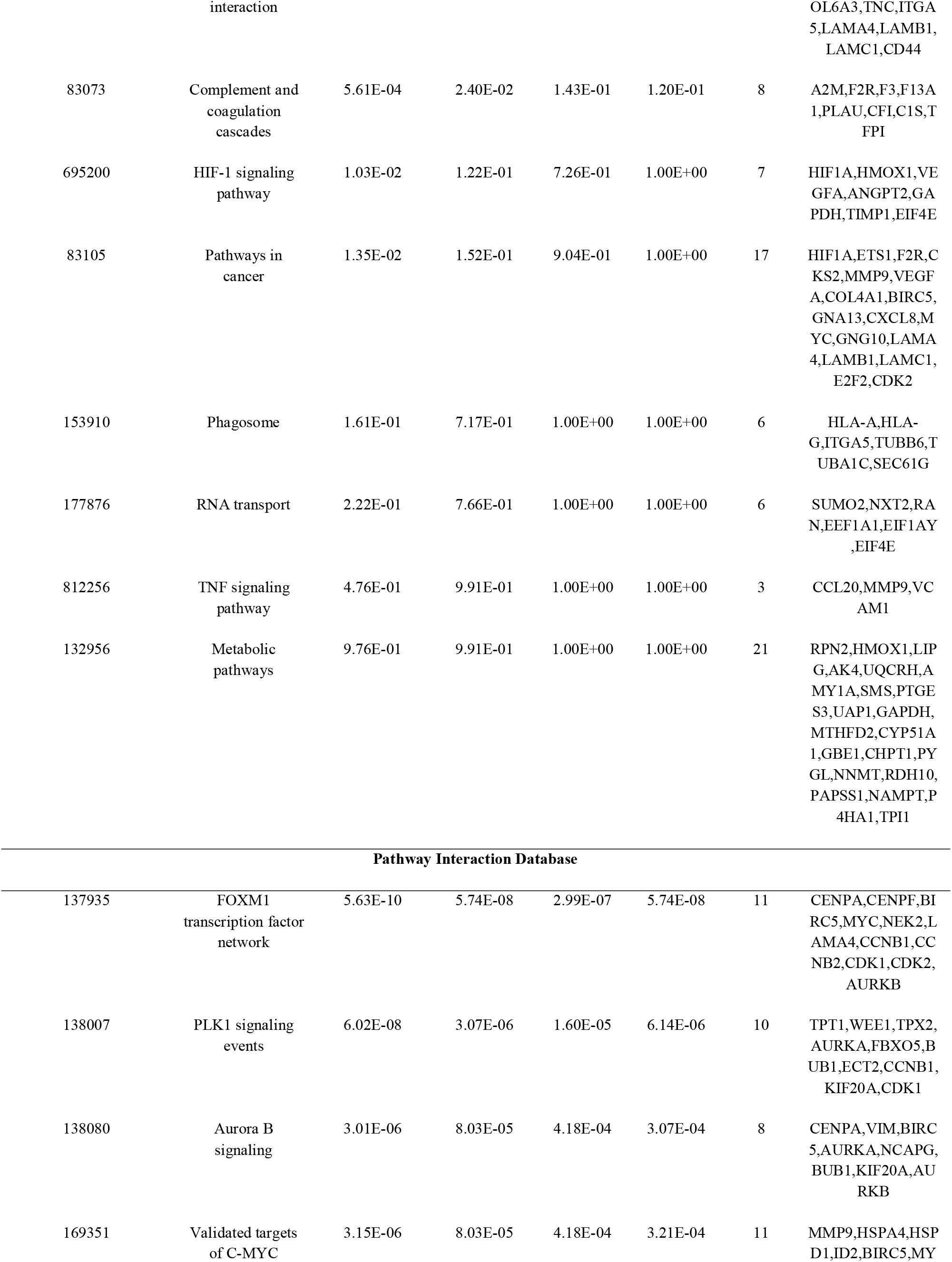
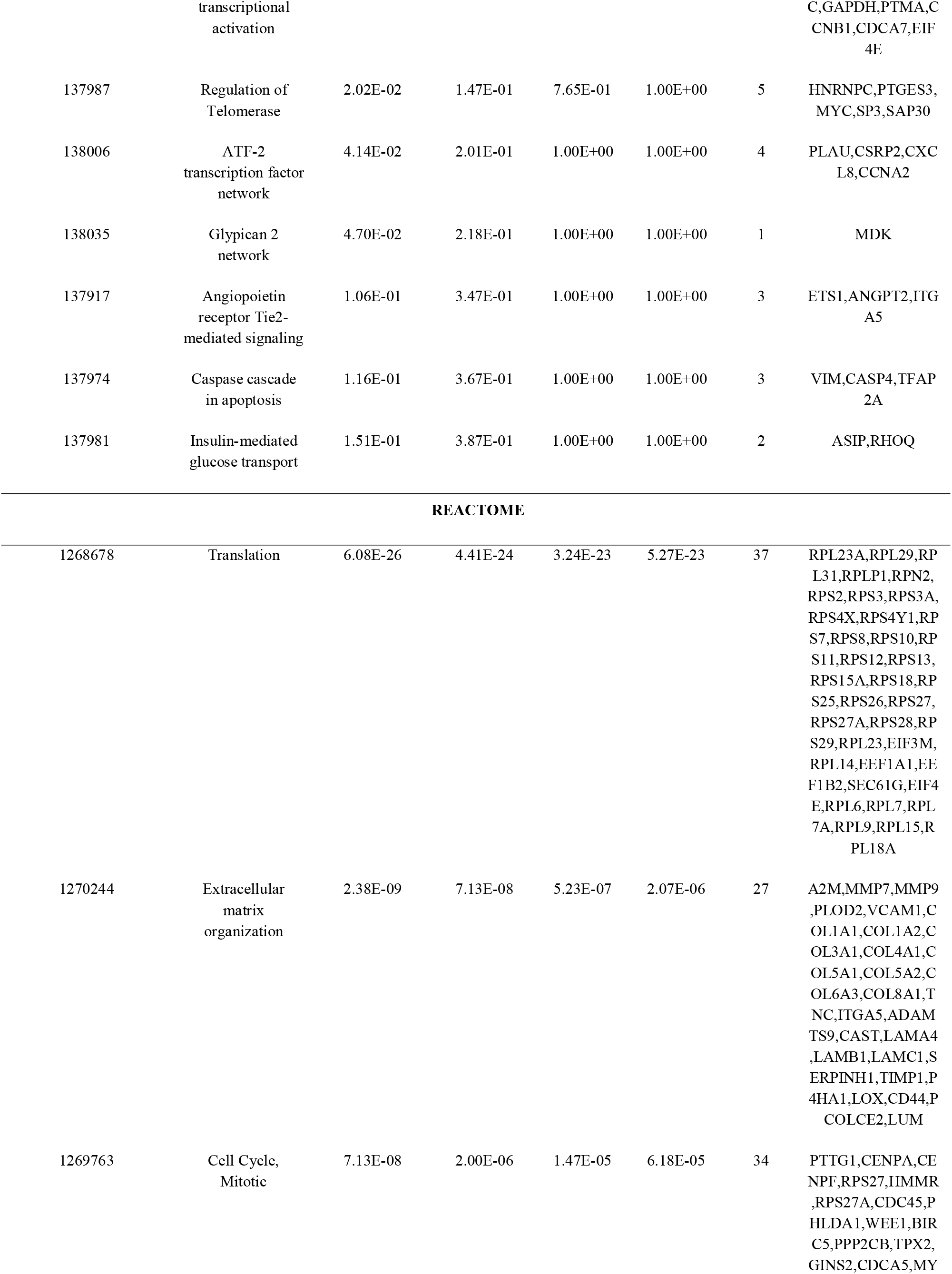
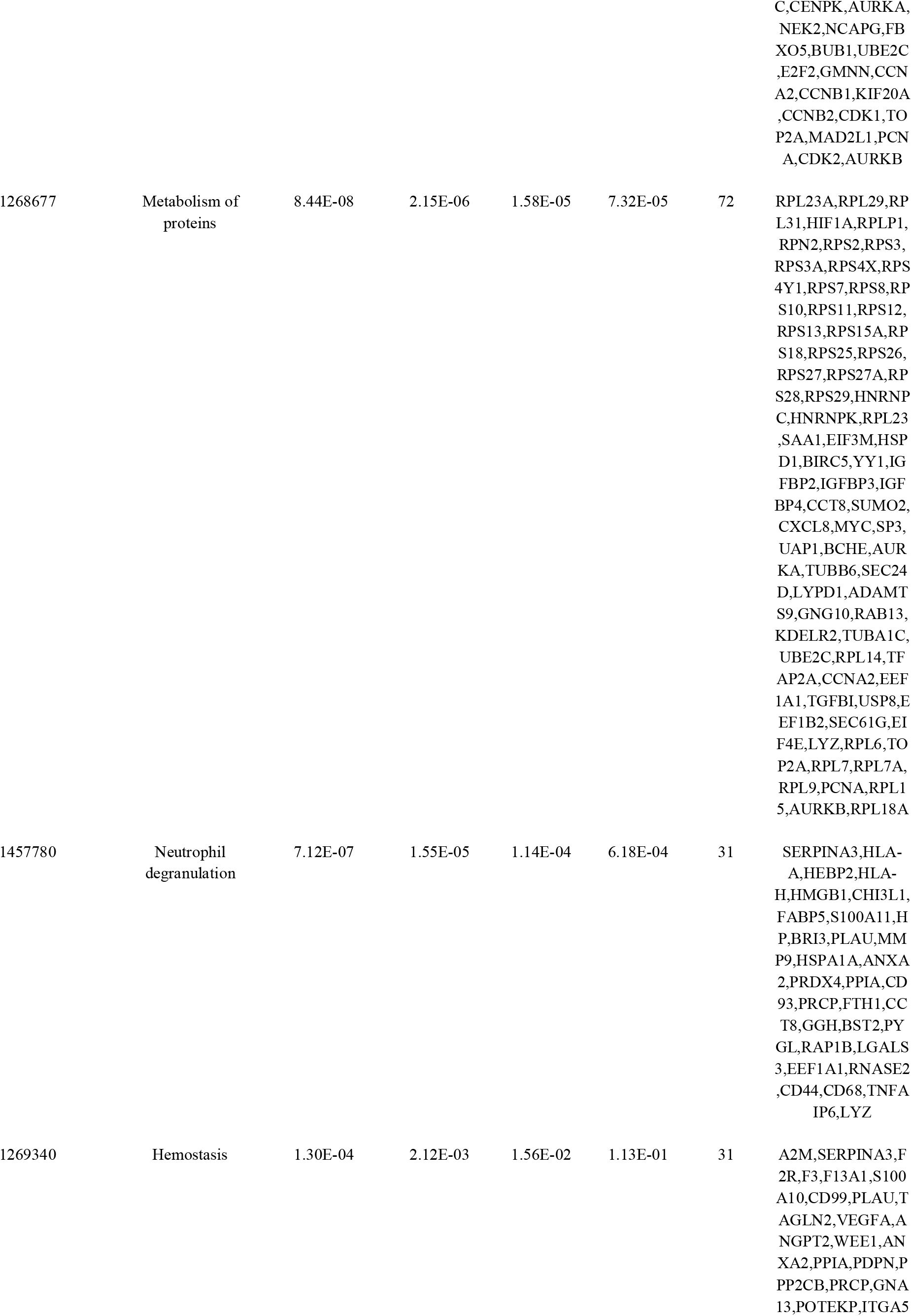
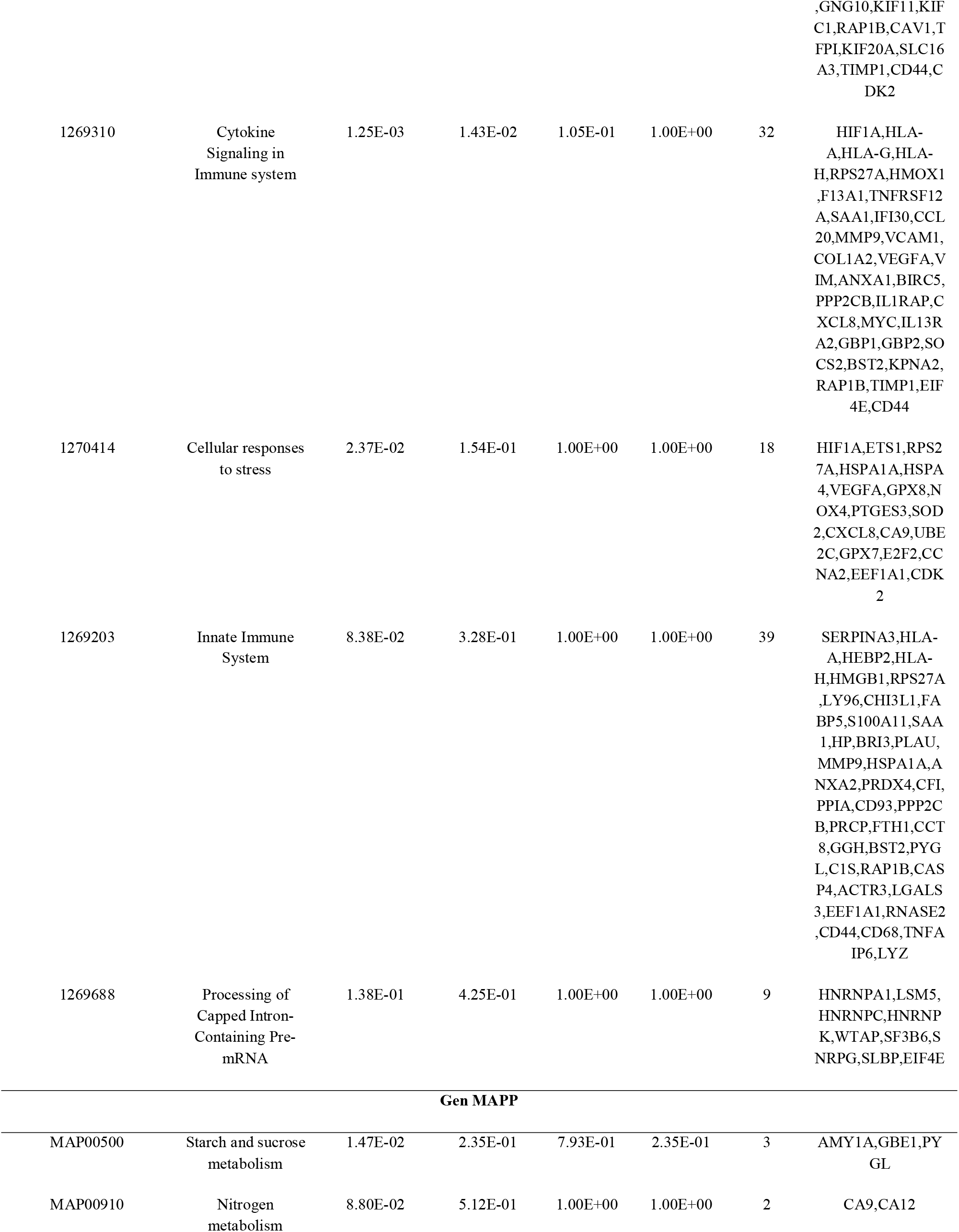
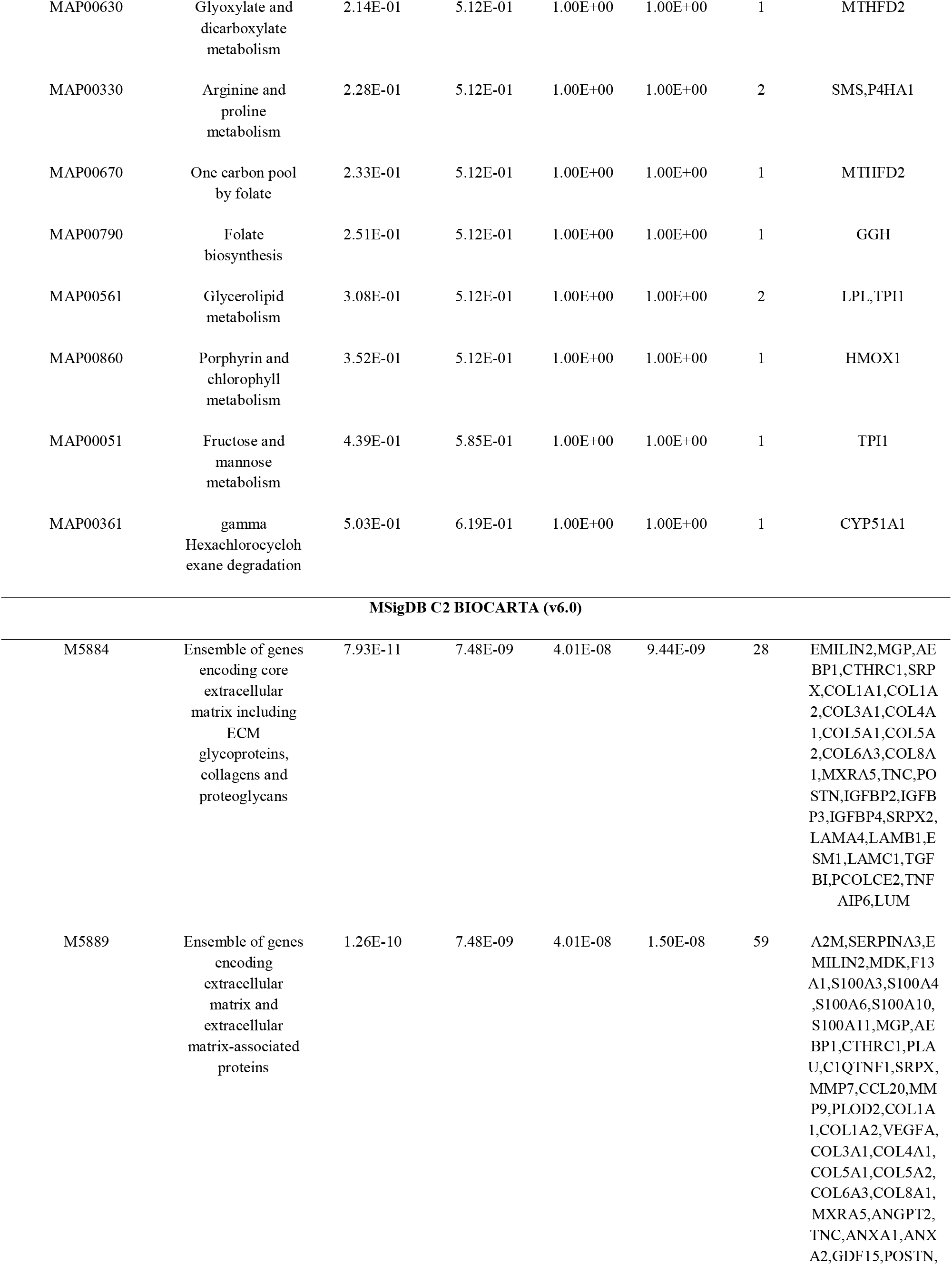
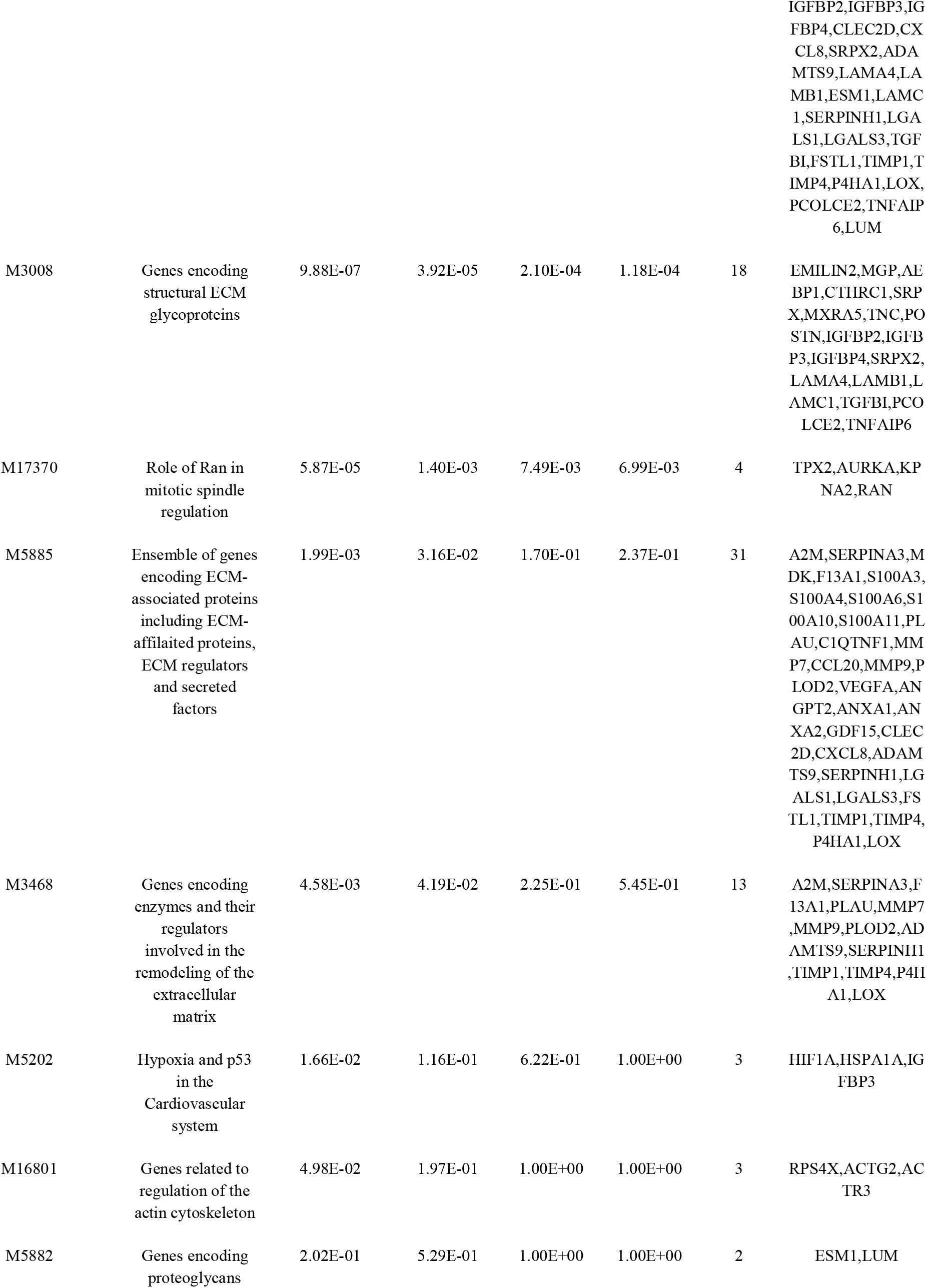
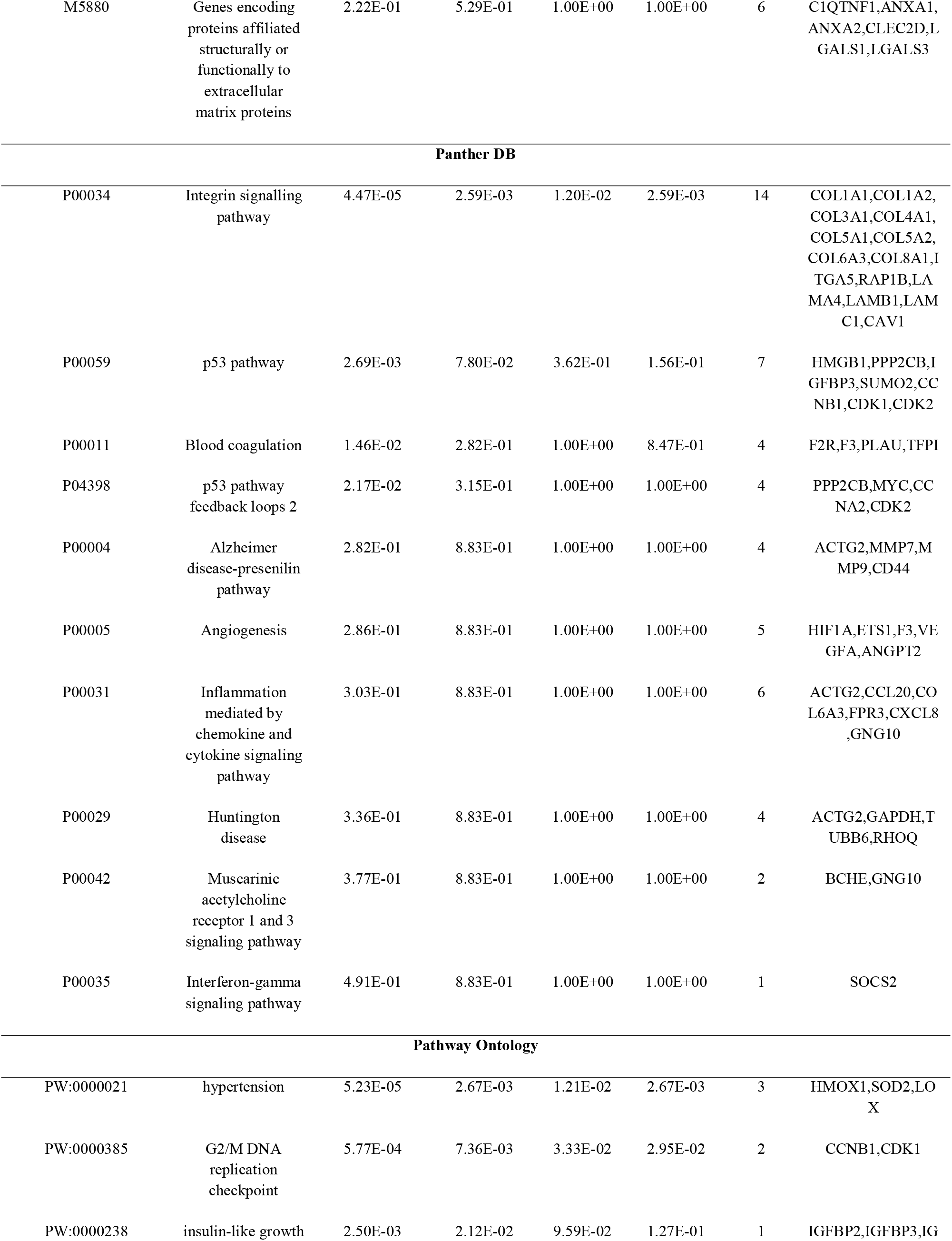
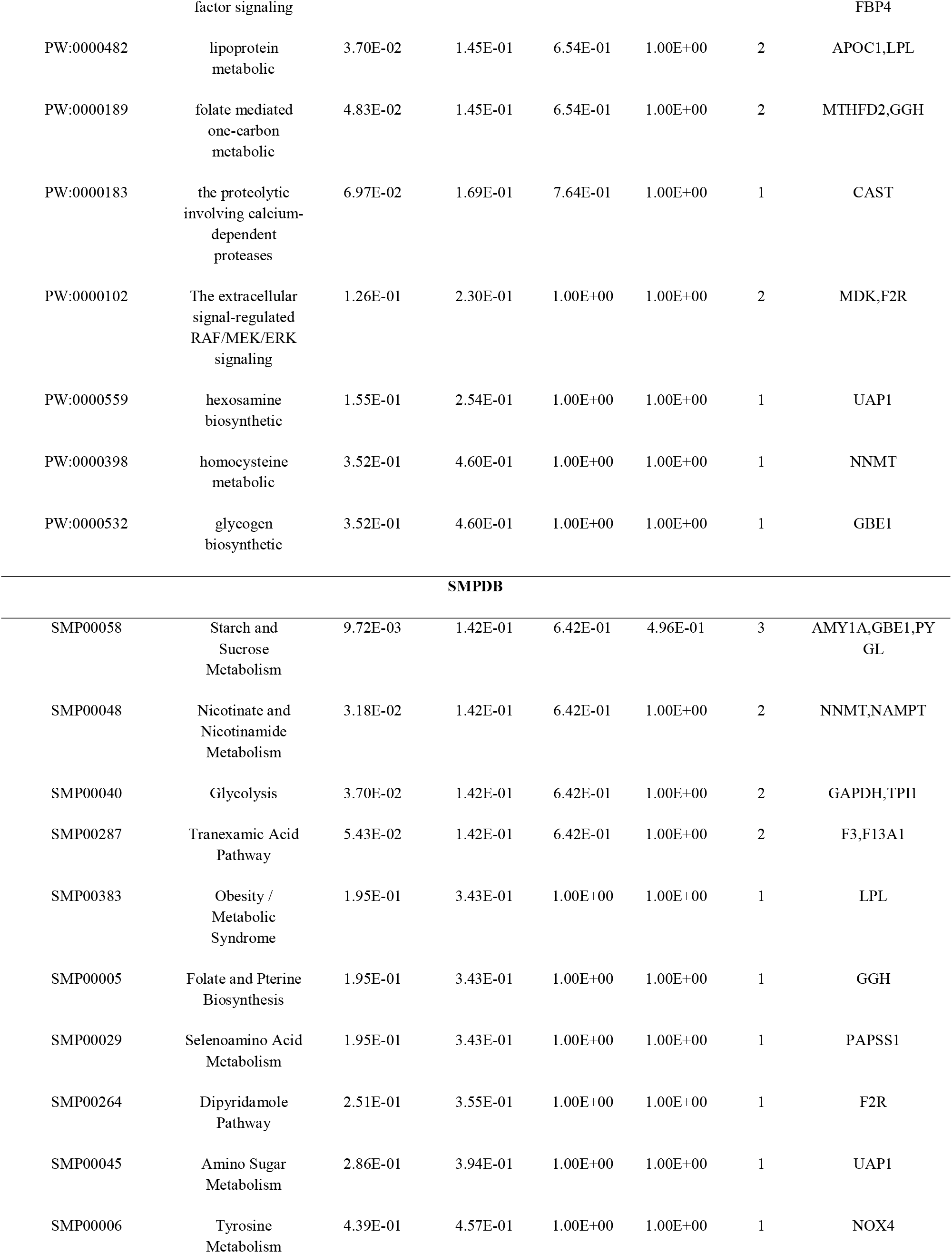
The enriched pathway terms of the up regulated differentially expressed genes

**Table 3.**
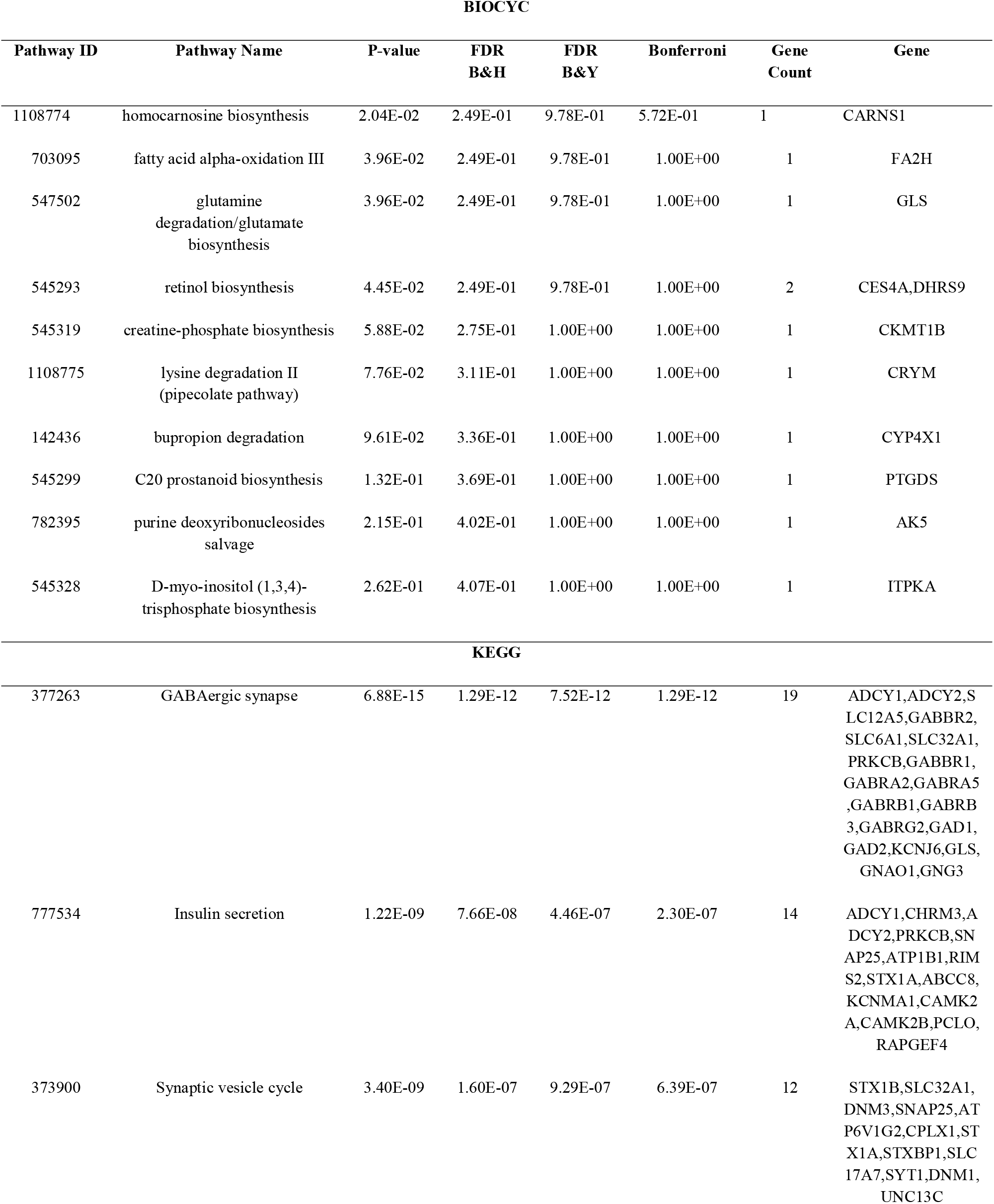
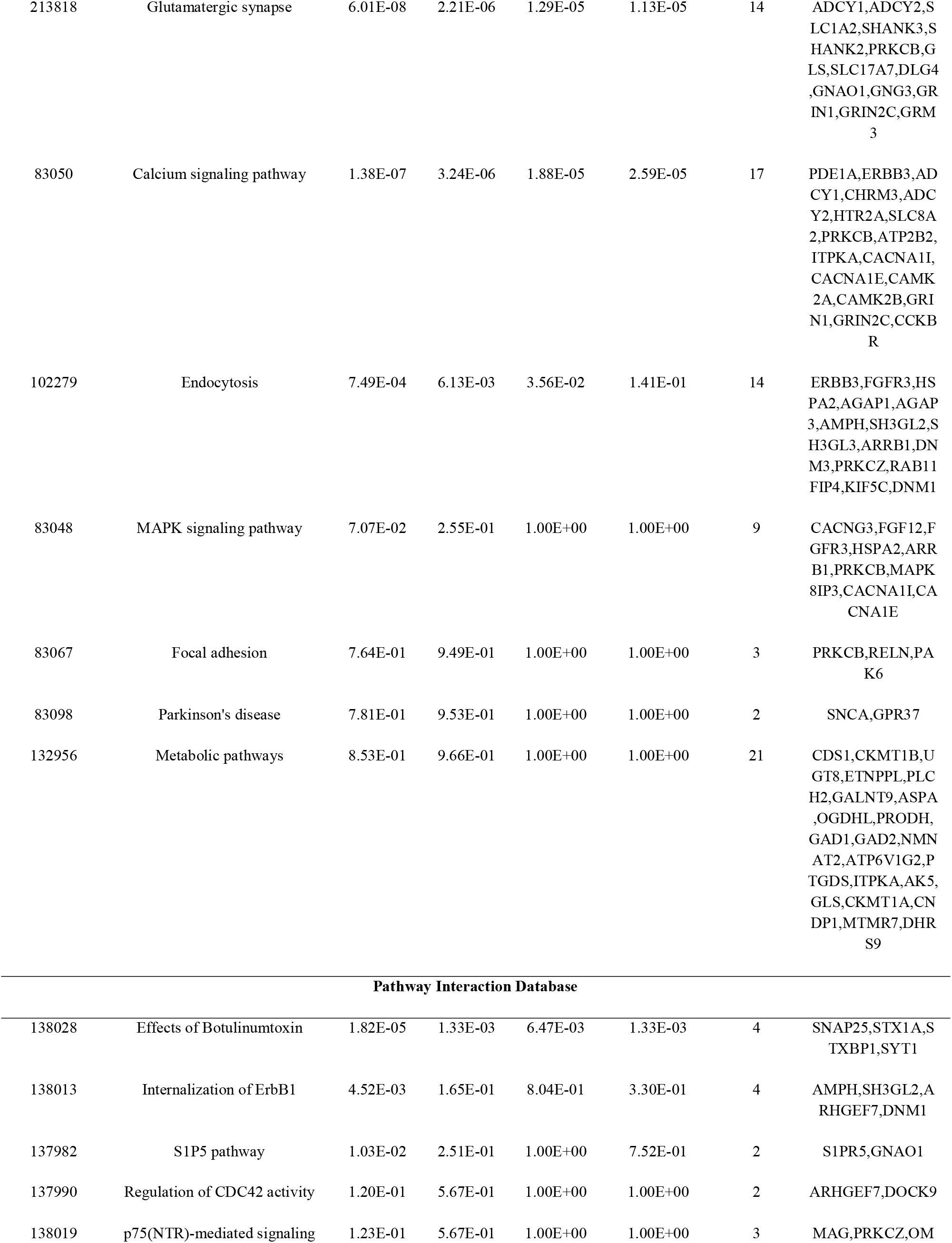
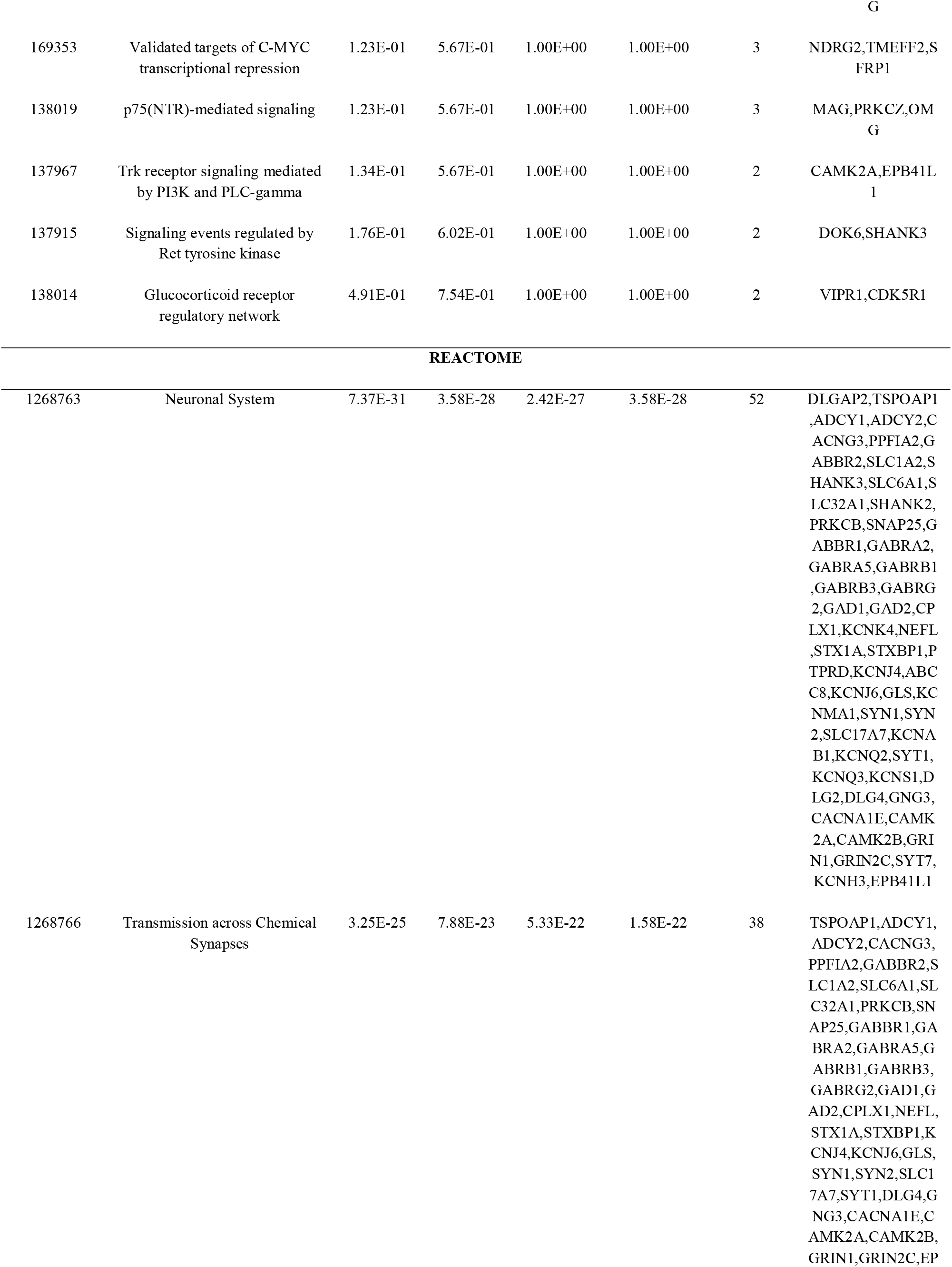
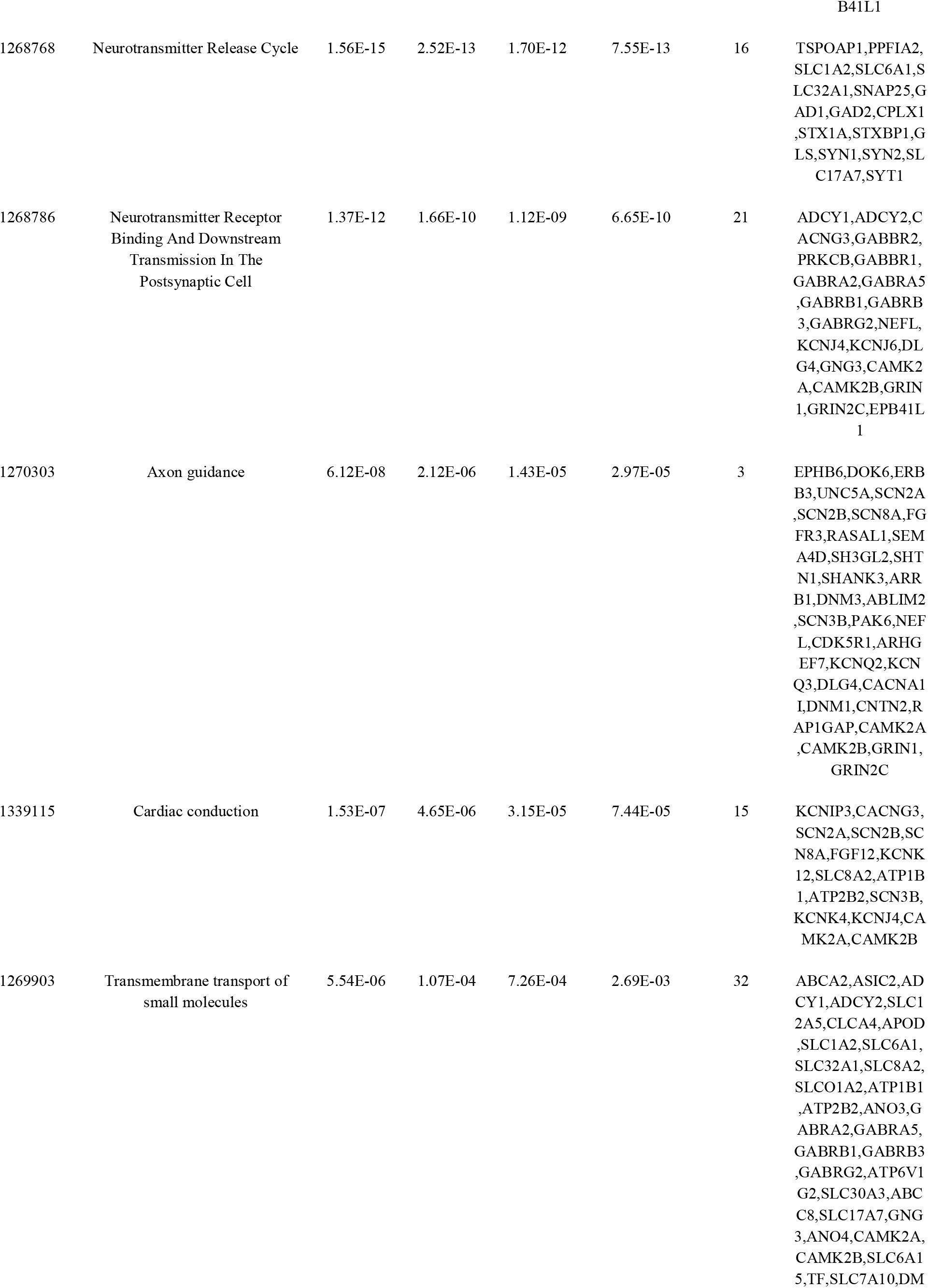
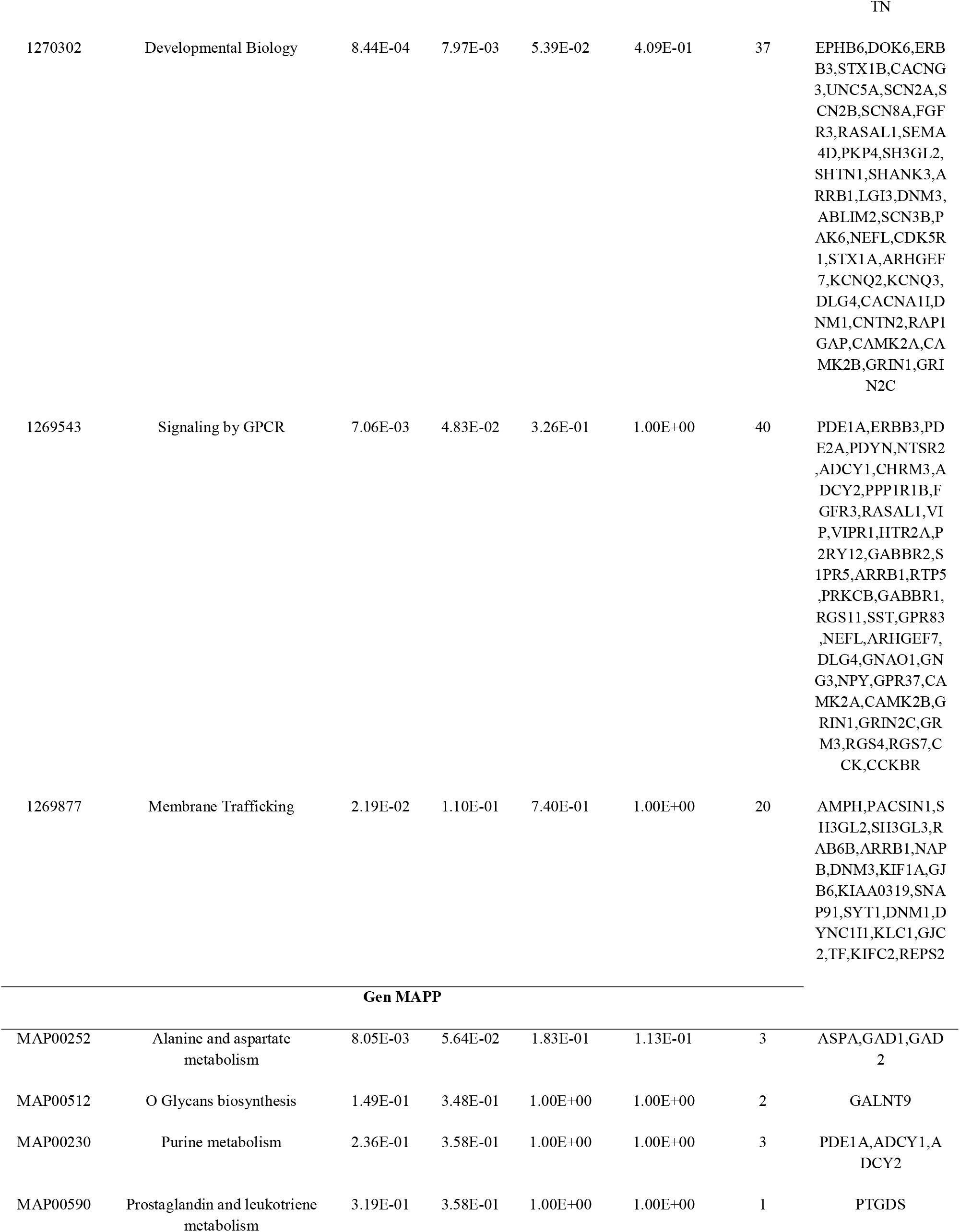
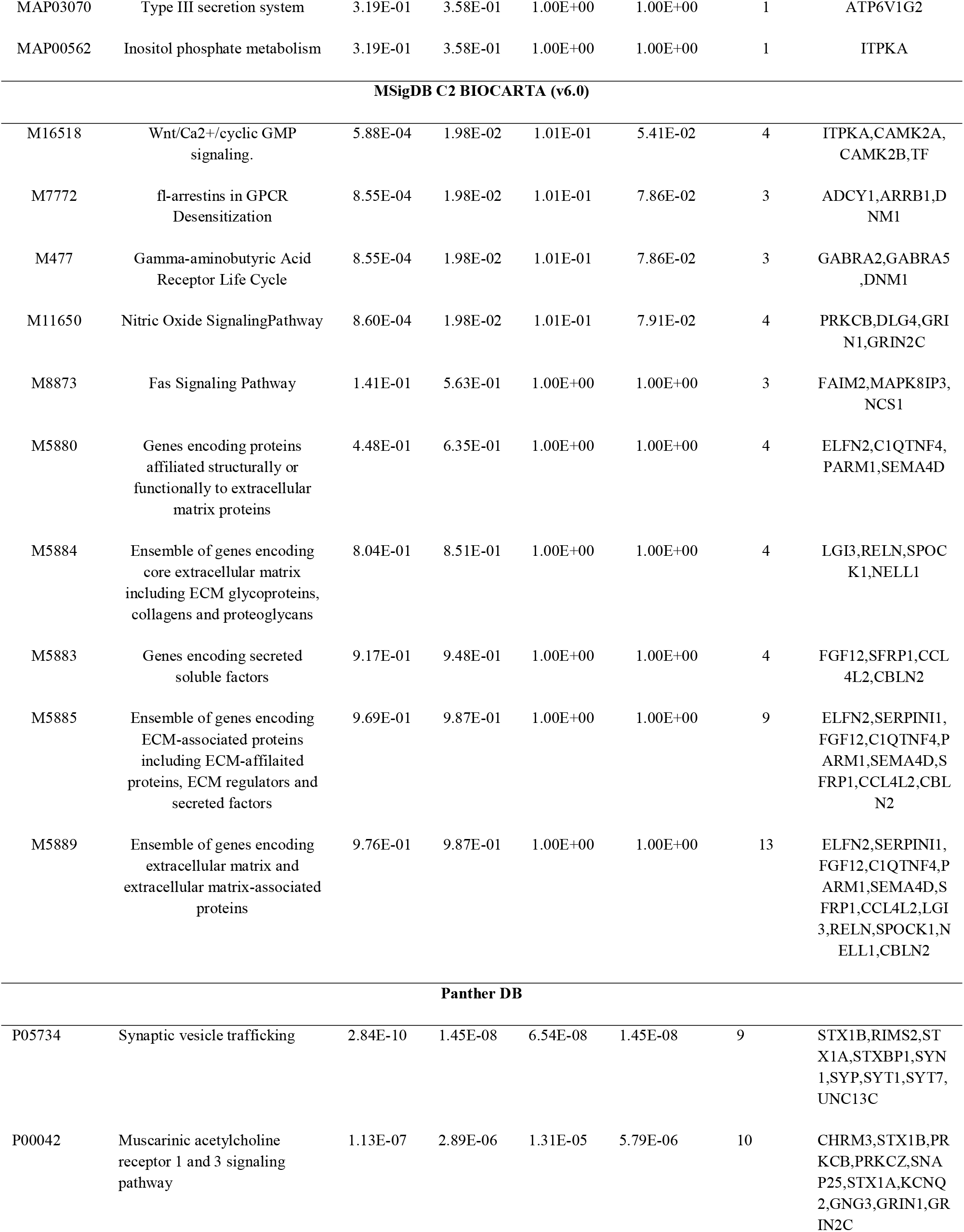
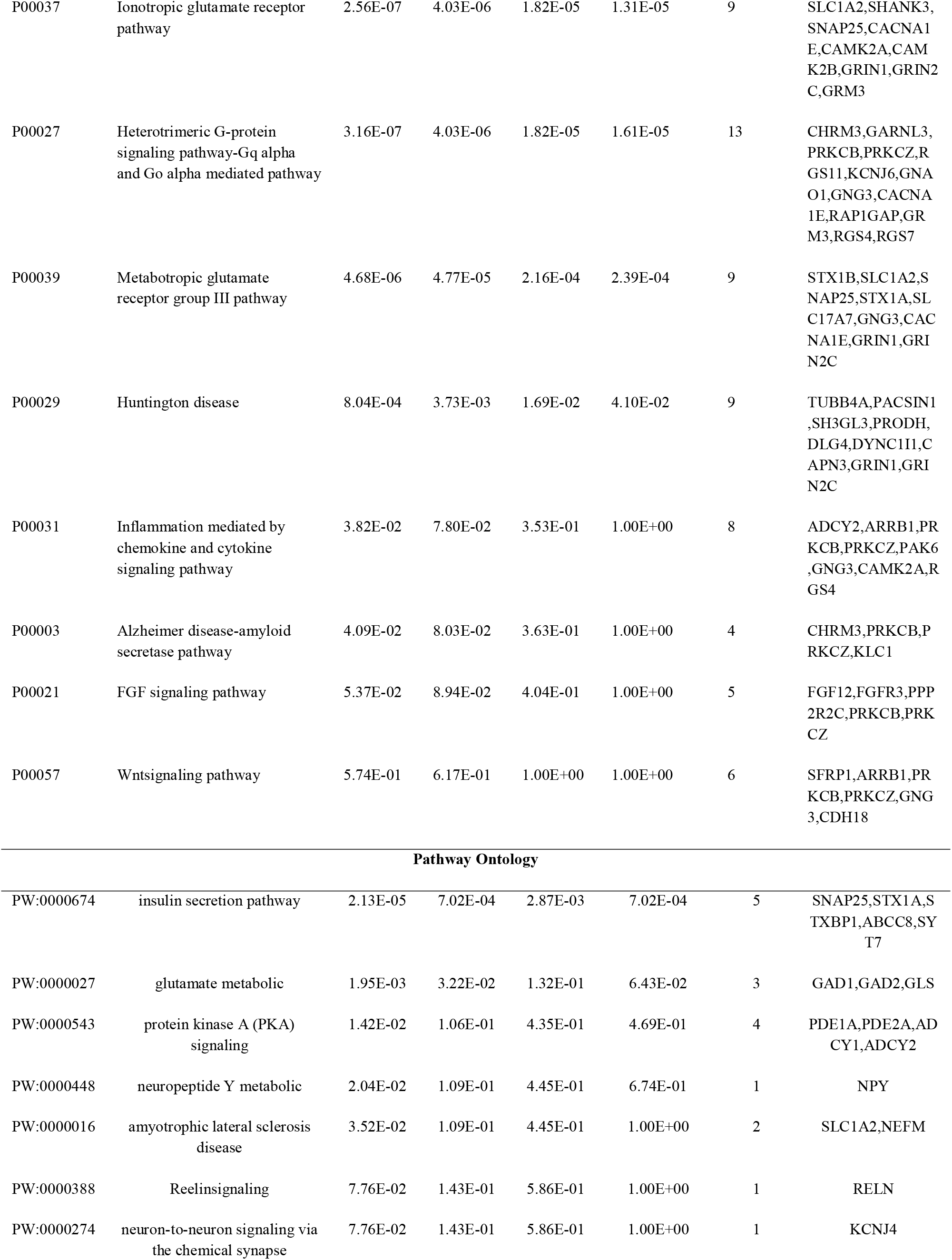
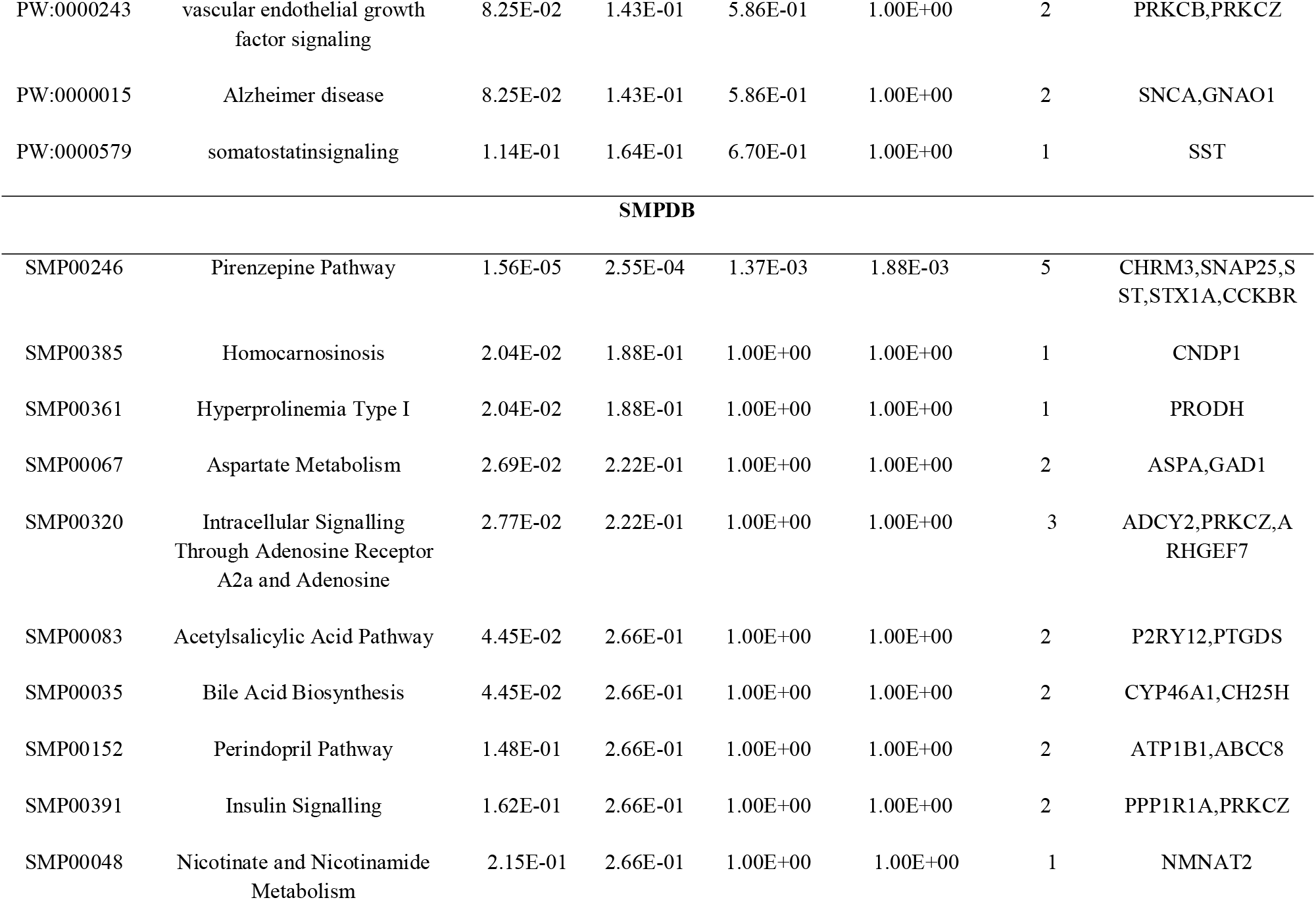
The enriched pathway terms of the down regulated differentially expressed genes

### Gene ontology enrichment analysis of DEGs

GO enrichment analysis was conducted using the ToppGene, and the results are illustrated in Table 4 and Table 5. For up regulated genes, the terms enriched in the BP category included macromolecule catabolic process and mitotic cell cycle. The GO CC category revealed enrichment in the cytosolic part and collagen-containing extracellular matrix. In addition, the MF category showed enrichment for factors involved in structural molecule activity and RNA binding. Down regulated genes showed enrichment in the BP category in processes such as synaptic signaling and cell-cell signaling. The enriched terms in the CC category mainly included synapse part and neuron projection. Additionally, the enriched MF was focused on ion gated channel activity and channel activity.

**Table 4.**
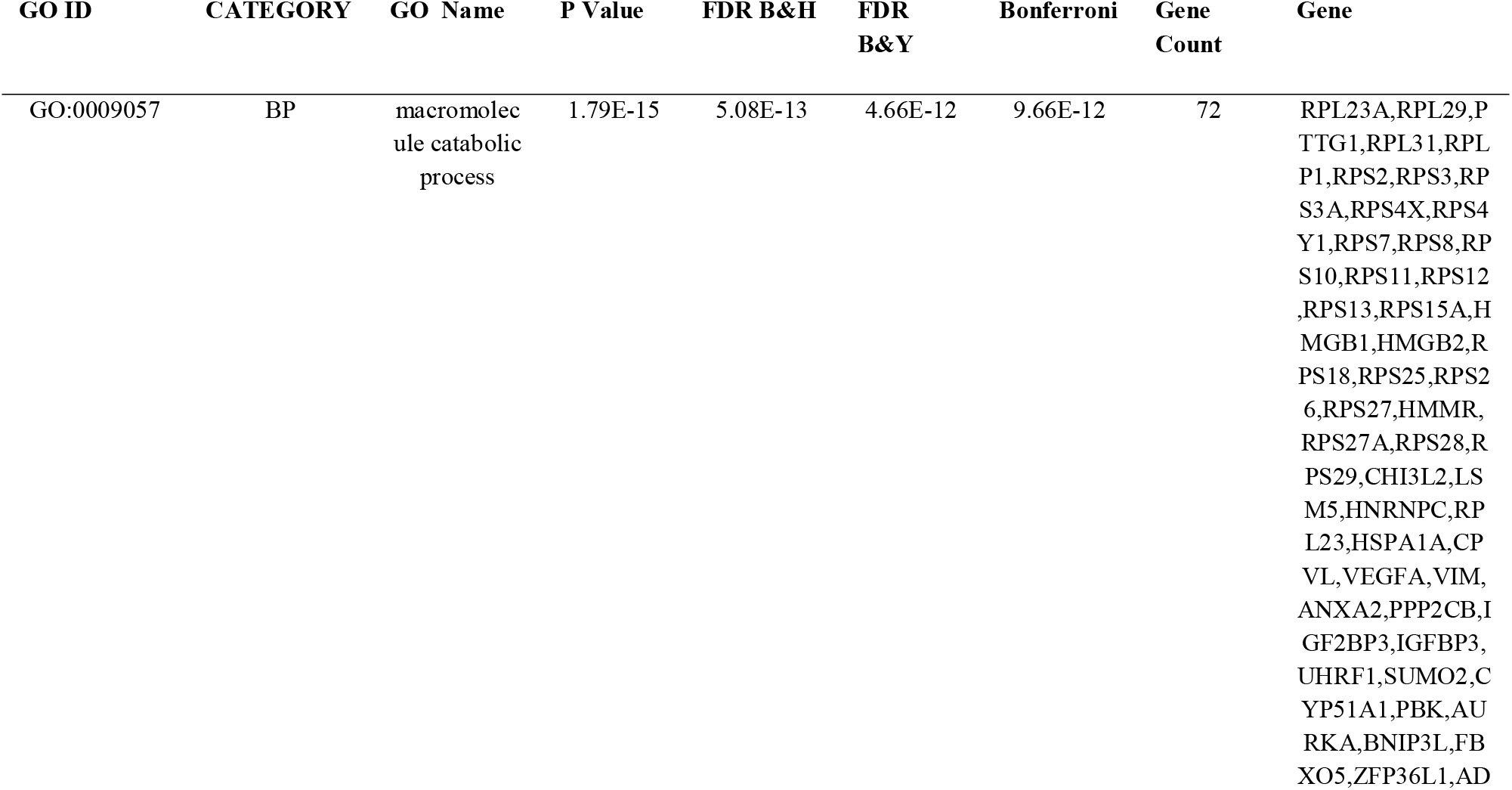
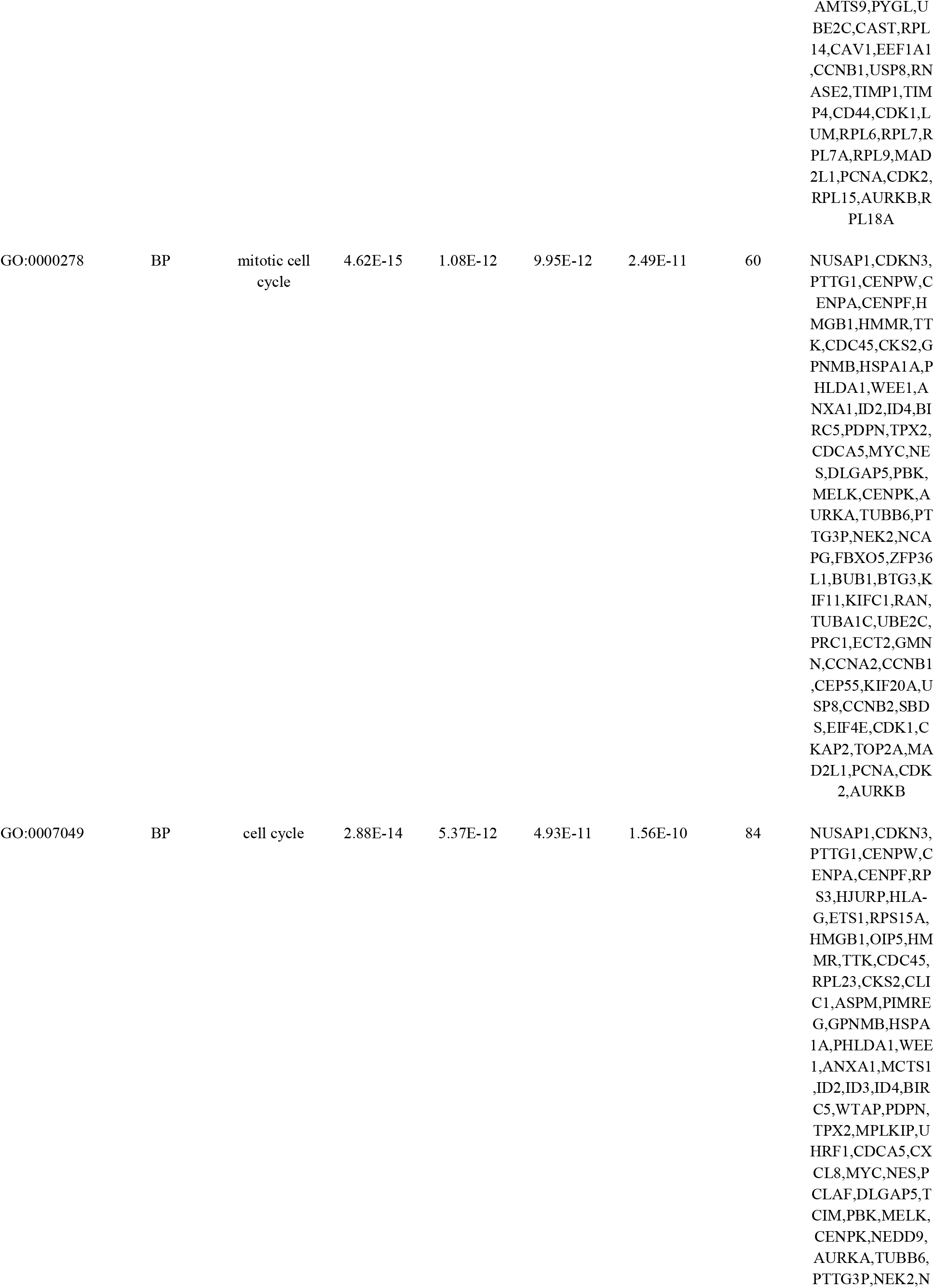
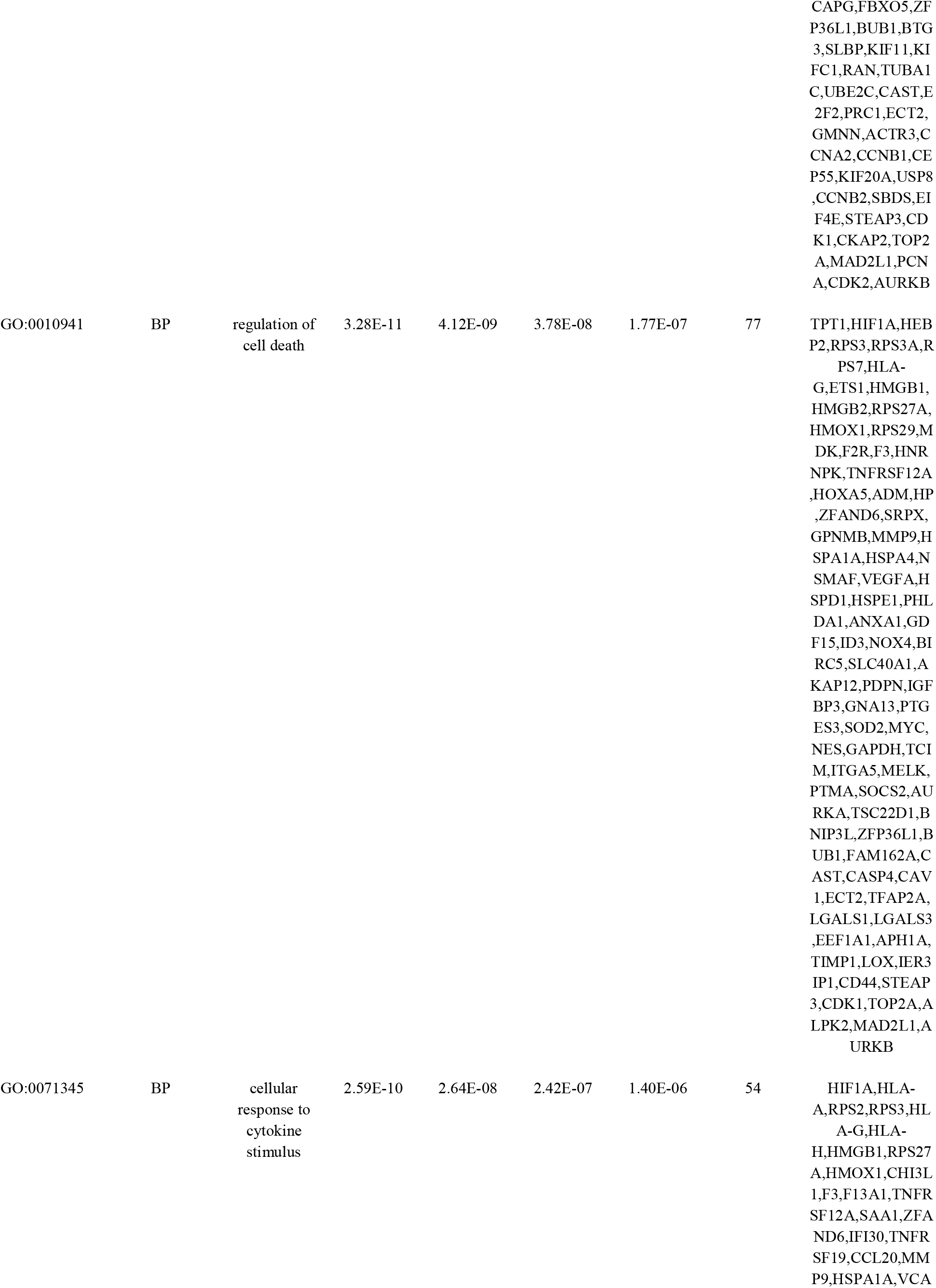
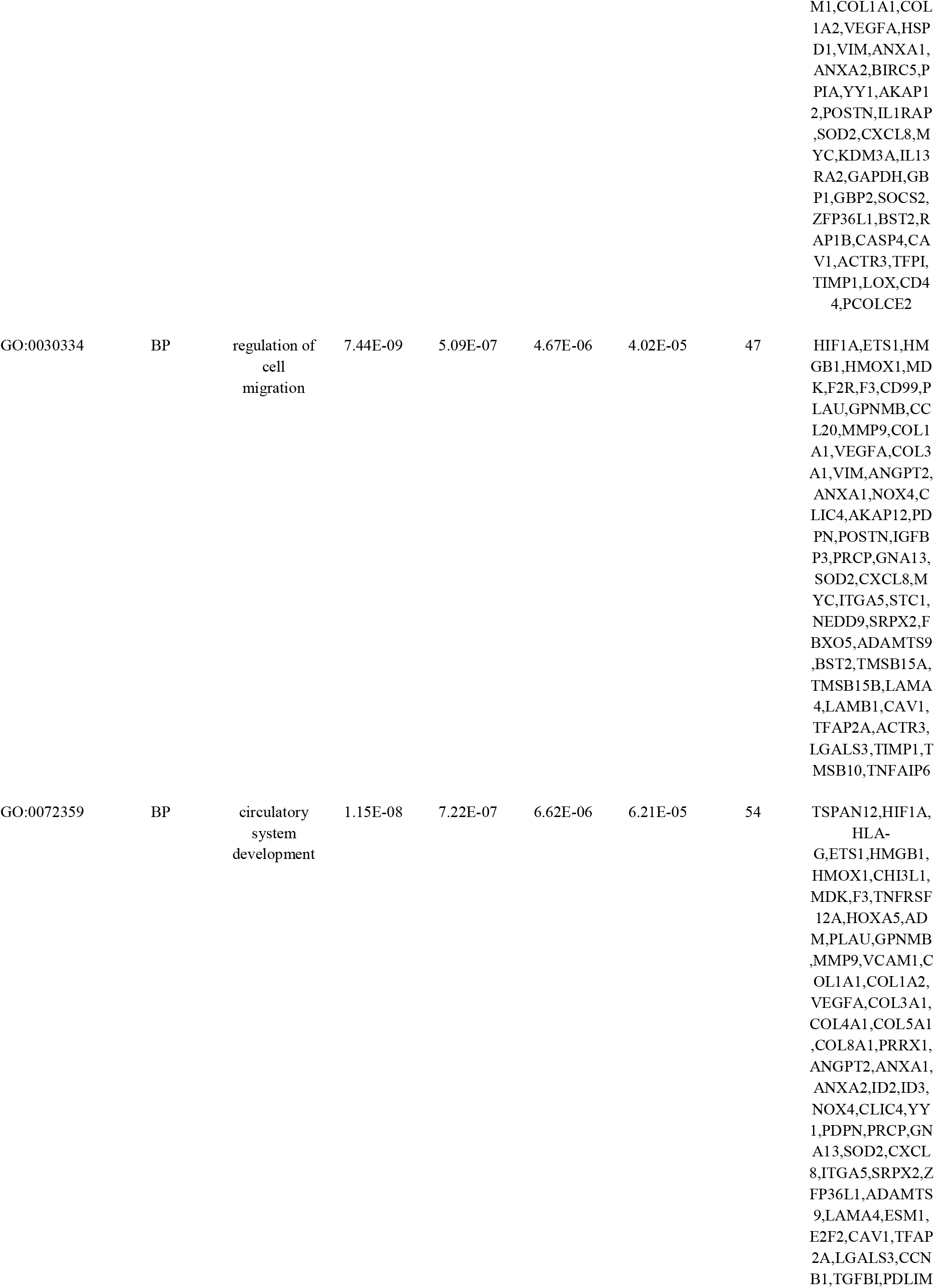
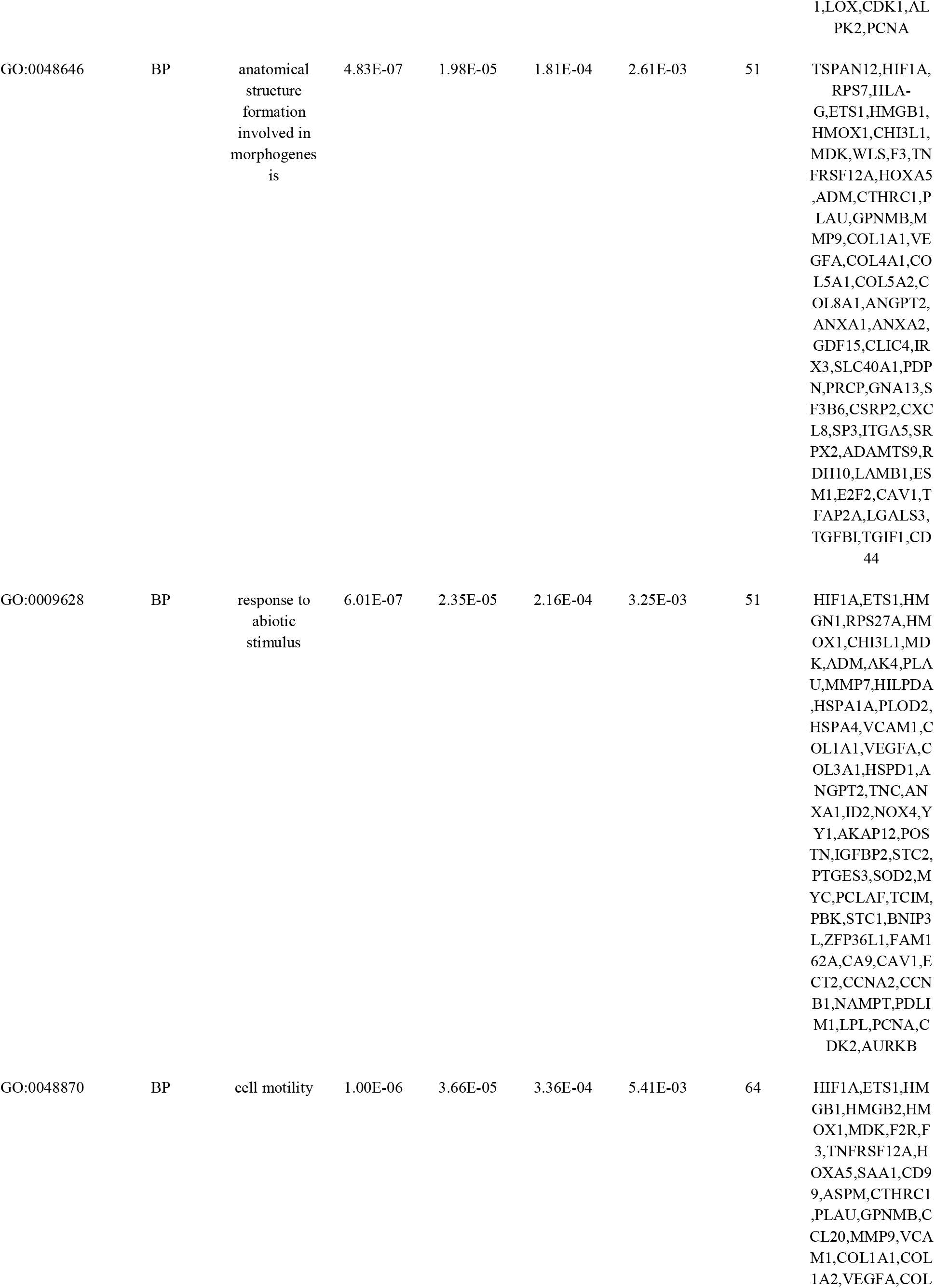
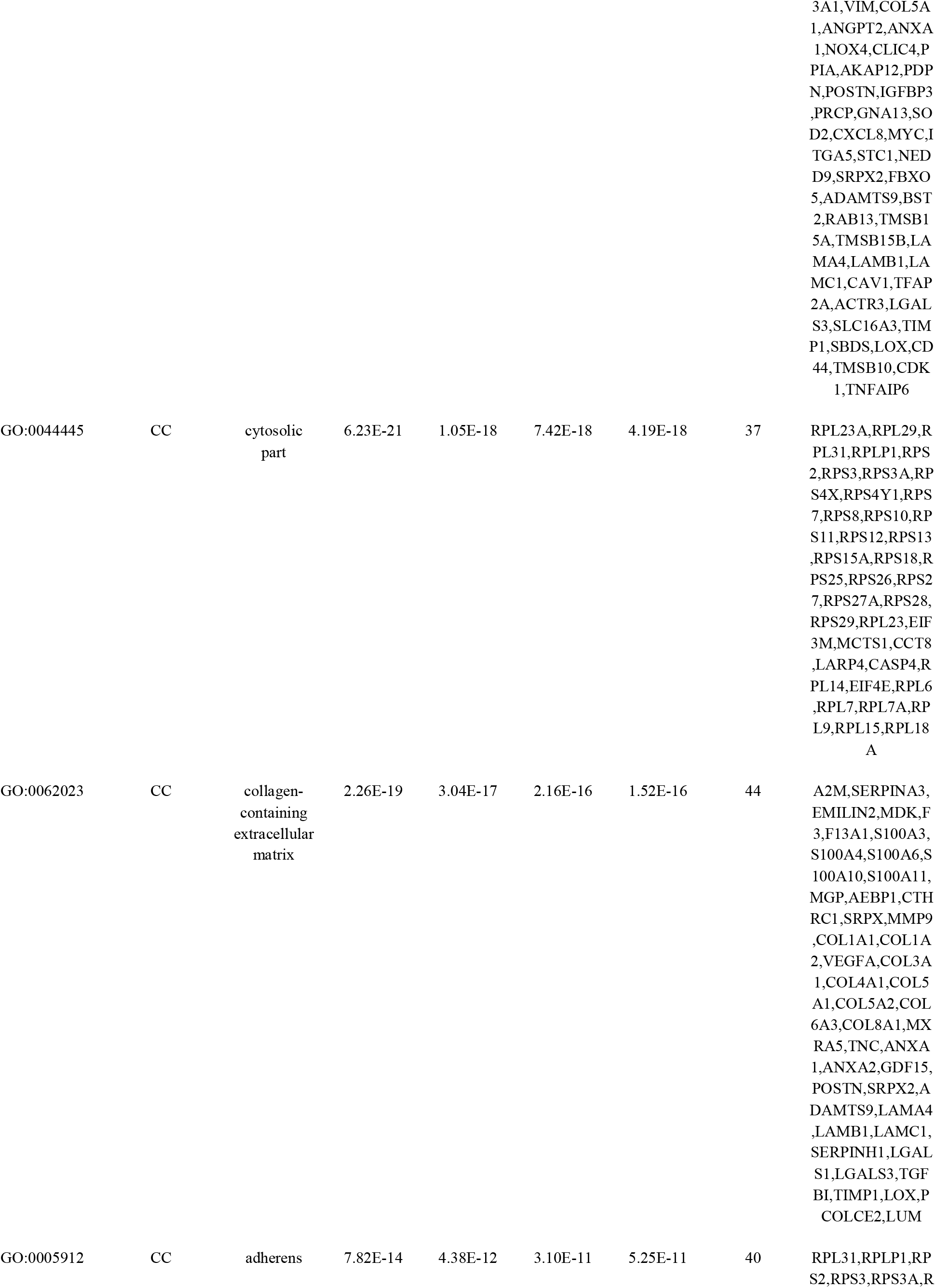
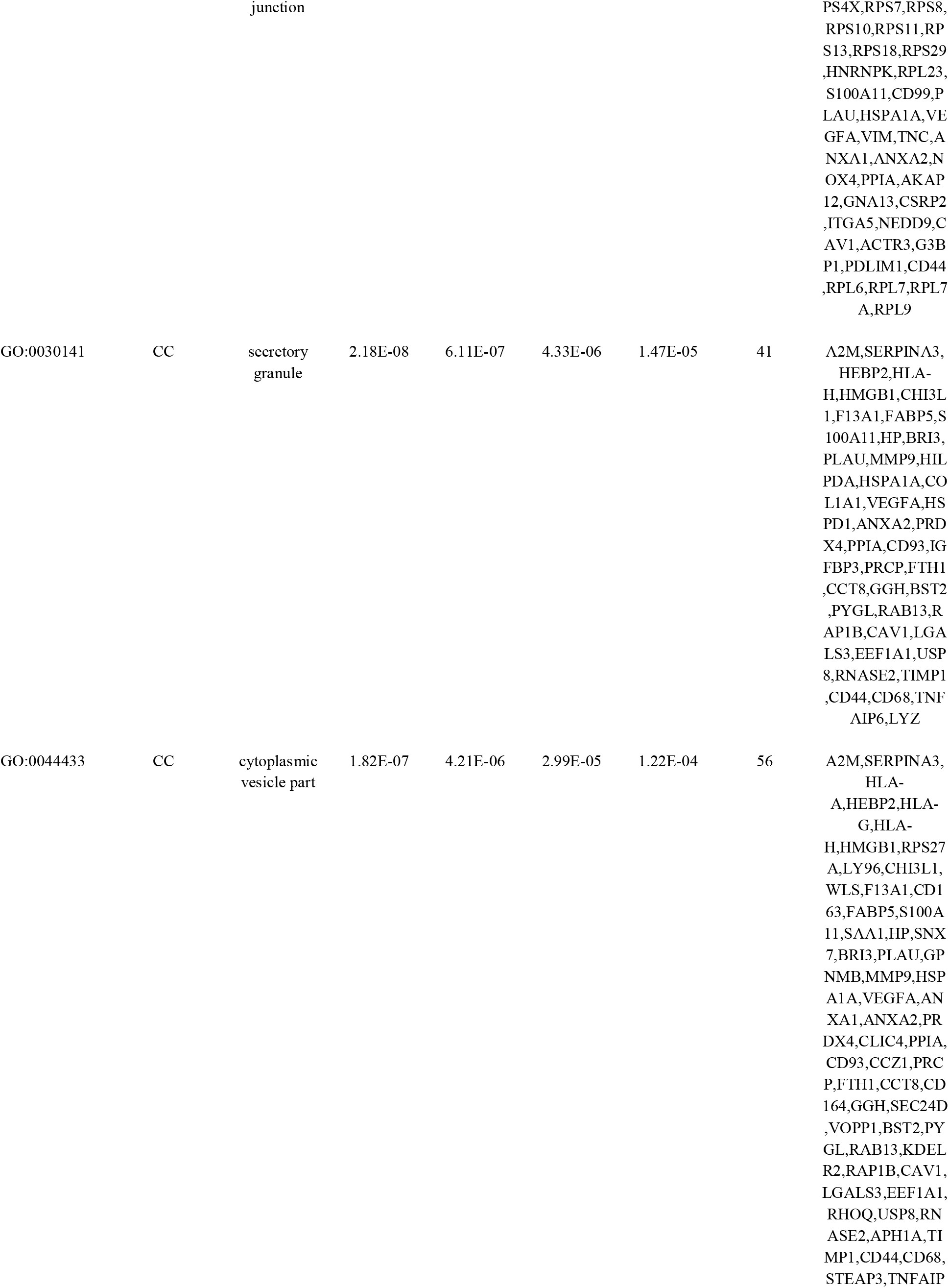
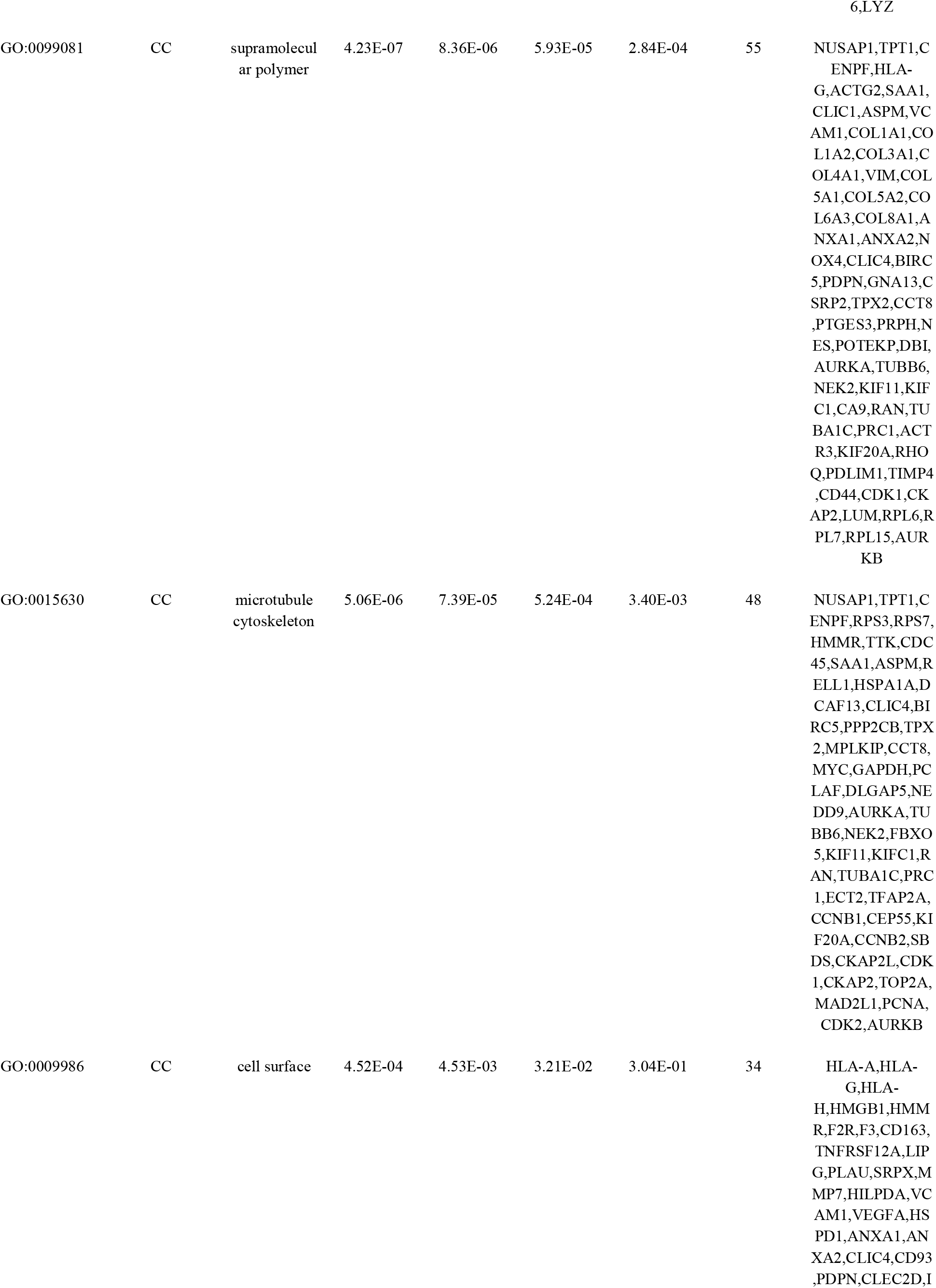
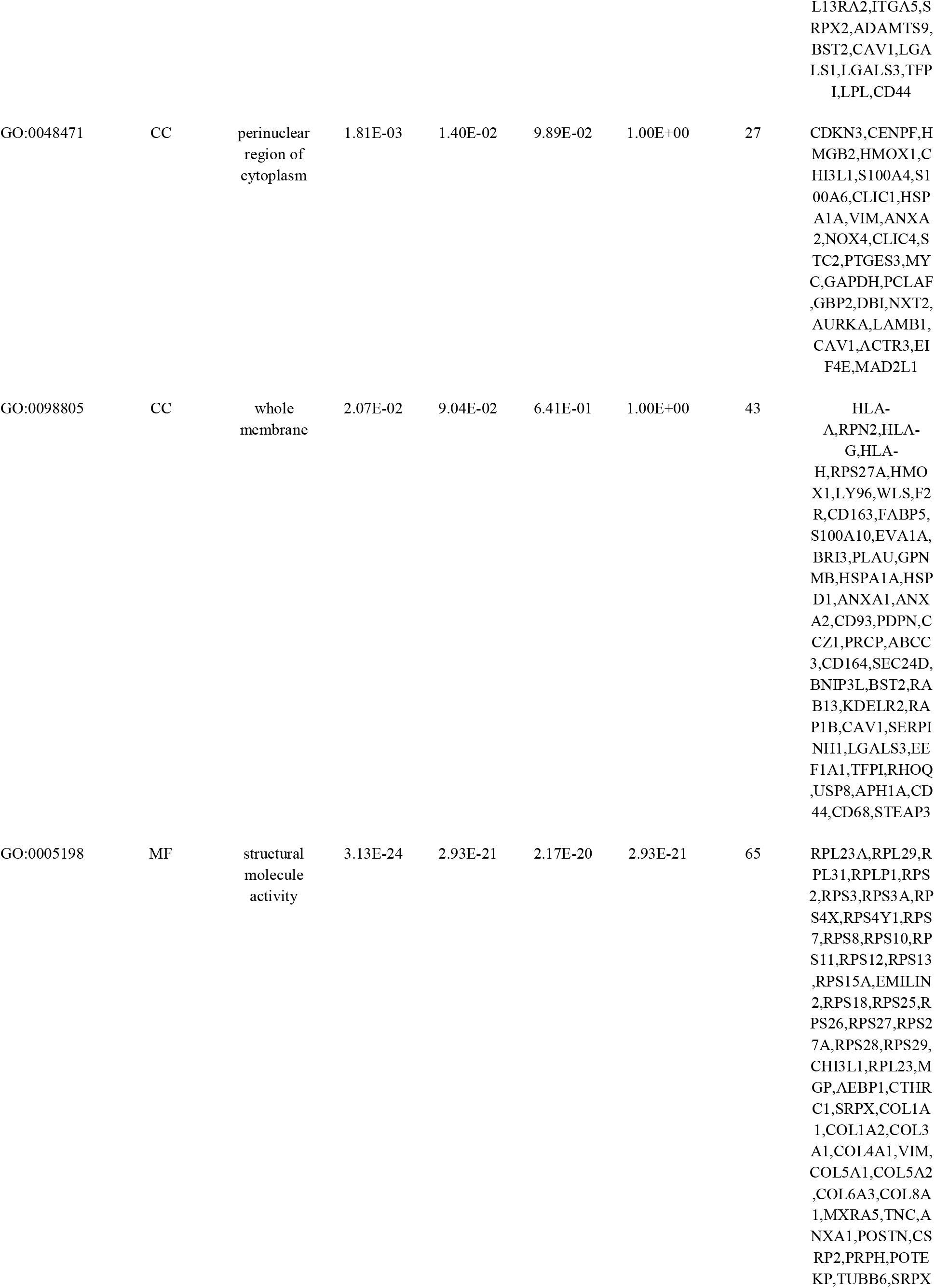
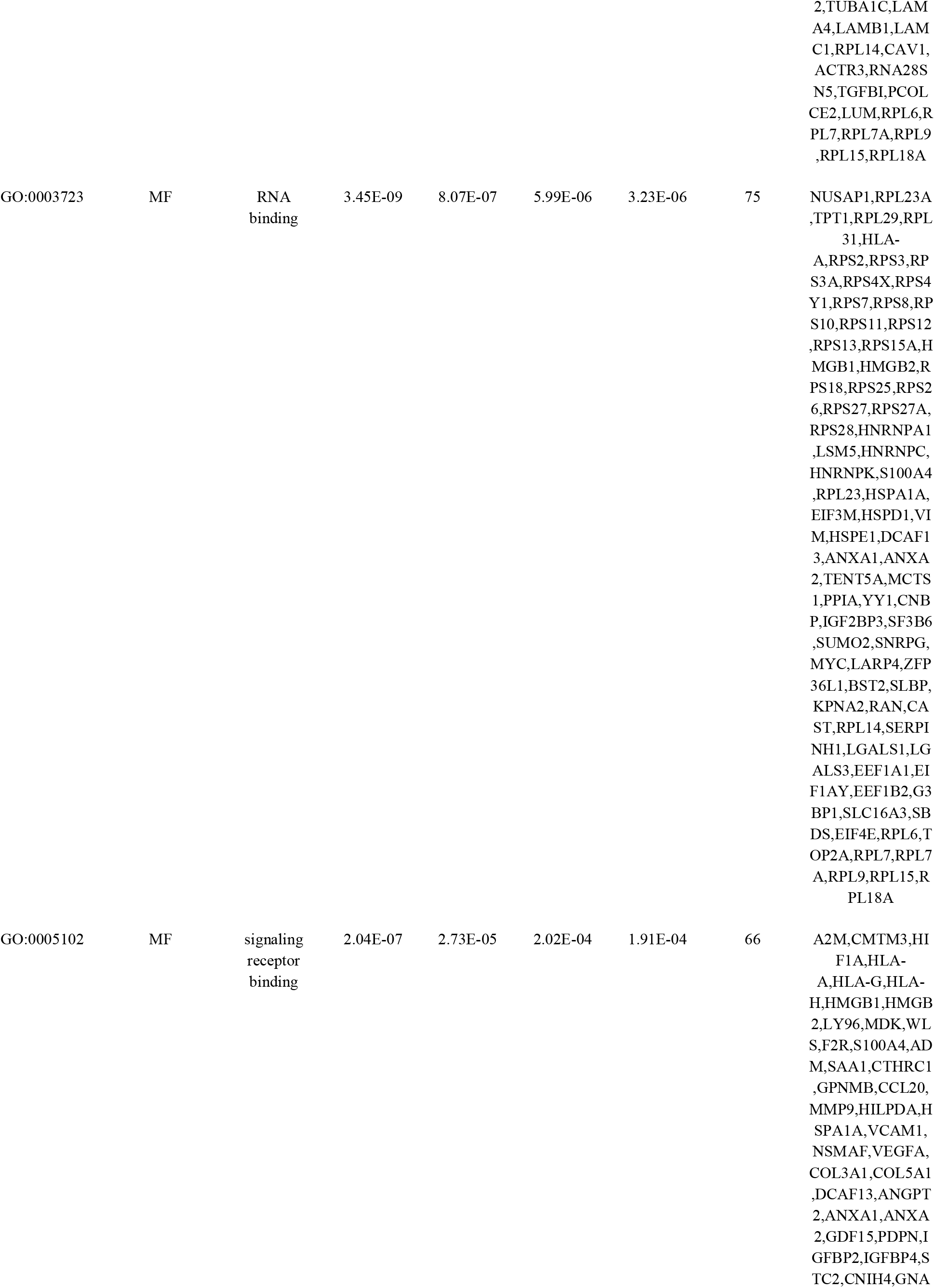
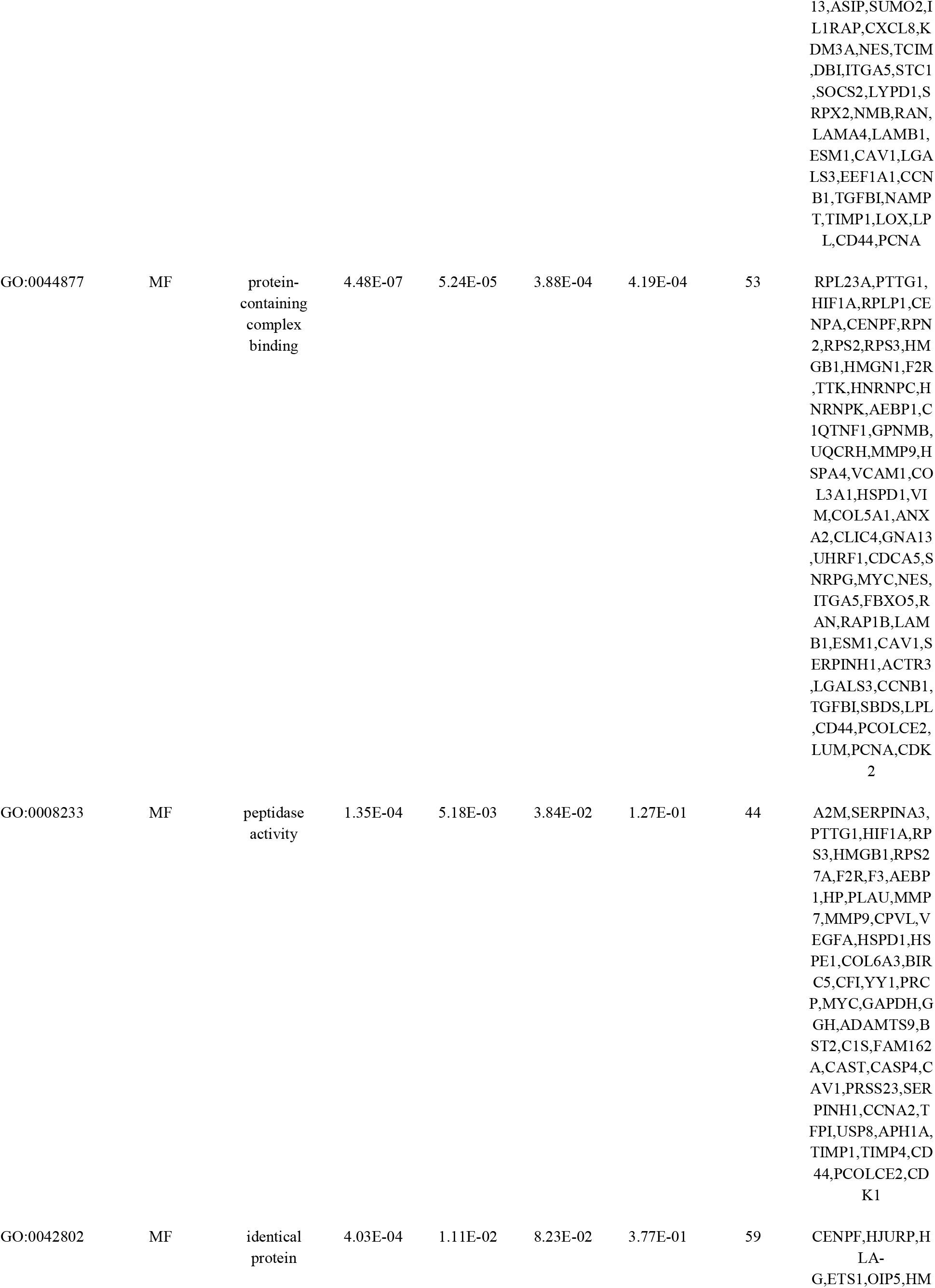
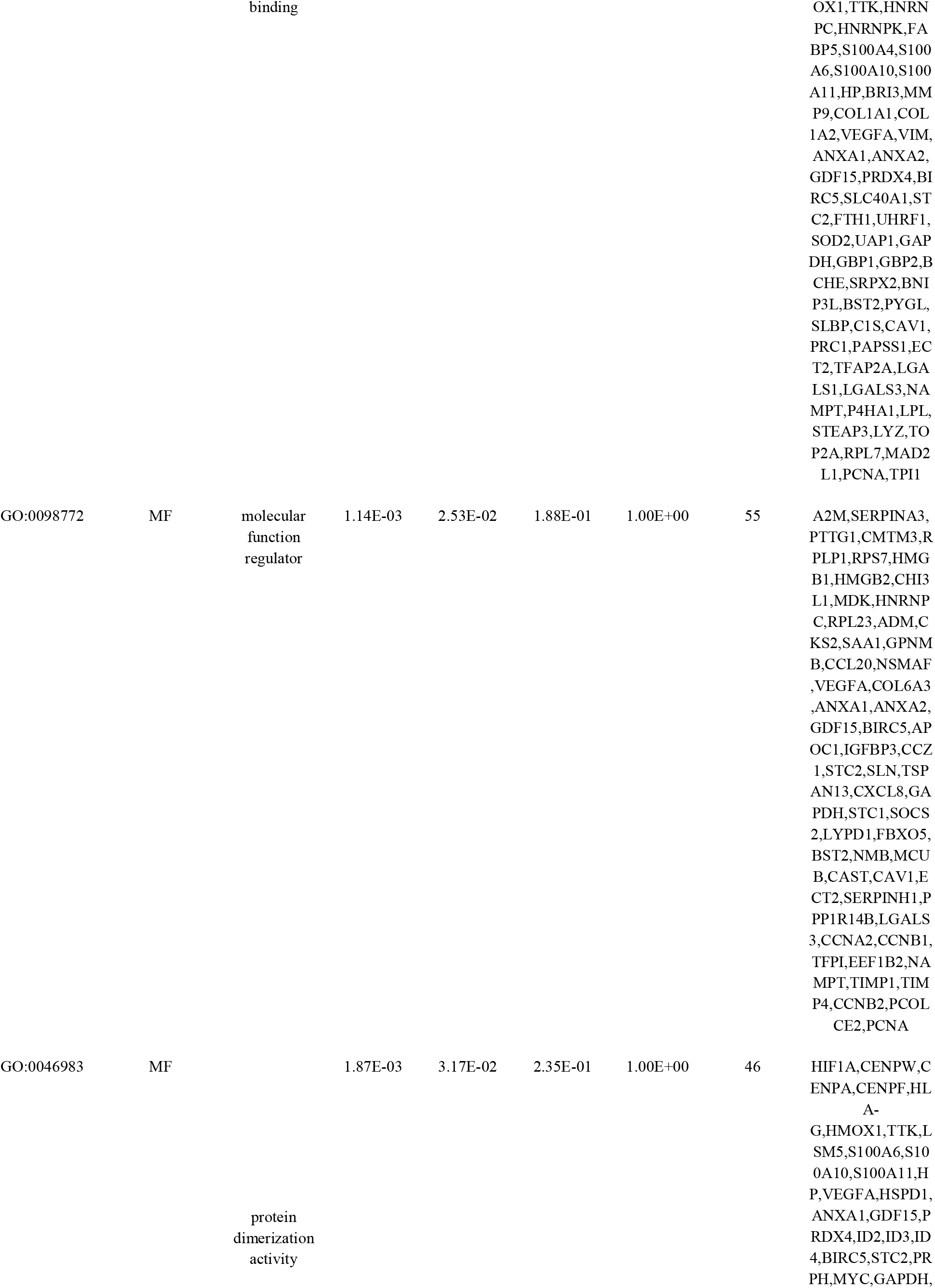
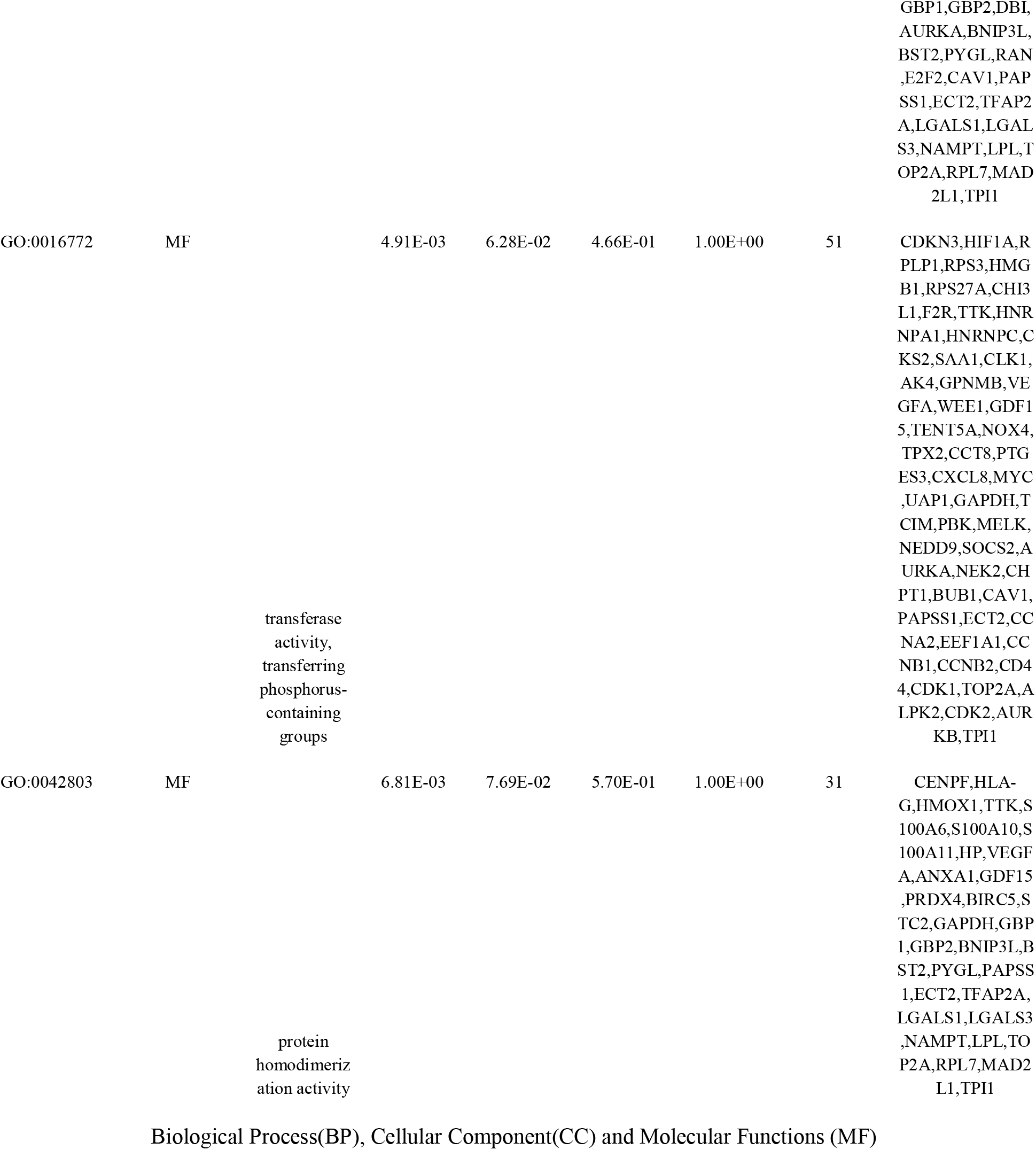
The enriched GO terms of the up regulated differentially expressed genes

**Table 5.**
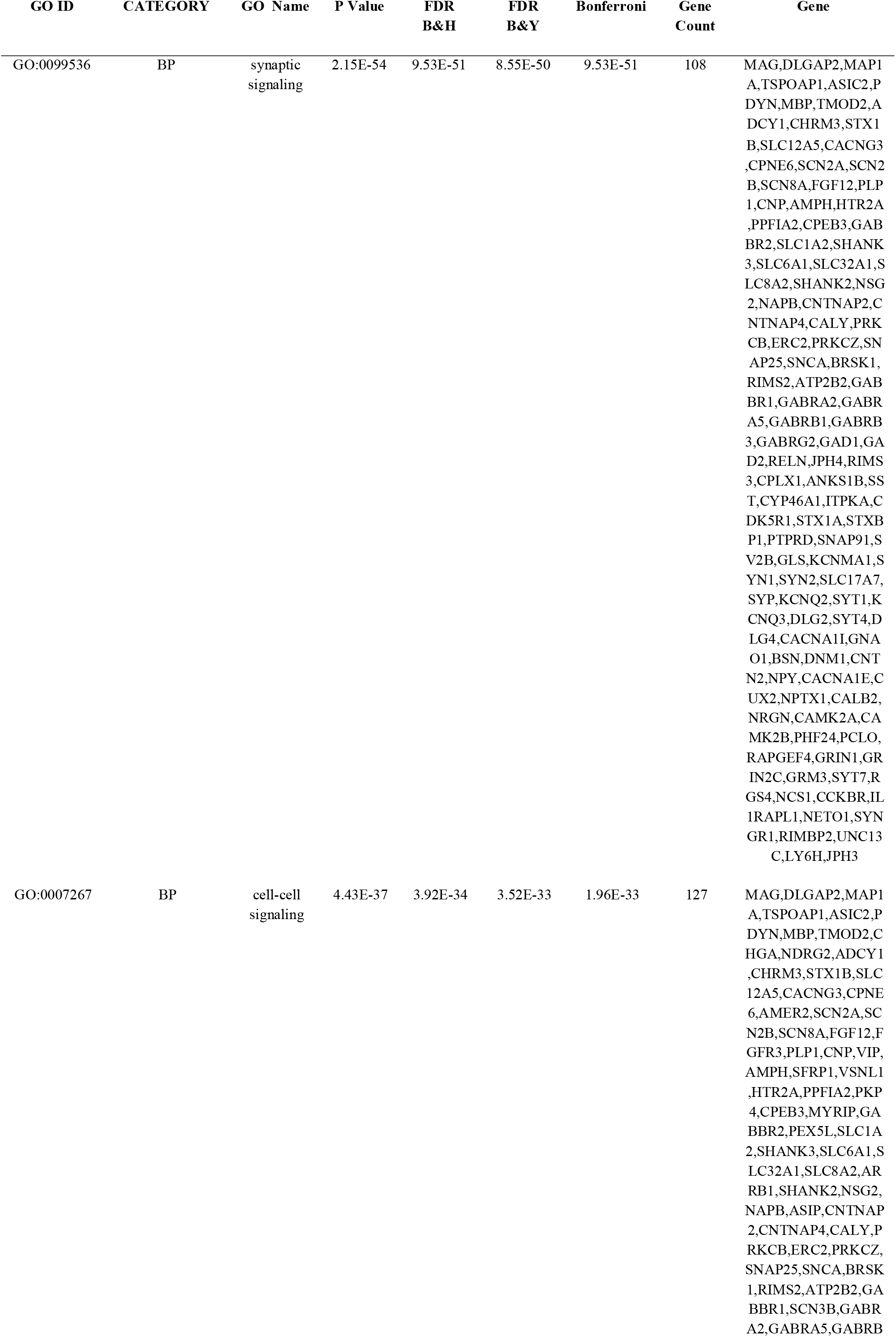
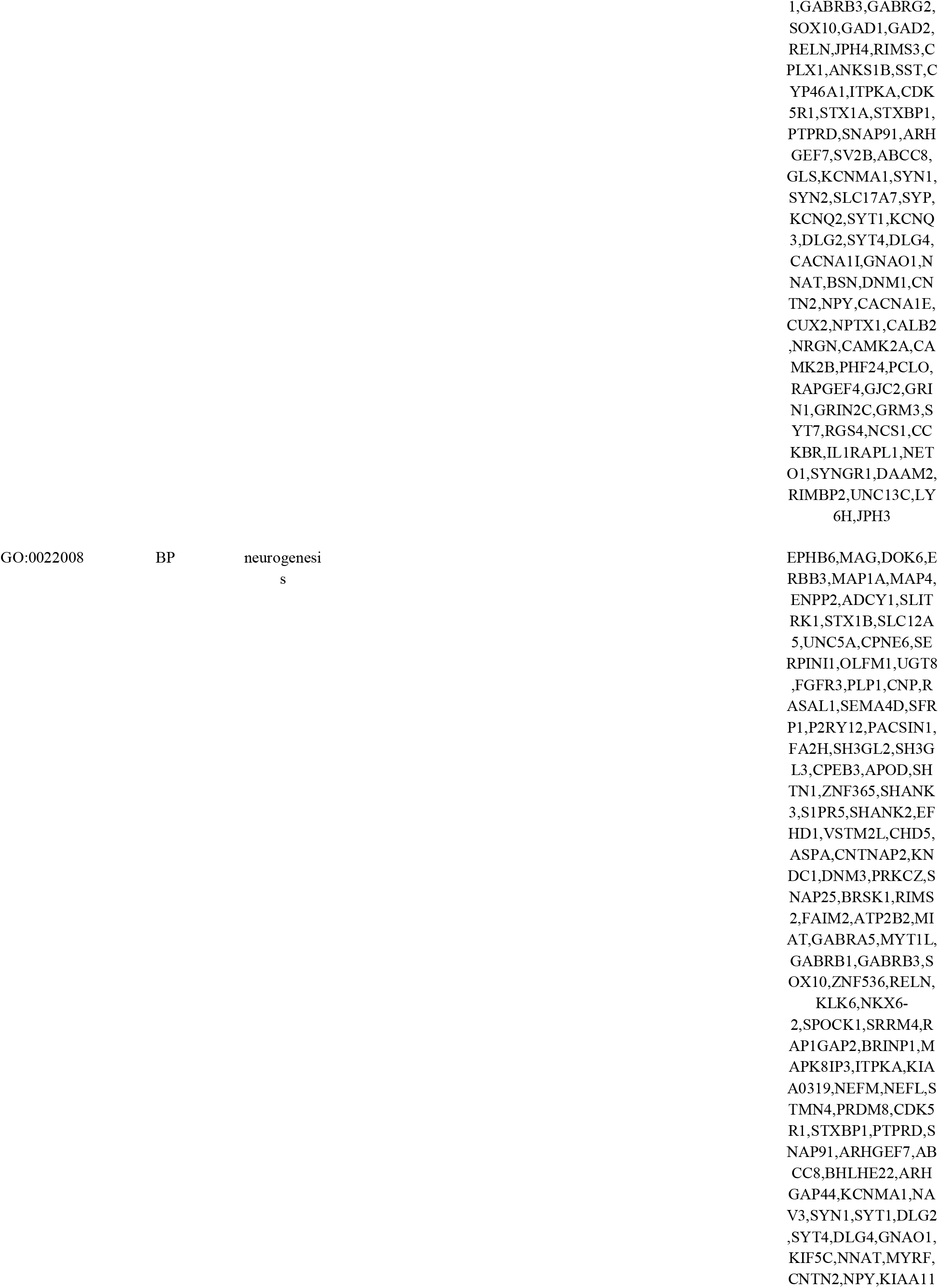
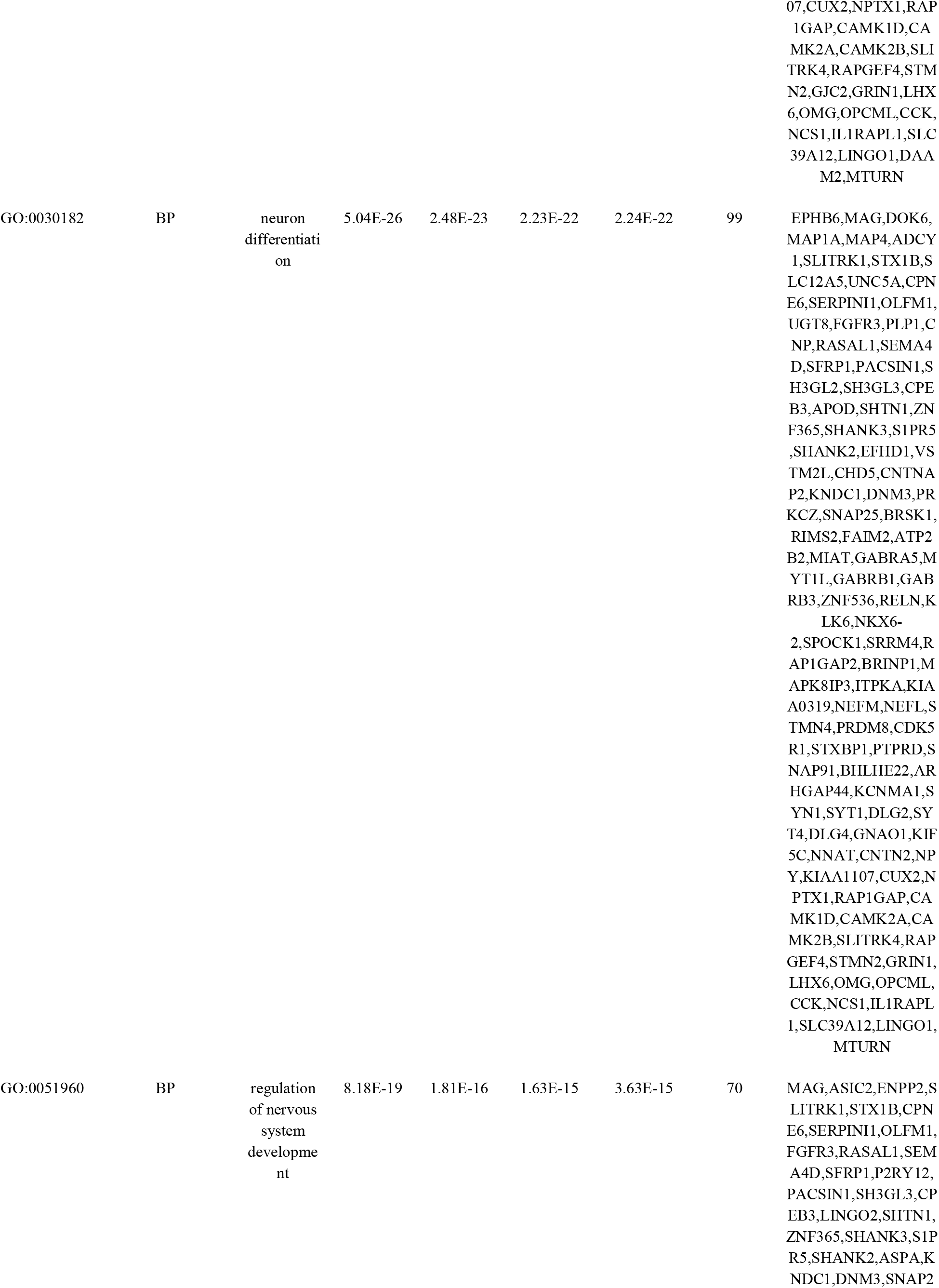
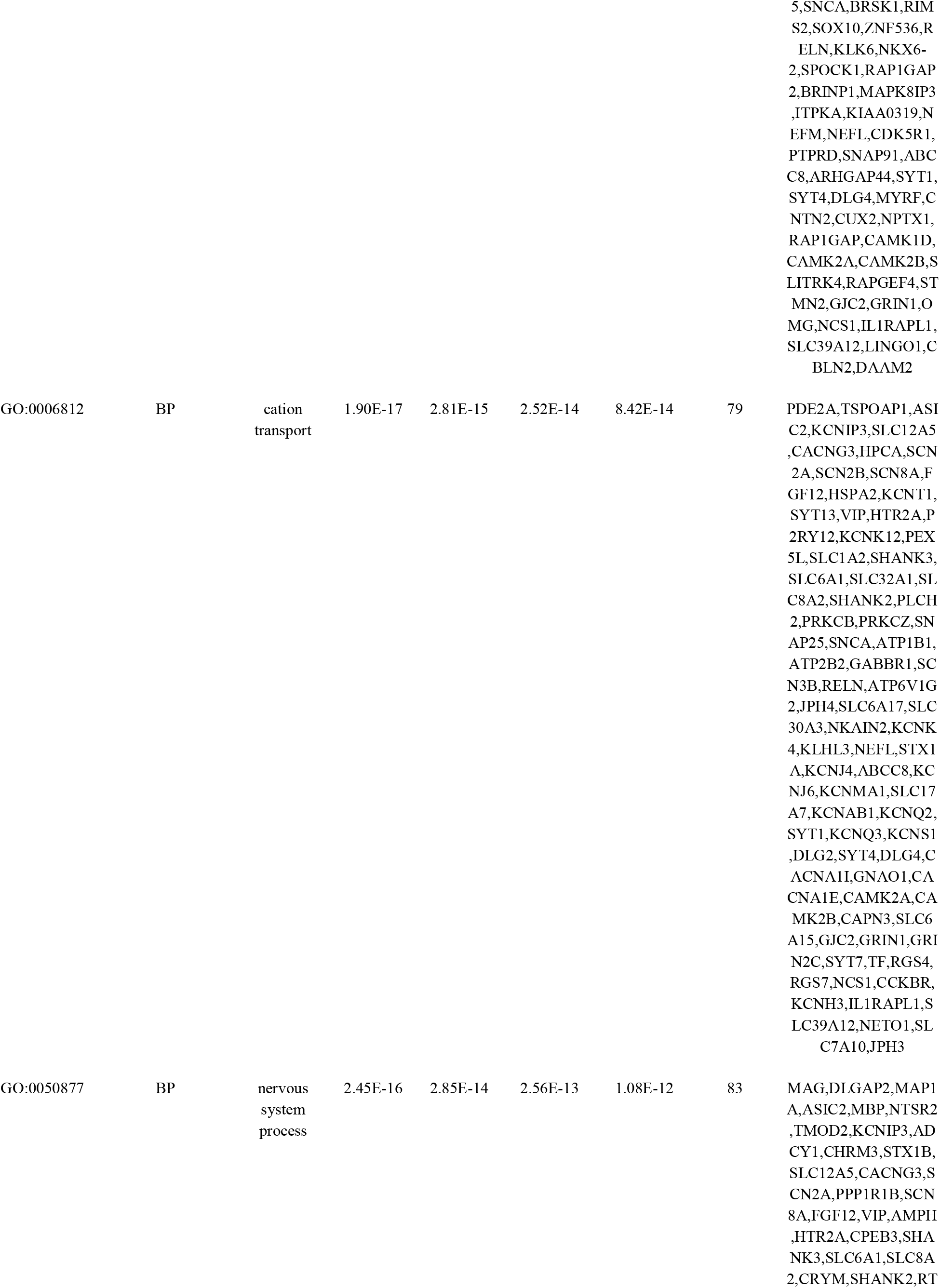
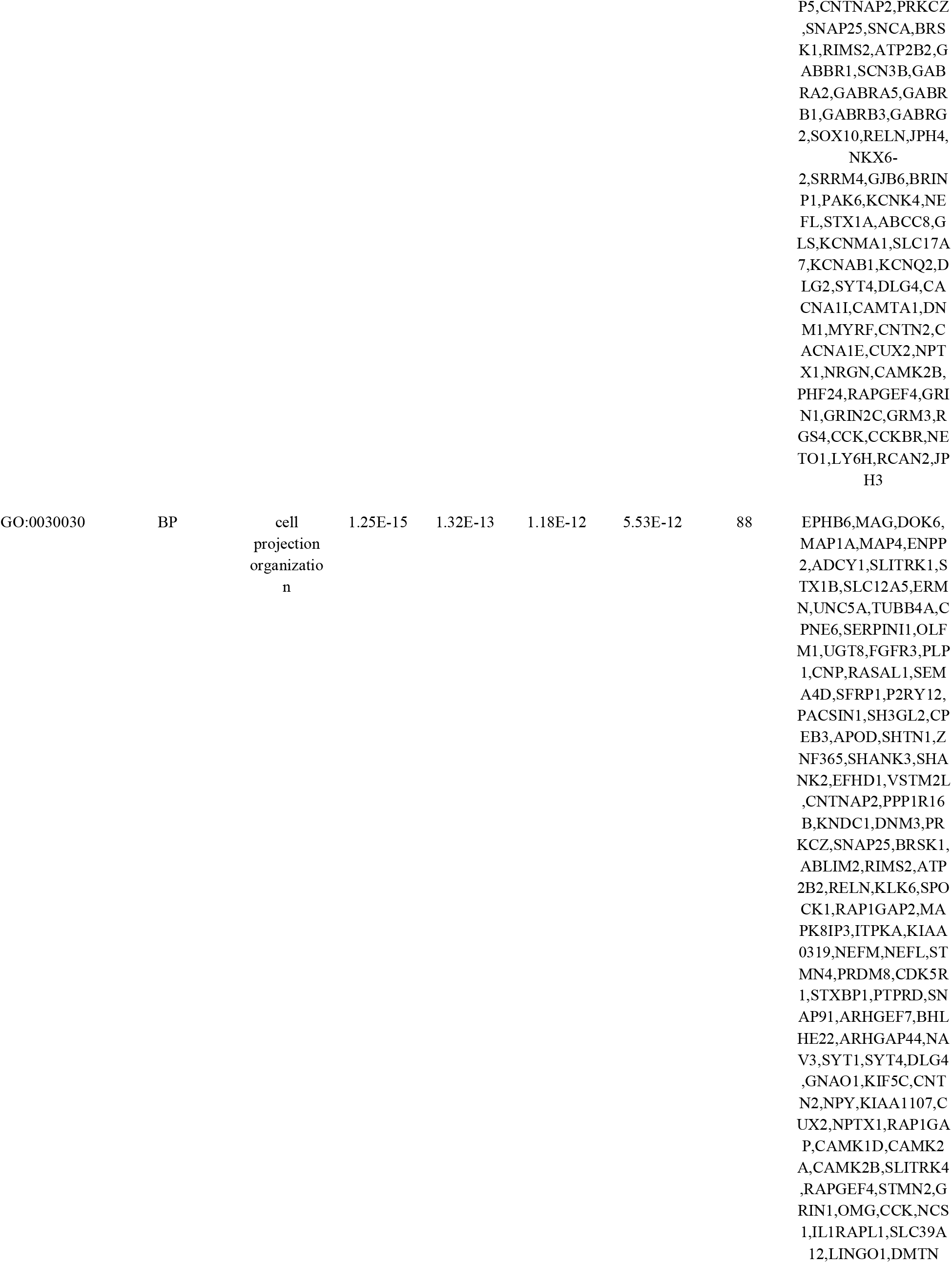
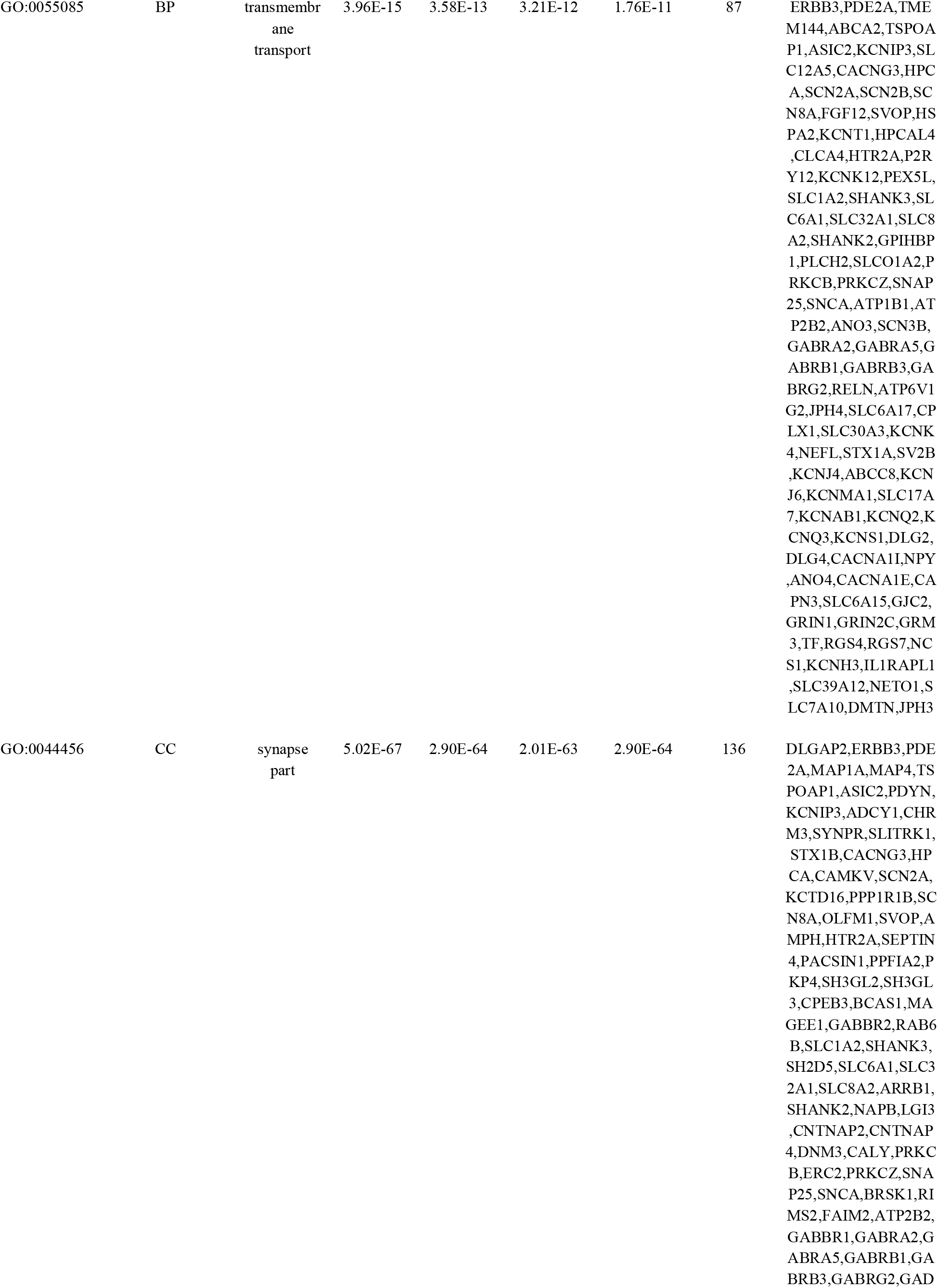
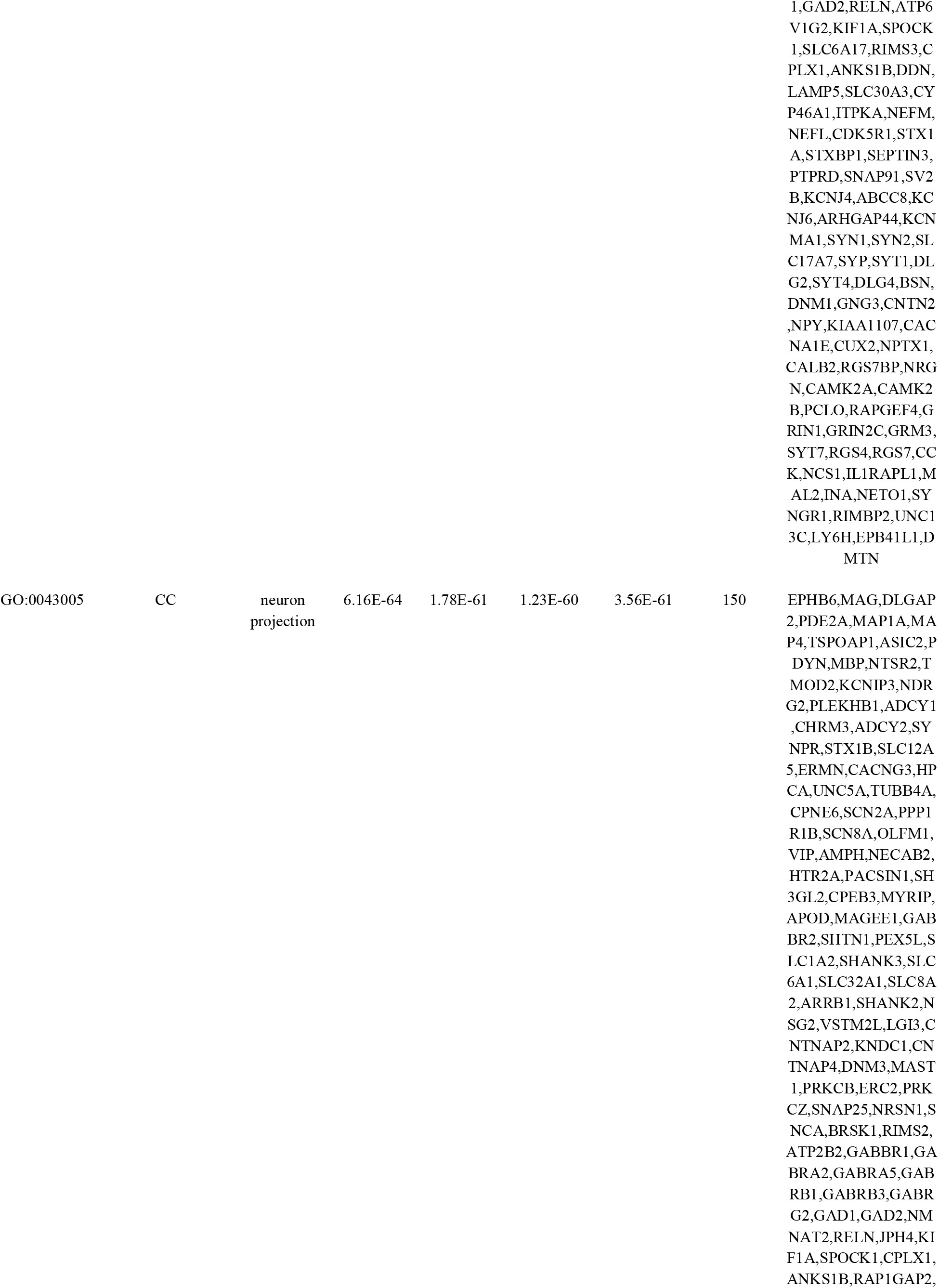
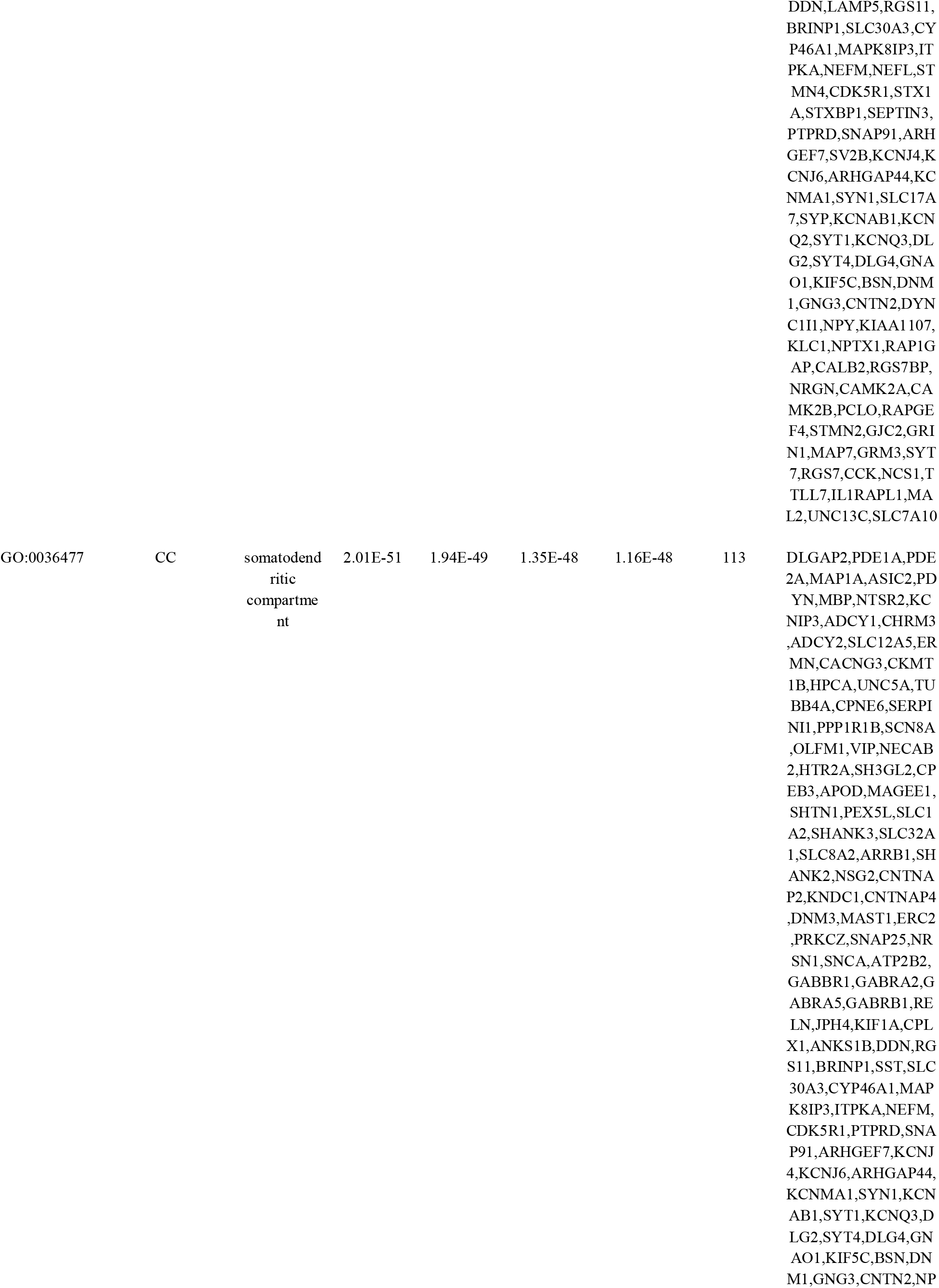
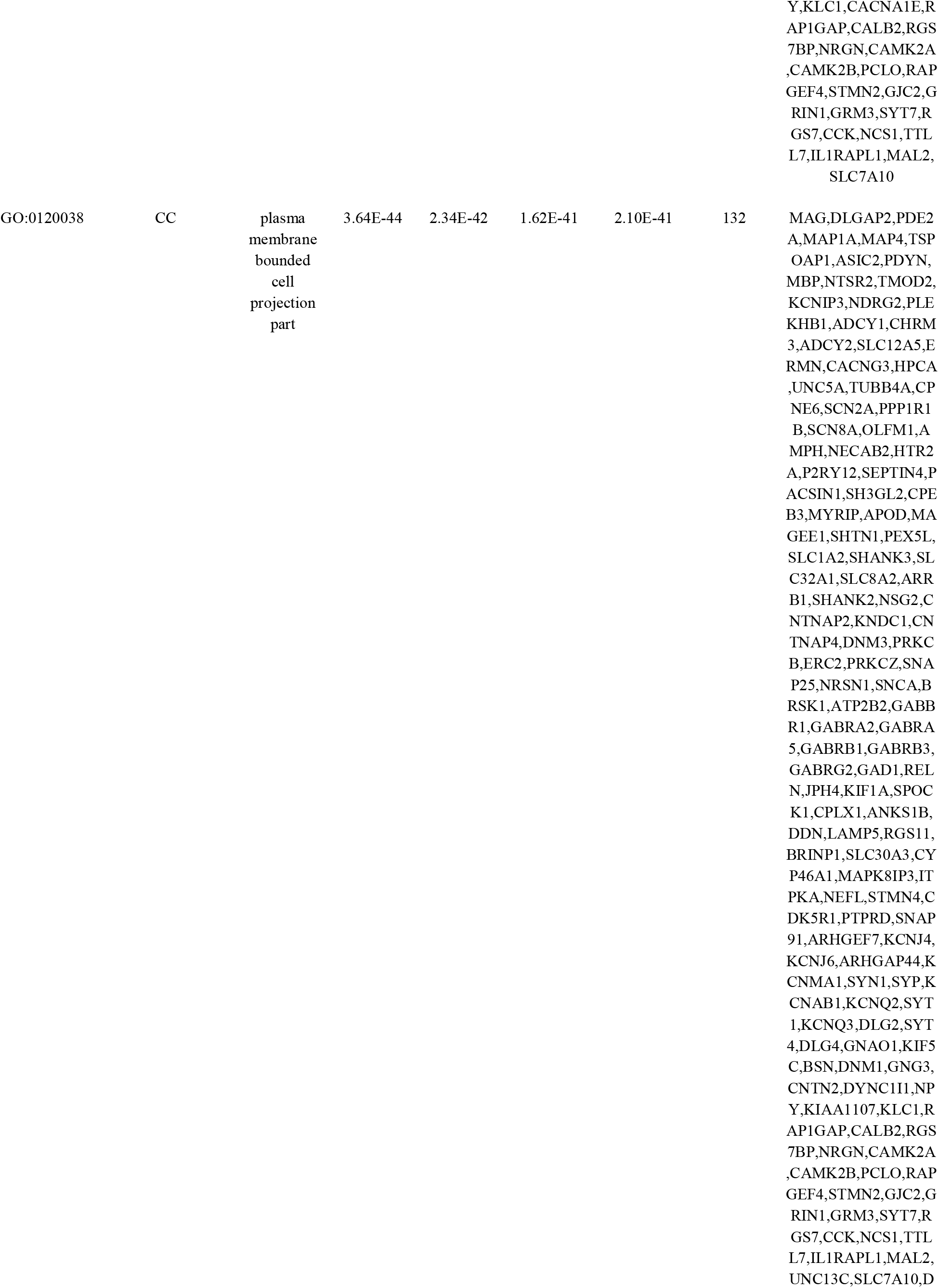
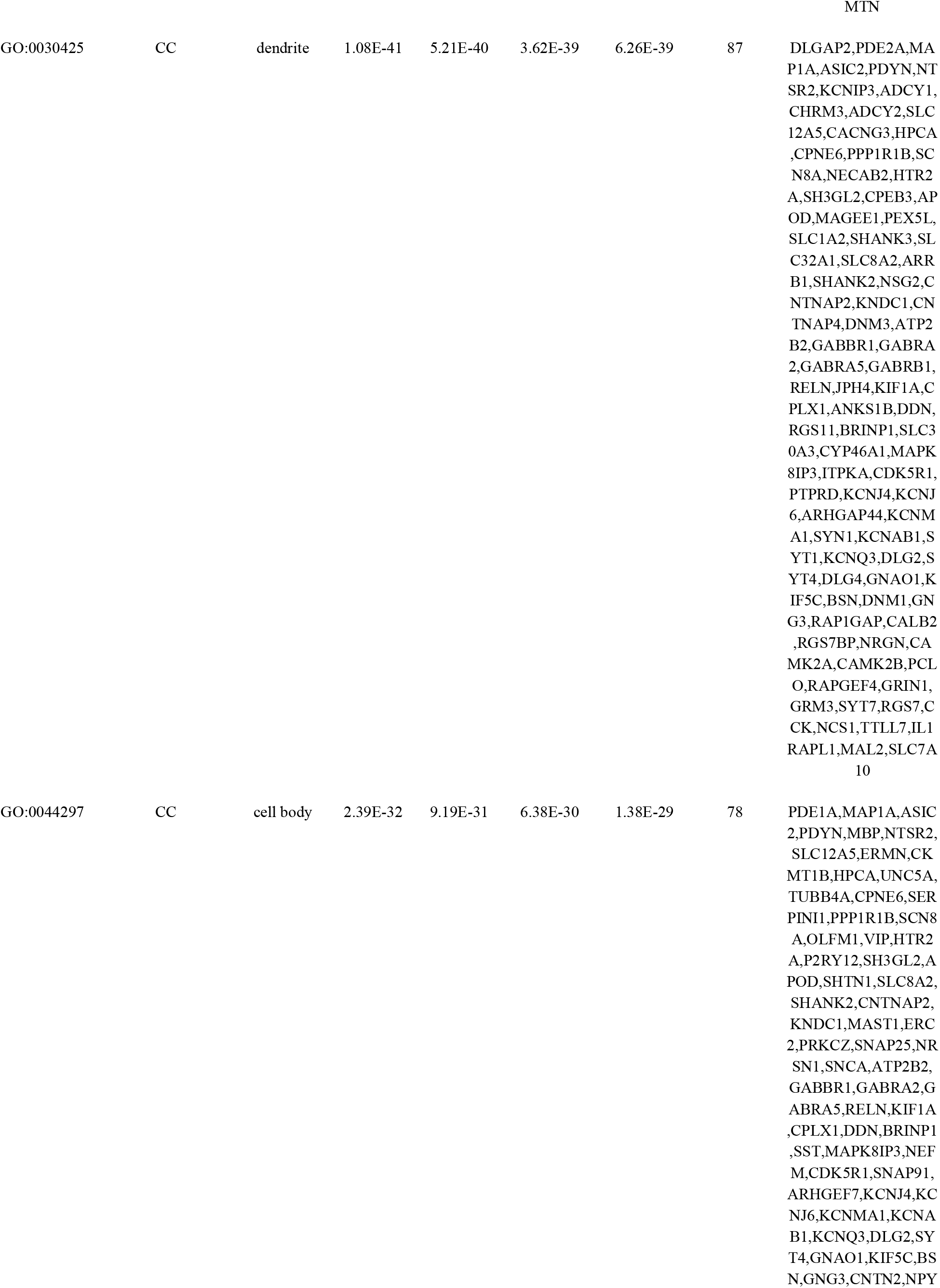
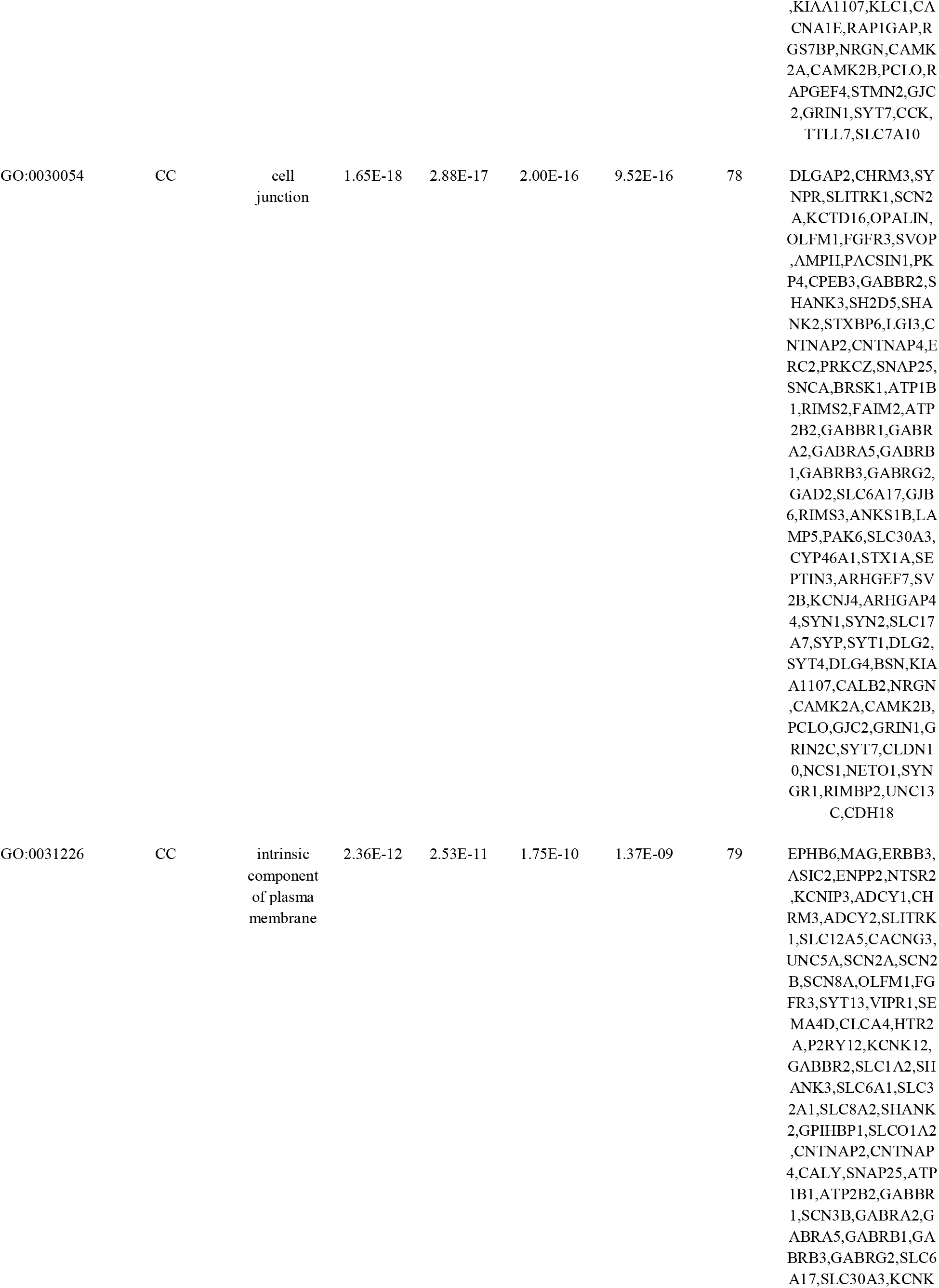
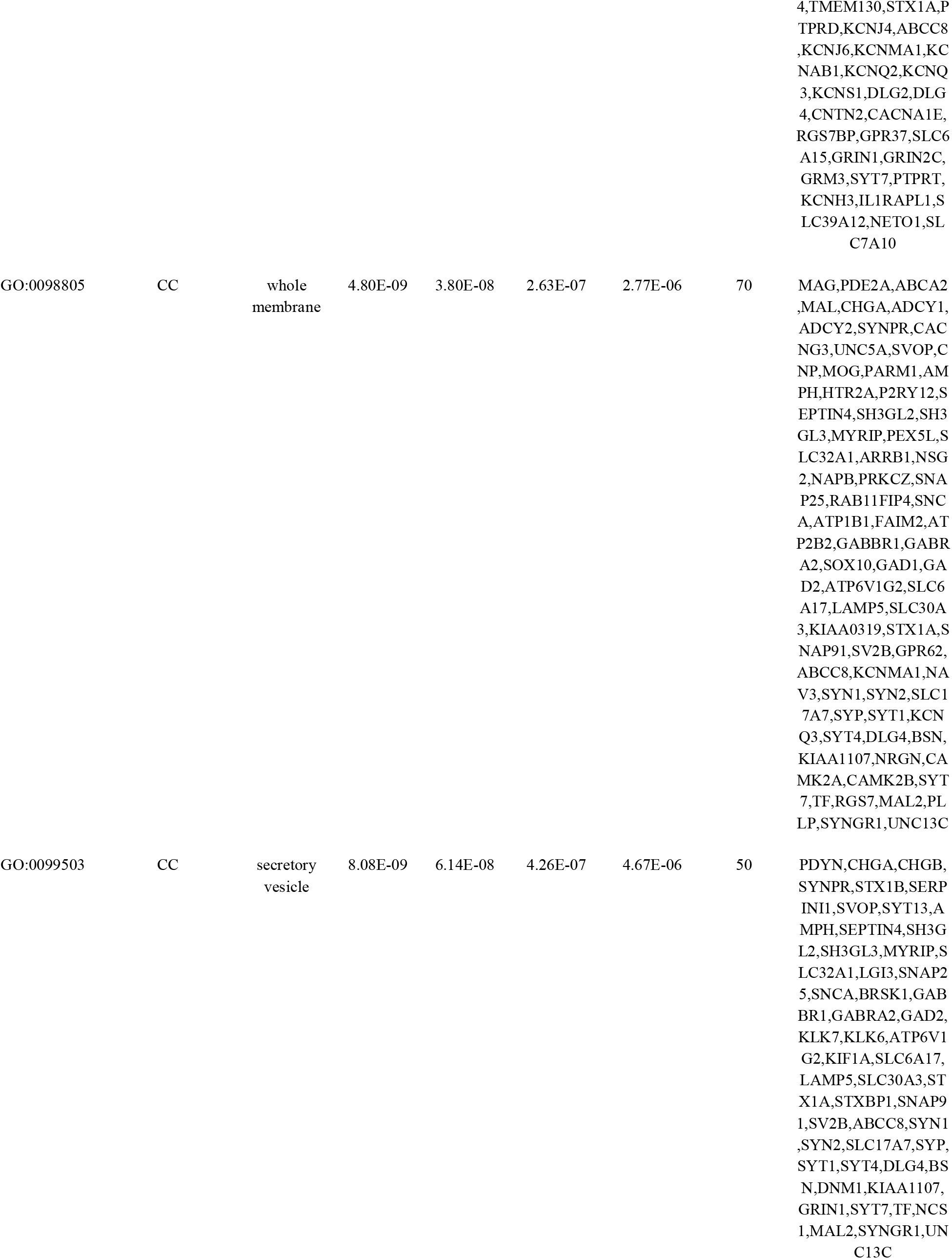
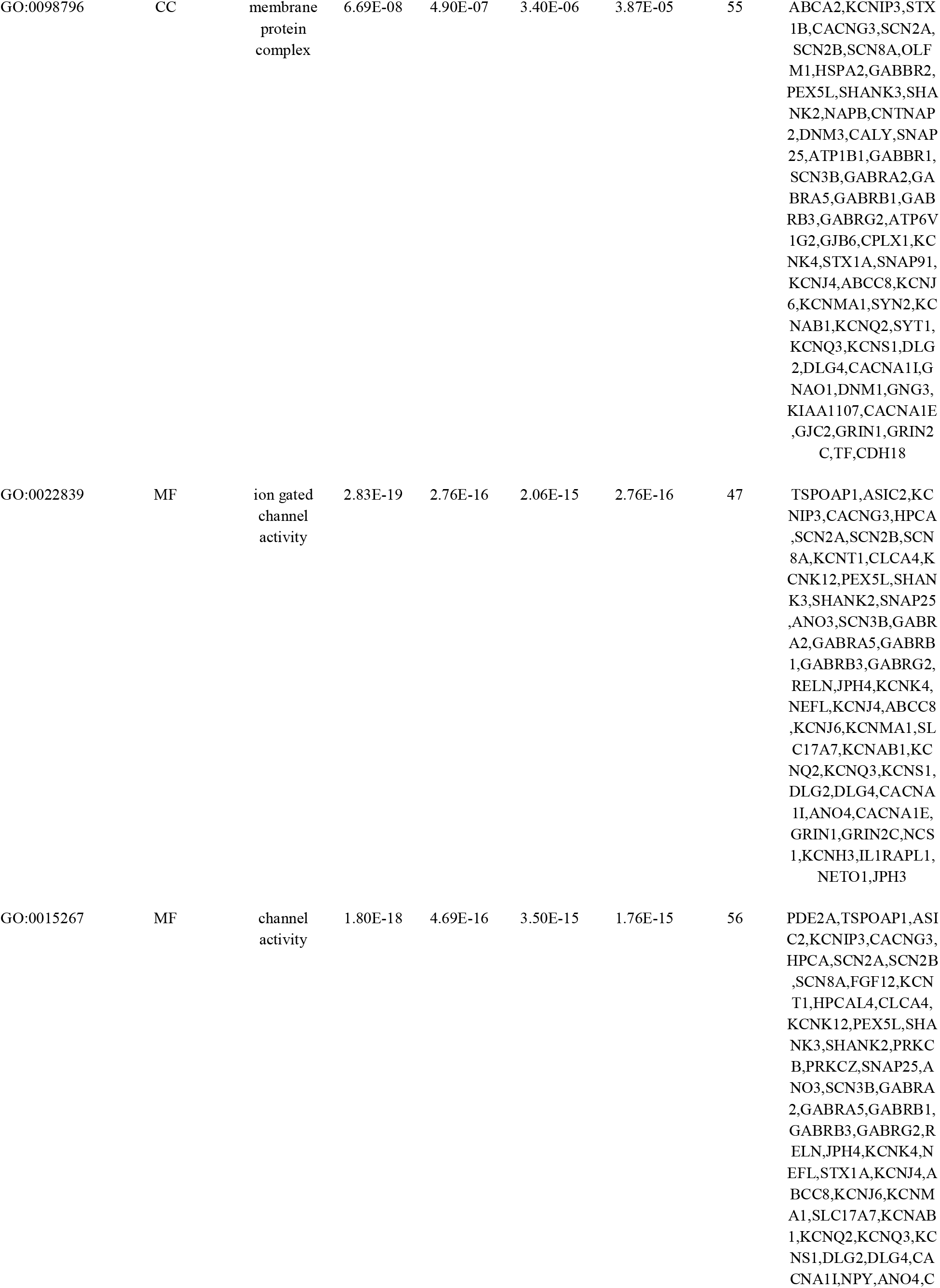
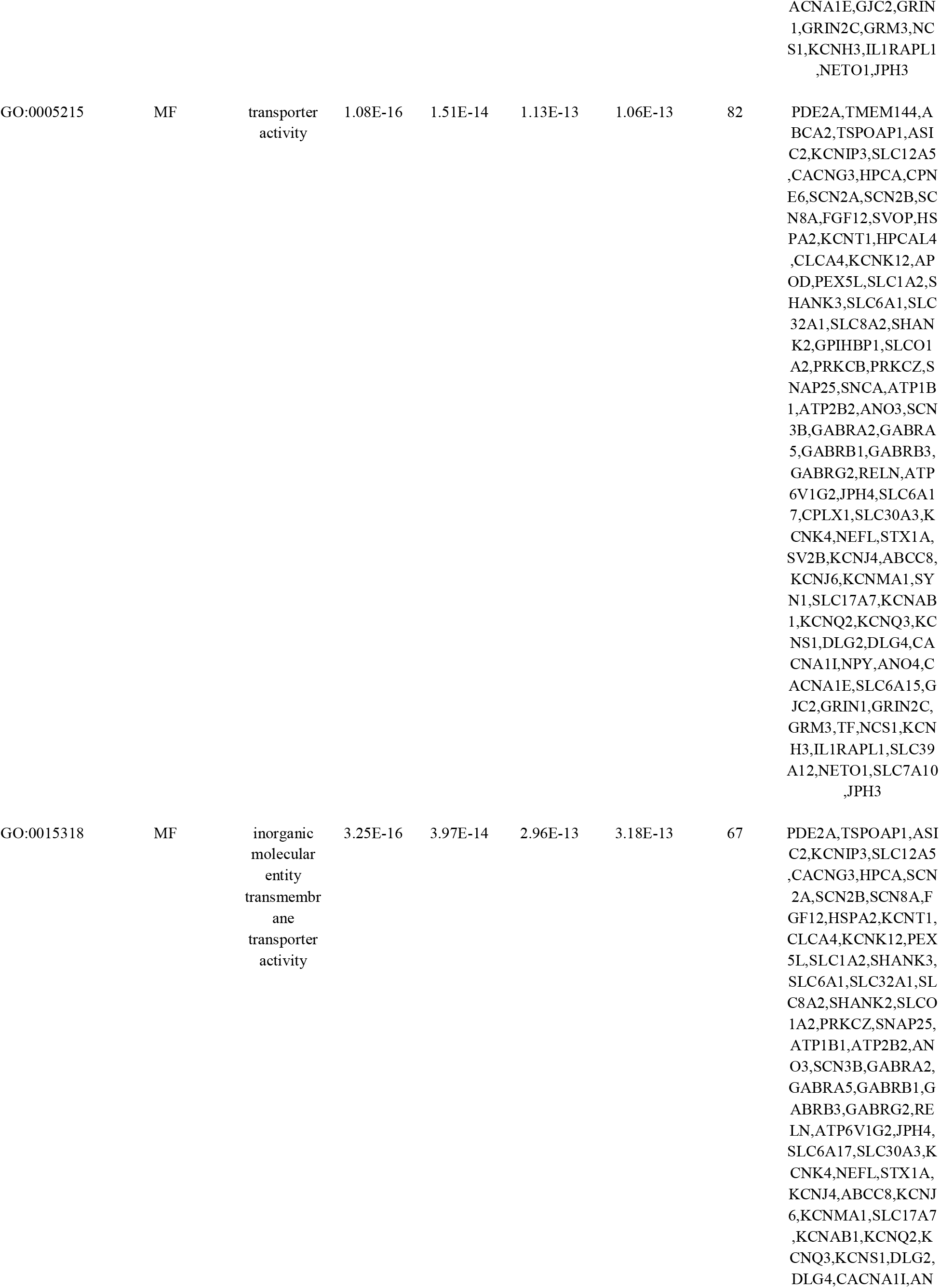
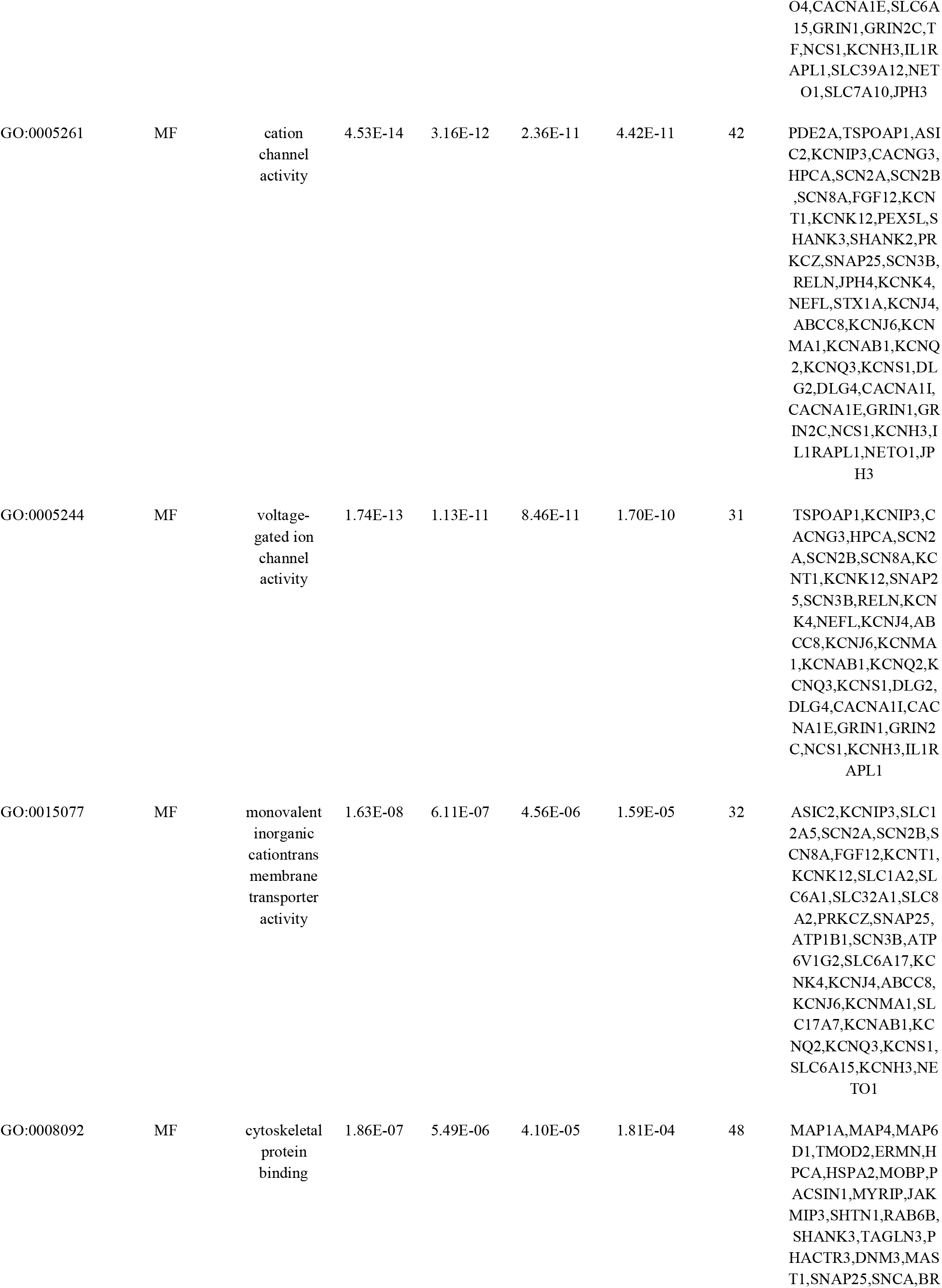
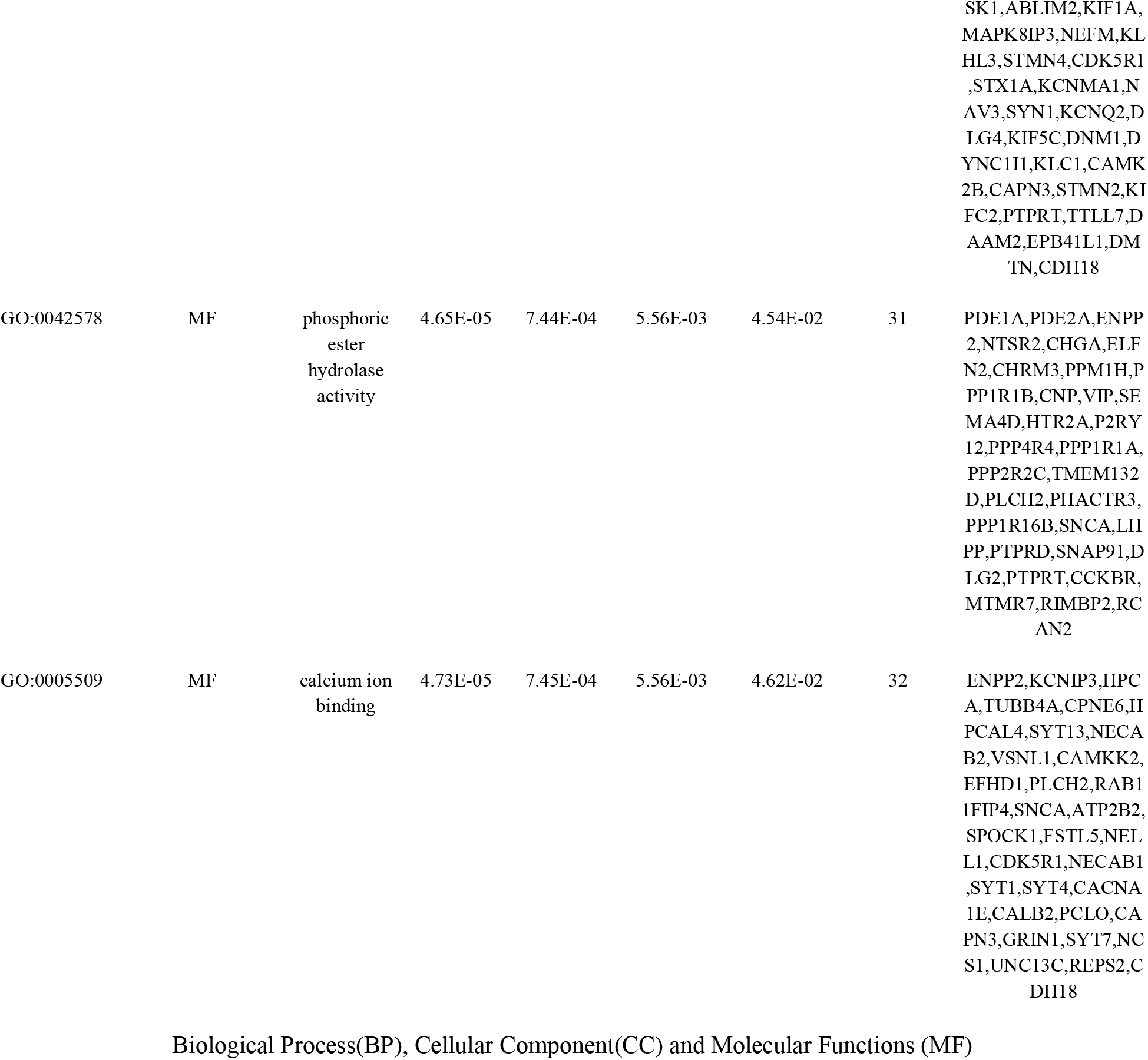
The enriched GO terms of the down-regulated differentially expressed genes

### PPI network construction and module analysis

There were 4162 nodes and 8776 relation pairs in PPI network for up regulated genes (Fig. 6). Hub genes in the network were analyzed, and the top nodes with the highest degree, betweenness centrality, stress centrality, closeness centrality score and lowest clustering coefficient score were MYC, VCAM1, CDK2, HNRNPA1, PCNA, CDK1, EEF1A1, HSPD1, HNRNPK, CEP55, A2M, CDCA5, ETS1 and PTGES3 are listed Table 6. The statistical results and scatter plot for node degree distribution, betweenness centrality, stress centrality, closeness centrality and clustring coefficient are shown in Fig. 7. These hub genes were enriched in cell cycle, TNF signaling pathway, FOXM1 transcription factor network, processing of capped intron-containing pre-mRNA, macromolecule catabolic process, mitotic cell cycle, regulation of cell death, validated targets of C-MYC transcriptional activation, metabolism of proteins, microtubule cytoskeleton, complement and coagulation cascades, protein-containing complex binding, pathways in cancer and C20 prostanoid biosynthesis. Similarly, there were 2392 nodes and 3196 relation pairs in PPI network for down regulated genes (Fig. 8). Hub genes in the network were analyzed, and the top nodes with the highest degree score were ARRB1, SNCA, ERBB3, PRKCZ, DLG4, SLC30A3, DNM1, FAM153B, RAPGEF5, EFHD1, PDYN, ZNF536 and TSPOAP1 are listed Table 6. The statistical results and scatter plot for node degree distribution, betweenness centrality, stress centrality, closeness centrality and clustring coefficient are shown in Fig. 9. These hub genes were enriched in endocytosis, Parkinson’s disease, calcium signaling pathway, synaptic signaling, glutamatergic synapse, transmembrane transport of small molecules, synaptic vesicle cycle, neurogenesis, signaling by GPCR, neuron differentiation and neuronal system.

**Fig. 6.**
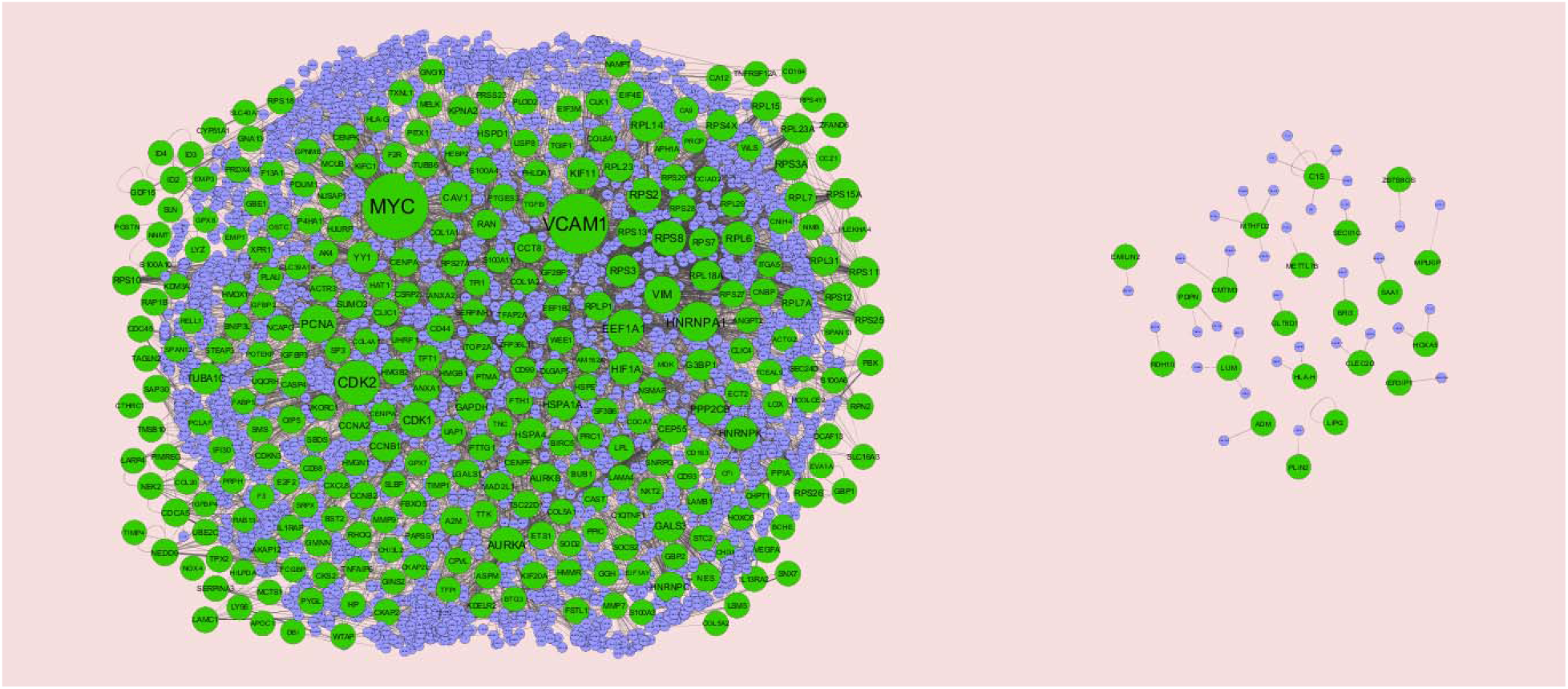
Protein–protein interaction network of differentially expressed genes (DEGs). Green nodes denotes up regulated genes.

**Fig. 7.**
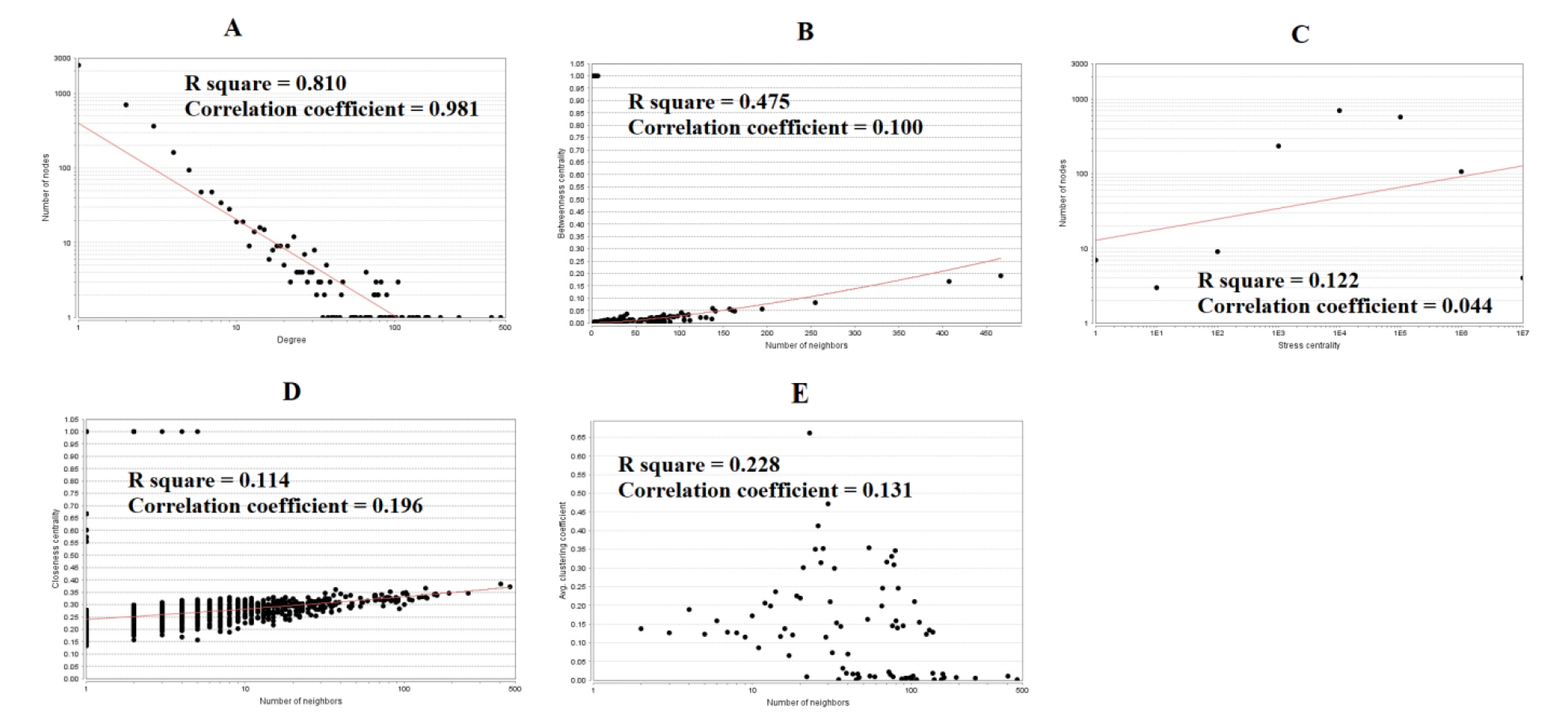
Scatter plot for up regulated genes. (A- Node degree; B- Betweenness centrality; C- Stress centrality; D- Closeness centrality; E- Clustering coefficient)

**Fig. 8.**
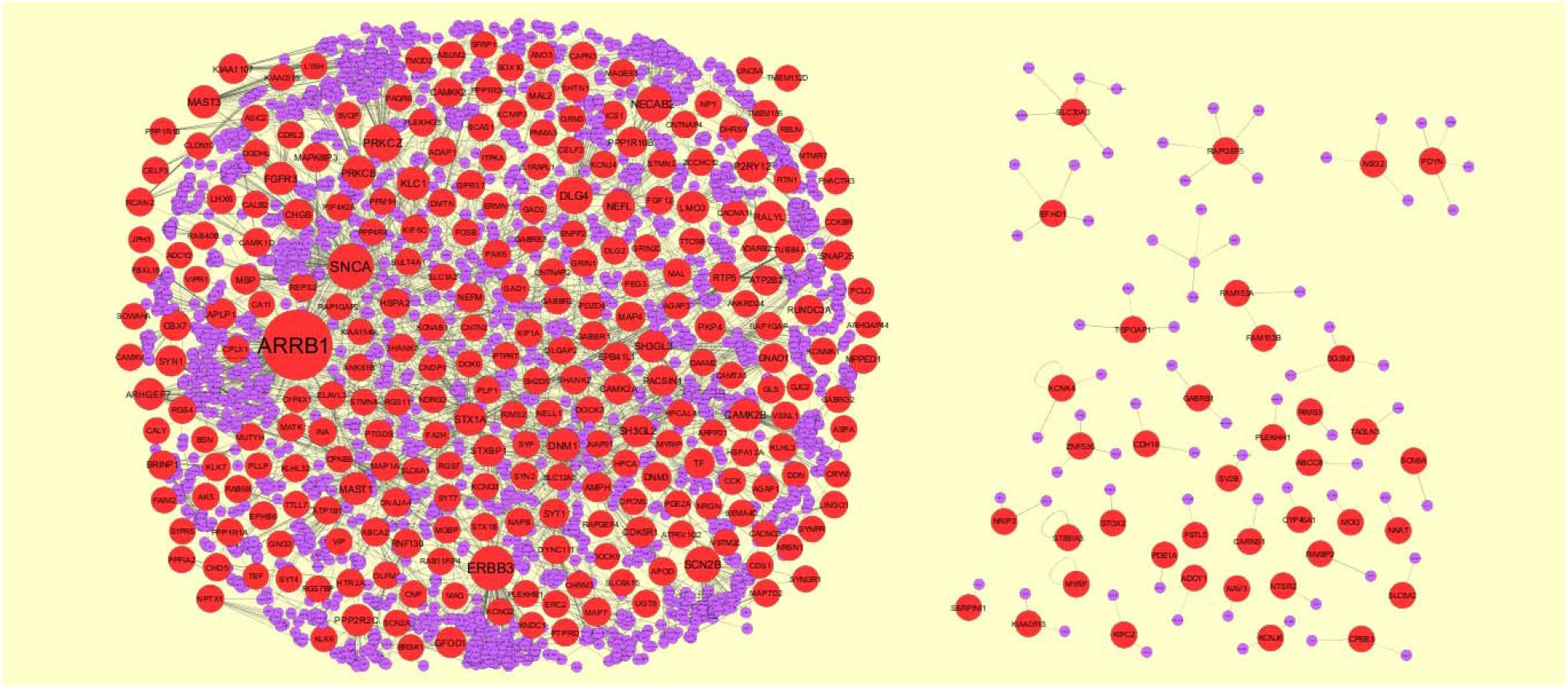
Protein–protein interaction network of differentially expressed genes (DEGs). Red nodes denotes down regulated genes.

**Fig. 9.**
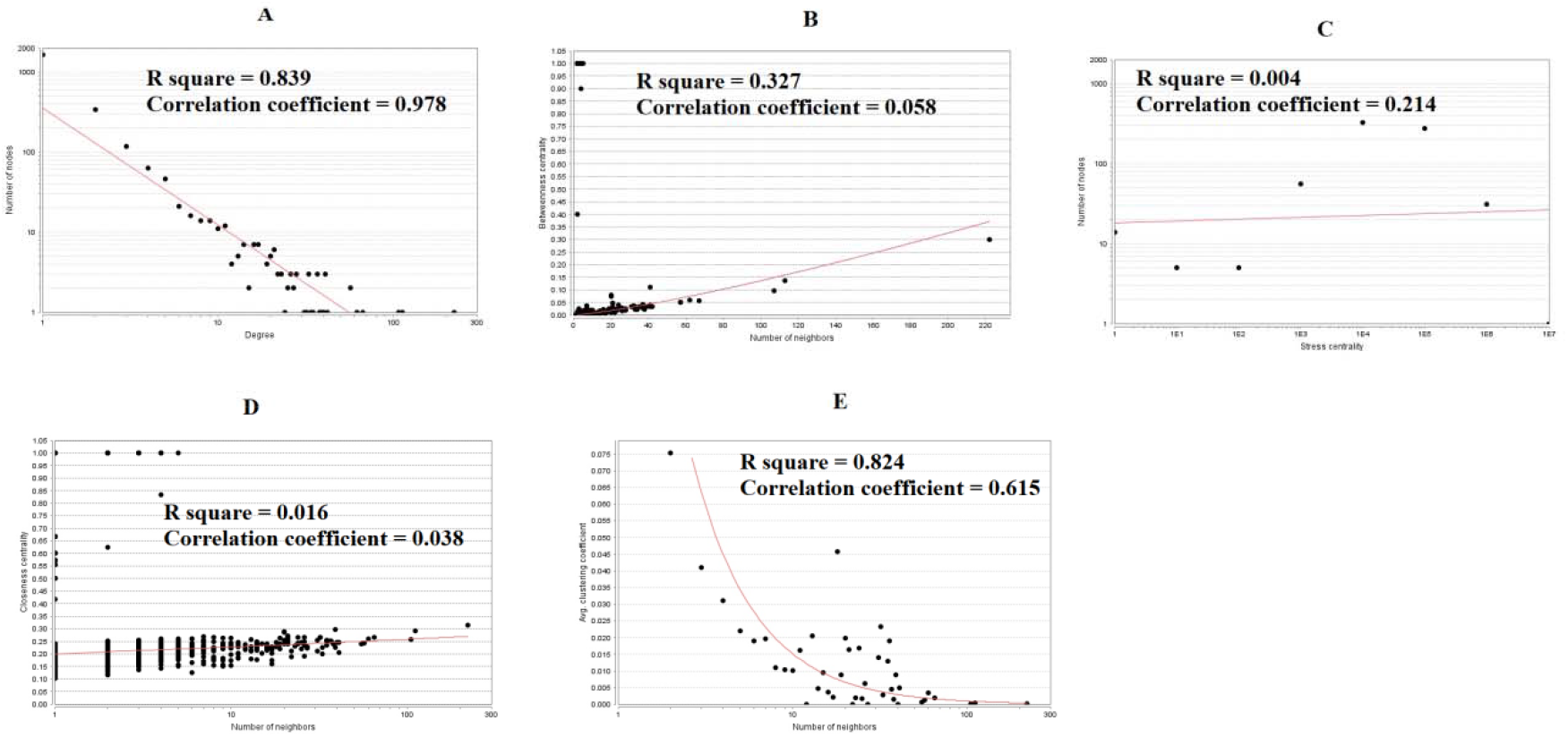
Scatter plot for down regulated genes. (A- Node degree; B- Betweenness centrality; C- Stress centrality; D- Closeness centrality; E- Clustering coefficient)

**Table 6.**
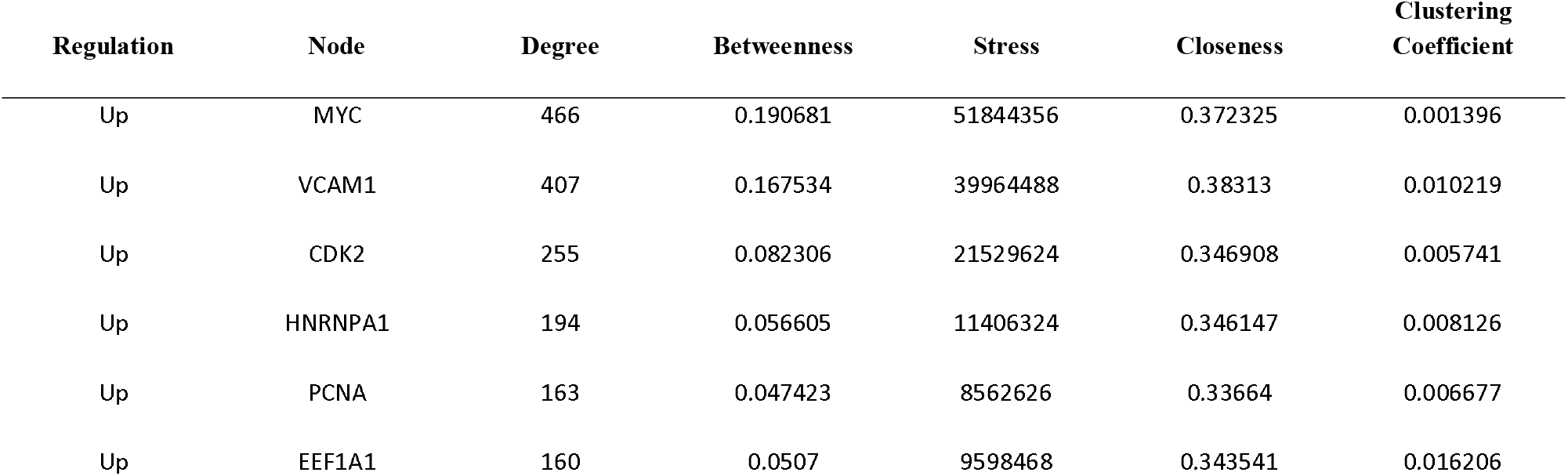
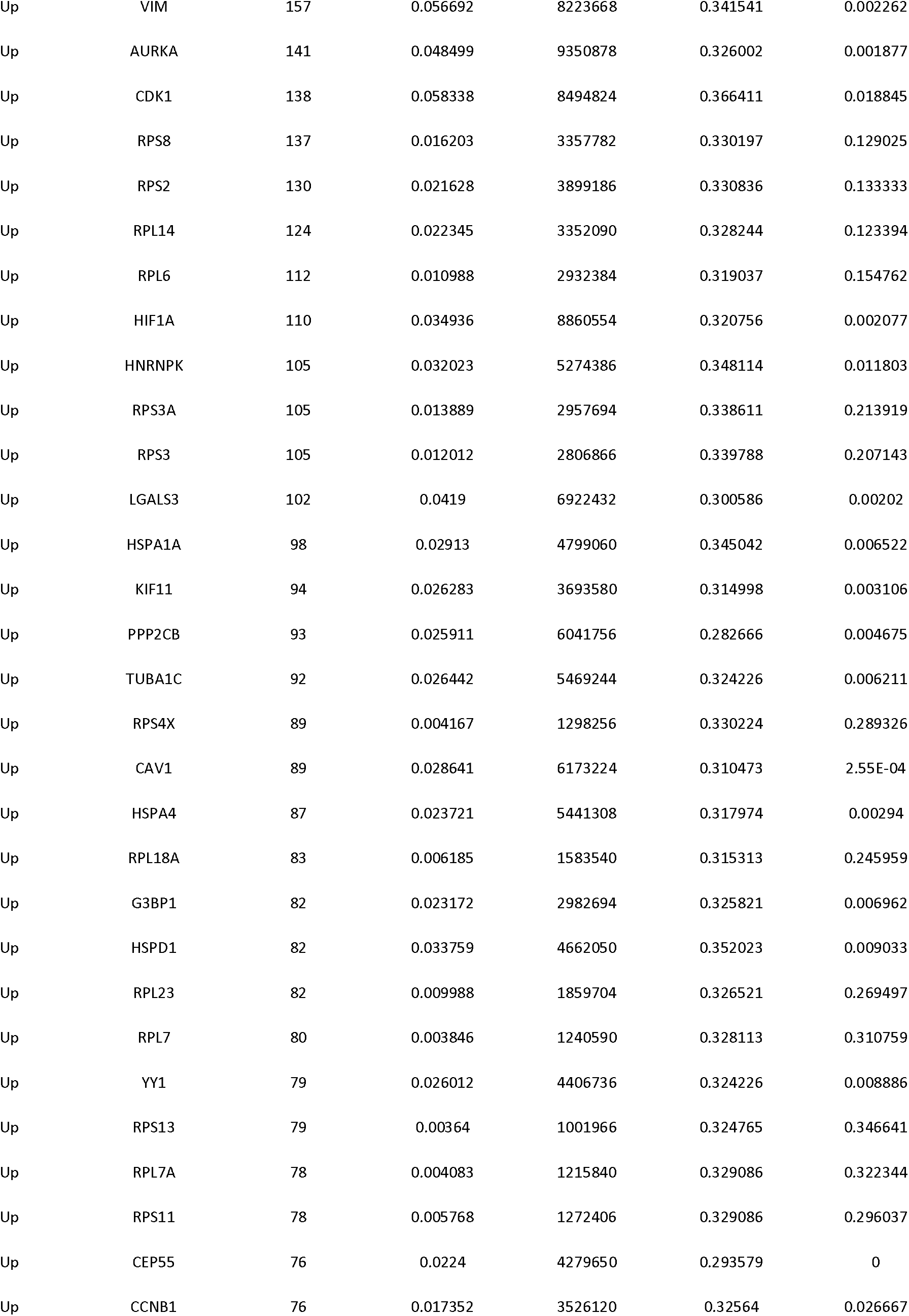
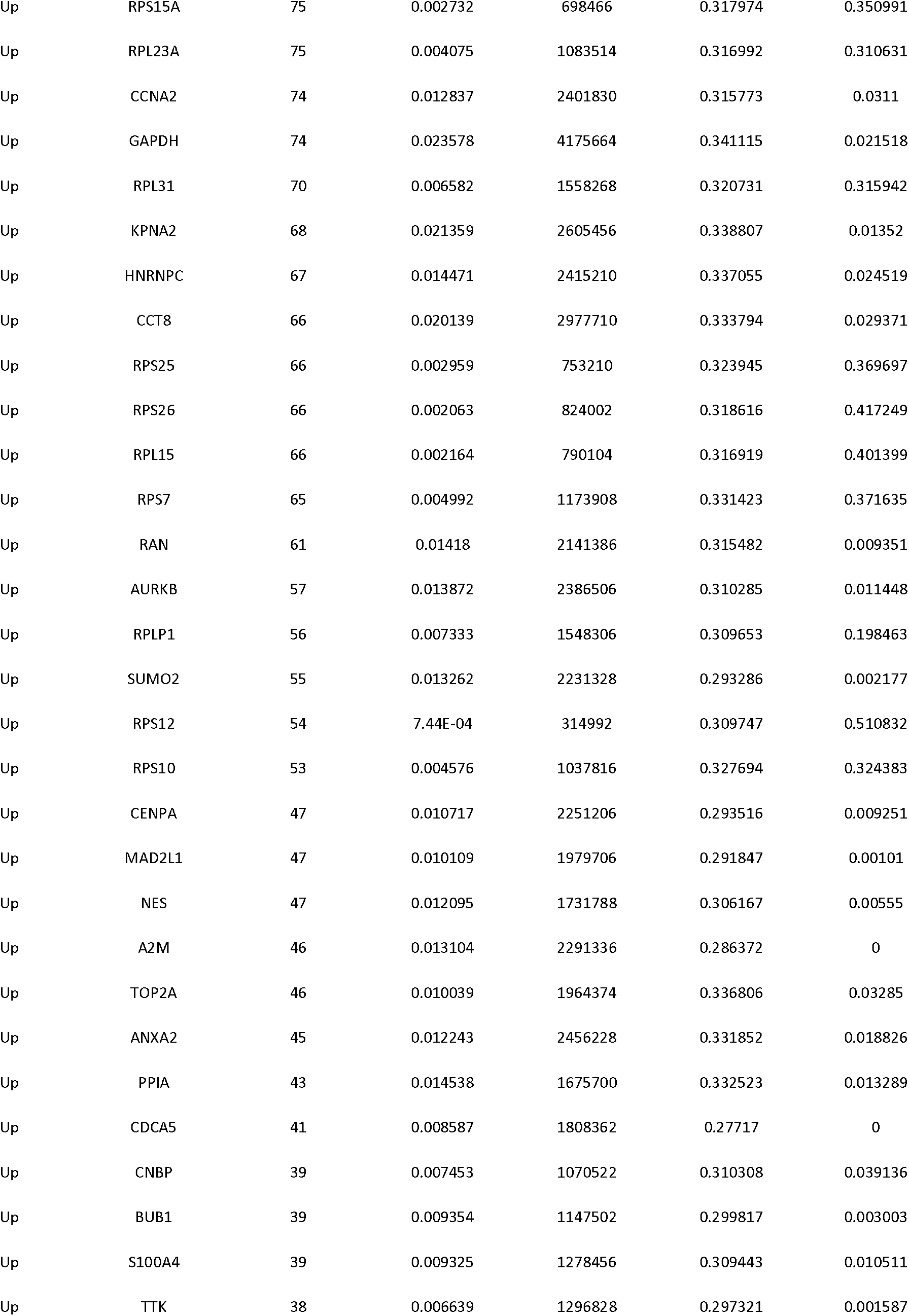
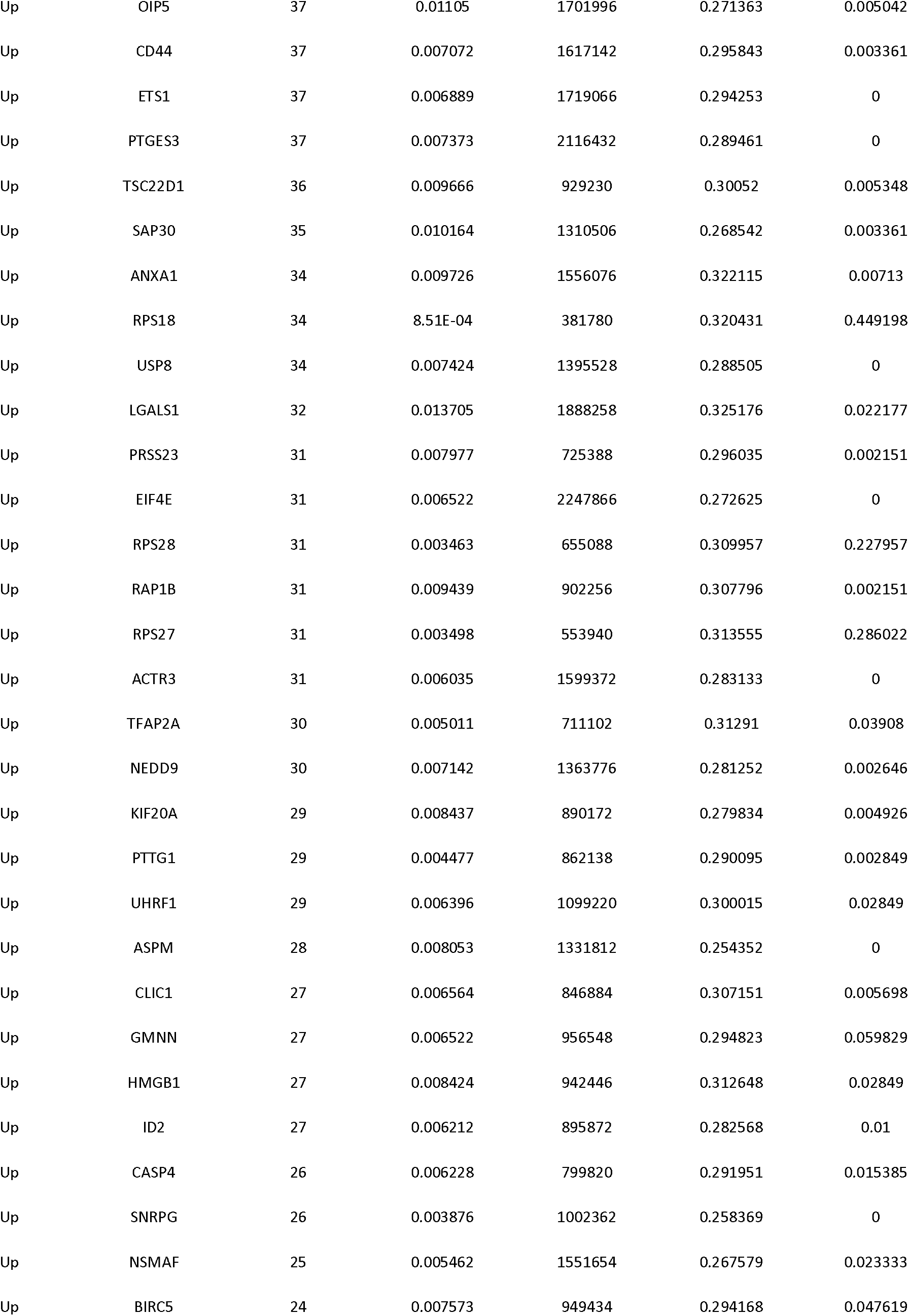
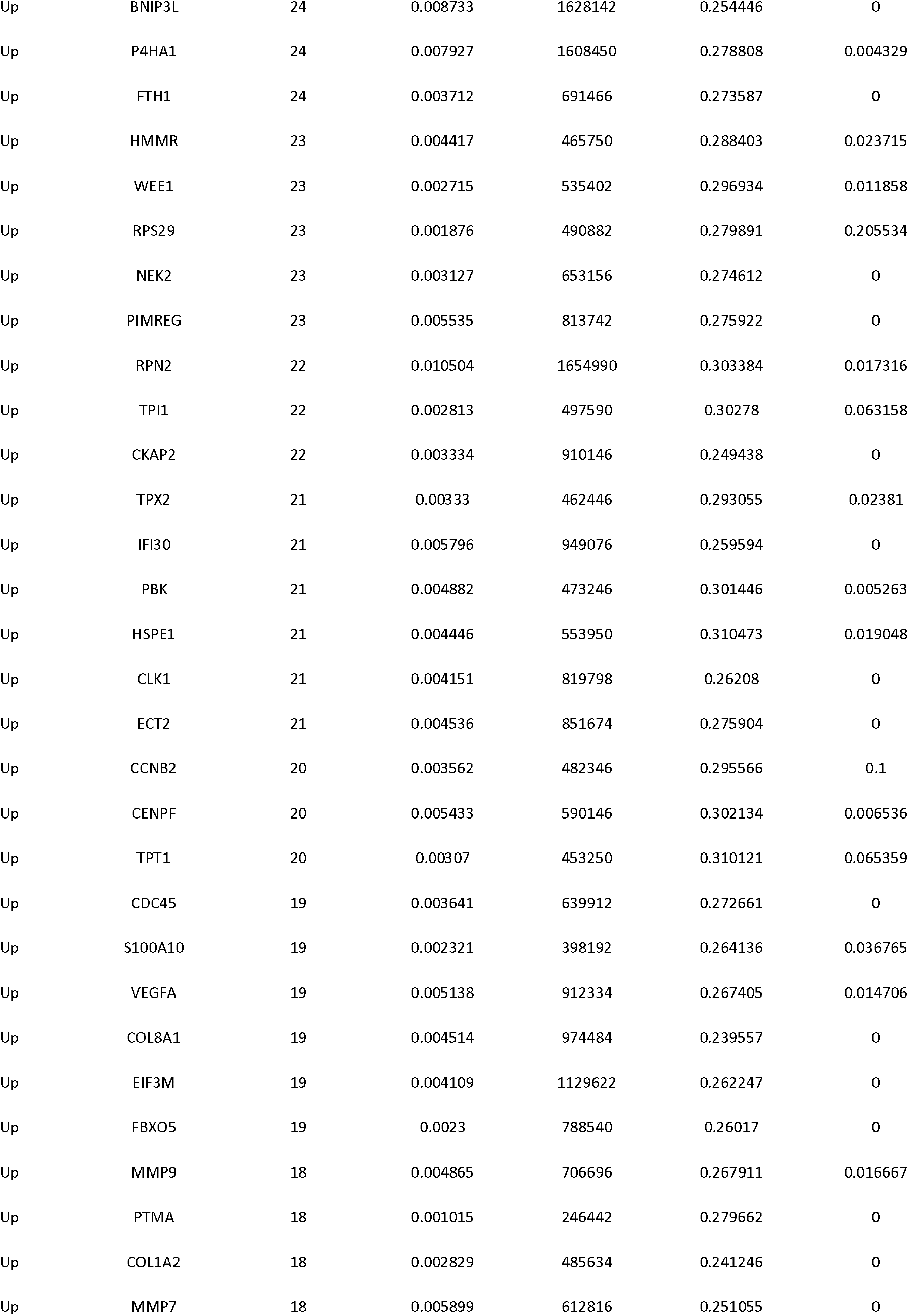
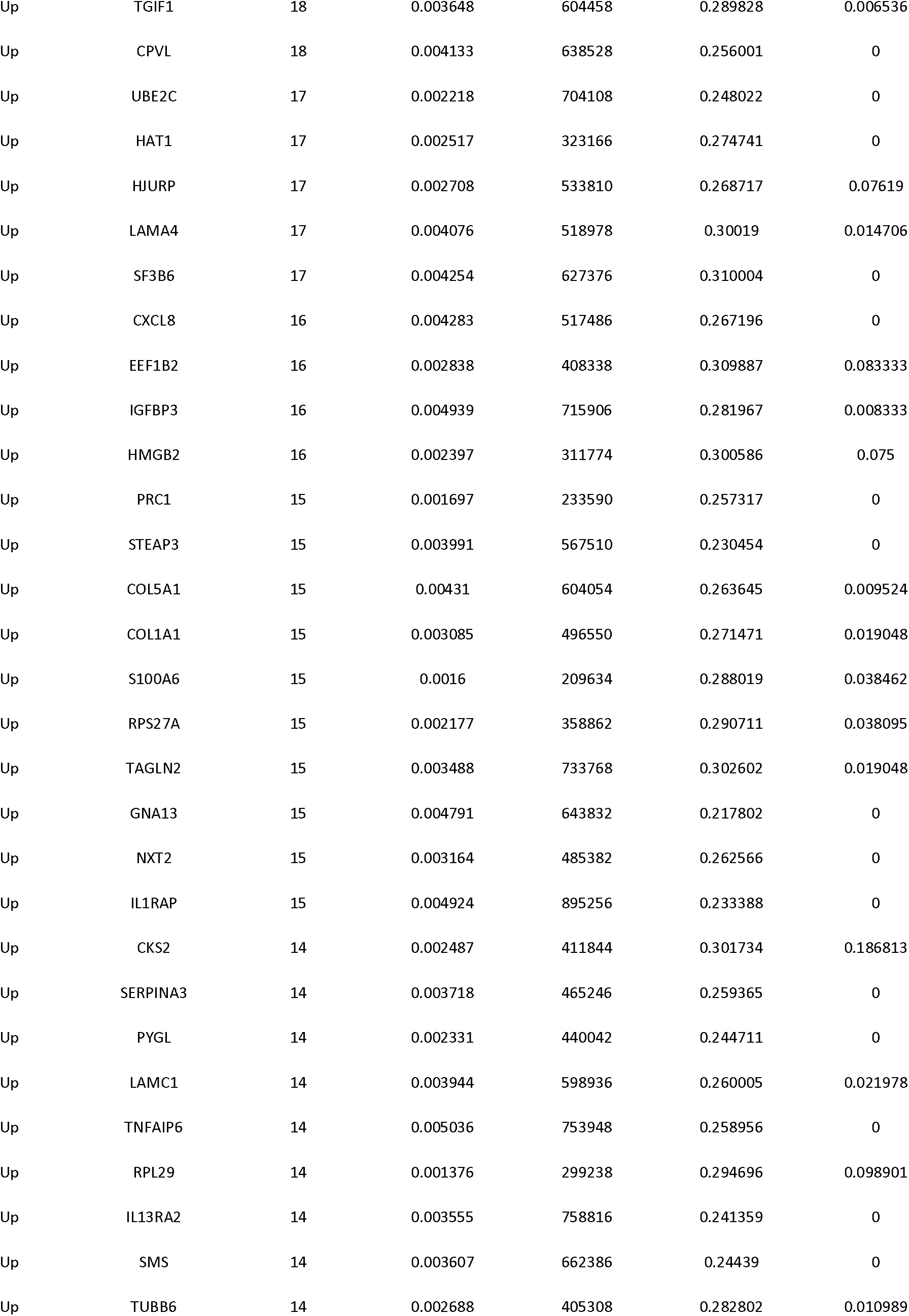
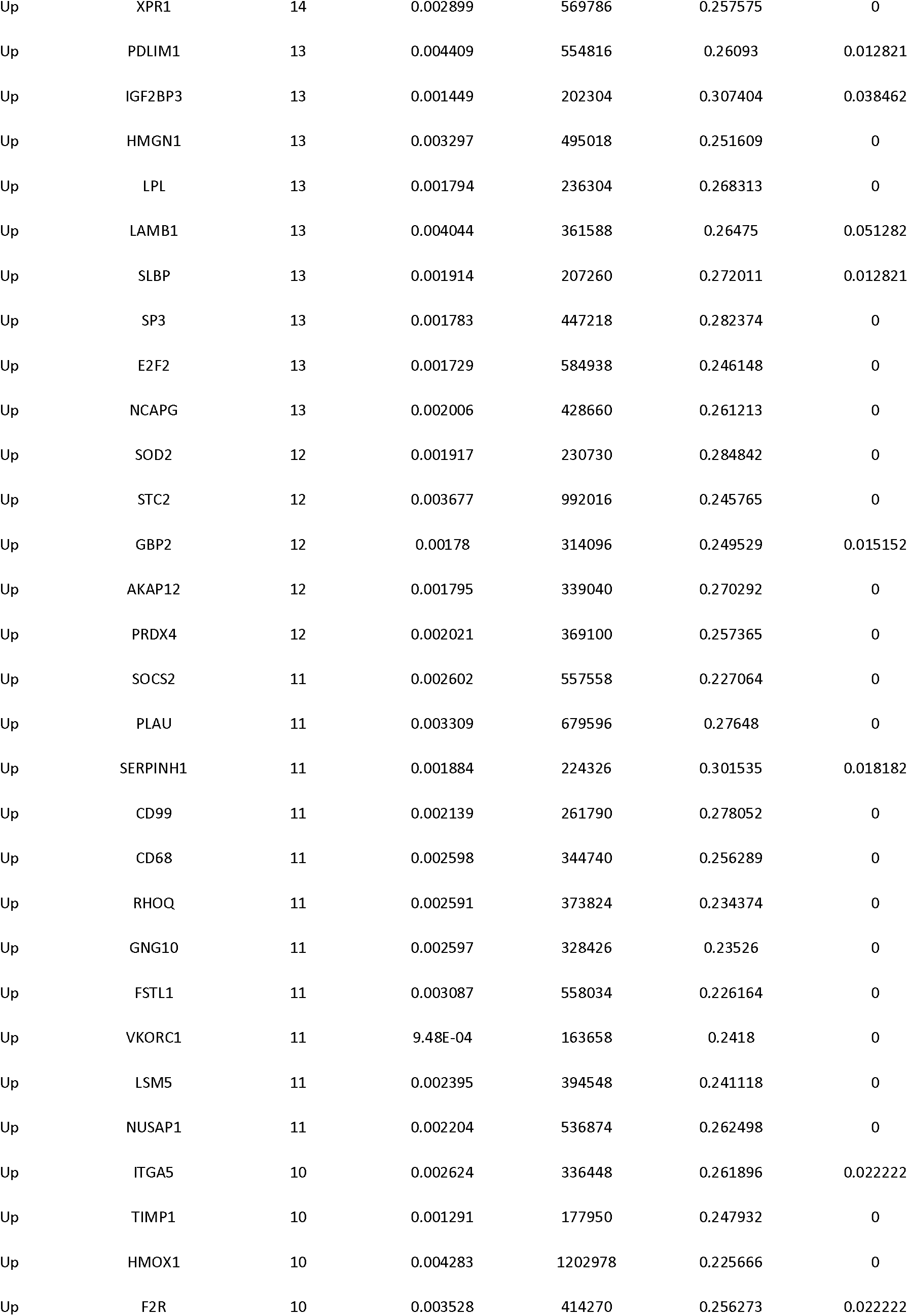
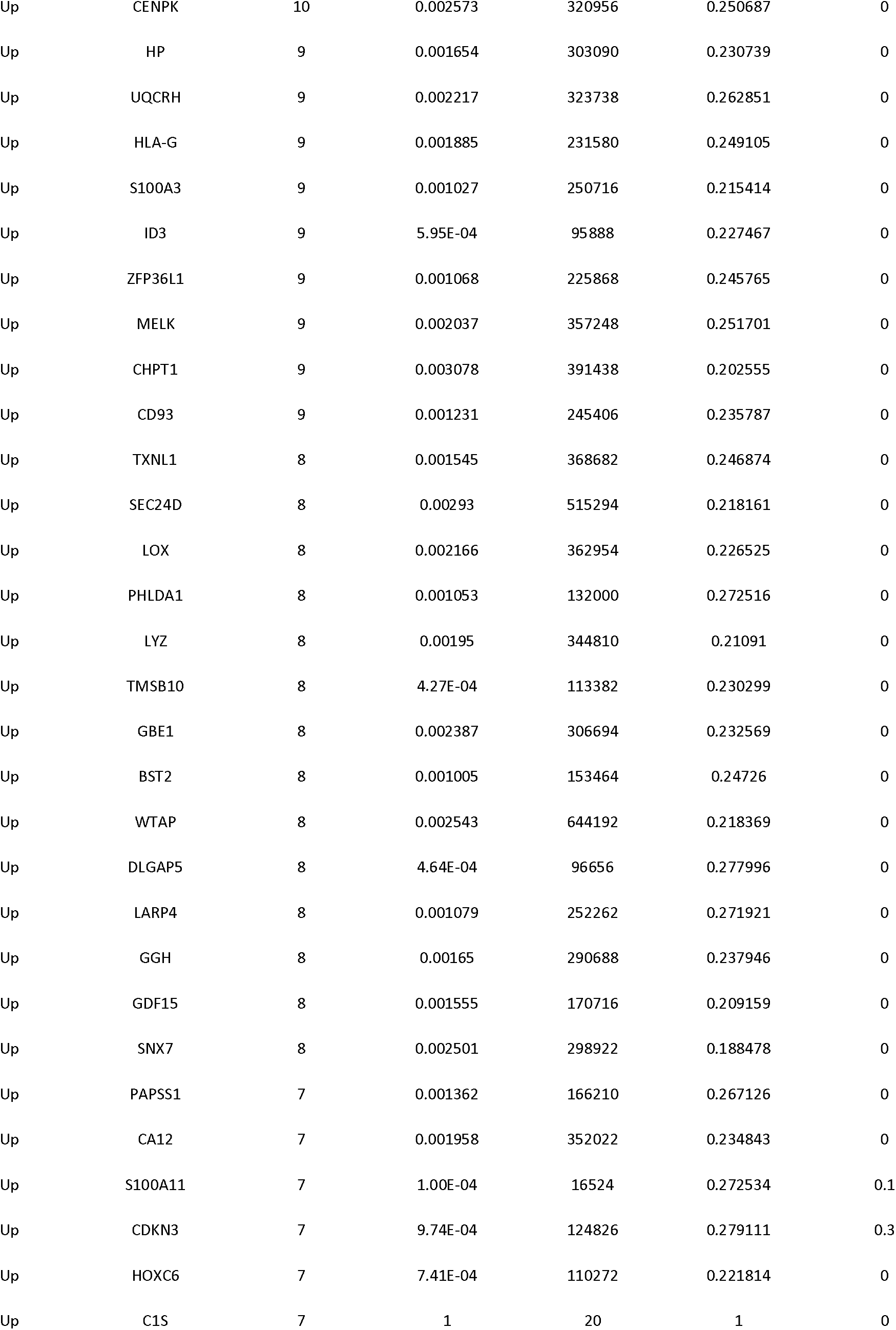
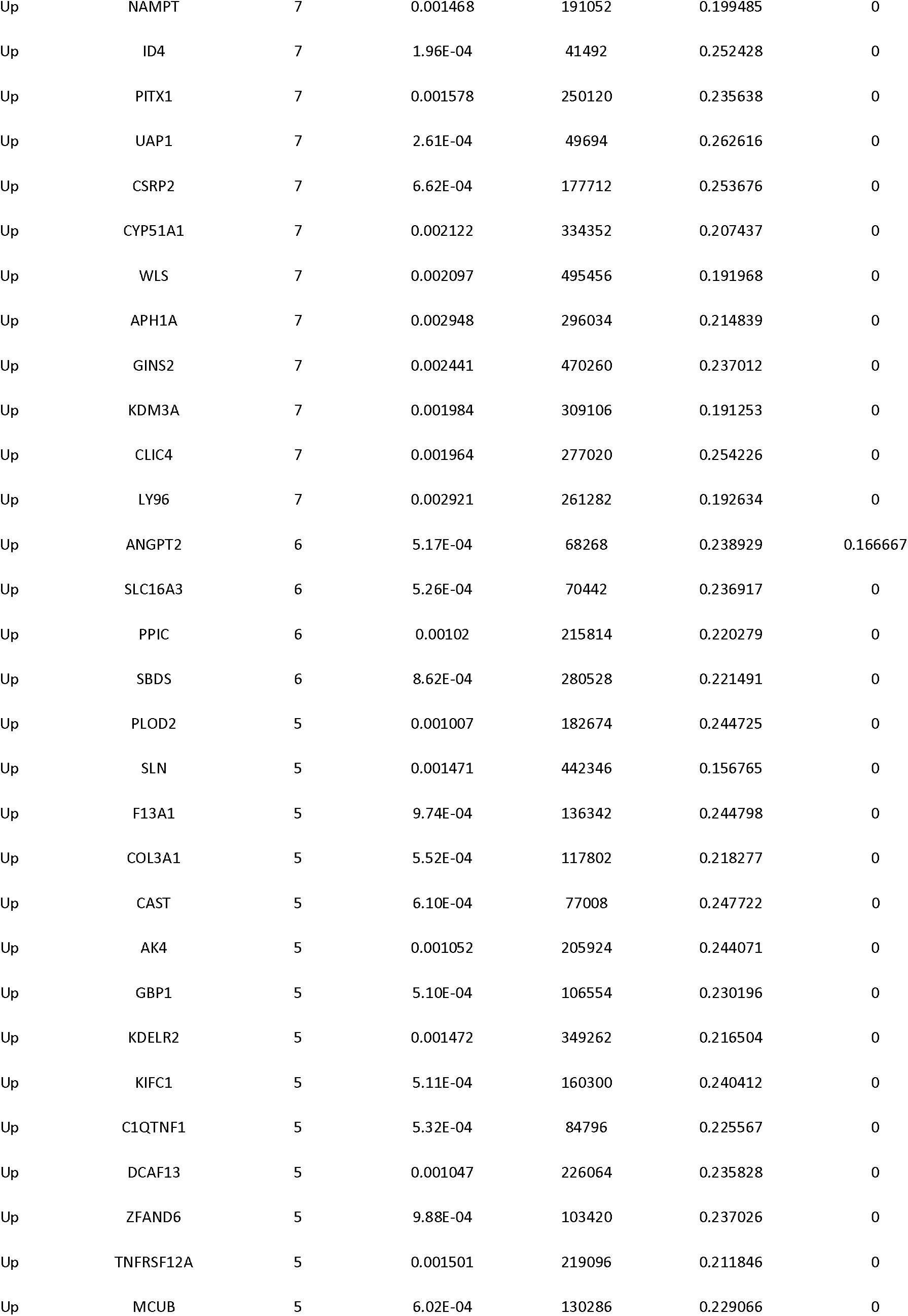
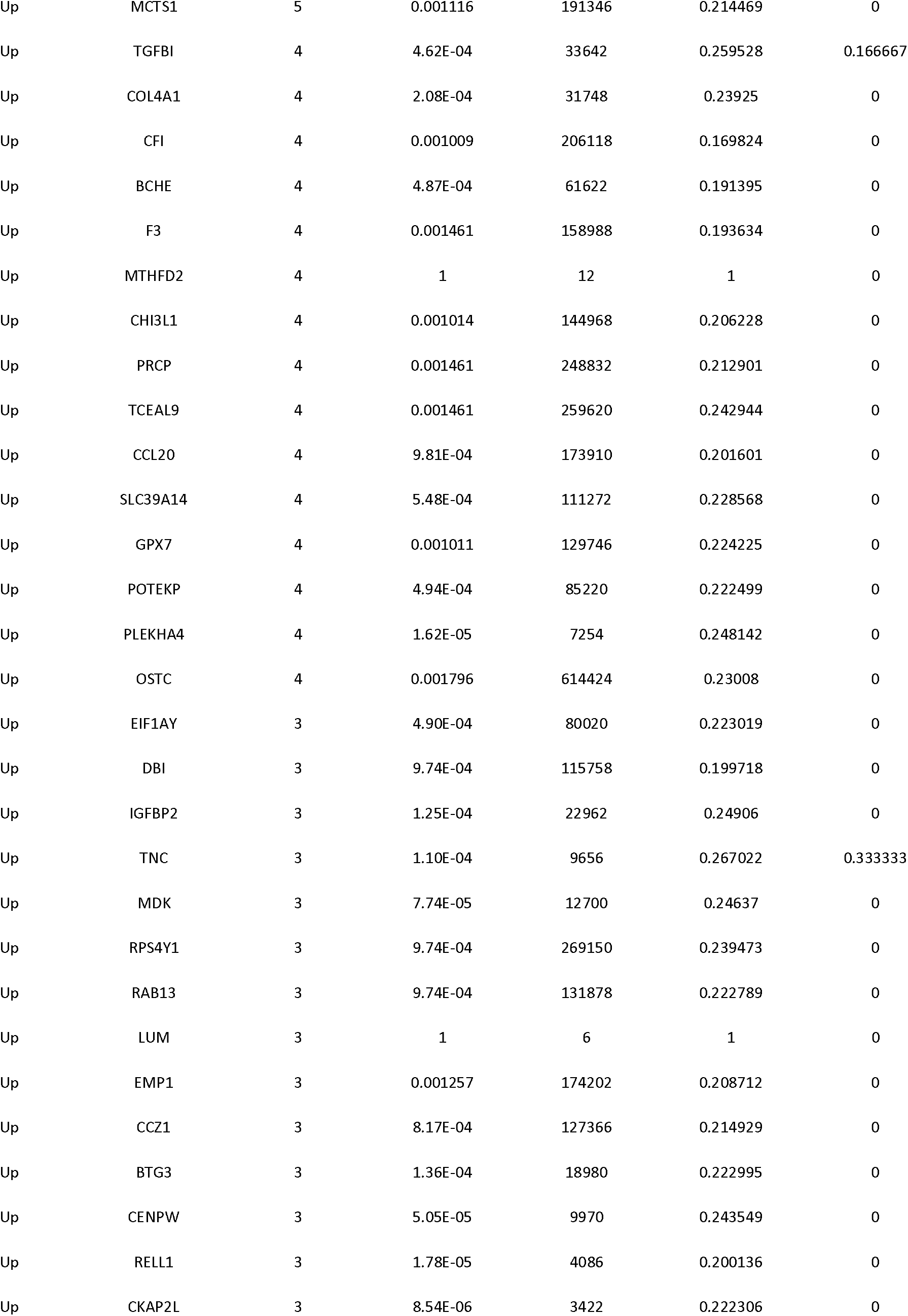
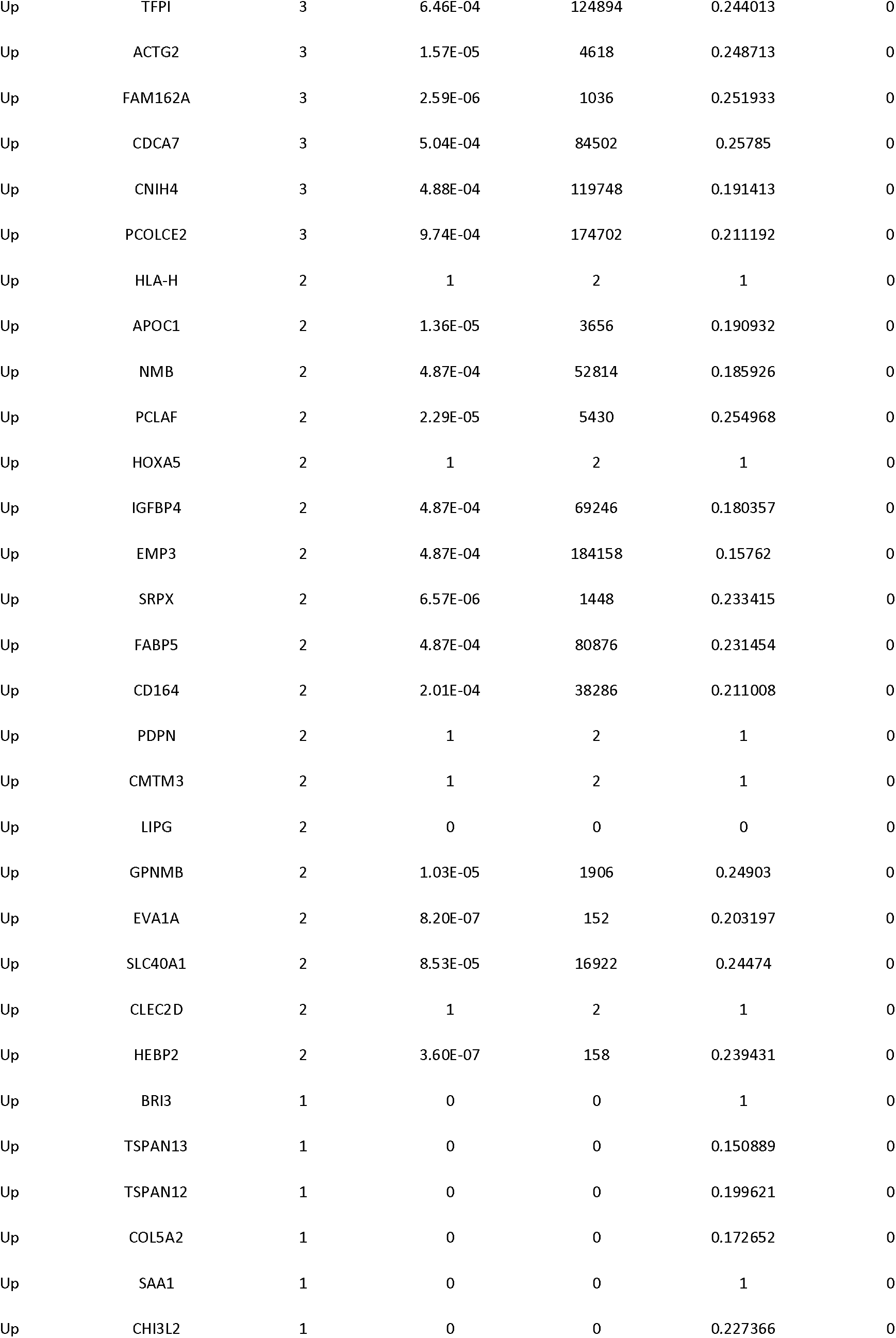
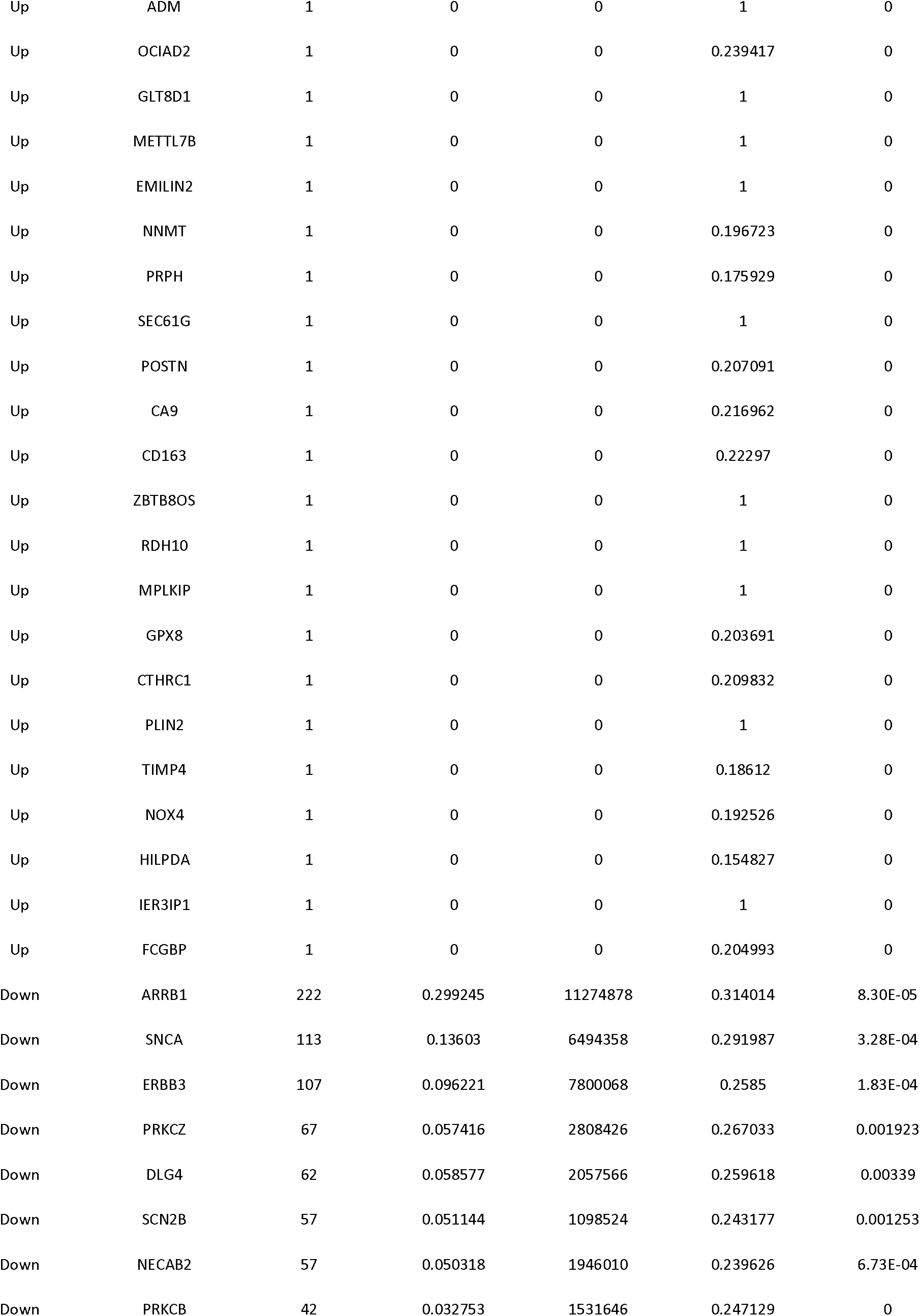
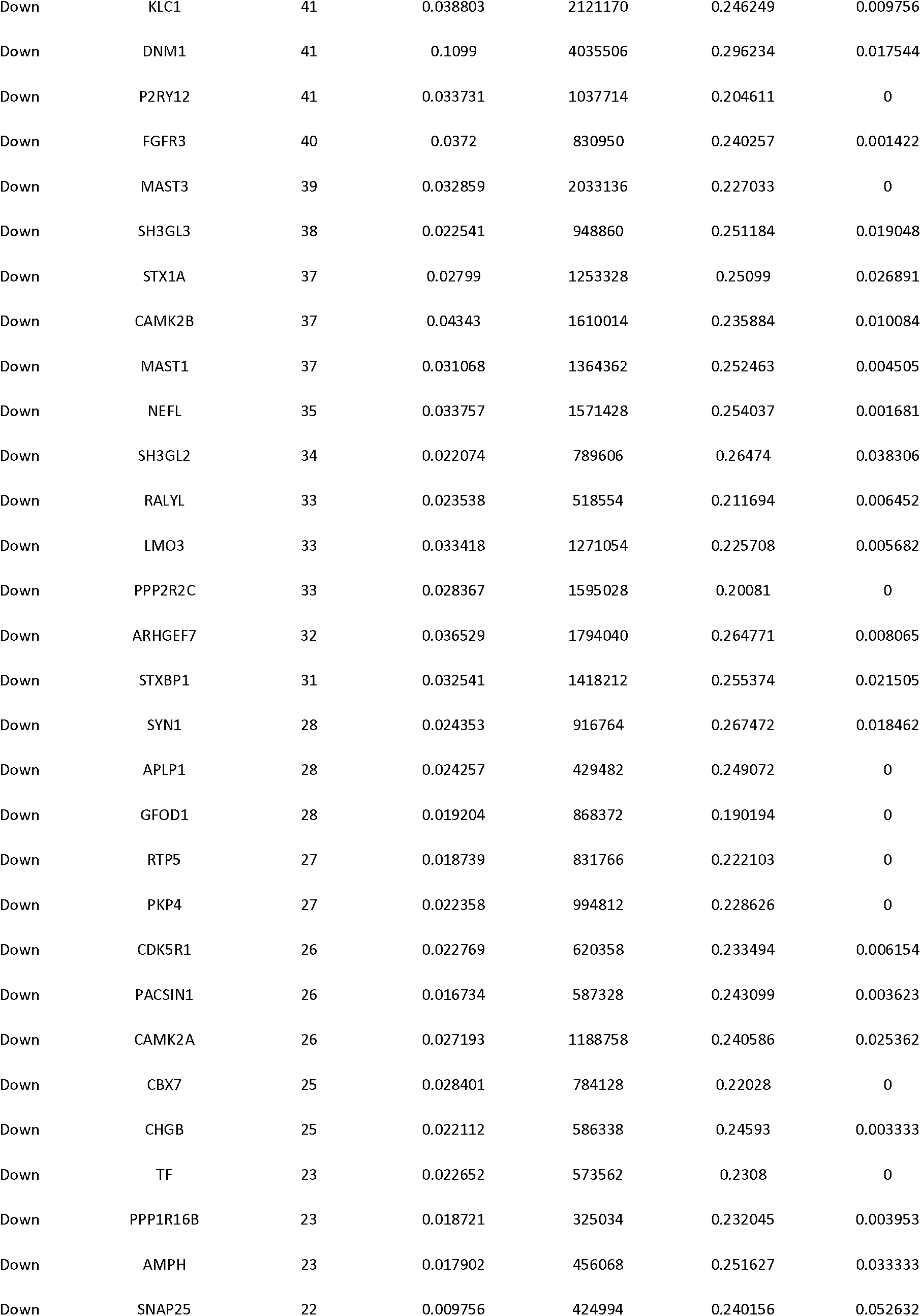
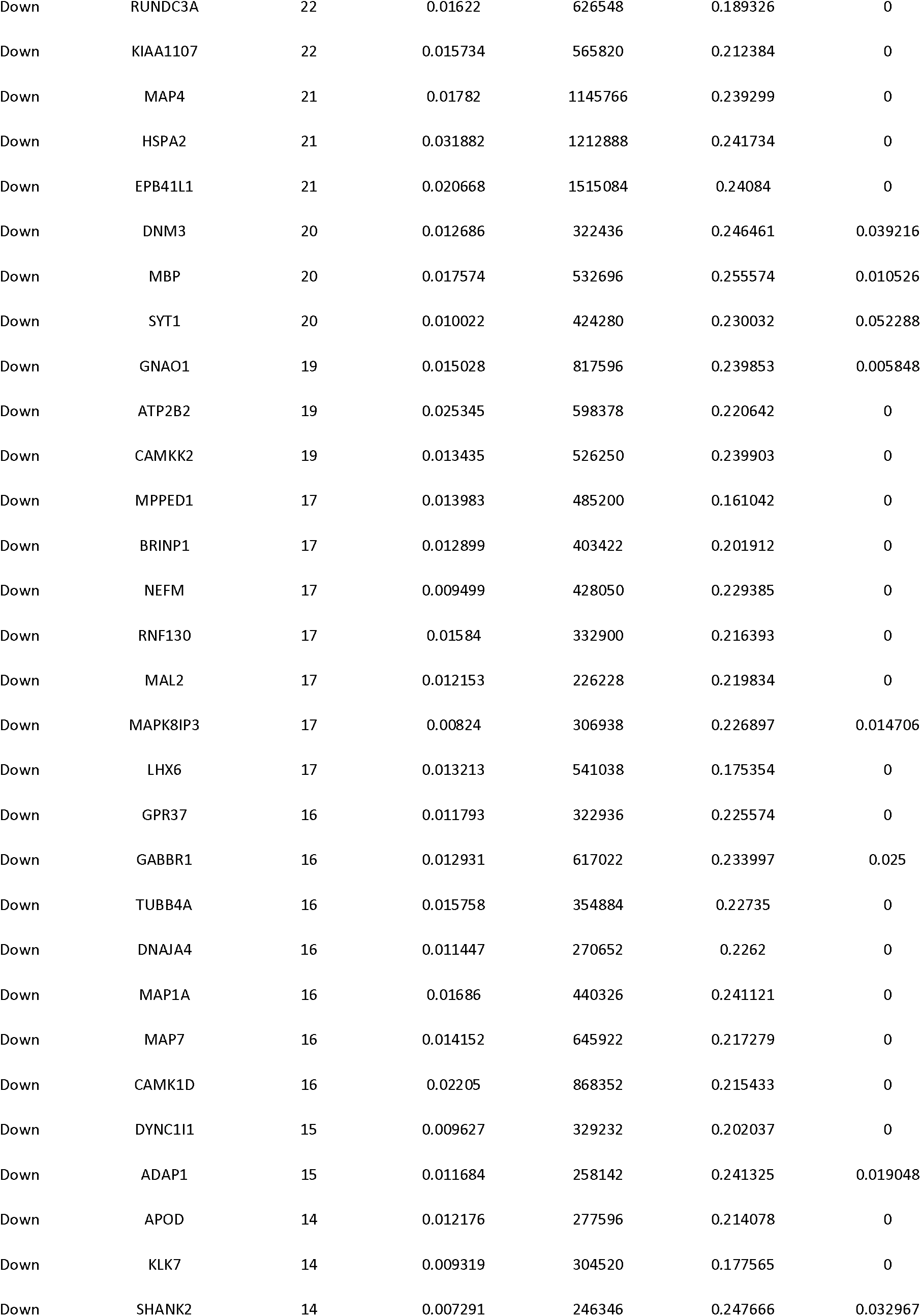
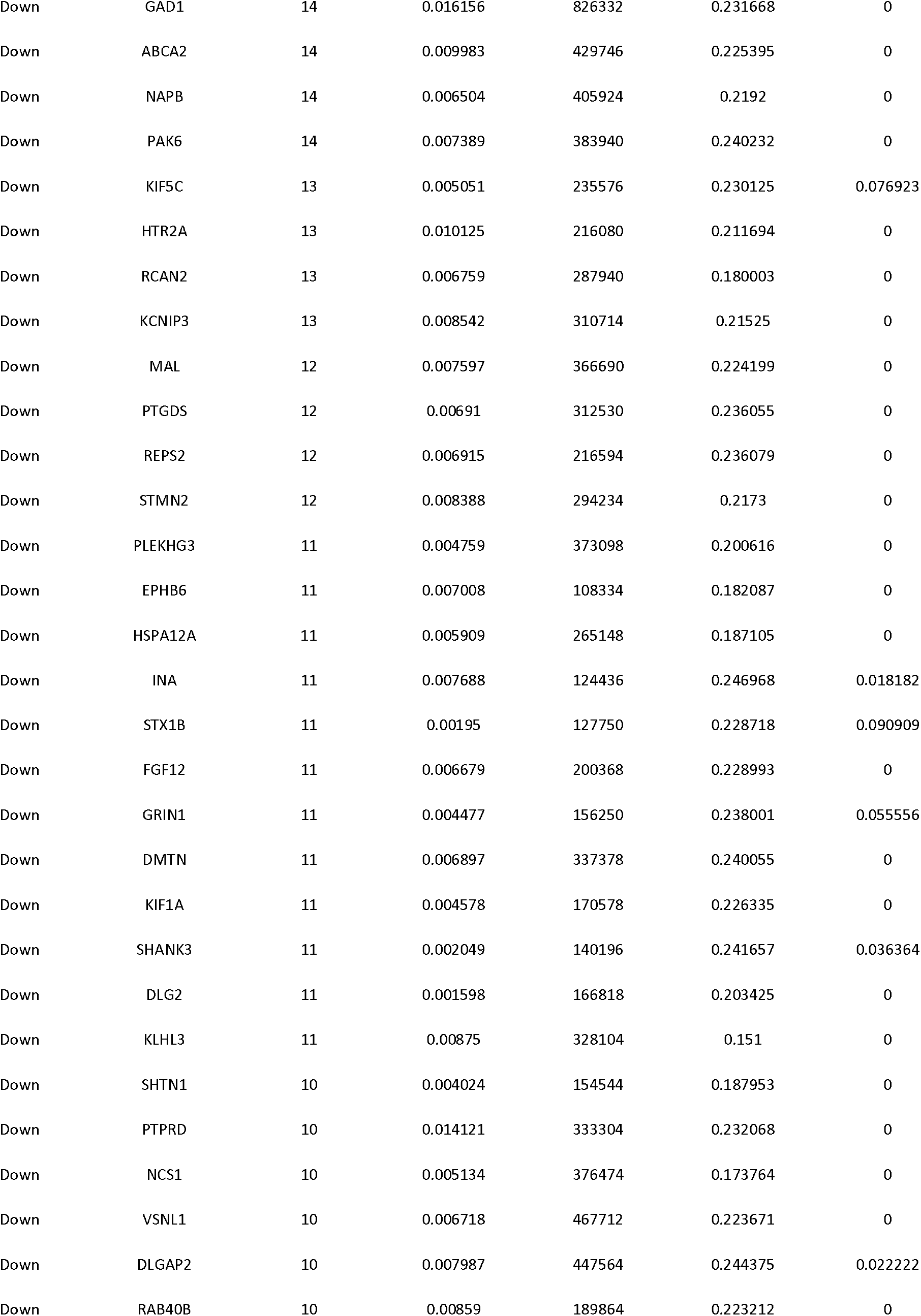
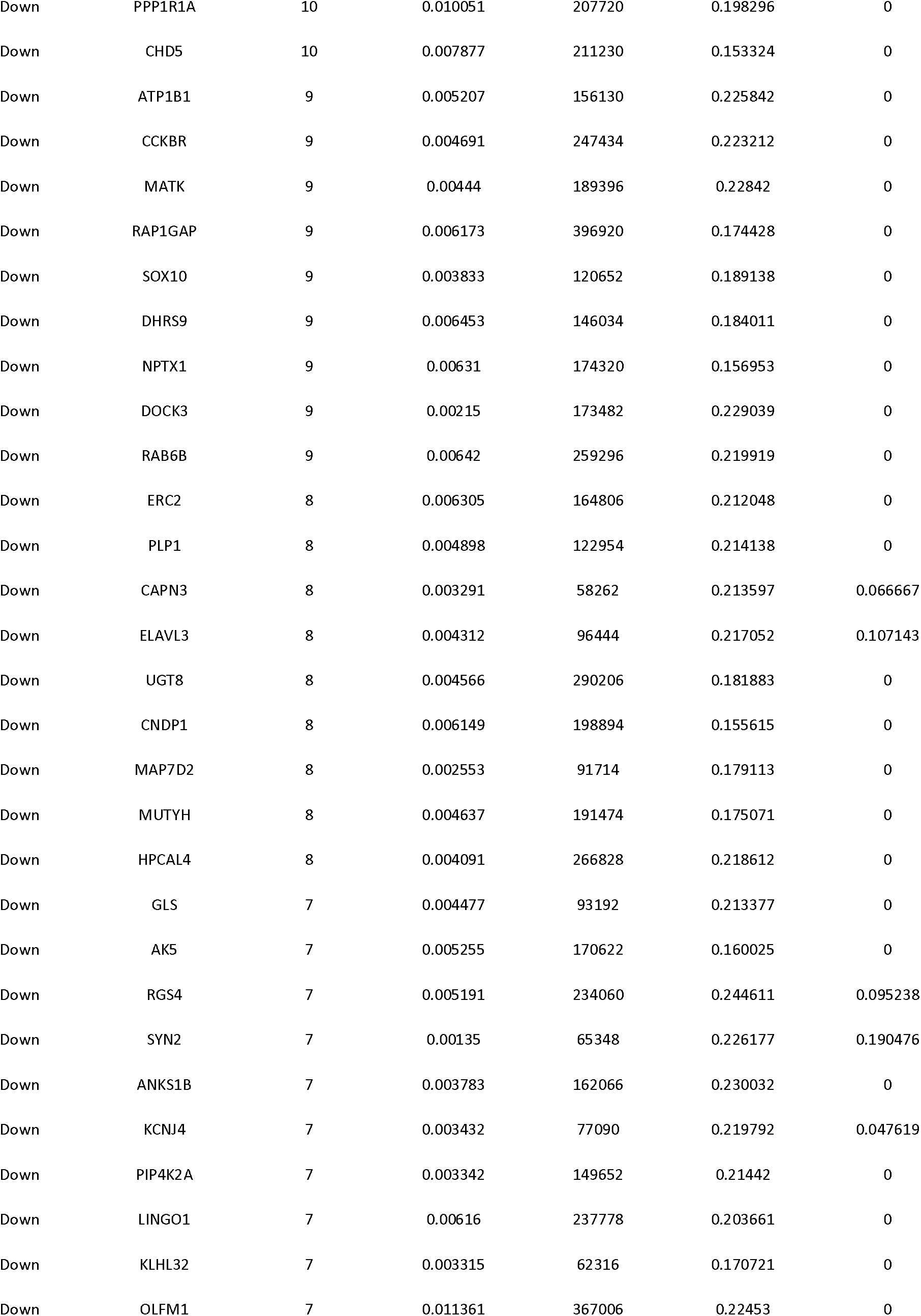
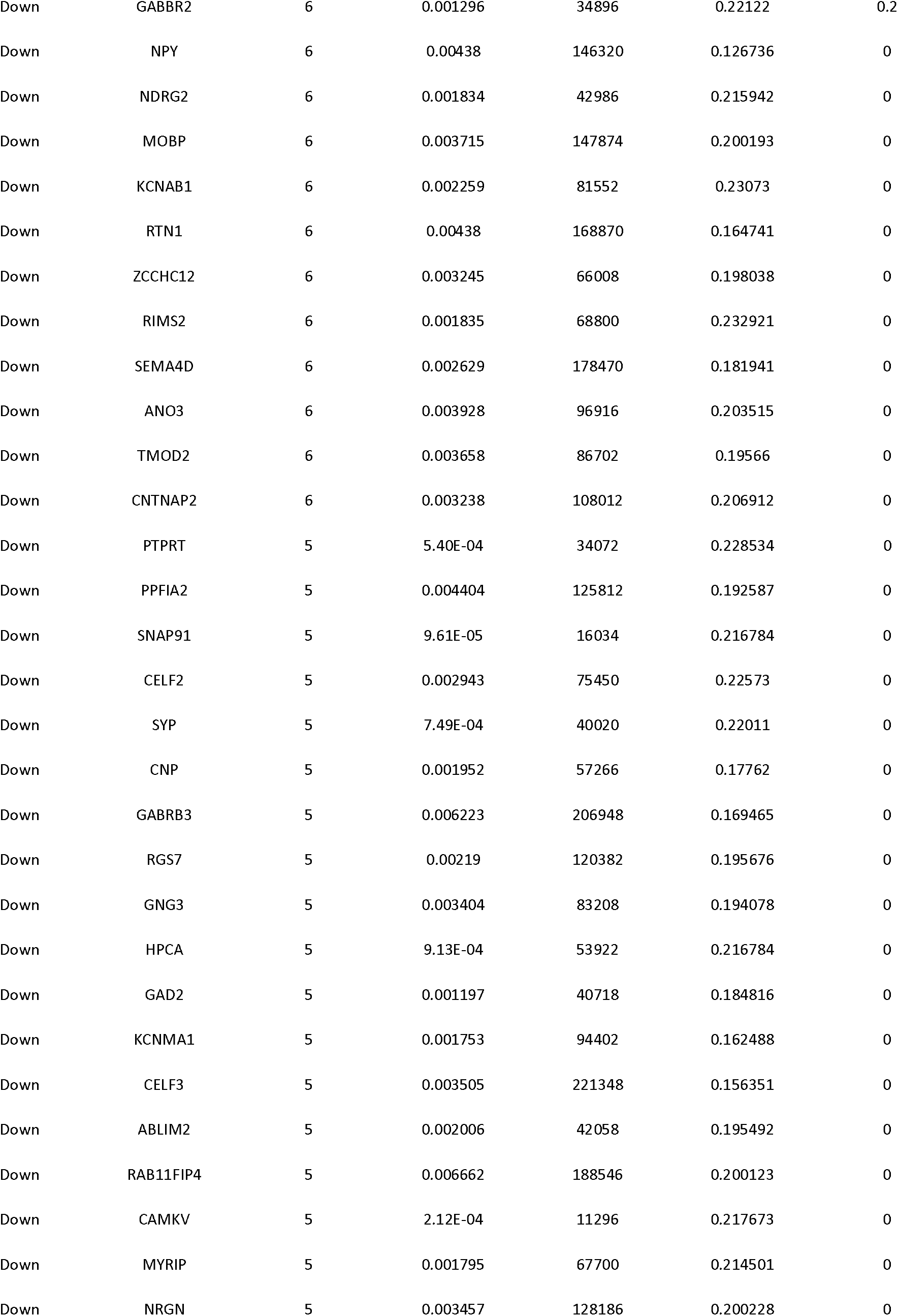
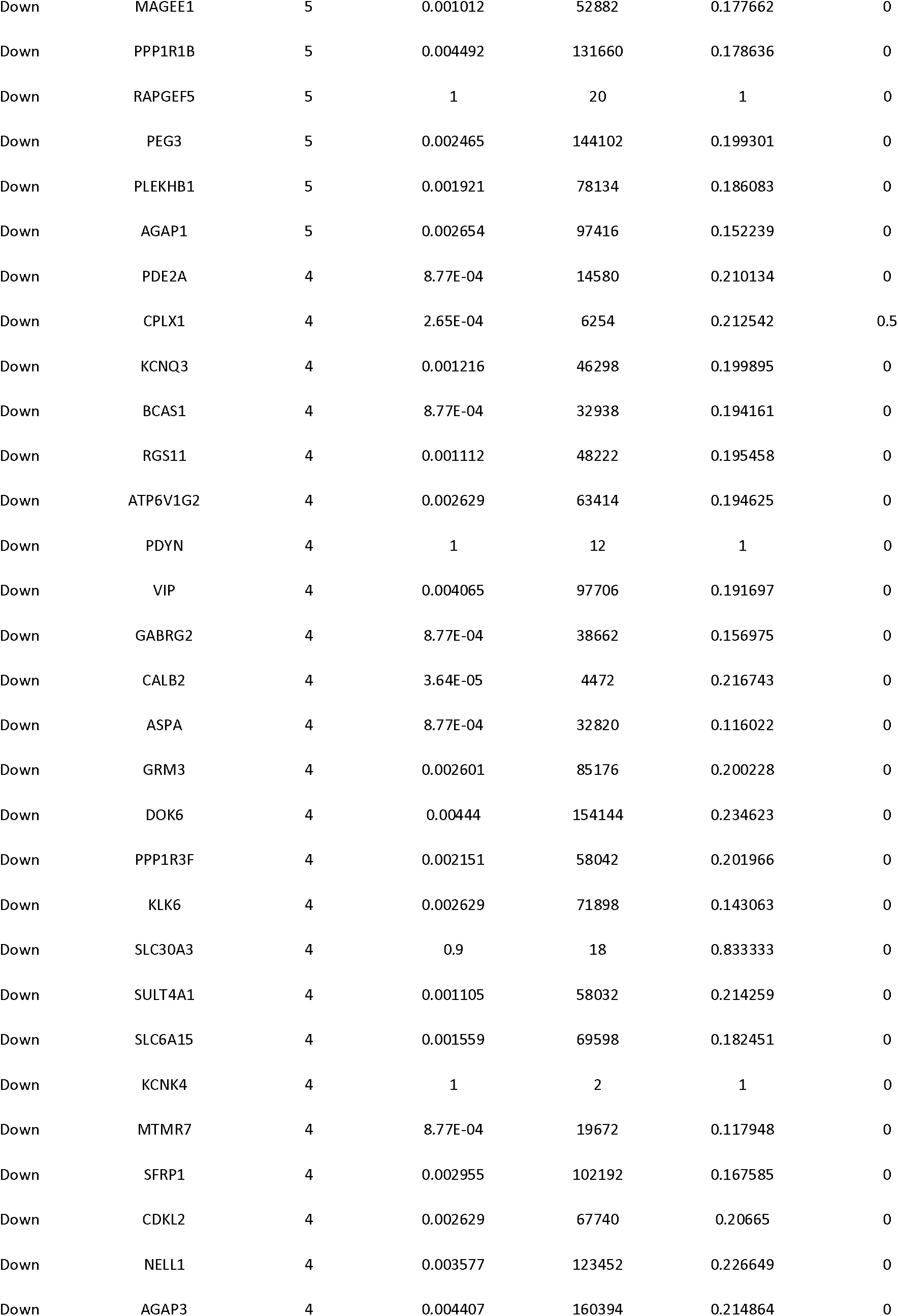
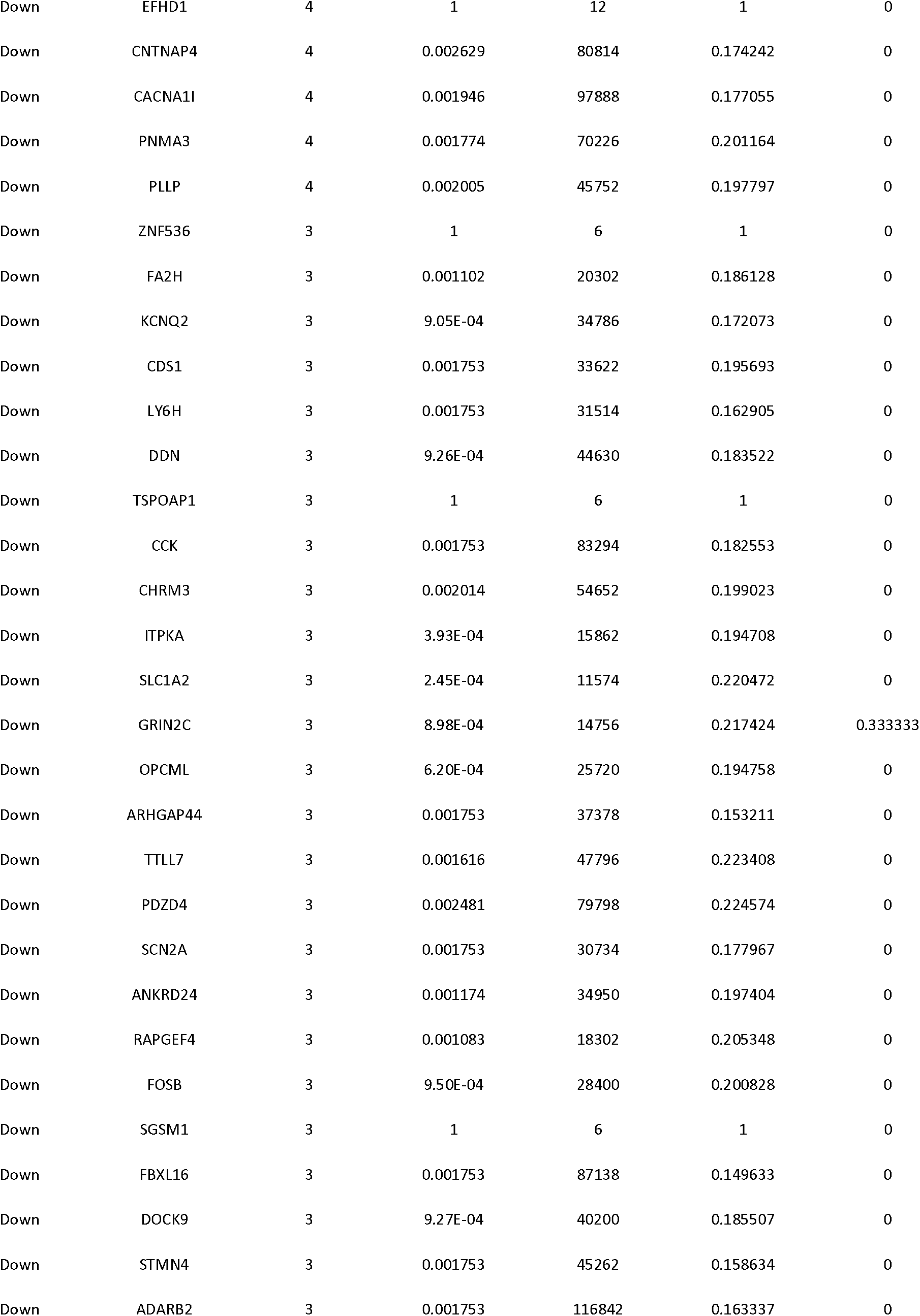
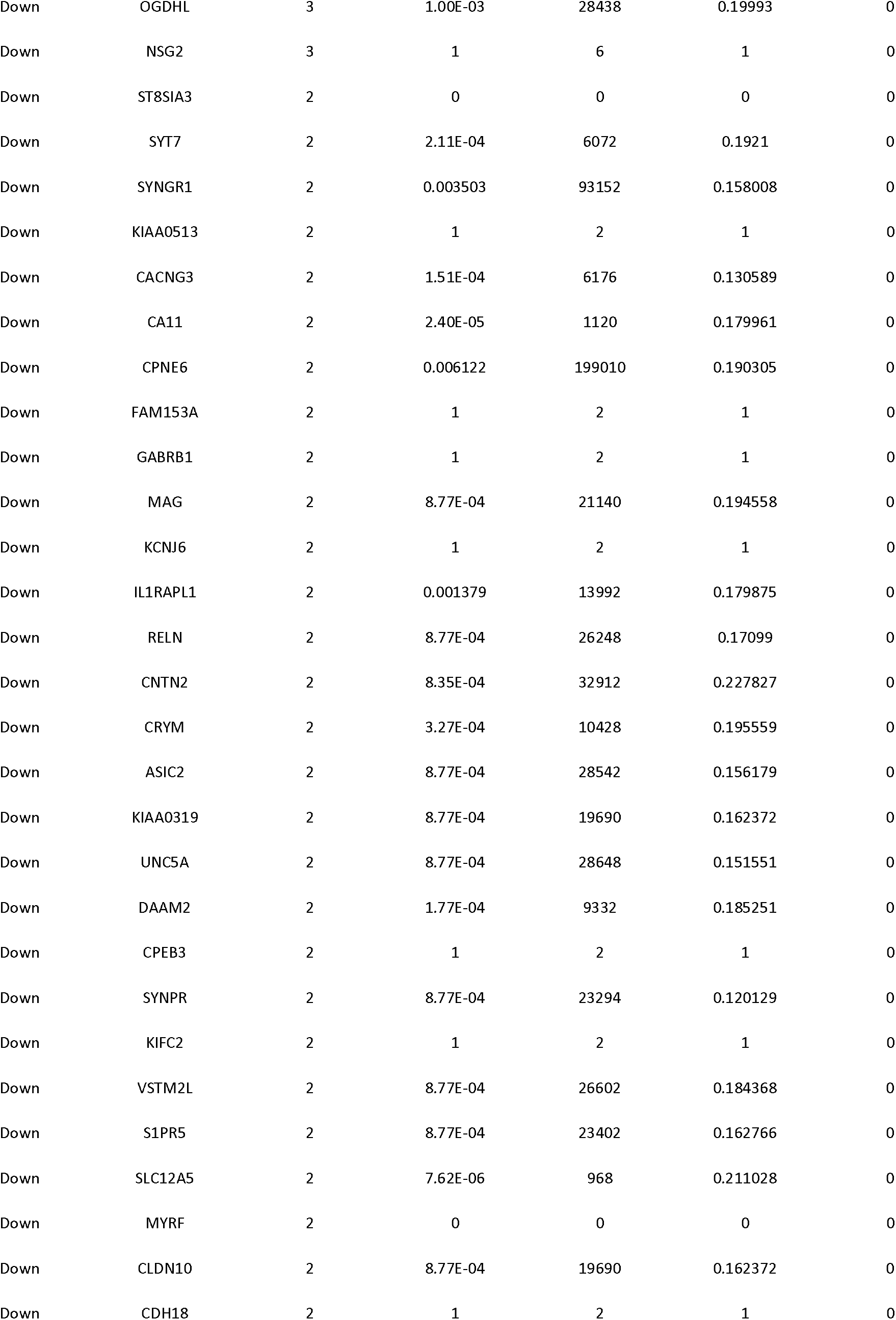
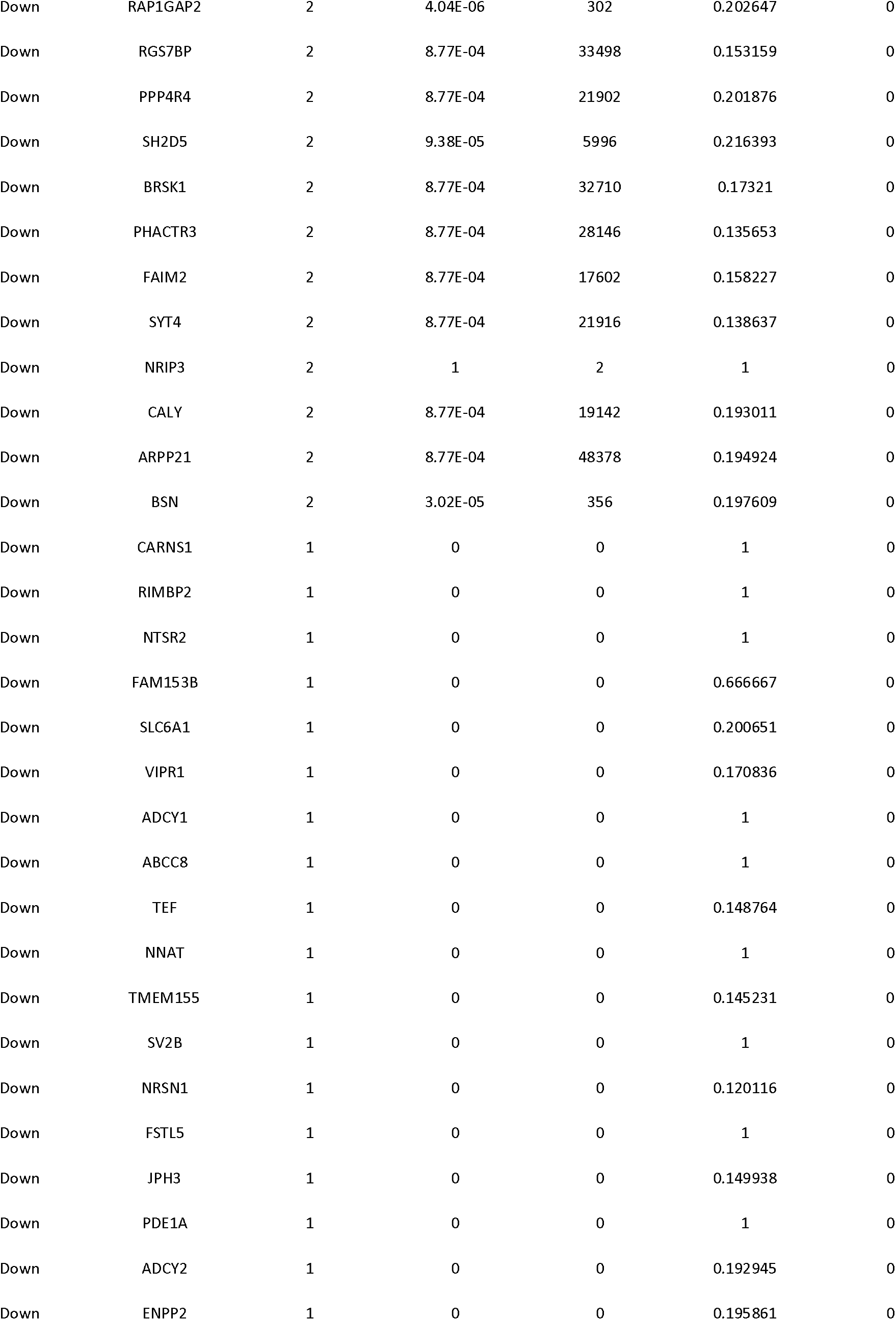
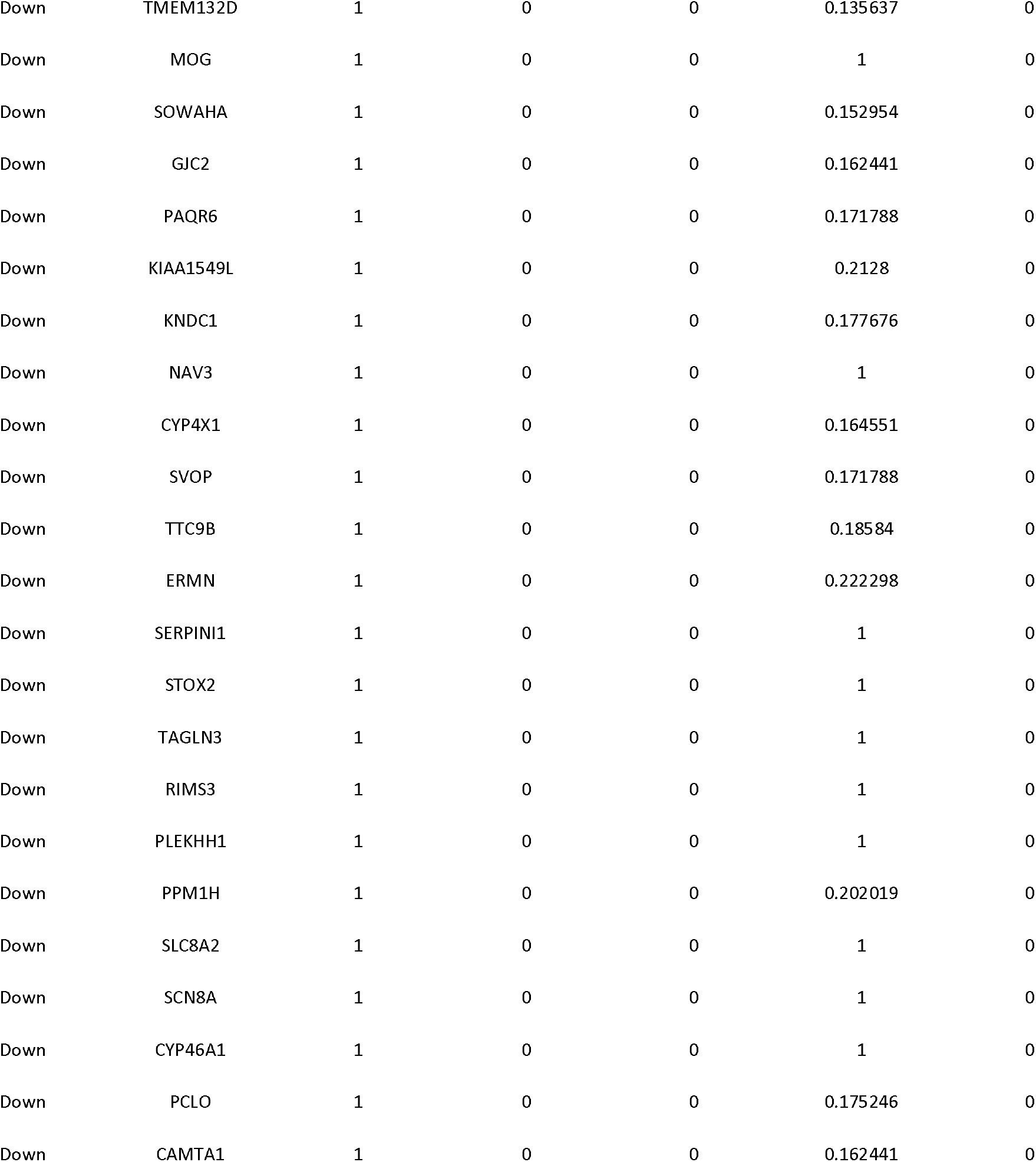
Topology table for up and down regulated genes

Subsequently, we performed module analysis of the whole network by the PEWCC1 plug-in. Total 849 modules were identified in PPI network for up regulated genes. Those hub genes were located at module 6, module 15, module 24 and module 51, are the most informative modules in PPI analysis (Fig. 10). These significant modules were proven to be associated with different pathways and GO categories such as ribosome, cell cycle, TNF signaling pathway, pathways in cancer, macromolecule catabolic process, mitotic cell cycle, RNA binding and cytosolic part. Similarly, total 201 modules were identified in PPI network for down regulated genes. Those hub genes were located at module 2, module 7, module 18 and module 22, are the most informative modules in PPI analysis (Fig. 11). These significant modules were proven to be associated with different pathways and GO categories such as insulin secretion, synaptic vesicle cycle, glutamatergic synapse, endocytosis, synaptic signaling, neurogenesis, cell-cell signaling and neuron differentiation.

**Fig. 10.**
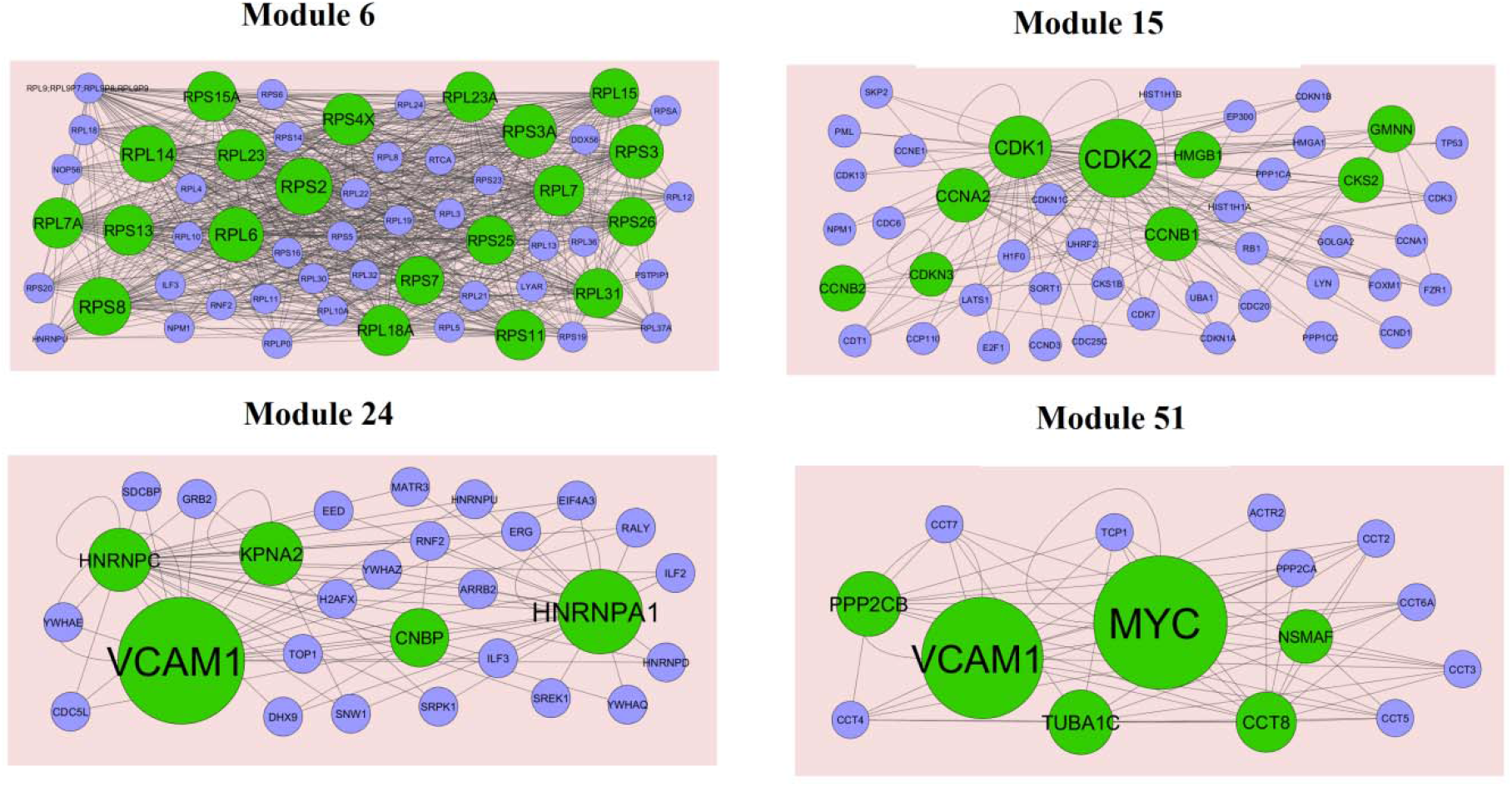
Modules in PPI network. The green nodes denote the up regulated genes

**Fig. 11.**
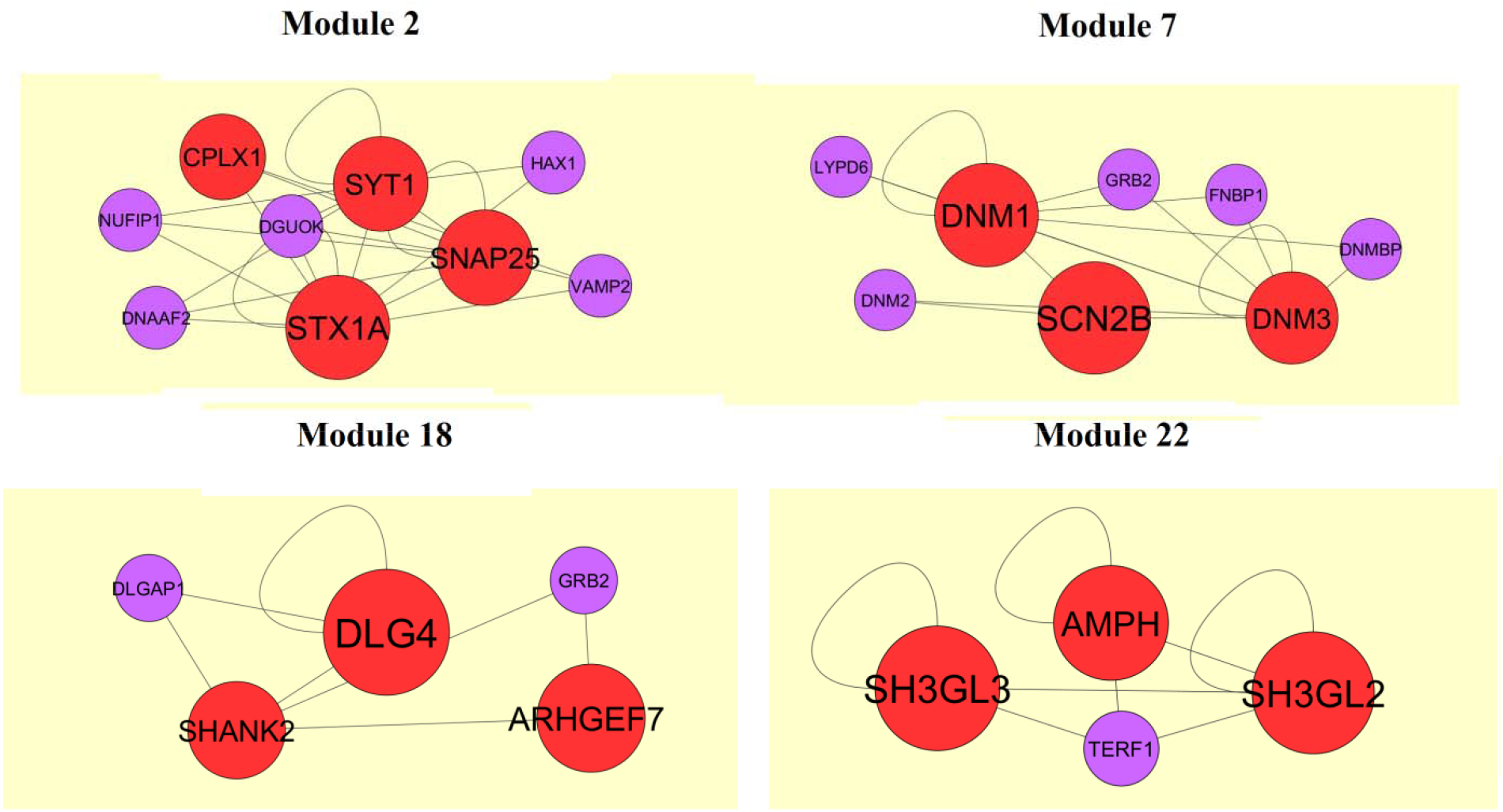
Modules in PPI network. The red nodes denote the down regulated genes

### Construction of target gene - miRNA regulatory network

For further research of the target genes (up and down regulated genes), target gene-related miRNAs were predicted by DIANA-TarBase and miRTarBase. Main miRNAs with interactions of target genes are listed in Table 7. Target genes were found to play a key role in regulating miRNAs. The target genes - miRNA regulatory network (up regulated genes) included 2440 nodes and 8546 edges (Fig.12). SOD2 was predicted to regulate 257 miRNAs (eg, hsa-mir-6077), WEE1 was predicted to regulate 167 miRNAs (eg, hsa-mir-4457), G3BP1 was predicted to regulate 158 miRNAs (eg, hsa-mir-4457), CNBP was predicted to regulate 153 miRNAs (eg, hsa-mir-4260) and HMGB1 was predicted to regulate 143 miRNAs (eg, hsa-mir-5193). These target genes were enriched in reactive oxygen species degradation, cell cycle, adherens junction, RNA binding and Neutrophil degranulation. The target genes - miRNA regulatory network (down regulated genes) included 2046 nodes and 4596 edges ((Fig.13). SVOP was predicted to regulate 107 miRNAs (eg, hsa-mir-3972), KCNJ6 was predicted to regulate 90 miRNAs (eg, hsa-mir-4287), SYT7 was predicted to regulate 75 miRNAs (eg, hsa-mir-4441), RAB11FIP4 was predicted to regulate 73 miRNAs (eg, hsa-mir-3176) and NPTX1 was predicted to regulate 73 miRNAs (eg, hsa-mir-3119). These target genes were enriched in transmembrane transport, synapse part, neuronal system, endocytosis and synaptic signaling.

**Fig. 12.**
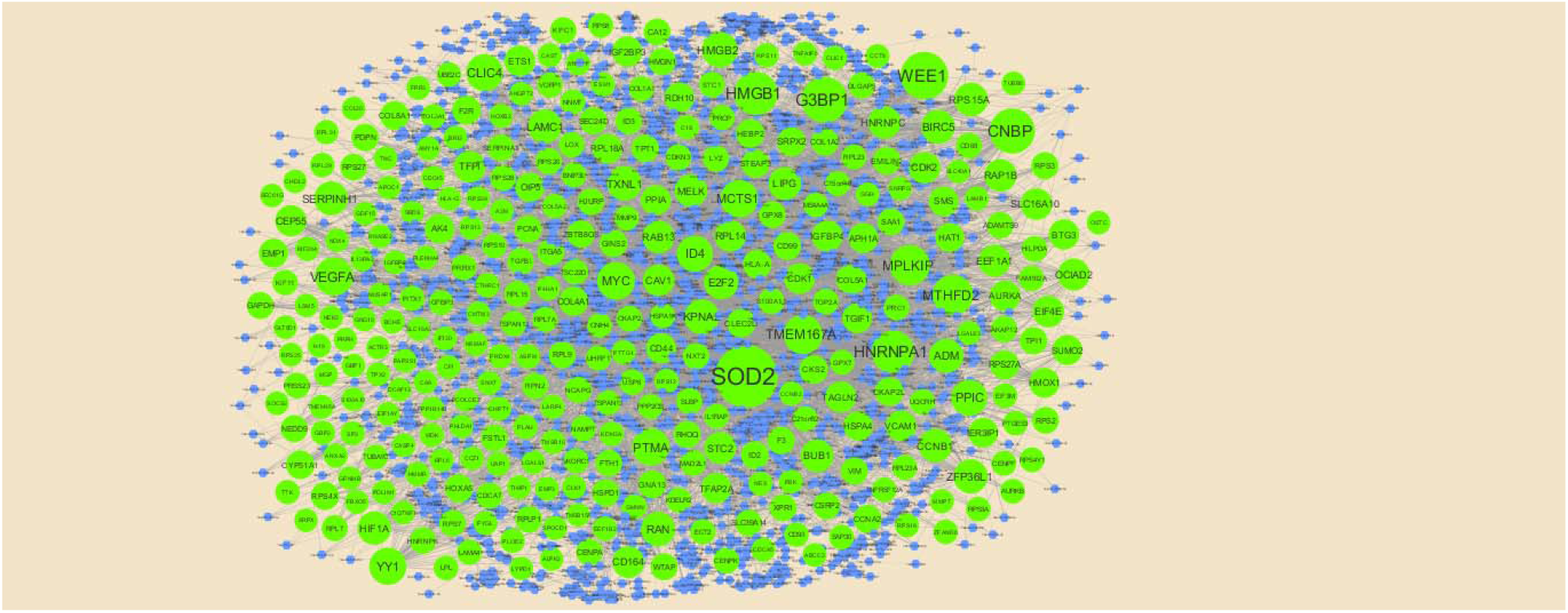
The network of up regulated genes and their related miRNAs. The green circles nodes are the up regulated genes, and blue diamond nodes are the miRNAs

**Fig. 13.**
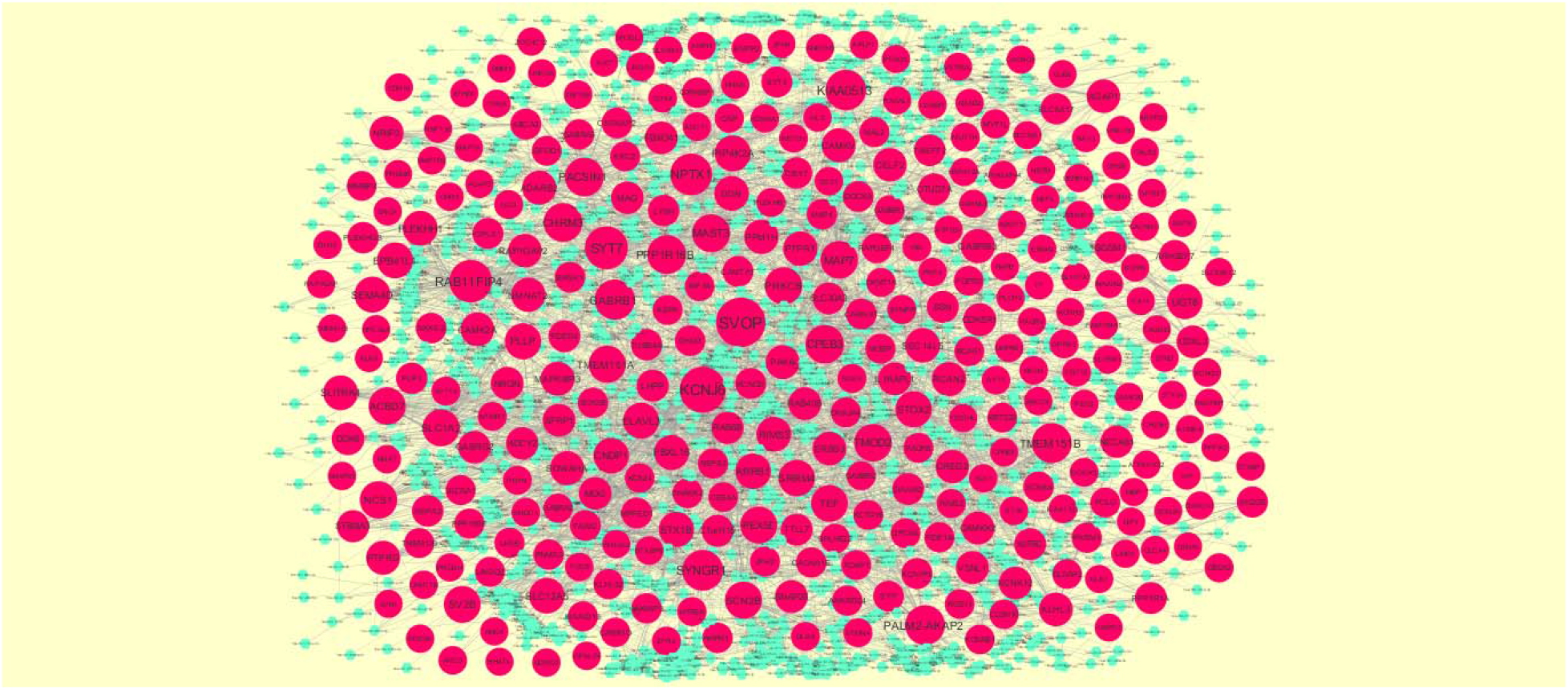
The network of down regulated genes and their related miRNAs. The pink circles nodes are the down regulated genes, and sky blue diamond nodes are the miRNAs

**Table 7.**
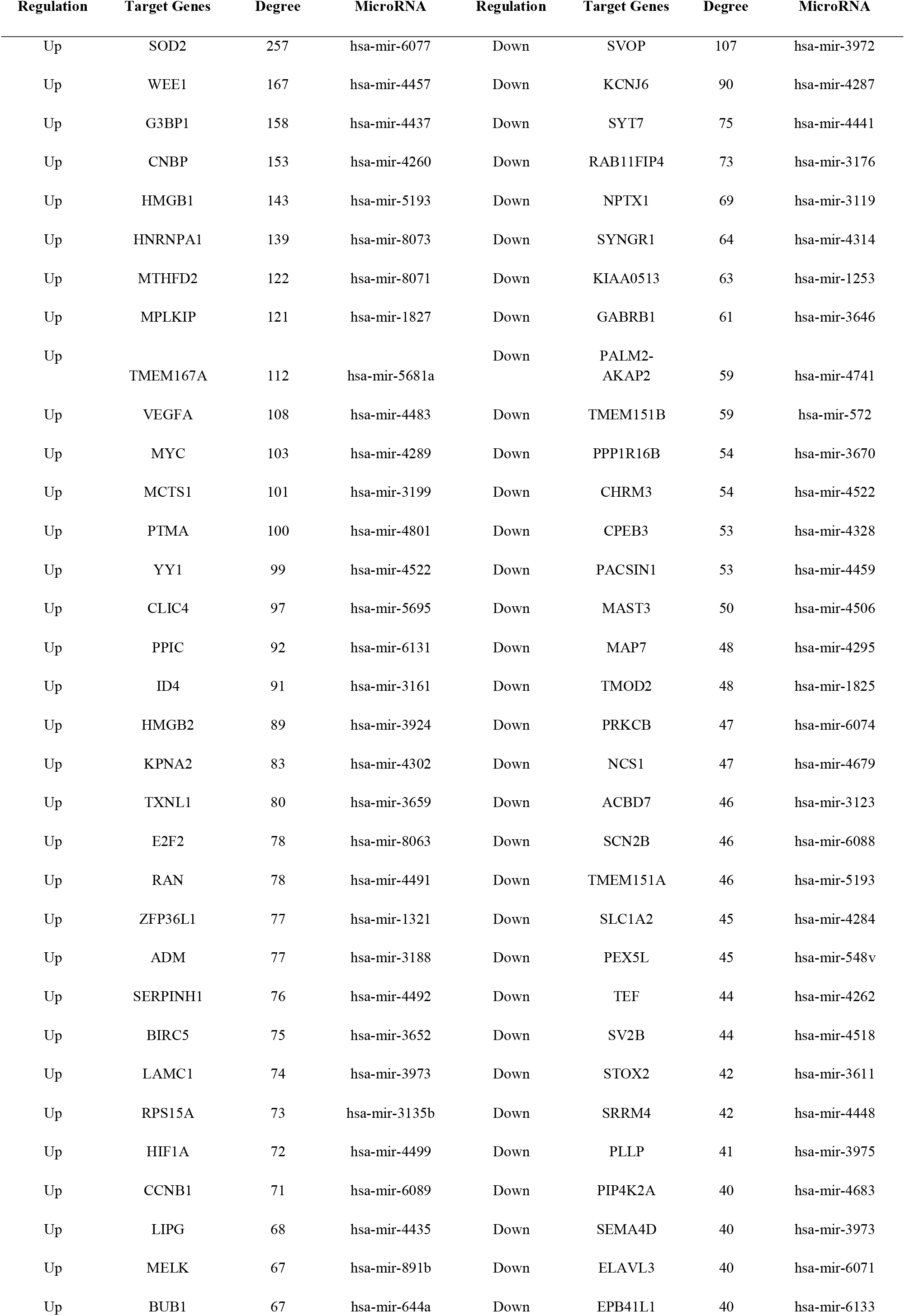
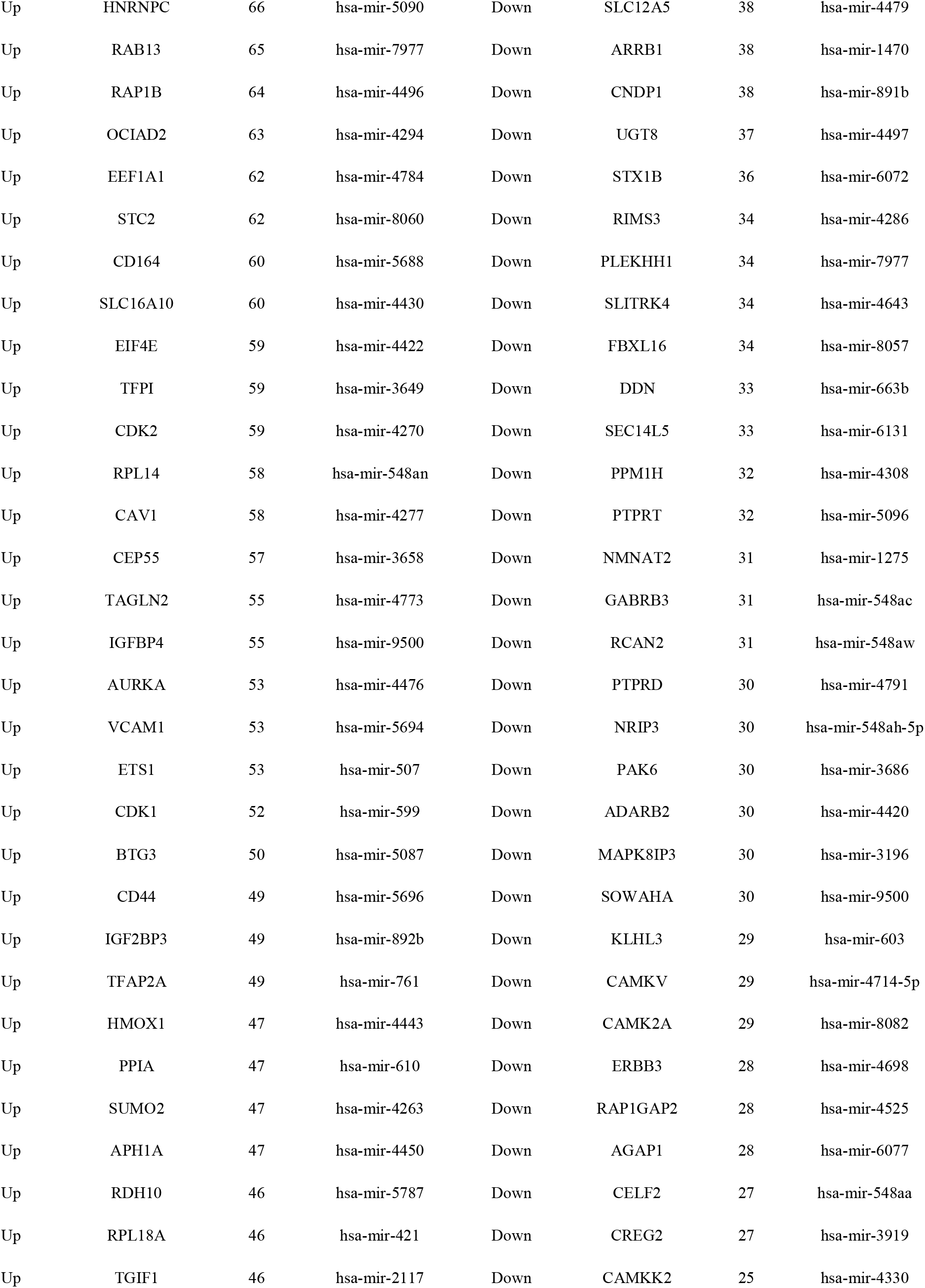
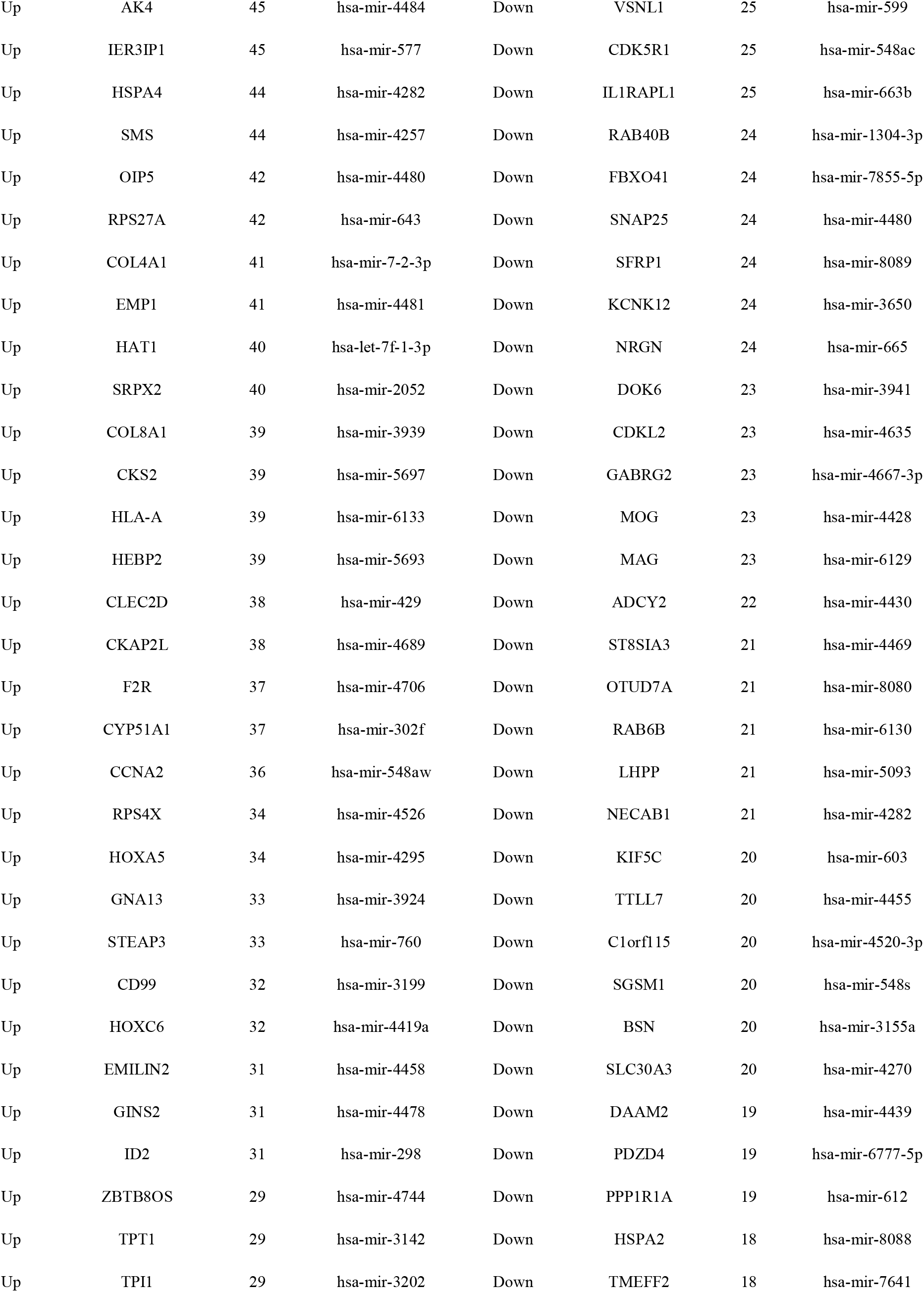
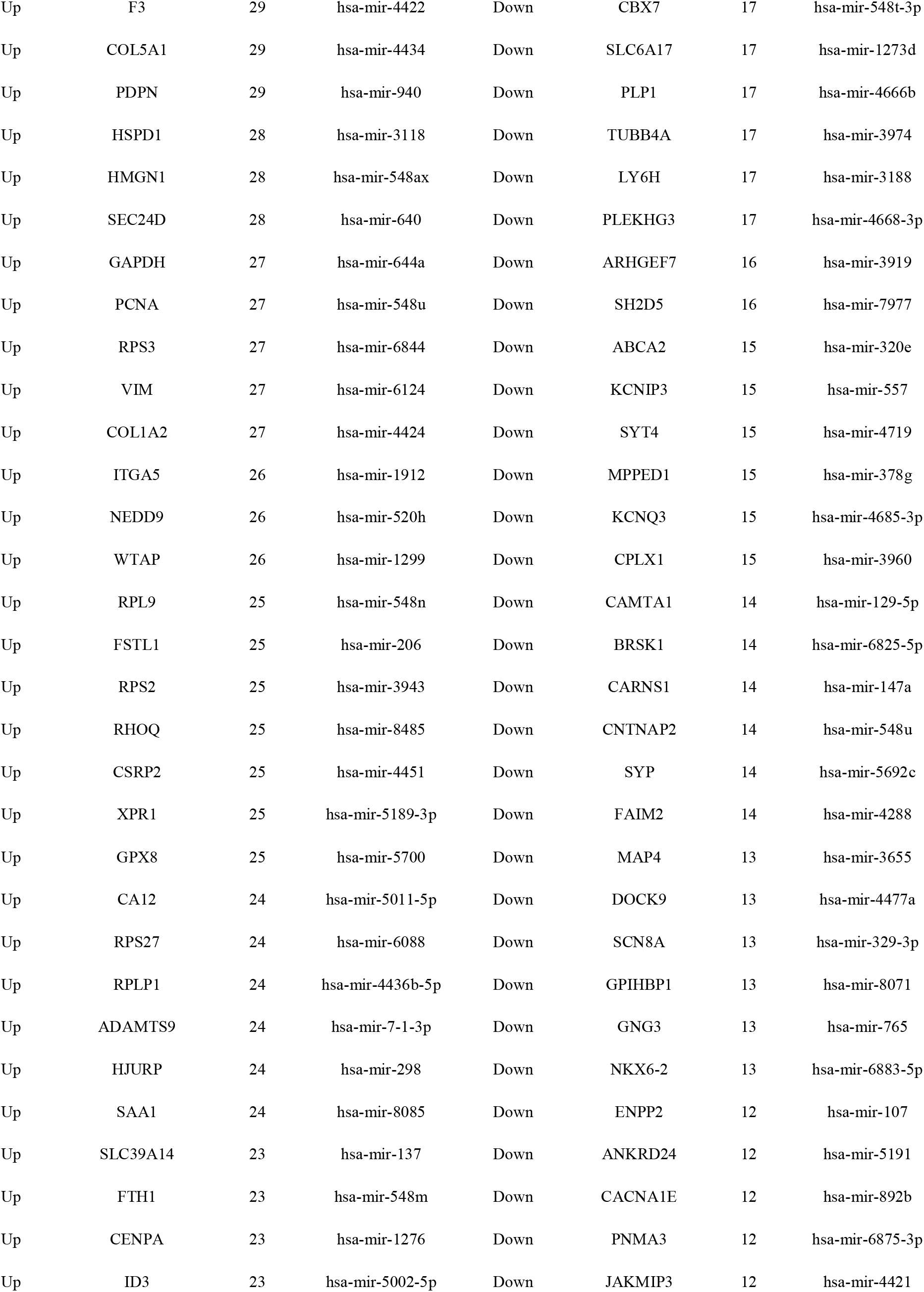
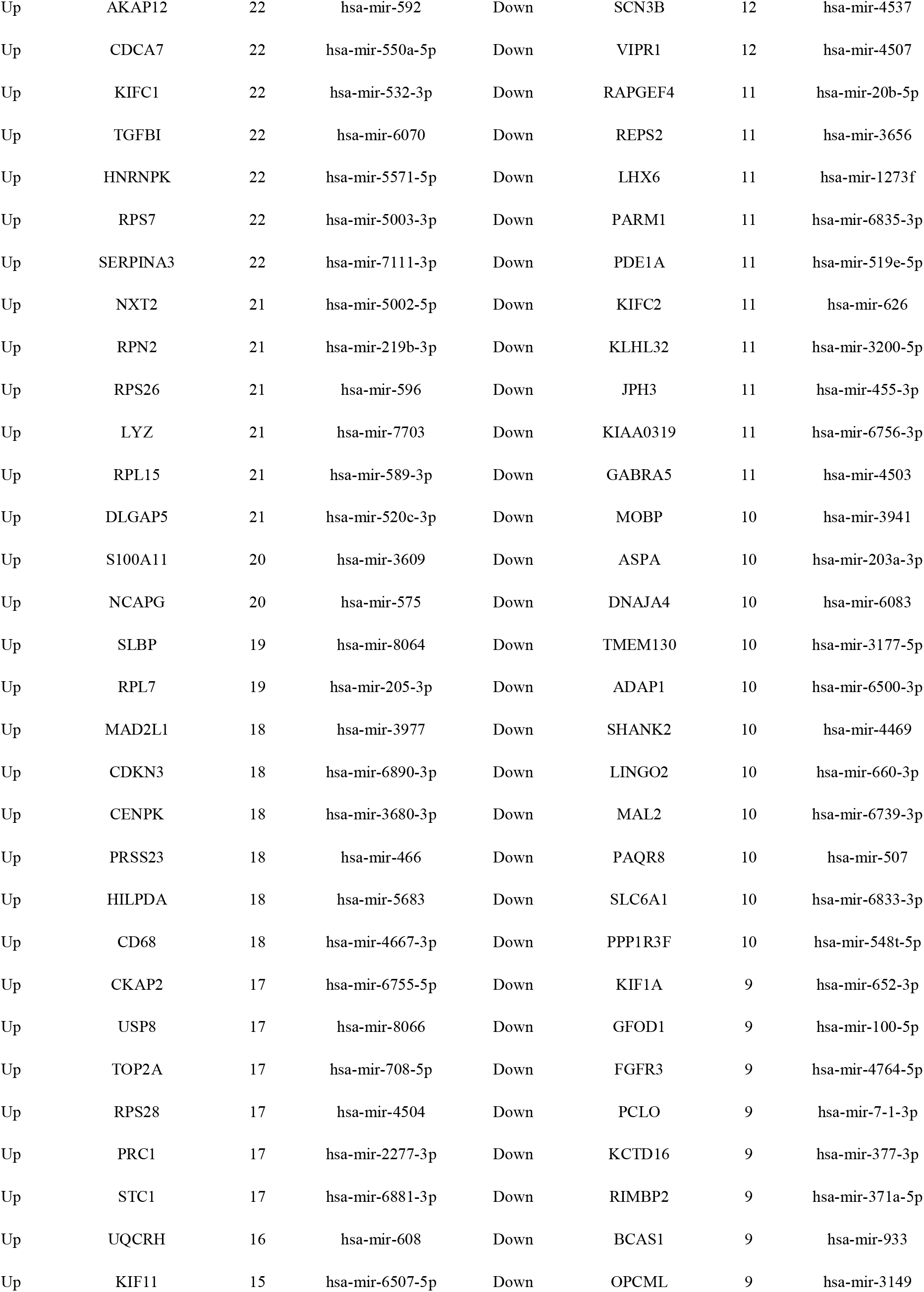
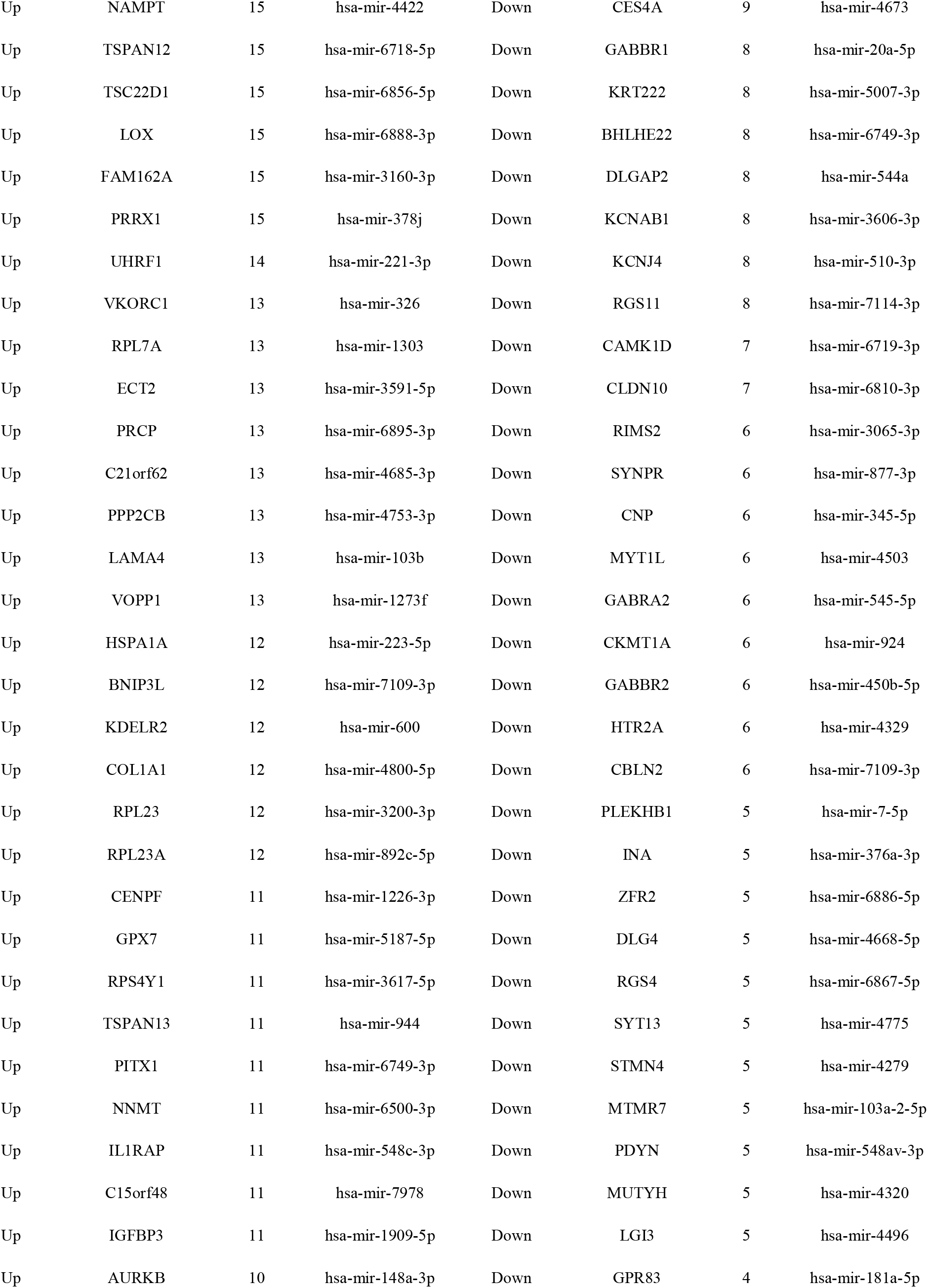
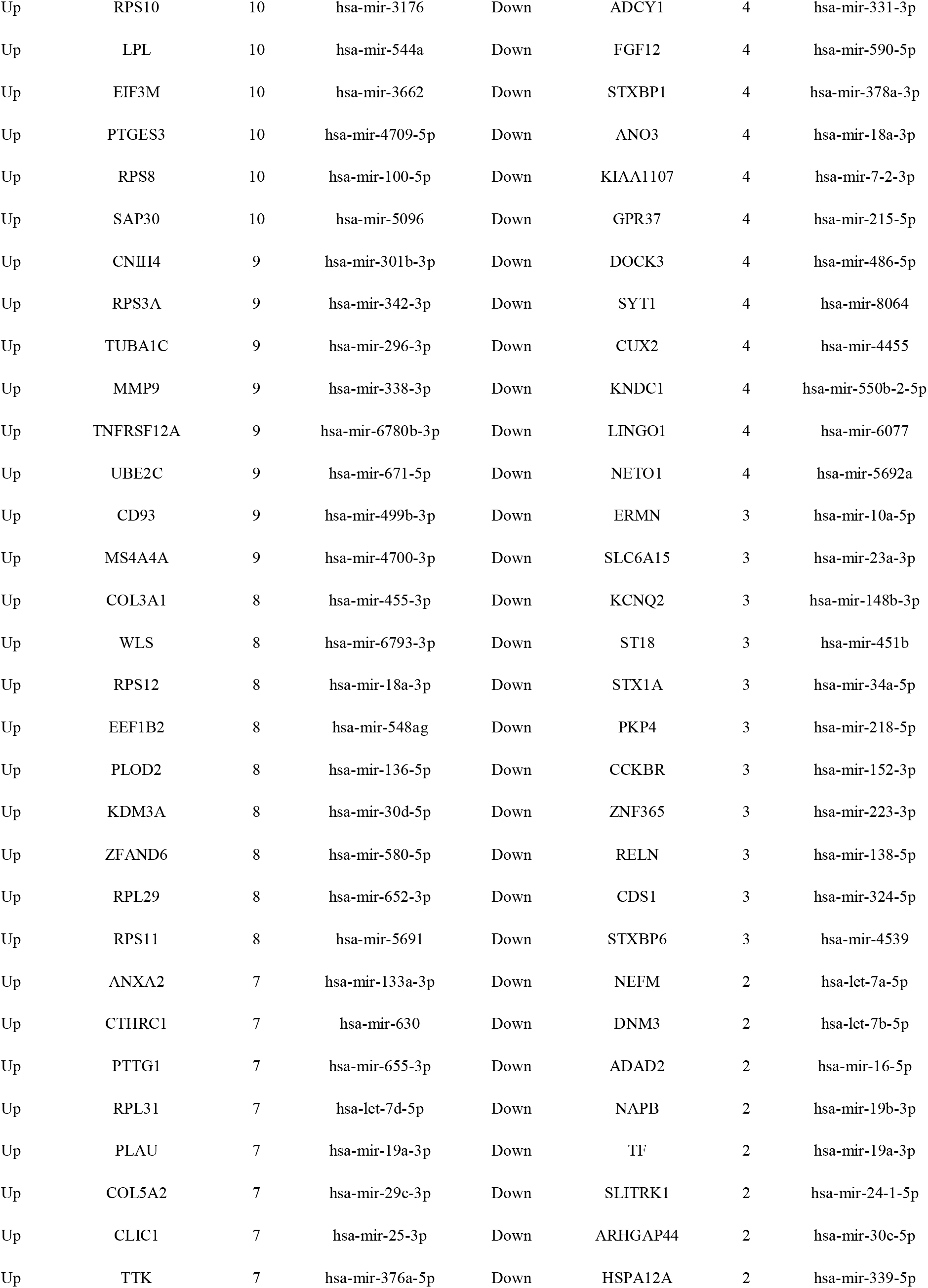
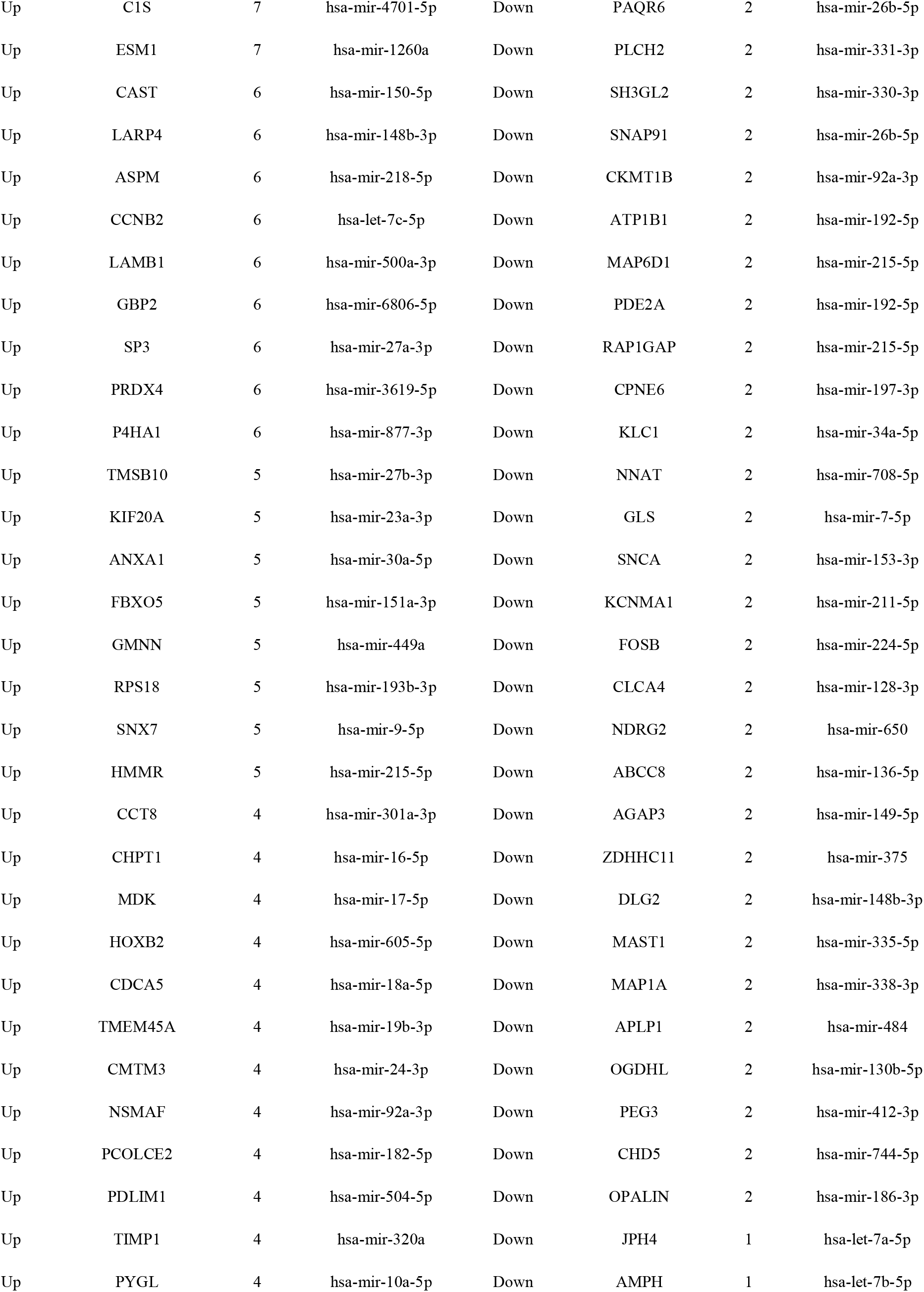
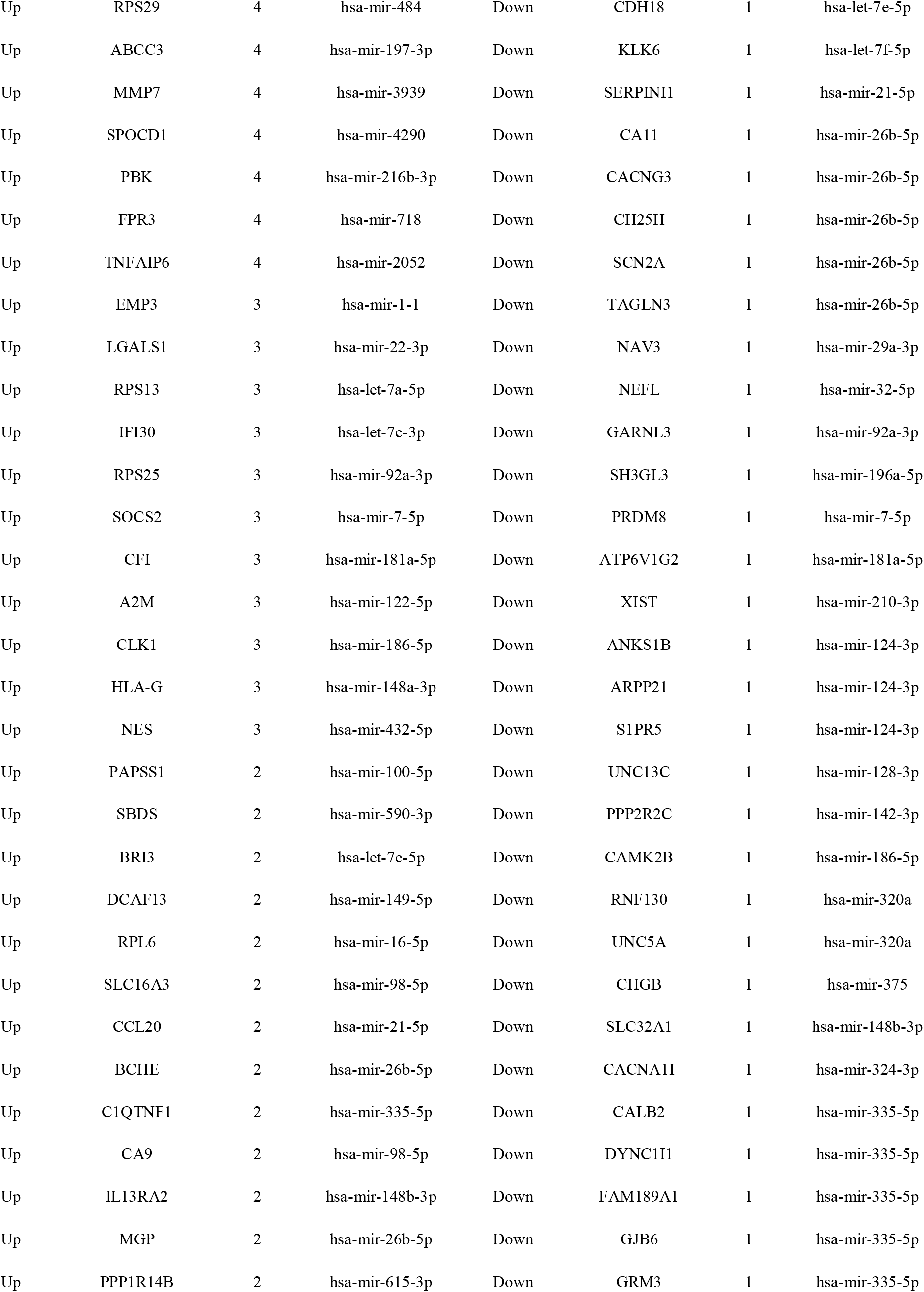
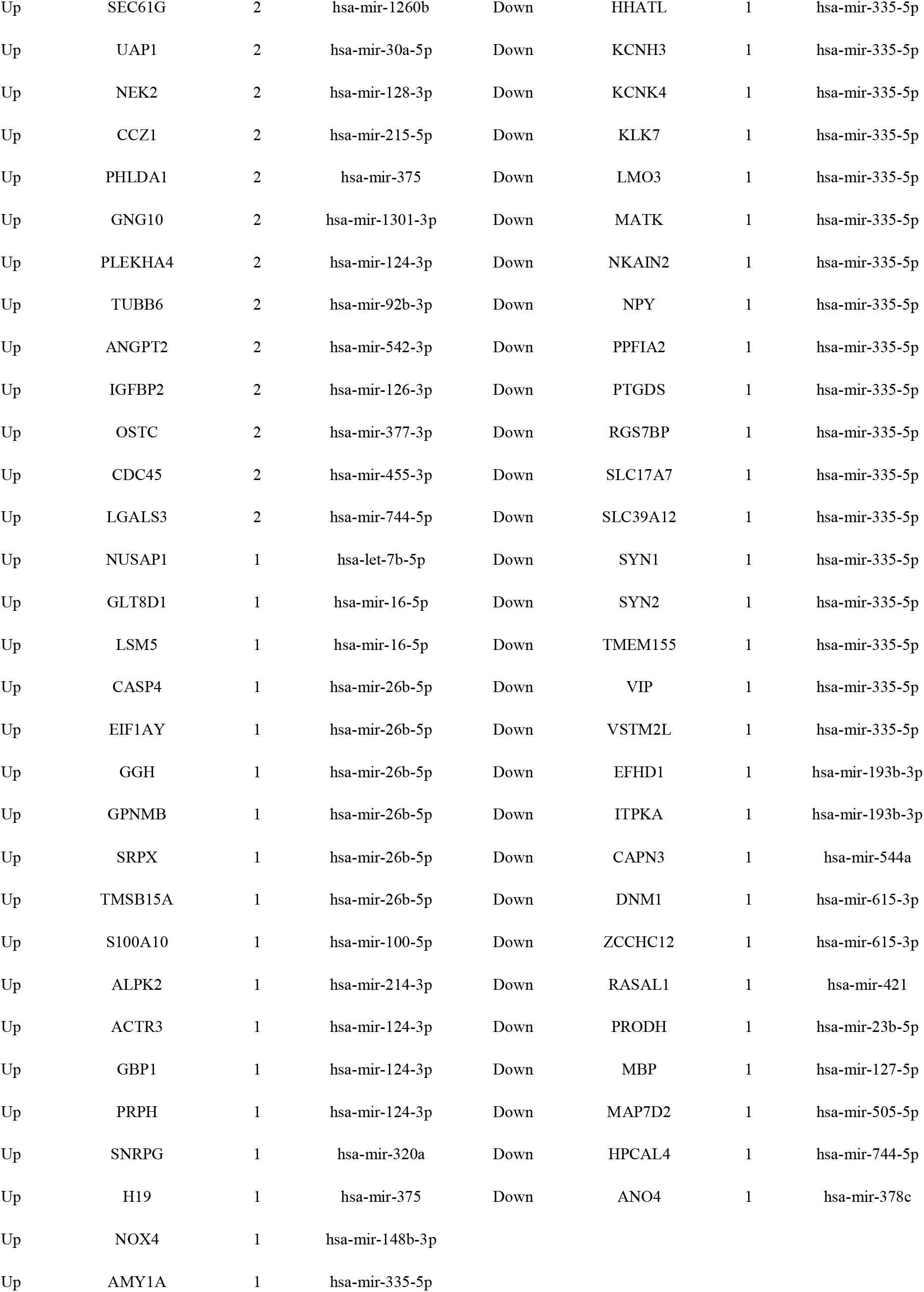
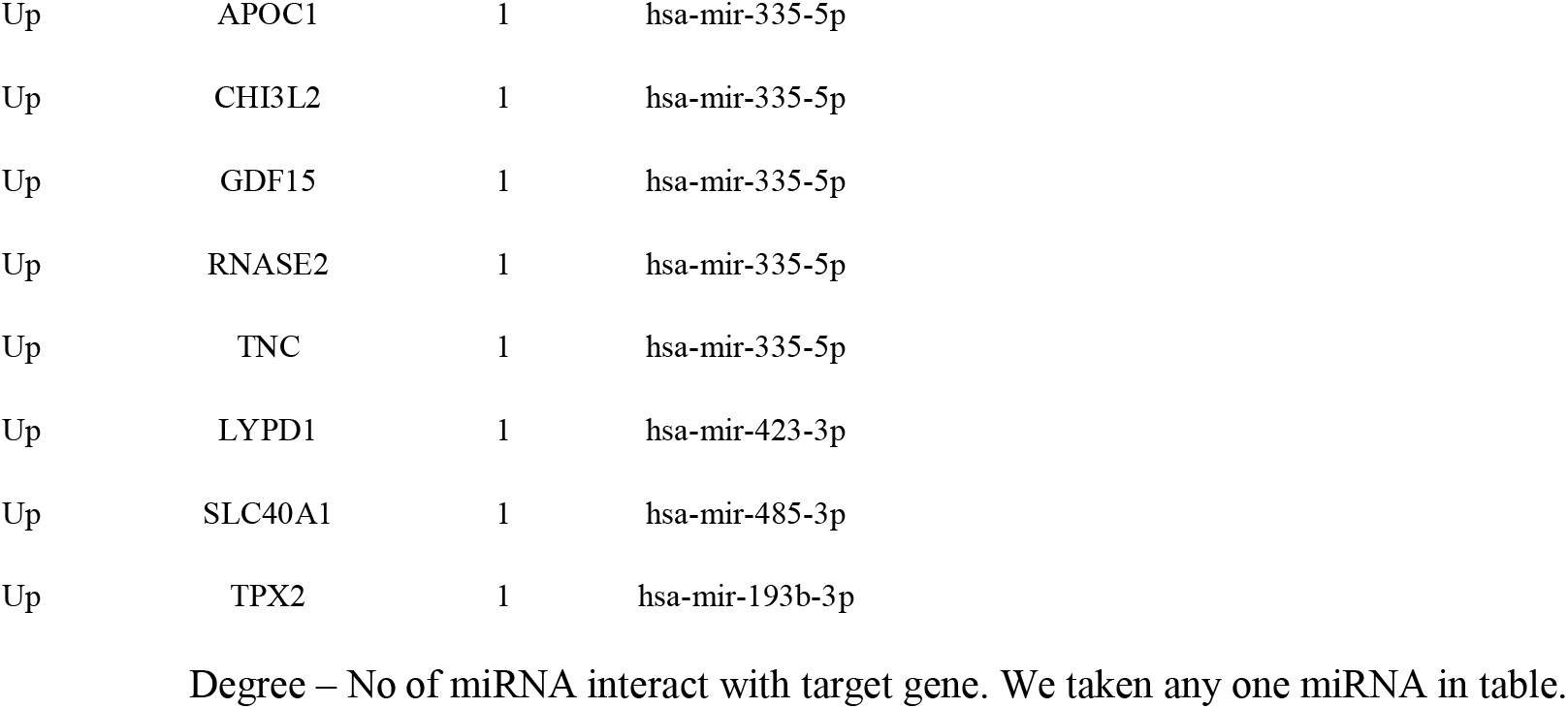
miRNA - target gene interaction table

| Regulation | Target Genes | Degree | MicroRNA | Regulation | Target Genes | Degree | MicroRNA |
| --- | --- | --- | --- | --- | --- | --- | --- |
| Up | SOD2 | 257 | hsa-mir-6077 | Down | SVOP | 107 | hsa-mir-3972 |
| Up | WEE1 | 167 | hsa-mir-4457 | Down | KCNJ6 | 90 | hsa-mir-4287 |
| Up | G3BP1 | 158 | hsa-mir-4437 | Down | SYT7 | 75 | hsa-mir-4441 |
| Up | CNBP | 153 | hsa-mir-4260 | Down | RAB11FIP4 | 73 | hsa-mir-3176 |
| Up | HMGB1 | 143 | hsa-mir-5193 | Down | NPTX1 | 69 | hsa-mir-3119 |
| Up | HNRNPA1 | 139 | hsa-mir-8073 | Down | SYNGR1 | 64 | hsa-mir-4314 |
| Up | MTHFD2 | 122 | hsa-mir-8071 | Down | KIAA0513 | 63 | hsa-mir-1253 |
| Up | MPLKIP | 121 | hsa-mir-1827 | Down | GABRB1 | 61 | hsa-mir-3646 |
| Up | TMEM167A | 112 | hsa-mir-5681a | Down | PALM2-AKAP2 | 59 | hsa-mir-4741 |
| Up | VEGFA | 108 | hsa-mir-4483 | Down | TMEM151B | 59 | hsa-mir-572 |
| Up | MYC | 103 | hsa-mir-4289 | Down | PPP1R16B | 54 | hsa-mir-3670 |
| Up | MCTS1 | 101 | hsa-mir-3199 | Down | CHRM3 | 54 | hsa-mir-4522 |
| Up | PTMA | 100 | hsa-mir-4801 | Down | CPEB3 | 53 | hsa-mir-4328 |
| Up | YY1 | 99 | hsa-mir-4522 | Down | PACSIN1 | 53 | hsa-mir-4459 |
| Up | CLIC4 | 97 | hsa-mir-5695 | Down | MAST3 | 50 | hsa-mir-4506 |
| Up | PPIC | 92 | hsa-mir-6131 | Down | MAP7 | 48 | hsa-mir-4295 |
| Up | ID4 | 91 | hsa-mir-3161 | Down | TMOD2 | 48 | hsa-mir-1825 |
| Up | HMGB2 | 89 | hsa-mir-3924 | Down | PRKCB | 47 | hsa-mir-6074 |
| Up | KPNA2 | 83 | hsa-mir-4302 | Down | NCS1 | 47 | hsa-mir-4679 |
| Up | TXNL1 | 80 | hsa-mir-3659 | Down | ACBD7 | 46 | hsa-mir-3123 |
| Up | E2F2 | 78 | hsa-mir-8063 | Down | SCN2B | 46 | hsa-mir-6088 |
| Up | RAN | 78 | hsa-mir-4491 | Down | TMEM151A | 46 | hsa-mir-5193 |
| Up | ZFP36L1 | 77 | hsa-mir-1321 | Down | SLC1A2 | 45 | hsa-mir-4284 |
| Up | ADM | 77 | hsa-mir-3188 | Down | PEX5L | 45 | hsa-mir-548v |
| Up | SERPINH1 | 76 | hsa-mir-4492 | Down | TEF | 44 | hsa-mir-4262 |
| Up | BIRC5 | 75 | hsa-mir-3652 | Down | SV2B | 44 | hsa-mir-4518 |
| Up | LAMC1 | 74 | hsa-mir-3973 | Down | STOX2 | 42 | hsa-mir-3611 |
| Up | RPS15A | 73 | hsa-mir-3135b | Down | SRRM4 | 42 | hsa-mir-4448 |
| Up | HIF1A | 72 | hsa-mir-4499 | Down | PLLP | 41 | hsa-mir-3975 |
| Up | CCNB1 | 71 | hsa-mir-6089 | Down | PIP4K2A | 40 | hsa-mir-4683 |
| Up | LIPG | 68 | hsa-mir-4435 | Down | SEMA4D | 40 | hsa-mir-3973 |
| Up | MELK | 67 | hsa-mir-891b | Down | ELAVL3 | 40 | hsa-mir-6071 |
| Up | BUB1 | 67 | hsa-mir-644a | Down | EPB41L1 | 40 | hsa-mir-6133 |
| Up | HNRNPC | 66 | hsa-mir-5090 | Down | SLC12A5 | 38 | hsa-mir-4479 |
| Up | RAB13 | 65 | hsa-mir-7977 | Down | ARRB1 | 38 | hsa-mir-1470 |
| Up | RAP1B | 64 | hsa-mir-4496 | Down | CNDP1 | 38 | hsa-mir-891b |
| Up | OCIAD2 | 63 | hsa-mir-4294 | Down | UGT8 | 37 | hsa-mir-4497 |
| Up | EEF1A1 | 62 | hsa-mir-4784 | Down | STX1B | 36 | hsa-mir-6072 |
| Up | STC2 | 62 | hsa-mir-8060 | Down | RIMS3 | 34 | hsa-mir-4286 |
| Up | CD164 | 60 | hsa-mir-5688 | Down | PLEKHH1 | 34 | hsa-mir-7977 |
| Up | SLC16A10 | 60 | hsa-mir-4430 | Down | SLITRK4 | 34 | hsa-mir-4643 |
| Up | EIF4E | 59 | hsa-mir-4422 | Down | FBXL16 | 34 | hsa-mir-8057 |
| Up | TFPI | 59 | hsa-mir-3649 | Down | DDN | 33 | hsa-mir-663b |
| Up | CDK2 | 59 | hsa-mir-4270 | Down | SEC14L5 | 33 | hsa-mir-6131 |
| Up | RPL14 | 58 | hsa-mir-548an | Down | PPM1H | 32 | hsa-mir-4308 |
| Up | CAV1 | 58 | hsa-mir-4277 | Down | PTPRT | 32 | hsa-mir-5096 |
| Up | CEP55 | 57 | hsa-mir-3658 | Down | NMNAT2 | 31 | hsa-mir-1275 |
| Up | TAGLN2 | 55 | hsa-mir-4773 | Down | GABRB3 | 31 | hsa-mir-548ac |
| Up | IGFBP4 | 55 | hsa-mir-9500 | Down | RCAN2 | 31 | hsa-mir-548aw |
| Up | AURKA | 53 | hsa-mir-4476 | Down | PTPRD | 30 | hsa-mir-4791 |
| Up | VCAM1 | 53 | hsa-mir-5694 | Down | NRIP3 | 30 | hsa-mir-548ah-5p |
| Up | ETS1 | 53 | hsa-mir-507 | Down | PAK6 | 30 | hsa-mir-3686 |
| Up | CDK1 | 52 | hsa-mir-599 | Down | ADARB2 | 30 | hsa-mir-4420 |
| Up | BTG3 | 50 | hsa-mir-5087 | Down | MAPK8IP3 | 30 | hsa-mir-3196 |
| Up | CD44 | 49 | hsa-mir-5696 | Down | SOWAHA | 30 | hsa-mir-9500 |
| Up | IGF2BP3 | 49 | hsa-mir-892b | Down | KLHL3 | 29 | hsa-mir-603 |
| Up | TFAP2A | 49 | hsa-mir-761 | Down | CAMKV | 29 | hsa-mir-4714-5p |
| Up | HMOX1 | 47 | hsa-mir-4443 | Down | CAMK2A | 29 | hsa-mir-8082 |
| Up | PPIA | 47 | hsa-mir-610 | Down | ERBB3 | 28 | hsa-mir-4698 |
| Up | SUMO2 | 47 | hsa-mir-4263 | Down | RAP1GAP2 | 28 | hsa-mir-4525 |
| Up | APH1A | 47 | hsa-mir-4450 | Down | AGAP1 | 28 | hsa-mir-6077 |
| Up | RDH10 | 46 | hsa-mir-5787 | Down | CELF2 | 27 | hsa-mir-548aa |
| Up | RPL18A | 46 | hsa-mir-421 | Down | CREG2 | 27 | hsa-mir-3919 |
| Up | TGIF1 | 46 | hsa-mir-2117 | Down | CAMKK2 | 25 | hsa-mir-4330 |
| Up | AK4 | 45 | hsa-mir-4484 | Down | VSNL1 | 25 | hsa-mir-599 |
| Up | IER3IP1 | 45 | hsa-mir-577 | Down | CDK5R1 | 25 | hsa-mir-548ac |
| Up | HSPA4 | 44 | hsa-mir-4282 | Down | IL1RAPL1 | 25 | hsa-mir-663b |
| Up | SMS | 44 | hsa-mir-4257 | Down | RAB40B | 24 | hsa-mir-1304-3p |
| Up | OIP5 | 42 | hsa-mir-4480 | Down | FBXO41 | 24 | hsa-mir-7855-5p |
| Up | RPS27A | 42 | hsa-mir-643 | Down | SNAP25 | 24 | hsa-mir-4480 |
| Up | COL4A1 | 41 | hsa-mir-7-2-3p | Down | SFRP1 | 24 | hsa-mir-8089 |
| Up | EMP1 | 41 | hsa-mir-4481 | Down | KCNK12 | 24 | hsa-mir-3650 |
| Up | HAT1 | 40 | hsa-let-7f-1-3p | Down | NRGN | 24 | hsa-mir-665 |
| Up | SRPX2 | 40 | hsa-mir-2052 | Down | DOK6 | 23 | hsa-mir-3941 |
| Up | COL8A1 | 39 | hsa-mir-3939 | Down | CDKL2 | 23 | hsa-mir-4635 |
| Up | CKS2 | 39 | hsa-mir-5697 | Down | GABRG2 | 23 | hsa-mir-4667-3p |
| Up | HLA-A | 39 | hsa-mir-6133 | Down | MOG | 23 | hsa-mir-4428 |
| Up | HEBP2 | 39 | hsa-mir-5693 | Down | MAG | 23 | hsa-mir-6129 |
| Up | CLEC2D | 38 | hsa-mir-429 | Down | ADCY2 | 22 | hsa-mir-4430 |
| Up | CKAP2L | 38 | hsa-mir-4689 | Down | ST8SIA3 | 21 | hsa-mir-4469 |
| Up | F2R | 37 | hsa-mir-4706 | Down | OTUD7A | 21 | hsa-mir-8080 |
| Up | CYP51A1 | 37 | hsa-mir-302f | Down | RAB6B | 21 | hsa-mir-6130 |
| Up | CCNA2 | 36 | hsa-mir-548aw | Down | LHPP | 21 | hsa-mir-5093 |
| Up | RPS4X | 34 | hsa-mir-4526 | Down | NECAB1 | 21 | hsa-mir-4282 |
| Up | HOXA5 | 34 | hsa-mir-4295 | Down | KIF5C | 20 | hsa-mir-603 |
| Up | GNA13 | 33 | hsa-mir-3924 | Down | TTLL7 | 20 | hsa-mir-4455 |
| Up | STEAP3 | 33 | hsa-mir-760 | Down | C1orf115 | 20 | hsa-mir-4520-3p |
| Up | CD99 | 32 | hsa-mir-3199 | Down | SGSM1 | 20 | hsa-mir-548s |
| Up | HOXC6 | 32 | hsa-mir-4419a | Down | BSN | 20 | hsa-mir-3155a |
| Up | EMILIN2 | 31 | hsa-mir-4458 | Down | SLC30A3 | 20 | hsa-mir-4270 |
| Up | GINS2 | 31 | hsa-mir-4478 | Down | DAAM2 | 19 | hsa-mir-4439 |
| Up | ID2 | 31 | hsa-mir-298 | Down | PDZD4 | 19 | hsa-mir-6777-5p |
| Up | ZBTB8OS | 29 | hsa-mir-4744 | Down | PPP1R1A | 19 | hsa-mir-612 |
| Up | TPT1 | 29 | hsa-mir-3142 | Down | HSPA2 | 18 | hsa-mir-8088 |
| Up | TPI1 | 29 | hsa-mir-3202 | Down | TMEFF2 | 18 | hsa-mir-7641 |
| Up | F3 | 29 | hsa-mir-4422 | Down | CBX7 | 17 | hsa-mir-548t-3p |
| Up | COL5A1 | 29 | hsa-mir-4434 | Down | SLC6A17 | 17 | hsa-mir-1273d |
| Up | PDPN | 29 | hsa-mir-940 | Down | PLP1 | 17 | hsa-mir-4666b |
| Up | HSPD1 | 28 | hsa-mir-3118 | Down | TUBB4A | 17 | hsa-mir-3974 |
| Up | HMGN1 | 28 | hsa-mir-548ax | Down | LY6H | 17 | hsa-mir-3188 |
| Up | SEC24D | 28 | hsa-mir-640 | Down | PLEKHG3 | 17 | hsa-mir-4668-3p |
| Up | GAPDH | 27 | hsa-mir-644a | Down | ARHGEF7 | 16 | hsa-mir-3919 |
| Up | PCNA | 27 | hsa-mir-548u | Down | SH2D5 | 16 | hsa-mir-7977 |
| Up | RPS3 | 27 | hsa-mir-6844 | Down | ABCA2 | 15 | hsa-mir-320e |
| Up | VIM | 27 | hsa-mir-6124 | Down | KCNIP3 | 15 | hsa-mir-557 |
| Up | COL1A2 | 27 | hsa-mir-4424 | Down | SYT4 | 15 | hsa-mir-4719 |
| Up | ITGA5 | 26 | hsa-mir-1912 | Down | MPPED1 | 15 | hsa-mir-378g |
| Up | NEDD9 | 26 | hsa-mir-520h | Down | KCNQ3 | 15 | hsa-mir-4685-3p |
| Up | WTAP | 26 | hsa-mir-1299 | Down | CPLX1 | 15 | hsa-mir-3960 |
| Up | RPL9 | 25 | hsa-mir-548n | Down | CAMTA1 | 14 | hsa-mir-129-5p |
| Up | FSTL1 | 25 | hsa-mir-206 | Down | BRSK1 | 14 | hsa-mir-6825-5p |
| Up | RPS2 | 25 | hsa-mir-3943 | Down | CARNS1 | 14 | hsa-mir-147a |
| Up | RHOQ | 25 | hsa-mir-8485 | Down | CNTNAP2 | 14 | hsa-mir-548u |
| Up | CSRP2 | 25 | hsa-mir-4451 | Down | SYP | 14 | hsa-mir-5692c |
| Up | XPR1 | 25 | hsa-mir-5189-3p | Down | FAIM2 | 14 | hsa-mir-4288 |
| Up | GPX8 | 25 | hsa-mir-5700 | Down | MAP4 | 13 | hsa-mir-3655 |
| Up | CA12 | 24 | hsa-mir-5011-5p | Down | DOCK9 | 13 | hsa-mir-4477a |
| Up | RPS27 | 24 | hsa-mir-6088 | Down | SCN8A | 13 | hsa-mir-329-3p |
| Up | RPLP1 | 24 | hsa-mir-4436b-5p | Down | GPIHBP1 | 13 | hsa-mir-8071 |
| Up | ADAMTS9 | 24 | hsa-mir-7-1-3p | Down | GNG3 | 13 | hsa-mir-765 |
| Up | HJURP | 24 | hsa-mir-298 | Down | NKX6-2 | 13 | hsa-mir-6883-5p |
| Up | SAA1 | 24 | hsa-mir-8085 | Down | ENPP2 | 12 | hsa-mir-107 |
| Up | SLC39A14 | 23 | hsa-mir-137 | Down | ANKRD24 | 12 | hsa-mir-5191 |
| Up | FTH1 | 23 | hsa-mir-548m | Down | CACNA1E | 12 | hsa-mir-892b |
| Up | CENPA | 23 | hsa-mir-1276 | Down | PNMA3 | 12 | hsa-mir-6875-3p |
| Up | ID3 | 23 | hsa-mir-5002-5p | Down | JAKMIP3 | 12 | hsa-mir-4421 |
| Up | AKAP12 | 22 | hsa-mir-592 | Down | SCN3B | 12 | hsa-mir-4537 |
| Up | CDCA7 | 22 | hsa-mir-550a-5p | Down | VIPR1 | 12 | hsa-mir-4507 |
| Up | KIFC1 | 22 | hsa-mir-532-3p | Down | RAPGEF4 | 11 | hsa-mir-20b-5p |
| Up | TGFB1 | 22 | hsa-mir-6070 | Down | REPS2 | 11 | hsa-mir-3656 |
| Up | HNRNPK | 22 | hsa-mir-5571-5p | Down | LHX6 | 11 | hsa-mir-1273f |
| Up | RPS7 | 22 | hsa-mir-5003-3p | Down | PARM1 | 11 | hsa-mir-6835-3p |
| Up | SERPINA3 | 22 | hsa-mir-7111-3p | Down | PDE1A | 11 | hsa-mir-519e-5p |
| Up | NXT2 | 21 | hsa-mir-5002-5p | Down | KIFC2 | 11 | hsa-mir-626 |
| Up | RPN2 | 21 | hsa-mir-219b-3p | Down | KLHL32 | 11 | hsa-mir-3200-5p |
| Up | RPS26 | 21 | hsa-mir-596 | Down | JPH3 | 11 | hsa-mir-455-3p |
| Up | LYZ | 21 | hsa-mir-7703 | Down | KIAA0319 | 11 | hsa-mir-6756-3p |
| Up | RPL15 | 21 | hsa-mir-589-3p | Down | GABRA5 | 11 | hsa-mir-4503 |
| Up | DLGAP5 | 21 | hsa-mir-520c-3p | Down | MOBP | 10 | hsa-mir-3941 |
| Up | S100A11 | 20 | hsa-mir-3609 | Down | ASPA | 10 | hsa-mir-203a-3p |
| Up | NCAPG | 20 | hsa-mir-575 | Down | DNAJA4 | 10 | hsa-mir-6083 |
| Up | SLBP | 19 | hsa-mir-8064 | Down | TMEM130 | 10 | hsa-mir-3177-5p |
| Up | RPL7 | 19 | hsa-mir-205-3p | Down | ADAP1 | 10 | hsa-mir-6500-3p |
| Up | MAD2L1 | 18 | hsa-mir-3977 | Down | SHANK2 | 10 | hsa-mir-4469 |
| Up | CDKN3 | 18 | hsa-mir-6890-3p | Down | LINGO2 | 10 | hsa-mir-660-3p |
| Up | CENPK | 18 | hsa-mir-3680-3p | Down | MAL2 | 10 | hsa-mir-6739-3p |
| Up | PRSS23 | 18 | hsa-mir-466 | Down | PAQR8 | 10 | hsa-mir-507 |
| Up | HILPDA | 18 | hsa-mir-5683 | Down | SLC6A1 | 10 | hsa-mir-6833-3p |
| Up | CD68 | 18 | hsa-mir-4667-3p | Down | PPP1R3F | 10 | hsa-mir-548t-5p |
| Up | CKAP2 | 17 | hsa-mir-6755-5p | Down | KIF1A | 9 | hsa-mir-652-3p |
| Up | USP8 | 17 | hsa-mir-8066 | Down | GFOD1 | 9 | hsa-mir-100-5p |
| Up | TOP2A | 17 | hsa-mir-708-5p | Down | FGFR3 | 9 | hsa-mir-4764-5p |
| Up | RPS28 | 17 | hsa-mir-4504 | Down | PCLO | 9 | hsa-mir-7-1-3p |
| Up | PRC1 | 17 | hsa-mir-2277-3p | Down | KCTD16 | 9 | hsa-mir-377-3p |
| Up | STC1 | 17 | hsa-mir-6881-3p | Down | RIMBP2 | 9 | hsa-mir-371a-5p |
| Up | UQCRH | 16 | hsa-mir-608 | Down | BCAS1 | 9 | hsa-mir-933 |
| Up | KIF11 | 15 | hsa-mir-6507-5p | Down | OPCML | 9 | hsa-mir-3149 |
| Up | NAMPT | 15 | hsa-mir-4422 | Down | CES4A | 9 | hsa-mir-4673 |
| Up | TSPAN12 | 15 | hsa-mir-6718-5p | Down | GABBR1 | 8 | hsa-mir-20a-5p |
| Up | TSC22D1 | 15 | hsa-mir-6856-5p | Down | KRT222 | 8 | hsa-mir-5007-3p |
| Up | LOX | 15 | hsa-mir-6888-3p | Down | BHLHE22 | 8 | hsa-mir-6749-3p |
| Up | FAM162A | 15 | hsa-mir-3160-3p | Down | DLGAP2 | 8 | hsa-mir-544a |
| Up | PRRX1 | 15 | hsa-mir-378j | Down | KCNAB1 | 8 | hsa-mir-3606-3p |
| Up | UHRF1 | 14 | hsa-mir-221-3p | Down | KCNJ4 | 8 | hsa-mir-510-3p |
| Up | VKORC1 | 13 | hsa-mir-326 | Down | RGS11 | 8 | hsa-mir-7114-3p |
| Up | RPL7A | 13 | hsa-mir-1303 | Down | CAMK1D | 7 | hsa-mir-6719-3p |
| Up | ECT2 | 13 | hsa-mir-3591-5p | Down | CLDN10 | 7 | hsa-mir-6810-3p |
| Up | PRCP | 13 | hsa-mir-6895-3p | Down | RIMS2 | 6 | hsa-mir-3065-3p |
| Up | C21orf62 | 13 | hsa-mir-4685-3p | Down | SYNPR | 6 | hsa-mir-877-3p |
| Up | PPP2CB | 13 | hsa-mir-4753-3p | Down | CNP | 6 | hsa-mir-345-5p |
| Up | LAMA4 | 13 | hsa-mir-103b | Down | MYT1L | 6 | hsa-mir-4503 |
| Up | VOPP1 | 13 | hsa-mir-1273f | Down | GABRA2 | 6 | hsa-mir-545-5p |
| Up | HSPA1A | 12 | hsa-mir-223-5p | Down | CKMT1A | 6 | hsa-mir-924 |
| Up | BNIP3L | 12 | hsa-mir-7109-3p | Down | GABBR2 | 6 | hsa-mir-450b-5p |
| Up | KDELR2 | 12 | hsa-mir-600 | Down | HTR2A | 6 | hsa-mir-4329 |
| Up | COL1A1 | 12 | hsa-mir-4800-5p | Down | CBLN2 | 6 | hsa-mir-7109-3p |
| Up | RPL23 | 12 | hsa-mir-3200-3p | Down | PLEKHB1 | 5 | hsa-mir-7-5p |
| Up | RPL23A | 12 | hsa-mir-892c-5p | Down | INA | 5 | hsa-mir-376a-3p |
| Up | CENPF | 11 | hsa-mir-1226-3p | Down | ZFR2 | 5 | hsa-mir-6886-5p |
| Up | GPX7 | 11 | hsa-mir-5187-5p | Down | DLG4 | 5 | hsa-mir-4668-5p |
| Up | RPS4Y1 | 11 | hsa-mir-3617-5p | Down | RGS4 | 5 | hsa-mir-6867-5p |
| Up | TSPAN13 | 11 | hsa-mir-944 | Down | SYT13 | 5 | hsa-mir-4775 |
| Up | PITX1 | 11 | hsa-mir-6749-3p | Down | STMN4 | 5 | hsa-mir-4279 |
| Up | NNMT | 11 | hsa-mir-6500-3p | Down | MTMR7 | 5 | hsa-mir-103a-2-5p |
| Up | IL1RAP | 11 | hsa-mir-548c-3p | Down | PDYN | 5 | hsa-mir-548av-3p |
| Up | C15orf48 | 11 | hsa-mir-7978 | Down | MUTYH | 5 | hsa-mir-4320 |
| Up | IGFBP3 | 11 | hsa-mir-1909-5p | Down | LGI3 | 5 | hsa-mir-4496 |
| Up | AURKB | 10 | hsa-mir-148a-3p | Down | GPR83 | 4 | hsa-mir-181a-5p |
| Up | RPS10 | 10 | hsa-mir-3176 | Down | ADCY1 | 4 | hsa-mir-331-3p |
| Up | LPL | 10 | hsa-mir-544a | Down | FGF12 | 4 | hsa-mir-590-5p |
| Up | EIF3M | 10 | hsa-mir-3662 | Down | STXBP1 | 4 | hsa-mir-378a-3p |
| Up | PTGES3 | 10 | hsa-mir-4709-5p | Down | ANO3 | 4 | hsa-mir-18a-3p |
| Up | RPS8 | 10 | hsa-mir-100-5p | Down | KIAA1107 | 4 | hsa-mir-7-2-3p |
| Up | SAP30 | 10 | hsa-mir-5096 | Down | GPR37 | 4 | hsa-mir-215-5p |
| Up | CNIH4 | 9 | hsa-mir-301b-3p | Down | DOCK3 | 4 | hsa-mir-486-5p |
| Up | RPS3A | 9 | hsa-mir-342-3p | Down | SYT1 | 4 | hsa-mir-8064 |
| Up | TUBA1C | 9 | hsa-mir-296-3p | Down | CUX2 | 4 | hsa-mir-4455 |
| Up | MMP9 | 9 | hsa-mir-338-3p | Down | KNDC1 | 4 | hsa-mir-550b-2-5p |
| Up | TNFRSF12A | 9 | hsa-mir-6780b-3p | Down | LINGO1 | 4 | hsa-mir-6077 |
| Up | UBE2C | 9 | hsa-mir-671-5p | Down | NETO1 | 4 | hsa-mir-5692a |
| Up | CD93 | 9 | hsa-mir-499b-3p | Down | ERMN | 3 | hsa-mir-10a-5p |
| Up | MS4A4A | 9 | hsa-mir-4700-3p | Down | SLC6A15 | 3 | hsa-mir-23a-3p |
| Up | COL3A1 | 8 | hsa-mir-455-3p | Down | KCNQ2 | 3 | hsa-mir-148b-3p |
| Up | WLS | 8 | hsa-mir-6793-3p | Down | ST18 | 3 | hsa-mir-451b |
| Up | RPS12 | 8 | hsa-mir-18a-3p | Down | STX1A | 3 | hsa-mir-34a-5p |
| Up | EEF1B2 | 8 | hsa-mir-548ag | Down | PKP4 | 3 | hsa-mir-218-5p |
| Up | PLOD2 | 8 | hsa-mir-136-5p | Down | CCKBR | 3 | hsa-mir-152-3p |
| Up | KDM3A | 8 | hsa-mir-30d-5p | Down | ZNF365 | 3 | hsa-mir-223-3p |
| Up | ZFAND6 | 8 | hsa-mir-580-5p | Down | RELN | 3 | hsa-mir-138-5p |
| Up | RPL29 | 8 | hsa-mir-652-3p | Down | CDS1 | 3 | hsa-mir-324-5p |
| Up | RPS11 | 8 | hsa-mir-5691 | Down | STXBP6 | 3 | hsa-mir-4539 |
| Up | ANXA2 | 7 | hsa-mir-133a-3p | Down | NEFM | 2 | hsa-let-7a-5p |
| Up | CTHRC1 | 7 | hsa-mir-630 | Down | DNM3 | 2 | hsa-let-7b-5p |
| Up | PTTG1 | 7 | hsa-mir-655-3p | Down | ADAD2 | 2 | hsa-mir-16-5p |
| Up | RPL31 | 7 | hsa-let-7d-5p | Down | NAPB | 2 | hsa-mir-19b-3p |
| Up | PLAU | 7 | hsa-mir-19a-3p | Down | TF | 2 | hsa-mir-19a-3p |
| Up | COL5A2 | 7 | hsa-mir-29c-3p | Down | SLITRK1 | 2 | hsa-mir-24-1-5p |
| Up | CLIC1 | 7 | hsa-mir-25-3p | Down | ARHGAP44 | 2 | hsa-mir-30c-5p |
| Up | TTK | 7 | hsa-mir-376a-5p | Down | HSPA12A | 2 | hsa-mir-339-5p |
| Up | C1S | 7 | hsa-mir-4701-5p | Down | PAQR6 | 2 | hsa-mir-26b-5p |
| Up | ESM1 | 7 | hsa-mir-1260a | Down | PLCH2 | 2 | hsa-mir-331-3p |
| Up | CAST | 6 | hsa-mir-150-5p | Down | SH3GL2 | 2 | hsa-mir-330-3p |
| Up | LARP4 | 6 | hsa-mir-148b-3p | Down | SNAP91 | 2 | hsa-mir-26b-5p |
| Up | ASPM | 6 | hsa-mir-218-5p | Down | CKMT1B | 2 | hsa-mir-92a-3p |
| Up | CCNB2 | 6 | hsa-let-7c-5p | Down | ATP1B1 | 2 | hsa-mir-192-5p |
| Up | LAMB1 | 6 | hsa-mir-500a-3p | Down | MAP6D1 | 2 | hsa-mir-215-5p |
| Up | GBP2 | 6 | hsa-mir-6806-5p | Down | PDE2A | 2 | hsa-mir-192-5p |
| Up | SP3 | 6 | hsa-mir-27a-3p | Down | RAP1GAP | 2 | hsa-mir-215-5p |
| Up | PRDX4 | 6 | hsa-mir-3619-5p | Down | CPNE6 | 2 | hsa-mir-197-3p |
| Up | P4HA1 | 6 | hsa-mir-877-3p | Down | KLC1 | 2 | hsa-mir-34a-5p |
| Up | TMSB10 | 5 | hsa-mir-27b-3p | Down | NNAT | 2 | hsa-mir-708-5p |
| Up | KIF20A | 5 | hsa-mir-23a-3p | Down | GLS | 2 | hsa-mir-7-5p |
| Up | ANXA1 | 5 | hsa-mir-30a-5p | Down | SNCA | 2 | hsa-mir-153-3p |
| Up | FBXO5 | 5 | hsa-mir-151a-3p | Down | KCNMA1 | 2 | hsa-mir-211-5p |
| Up | GMNN | 5 | hsa-mir-449a | Down | FOSB | 2 | hsa-mir-224-5p |
| Up | RPS18 | 5 | hsa-mir-193b-3p | Down | CLCA4 | 2 | hsa-mir-128-3p |
| Up | SNX7 | 5 | hsa-mir-9-5p | Down | NDRG2 | 2 | hsa-mir-650 |
| Up | HMMR | 5 | hsa-mir-215-5p | Down | ABCC8 | 2 | hsa-mir-136-5p |
| Up | CCT8 | 4 | hsa-mir-301a-3p | Down | AGAP3 | 2 | hsa-mir-149-5p |
| Up | CHPT1 | 4 | hsa-mir-16-5p | Down | ZDHHC11 | 2 | hsa-mir-375 |
| Up | MDK | 4 | hsa-mir-17-5p | Down | DLG2 | 2 | hsa-mir-148b-3p |
| Up | HOXB2 | 4 | hsa-mir-605-5p | Down | MAST1 | 2 | hsa-mir-335-5p |
| Up | CDCA5 | 4 | hsa-mir-18a-5p | Down | MAP1A | 2 | hsa-mir-338-3p |
| Up | TMEM45A | 4 | hsa-mir-19b-3p | Down | APLP1 | 2 | hsa-mir-484 |
| Up | CMTM3 | 4 | hsa-mir-24-3p | Down | OGDHL | 2 | hsa-mir-130b-5p |
| Up | NSMAF | 4 | hsa-mir-92a-3p | Down | PEG3 | 2 | hsa-mir-412-3p |
| Up | PCOLCE2 | 4 | hsa-mir-182-5p | Down | CHD5 | 2 | hsa-mir-744-5p |
| Up | PDLIM1 | 4 | hsa-mir-504-5p | Down | OPALIN | 2 | hsa-mir-186-3p |
| Up | TIMP1 | 4 | hsa-mir-320a | Down | JPH4 | 1 | hsa-let-7a-5p |
| Up | PYGL | 4 | hsa-mir-10a-5p | Down | AMPH | 1 | hsa-let-7b-5p |
| Up | RPS29 | 4 | hsa-mir-484 | Down | CDH18 | 1 | hsa-let-7e-5p |
| Up | ABCC3 | 4 | hsa-mir-197-3p | Down | KLK6 | 1 | hsa-let-7f-5p |
| Up | MMP7 | 4 | hsa-mir-3939 | Down | SERPINI1 | 1 | hsa-mir-21-5p |
| Up | SPOCD1 | 4 | hsa-mir-4290 | Down | CA11 | 1 | hsa-mir-26b-5p |
| Up | PBK | 4 | hsa-mir-216b-3p | Down | CACNG3 | 1 | hsa-mir-26b-5p |
| Up | FPR3 | 4 | hsa-mir-718 | Down | CH25H | 1 | hsa-mir-26b-5p |
| Up | TNFAIP6 | 4 | hsa-mir-2052 | Down | SCN2A | 1 | hsa-mir-26b-5p |
| Up | EMP3 | 3 | hsa-mir-1-1 | Down | TAGLN3 | 1 | hsa-mir-26b-5p |
| Up | LGALS1 | 3 | hsa-mir-22-3p | Down | NAV3 | 1 | hsa-mir-29a-3p |
| Up | RPS13 | 3 | hsa-let-7a-5p | Down | NEFL | 1 | hsa-mir-32-5p |
| Up | IFI30 | 3 | hsa-let-7c-3p | Down | GARNL3 | 1 | hsa-mir-92a-3p |
| Up | RPS25 | 3 | hsa-mir-92a-3p | Down | SH3GL3 | 1 | hsa-mir-196a-5p |
| Up | SOCS2 | 3 | hsa-mir-7-5p | Down | PRDM8 | 1 | hsa-mir-7-5p |
| Up | CFI | 3 | hsa-mir-181a-5p | Down | ATP6V1G2 | 1 | hsa-mir-181a-5p |
| Up | A2M | 3 | hsa-mir-122-5p | Down | XIST | 1 | hsa-mir-210-3p |
| Up | CLK1 | 3 | hsa-mir-186-5p | Down | ANKS1B | 1 | hsa-mir-124-3p |
| Up | HLA-G | 3 | hsa-mir-148a-3p | Down | ARPP21 | 1 | hsa-mir-124-3p |
| Up | NES | 3 | hsa-mir-432-5p | Down | S1PR5 | 1 | hsa-mir-124-3p |
| Up | PAPSS1 | 2 | hsa-mir-100-5p | Down | UNC13C | 1 | hsa-mir-128-3p |
| Up | SBDS | 2 | hsa-mir-590-3p | Down | PPP2R2C | 1 | hsa-mir-142-3p |
| Up | BRI3 | 2 | hsa-let-7e-5p | Down | CAMK2B | 1 | hsa-mir-186-5p |
| Up | DCAF13 | 2 | hsa-mir-149-5p | Down | RNF130 | 1 | hsa-mir-320a |
| Up | RPL6 | 2 | hsa-mir-16-5p | Down | UNC5A | 1 | hsa-mir-320a |
| Up | SLC16A3 | 2 | hsa-mir-98-5p | Down | CHGB | 1 | hsa-mir-375 |
| Up | CCL20 | 2 | hsa-mir-21-5p | Down | SLC32A1 | 1 | hsa-mir-148b-3p |
| Up | BCHE | 2 | hsa-mir-26b-5p | Down | CACNA1I | 1 | hsa-mir-324-3p |
| Up | C1QTNF1 | 2 | hsa-mir-335-5p | Down | CALB2 | 1 | hsa-mir-335-5p |
| Up | CA9 | 2 | hsa-mir-98-5p | Down | DYNC1I1 | 1 | hsa-mir-335-5p |
| Up | IL13RA2 | 2 | hsa-mir-148b-3p | Down | FAM189A1 | 1 | hsa-mir-335-5p |
| Up | MGP | 2 | hsa-mir-26b-5p | Down | GJB6 | 1 | hsa-mir-335-5p |
| Up | PPP1R14B | 2 | hsa-mir-615-3p | Down | GRM3 | 1 | hsa-mir-335-5p |
| Up | SEC61G | 2 | hsa-mir-1260b | Down | HHATL | 1 | hsa-mir-335-5p |
| Up | UAP1 | 2 | hsa-mir-30a-5p | Down | KCNH3 | 1 | hsa-mir-335-5p |
| Up | NEK2 | 2 | hsa-mir-128-3p | Down | KCNK4 | 1 | hsa-mir-335-5p |
| Up | CCZ1 | 2 | hsa-mir-215-5p | Down | KLK7 | 1 | hsa-mir-335-5p |
| Up | PHLDA1 | 2 | hsa-mir-375 | Down | LMO3 | 1 | hsa-mir-335-5p |
| Up | GNG10 | 2 | hsa-mir-1301-3p | Down | MATK | 1 | hsa-mir-335-5p |
| Up | PLEKHA4 | 2 | hsa-mir-124-3p | Down | NKAIN2 | 1 | hsa-mir-335-5p |
| Up | TUBB6 | 2 | hsa-mir-92b-3p | Down | NPY | 1 | hsa-mir-335-5p |
| Up | ANGPT2 | 2 | hsa-mir-542-3p | Down | PPFIA2 | 1 | hsa-mir-335-5p |
| Up | IGFBP2 | 2 | hsa-mir-126-3p | Down | PTGDS | 1 | hsa-mir-335-5p |
| Up | OSTC | 2 | hsa-mir-377-3p | Down | RGS7BP | 1 | hsa-mir-335-5p |
| Up | CDC45 | 2 | hsa-mir-455-3p | Down | SLC17A7 | 1 | hsa-mir-335-5p |
| Up | LGALS3 | 2 | hsa-mir-744-5p | Down | SLC39A12 | 1 | hsa-mir-335-5p |
| Up | NUSAP1 | 1 | hsa-let-7b-5p | Down | SYN1 | 1 | hsa-mir-335-5p |
| Up | GLT8D1 | 1 | hsa-mir-16-5p | Down | SYN2 | 1 | hsa-mir-335-5p |
| Up | LSM5 | 1 | hsa-mir-16-5p | Down | TMEM155 | 1 | hsa-mir-335-5p |
| Up | CASP4 | 1 | hsa-mir-26b-5p | Down | VIP | 1 | hsa-mir-335-5p |
| Up | EIF1AY | 1 | hsa-mir-26b-5p | Down | VSTM2L | 1 | hsa-mir-335-5p |
| Up | GGH | 1 | hsa-mir-26b-5p | Down | EFHD1 | 1 | hsa-mir-193b-3p |
| Up | GPNMB | 1 | hsa-mir-26b-5p | Down | ITPKA | 1 | hsa-mir-193b-3p |
| Up | SRPX | 1 | hsa-mir-26b-5p | Down | CAPN3 | 1 | hsa-mir-544a |
| Up | TMSB15A | 1 | hsa-mir-26b-5p | Down | DNM1 | 1 | hsa-mir-615-3p |
| Up | S100A10 | 1 | hsa-mir-100-5p | Down | ZCCHC12 | 1 | hsa-mir-615-3p |
| Up | ALPK2 | 1 | hsa-mir-214-3p | Down | RASAL1 | 1 | hsa-mir-421 |
| Up | ACTR3 | 1 | hsa-mir-124-3p | Down | PRODH | 1 | hsa-mir-23b-5p |
| Up | GBP1 | 1 | hsa-mir-124-3p | Down | MBP | 1 | hsa-mir-127-5p |
| Up | PRPH | 1 | hsa-mir-124-3p | Down | MAP7D2 | 1 | hsa-mir-505-5p |
| Up | SNRPG | 1 | hsa-mir-320a | Down | HPCAL4 | 1 | hsa-mir-744-5p |
| Up | H19 | 1 | hsa-mir-375 | Down | ANO4 | 1 | hsa-mir-378c |
| Up | NOX4 | 1 | hsa-mir-148b-3p |  |  |  |  |
| Up | AMY1A | 1 | hsa-mir-335-5p |  |  |  |  |

|  |  |  |  |
| --- | --- | --- | --- |
| Up | APOC1 | 1 | hsa-mir-335-5p |
| Up | CHI3L2 | 1 | hsa-mir-335-5p |
| Up | GDF15 | 1 | hsa-mir-335-5p |
| Up | RNASE2 | 1 | hsa-mir-335-5p |
| Up | TNC | 1 | hsa-mir-335-5p |
| Up | LYPD1 | 1 | hsa-mir-423-3p |
| Up | SLC40A1 | 1 | hsa-mir-485-3p |
| Up | TPX2 | 1 | hsa-mir-193b-3p |

Degree – No of miRNA interact with target gene. We taken any one miRNA in table.

### Construction of target gene - TF regulatory network

For further research of the target genes (up and down regulated genes), target gene-related TFs were predicted by ChEA database. Main TFs with interactions of target genes are listed in Table 8. Target genes were found to play a key role in regulating TFs. The target genes - TF regulatory network (up regulated genes) included 555 nodes and 9100 edges (Fig.14). ABCC3 was predicted to regulate 225 TFs (eg, SOX2), VKORC1 was predicted to regulate 180 TFs (eg, NANOG), MCTS1 was predicted to regulate 171 TFs (eg, SPI1), TNFRSF12A was predicted to regulate 167 TFs (eg, E2F1) and C15orf48 was predicted to regulate 155 TFs (eg, POU5F1). These target genes were enriched in whole membrane, cell cycle and cytokine signaling in immune system. The target genes - TF regulatory network (down regulated genes) included 576 nodes and 8171 edges (Fig.15). ABCA2 was predicted to regulate 234 TFs (eg, SUZ12), MOBP was predicted to regulate 201 TFs (eg, REST), PLEKHG3 was predicted to regulate 198 TFs (eg, EGR1), TTLL7 was predicted to regulate 188 TFs (eg, SOX2) and CAPN3 was predicted to regulate 178 TFs (eg, AR). These target genes were enriched in transmembrane transport of small molecules, cytoskeletal protein binding, neuron projection and Huntington disease.

**Fig. 14.**
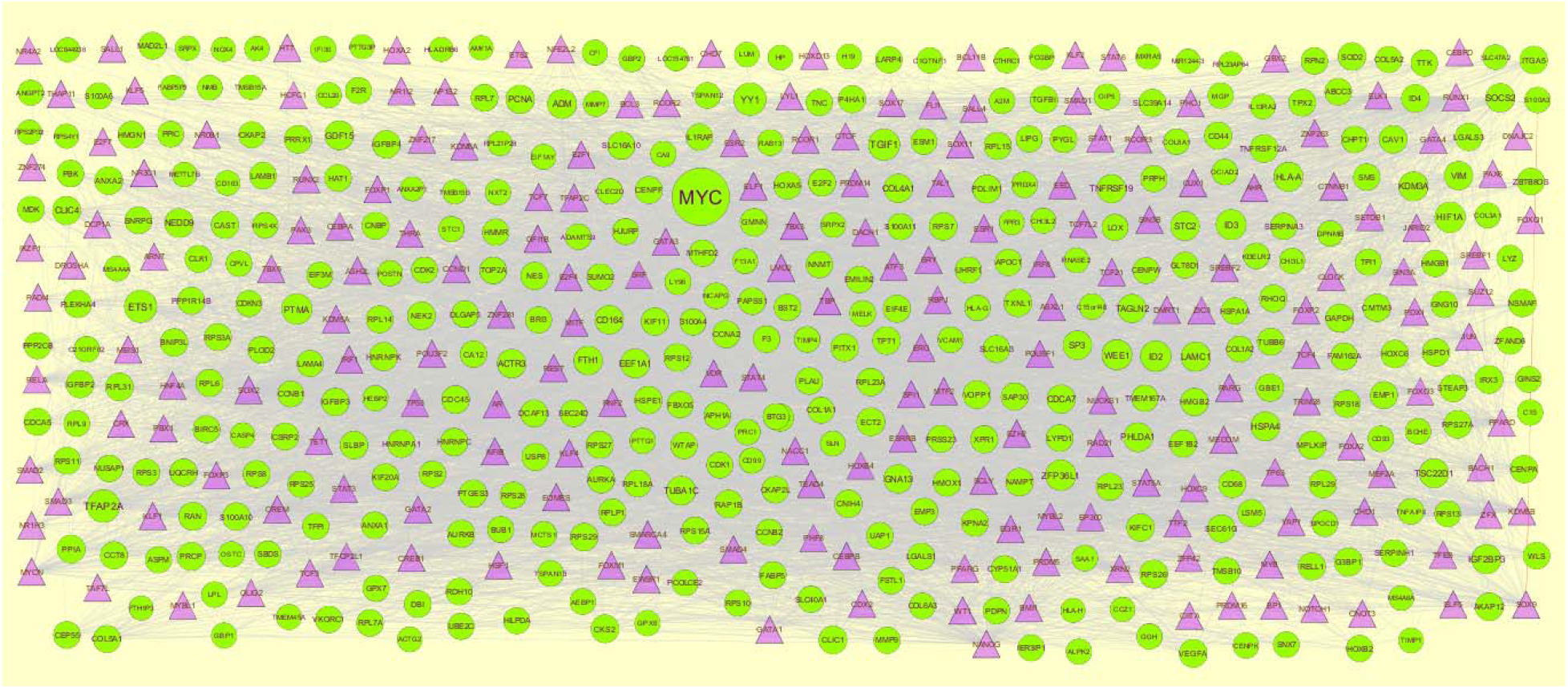
The network of up regulated genes and their related TFs. The green circles nodes are the up regulated genes, and purple triangle nodes are the TFs

**Fig. 15.**
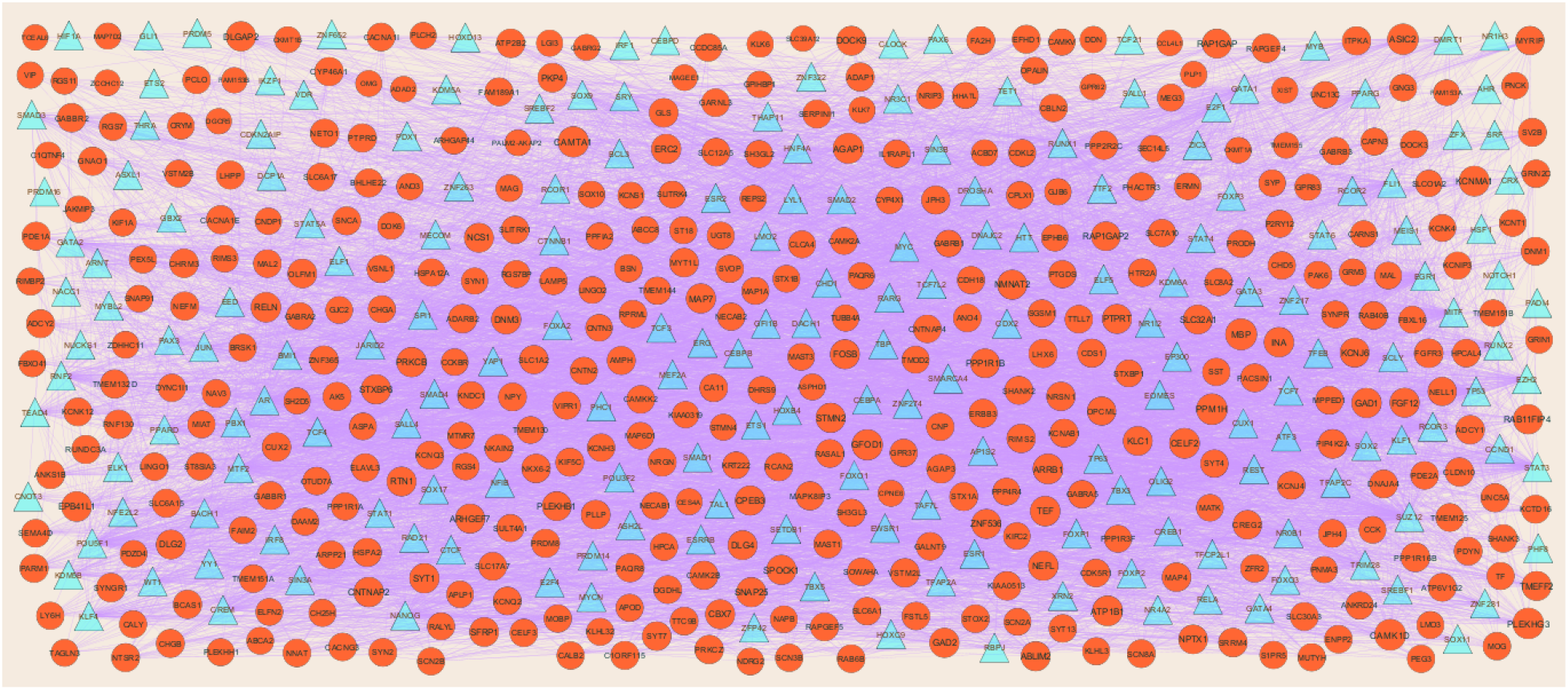
The network of down regulated genes and their related TFs. The green circles nodes are the down regulated genes, and blue triangle nodes are the TFs

**Table 8.**
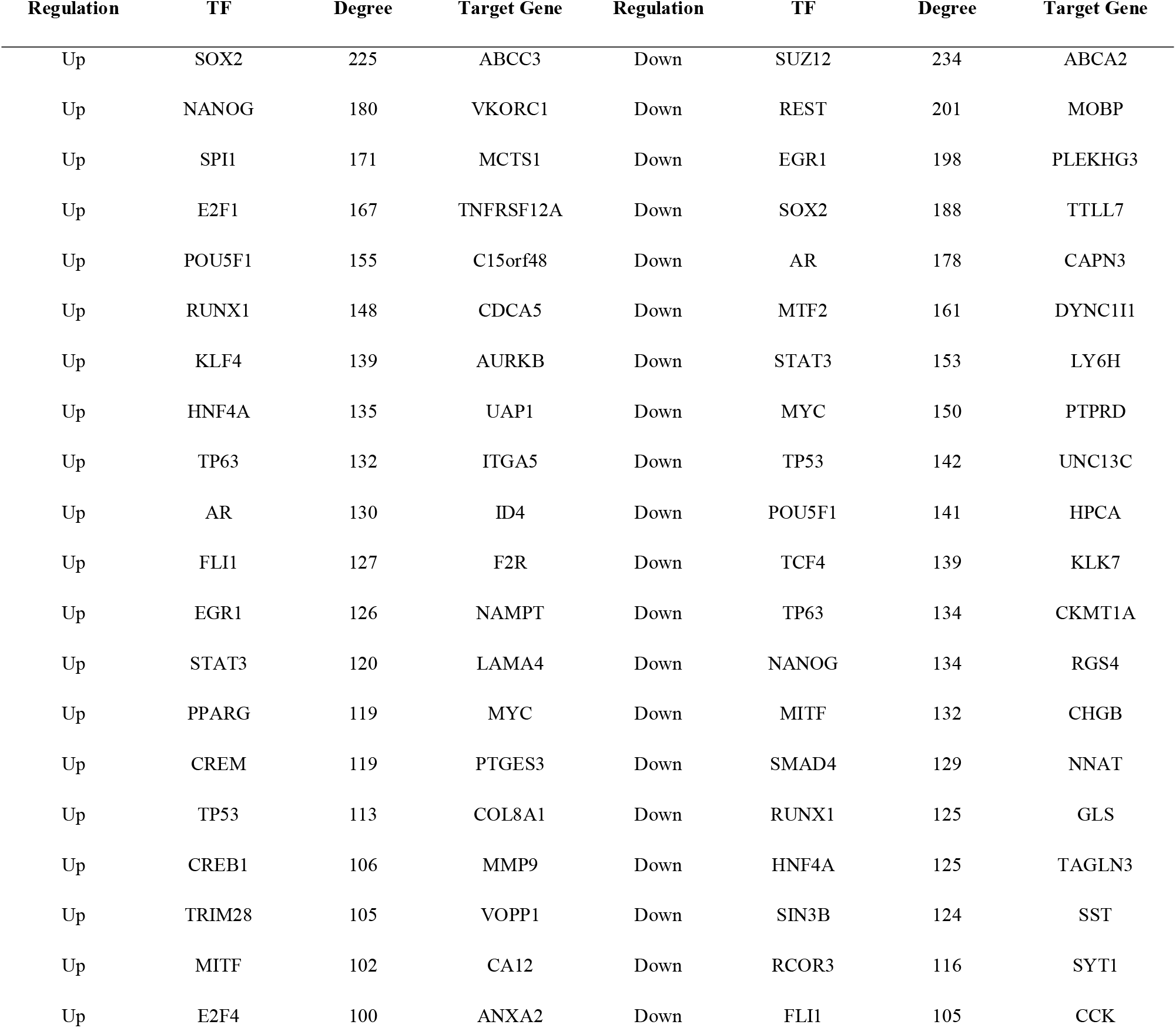
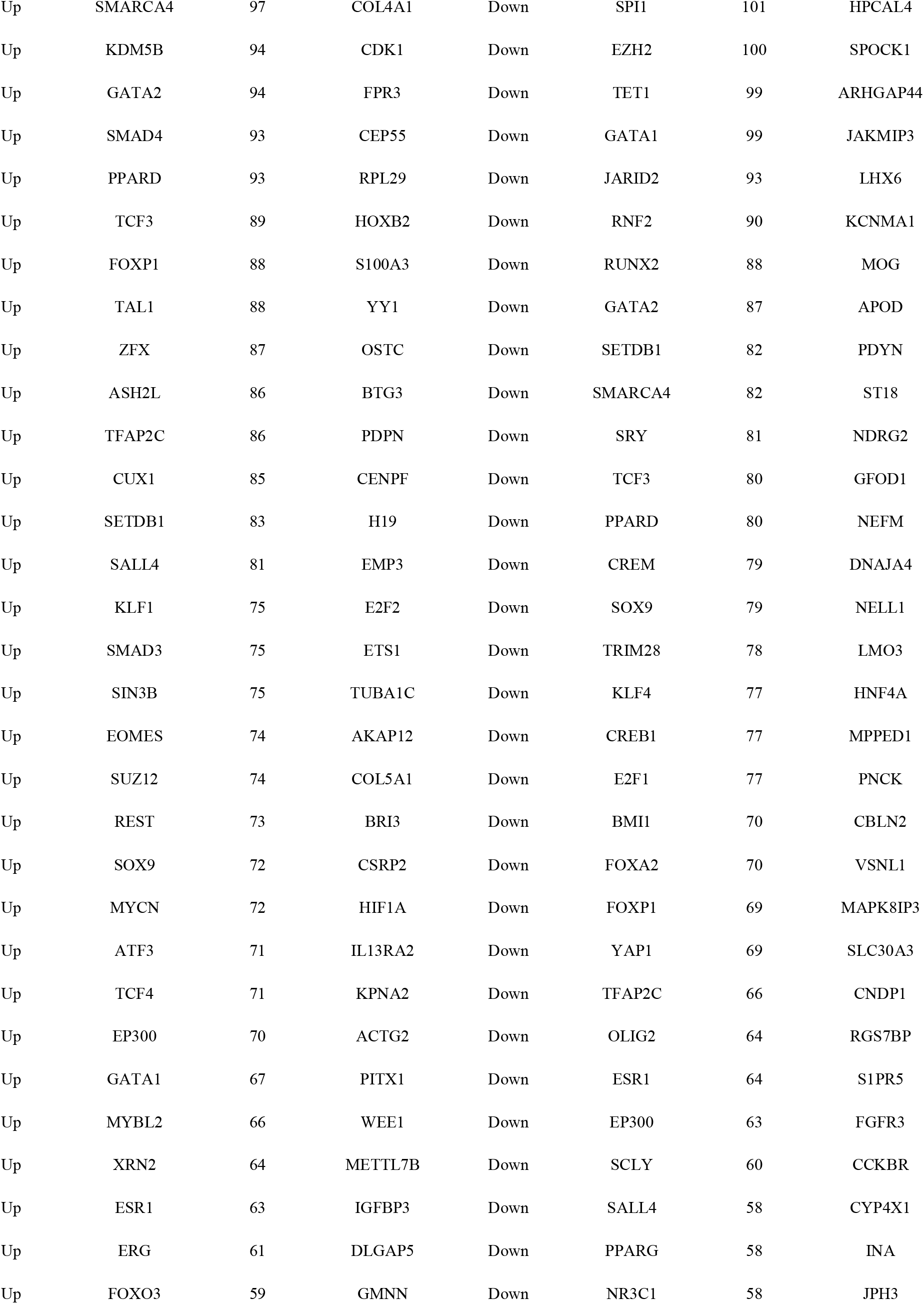
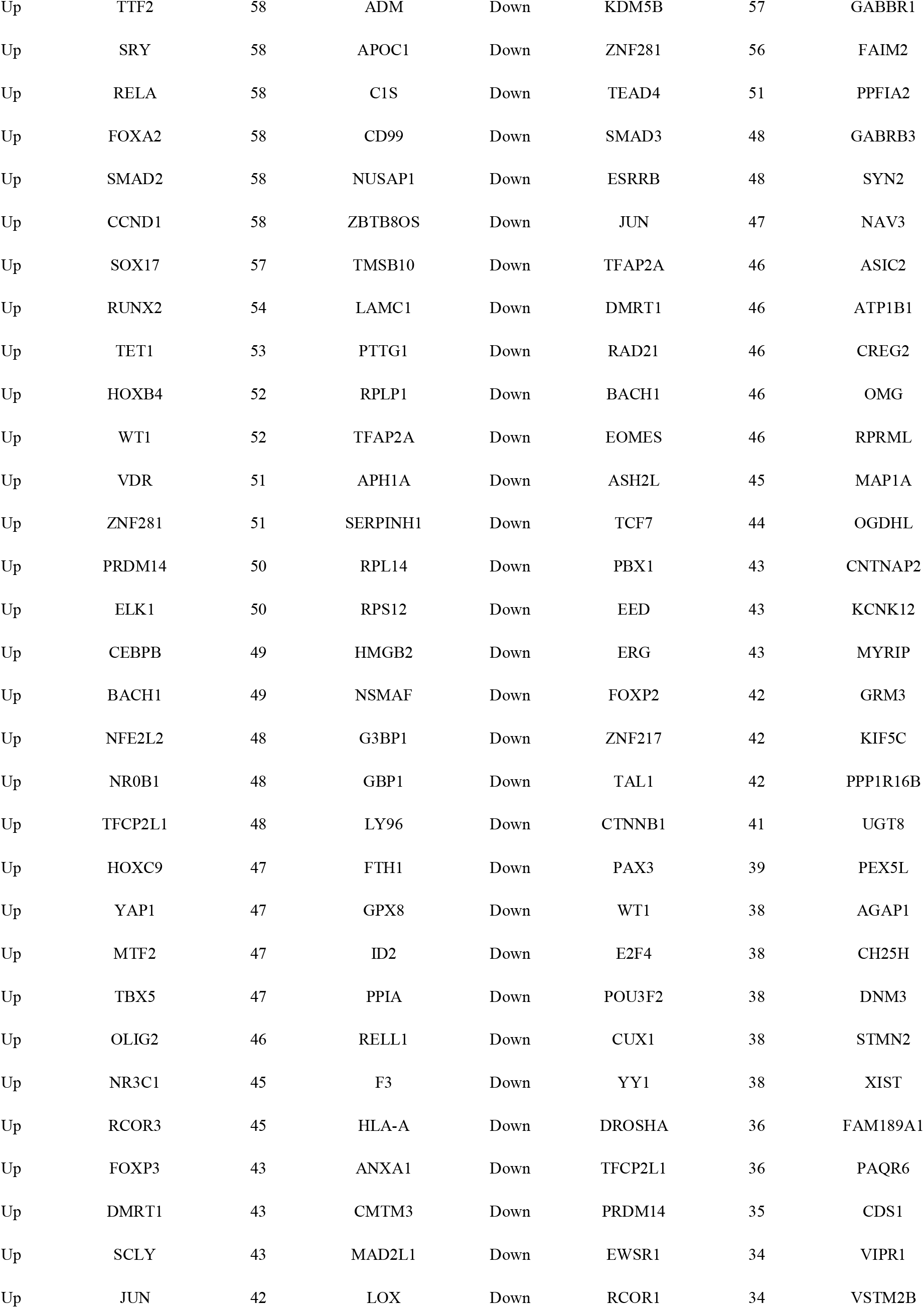
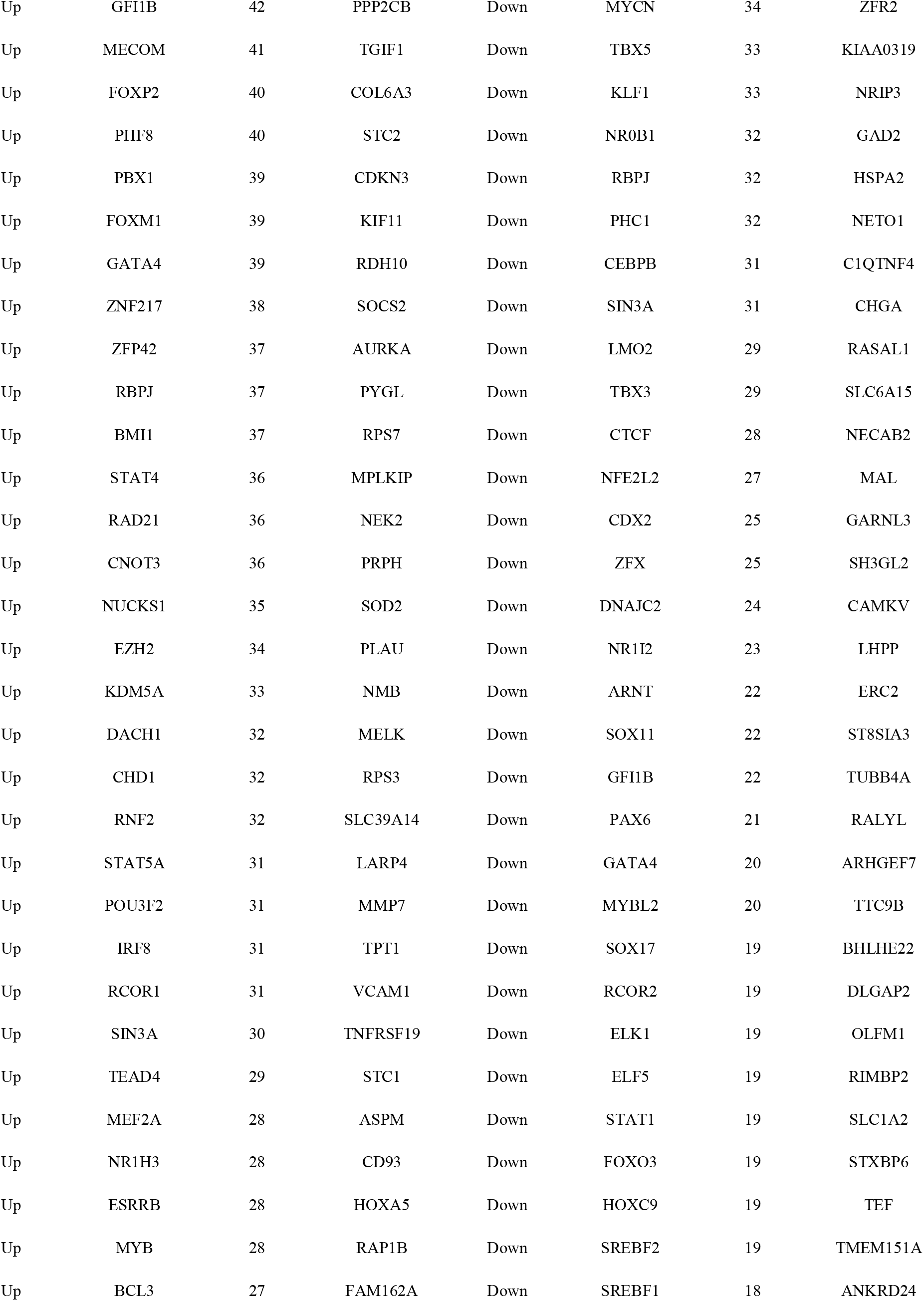
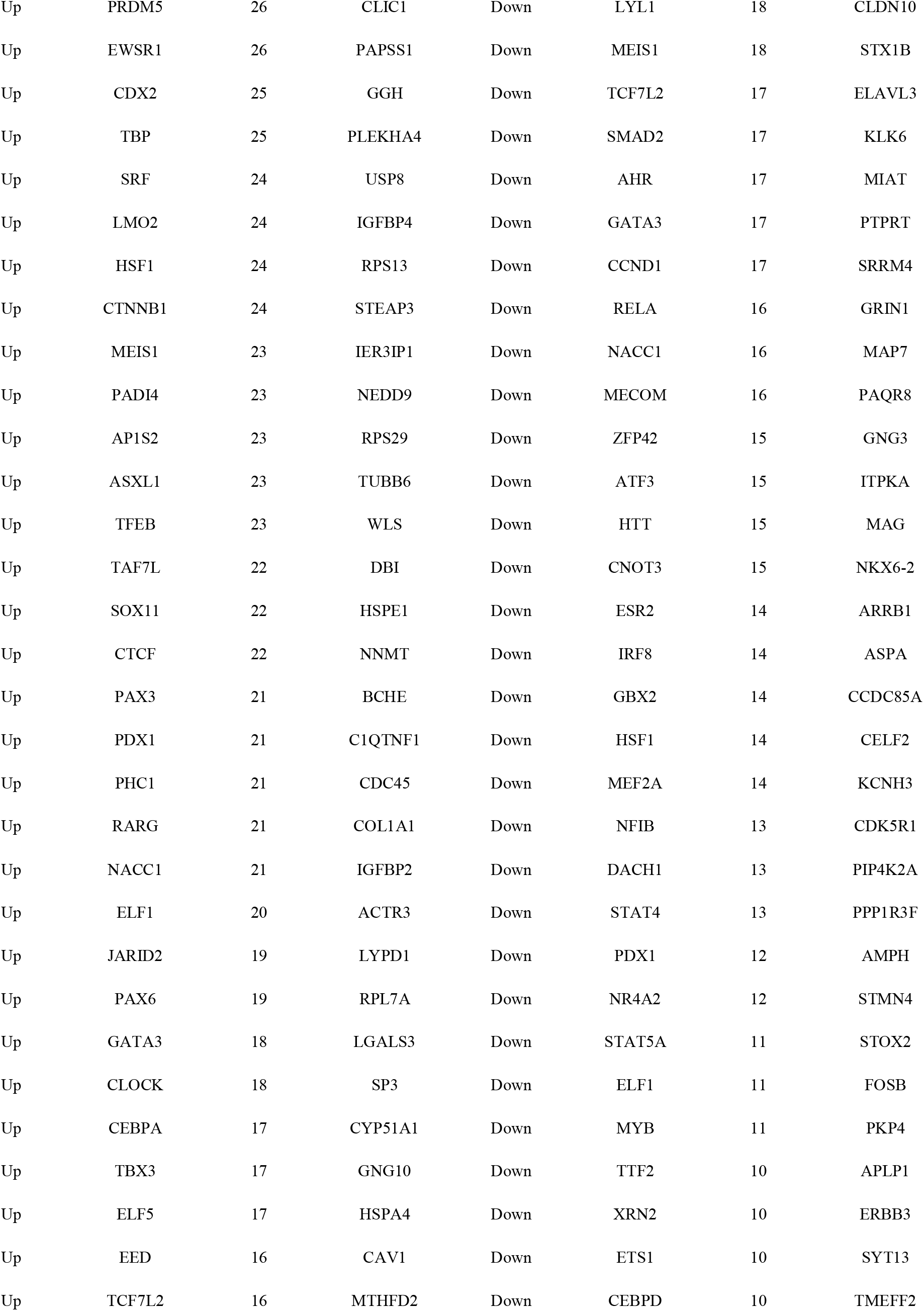
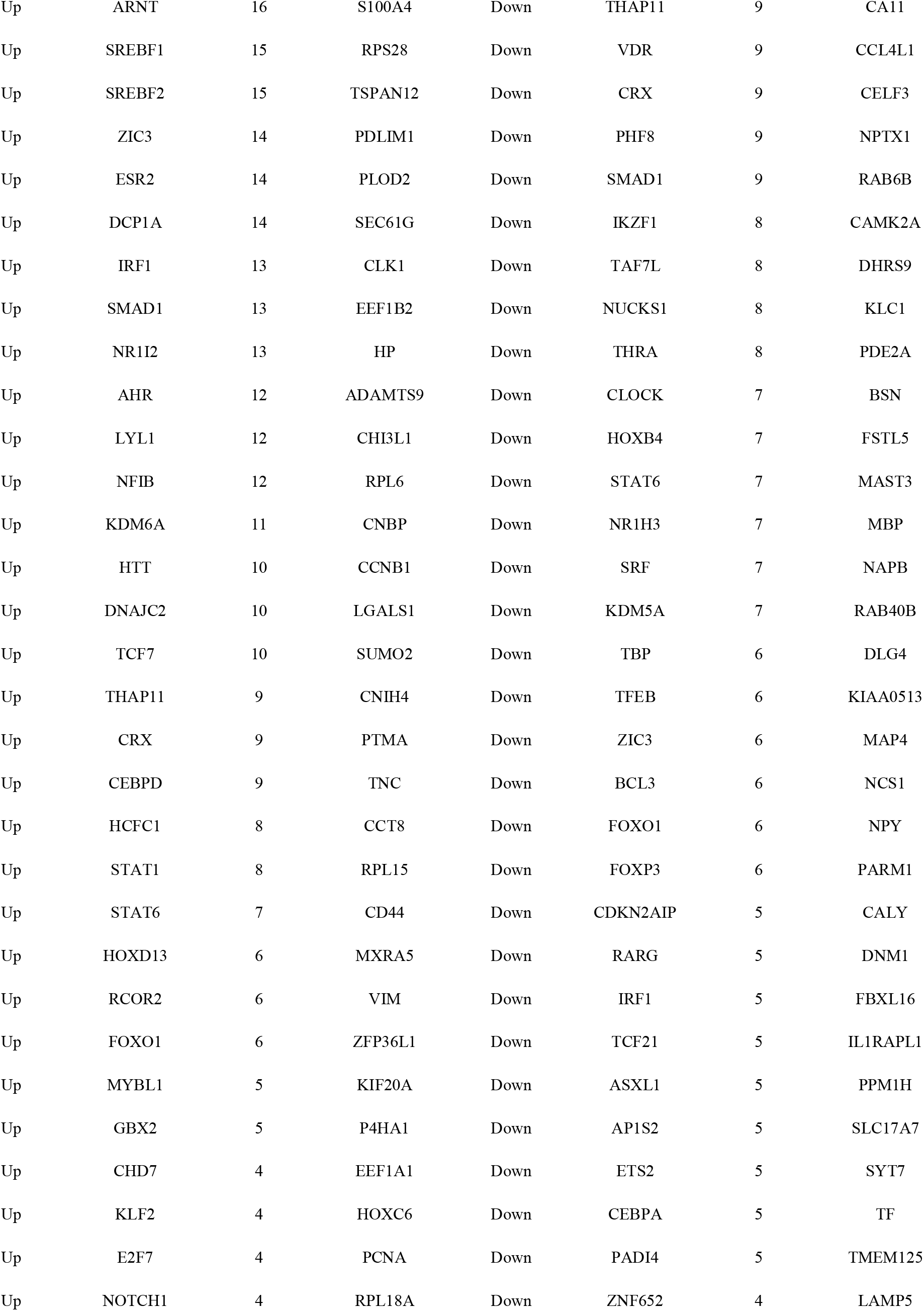
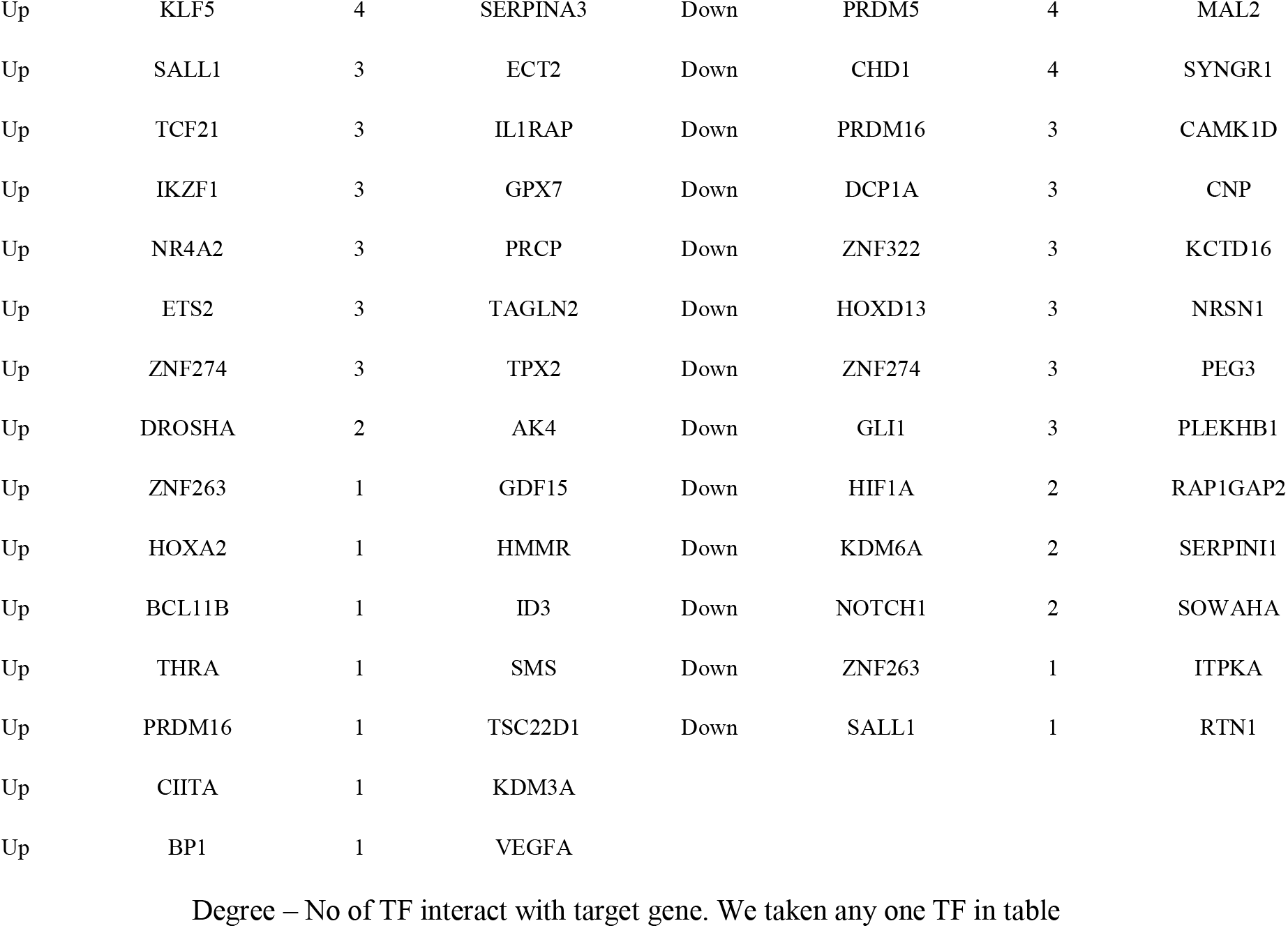
TF - target gene interaction table

### Validation of hub genes and clinical significance

UALCAN, the online tool with data sourced from TCGA, was used to validate the expression of these hub genes in GBM. Survival analysis (P<0.05) (Fig. 16); highly expressing TUBA1C, CAV1, S100A4, DNAJA4, PAK6, NELL1 and ITPKA tends to have poor survival outcomes in GBM. However, low expressing RPL23, YY1 and ARHGEF7 tends to have poor survival outcomes in GBM. As shown in Fig 17, the expression of the up regulated hub genes TUBA1C, CAV1, RPL23, YY1 and S100A4 in GBM tissue were significantly elevated compared with normal brain tissues. Furthermore, the expressions of down regulated hub genes ARHGEF7, DNAJA4, PAK6, NELL1 and ITPKA in GBM tissue were significantly decreased compared with normal brain tissues. The expression of each hub gene in GBM patients was analyzed according to the patient’s age. As shown in Fig 18, the expression of TUBA1C, CAV1, RPL23, YY1 and S100A4 were higher in patients with age (21-40 years, 21-60 years, 61-80 years, 81-100 years), which revealed that these up regulated hub genes might be associated with GBM advancement positively. Similarly, the expression of ARHGEF7, DNAJA4, PAK6, NELL1 and ITPKA were lower in patients age (21-40 years, 21-60 years, 61-80 years, 81-100 years), which revealed that these down regulated hub genes might be linked with GBM advancement positively. Fig. 19 presented the mutation information of the ten hub genes. TUBA1C, CAV1, RPL23, YY1, S100A4, ARHGEF7, DNAJA4, PAK6, NELL1 and ITPKA were changed most often (0.7%, 0.4%, 0.4%, 0.7%, 0.4%, 0.7%, 0.4%, 0.7%, 1.5% and 0.4%), these include amplification, deep deletion, missense mutation and truncating mutation. The Human Protein Atlas (THPA) demonstrated that the expression of TUBA1C, CAV1, RPL23, YY1 and S100A4 were highly expressed in GBM tissues, whereas ARHGEF, DNAJA4, PAK6, NELL1 and ITPKA were low expressed in GBM tissue (Fig. 20). To verify the diagnostic value of the hub genes, expression levels in GBM were evaluated using ROC curves. As presented in Fig. 21, the area under the curve (AUC) for TUBA1C, CAV1, RPL23, YY1, S100A4, ARHGEF7, DNAJA4, PAK6, NELL1 and ITPKA in GBM and normal control tissue determined for the GSE116520 dataset were 0.963, 0.971, 0.993, 0.963, 0.971, 0.963, 0.985, 0.971, 0.978 and 0.985, respectively. RT-PCR demonstrated that the relative expression levels of TUBA1C, CAV1, RPL23, YY1 and S100A4 in GBM tissues were significantly higher compared with those in normal tissue (Fig. 22A - E), whereas expression levels of ARHGEF7, DNAJA4, PAK6, NELL1 and ITPKA in GBM tissue were significantly lower compared with those in normal tissue (Fig. 22 F-J). The PCR primers are listed in Table 9. To investigate the immune infiltration analysis of the ten potential hub genes, the TIMER bioinformatics analysis platform was used. We found that the high expression of hub genes (TUBA1C, CAV1, RPL23, YY1 and S100A4) were negatively associated with tumor purity (Fig. 23A - E), where as low expression of hub genes (ARHGEF7, DNAJA4, PAK6, NELL1 and ITPKA) were positively associated with tumor purity (Fig. 23A - E).

**Fig. 16.**
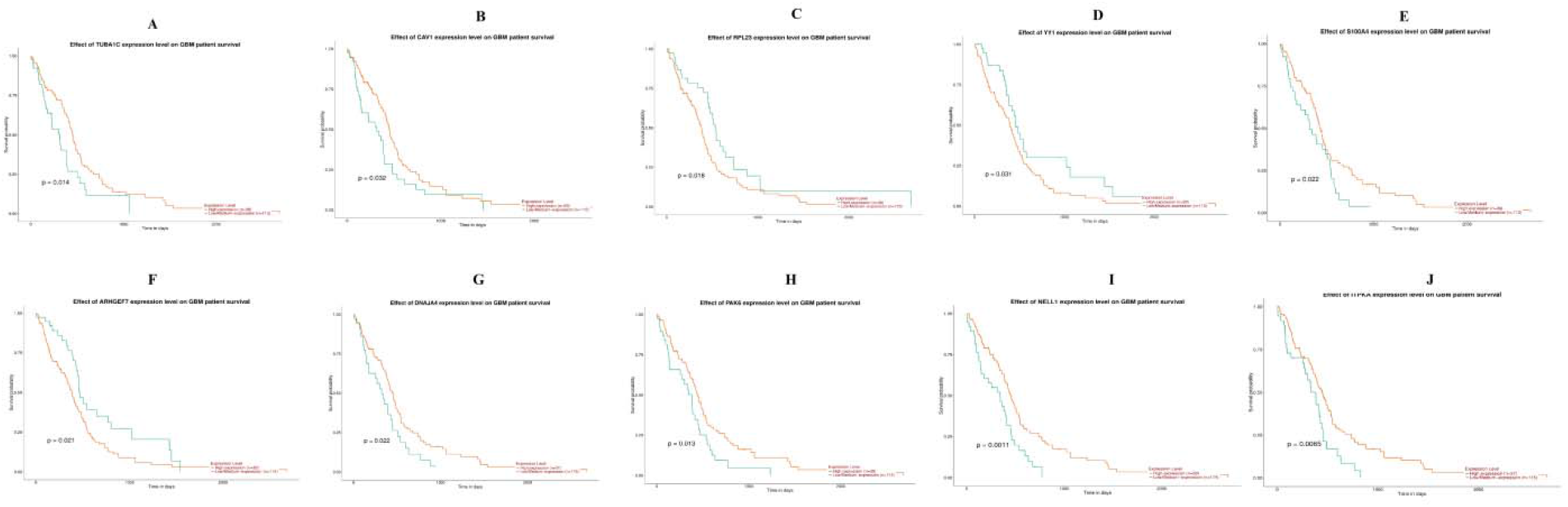
Overall survival analysis of hub genes. Overall survival analyses were performed using the UALCAN online platform A) TUBA1C B) CAV1 C) RPL23 D) YY1 E) S100A4 F) ARHGEF7 G) DNAJA4 H) PAK6 I) NELL1 J) ITPKA

**Fig. 17.**
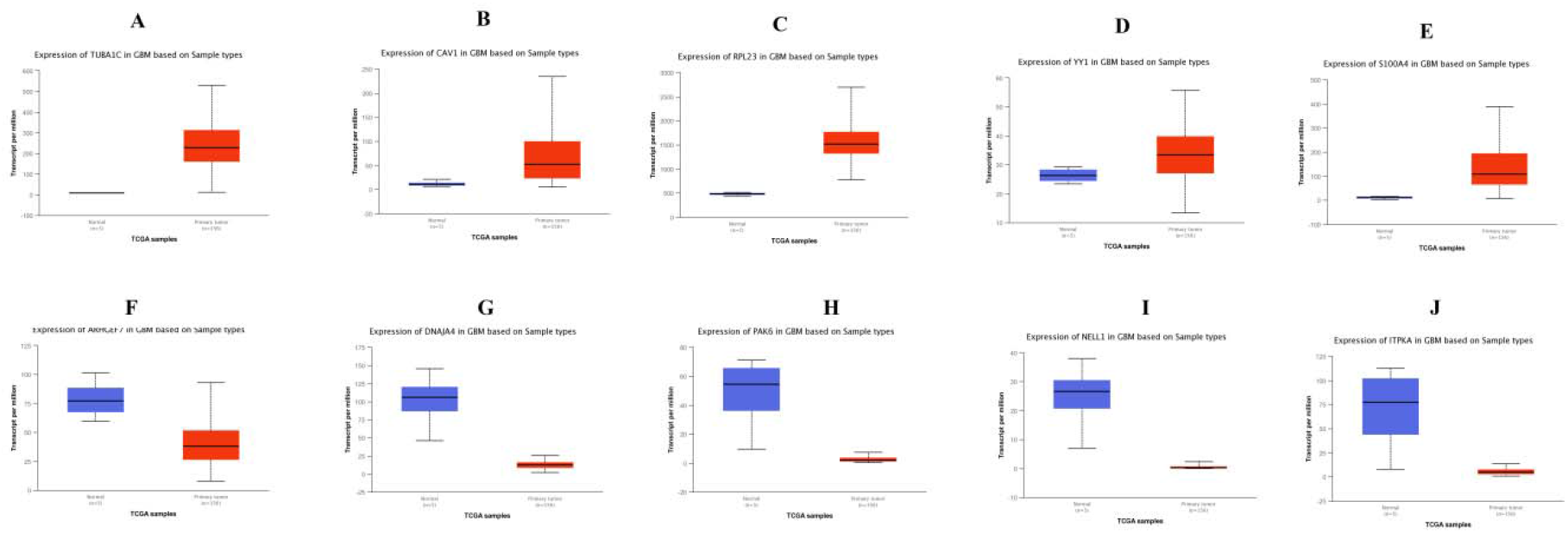
Box plots (expression analysis) hub genes were produced using the UALCAN platform A) TUBA1C B) CAV1 C) RPL23 D) YY1 E) S100A4 F) ARHGEF7 G) DNAJA4 H) PAK6 I) NELL1 J) ITPKA

**Fig. 18.**
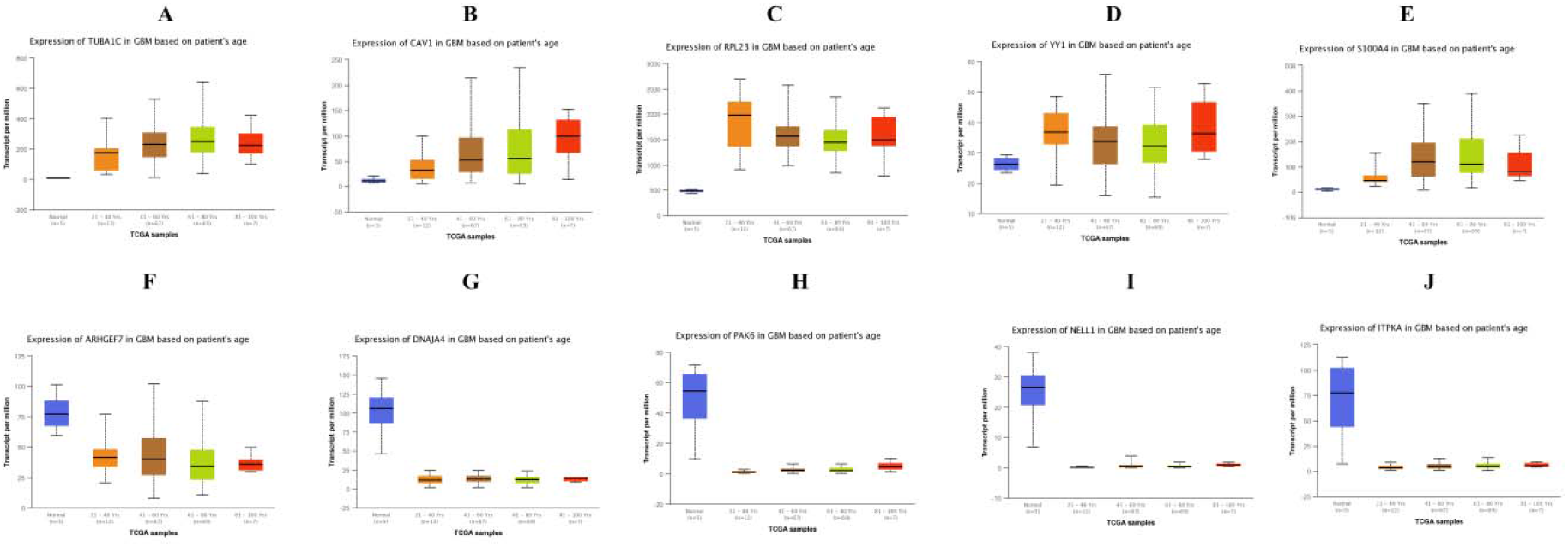
Box plots (age analysis) of hub genes were produced using the UALCAN platform A) TUBA1C B) CAV1 C) RPL23 D) YY1 E) S100A4 F) ARHGEF7 G) DNAJA4 H) PAK6 I) NELL1 J) ITPKA

**Fig. 19.**
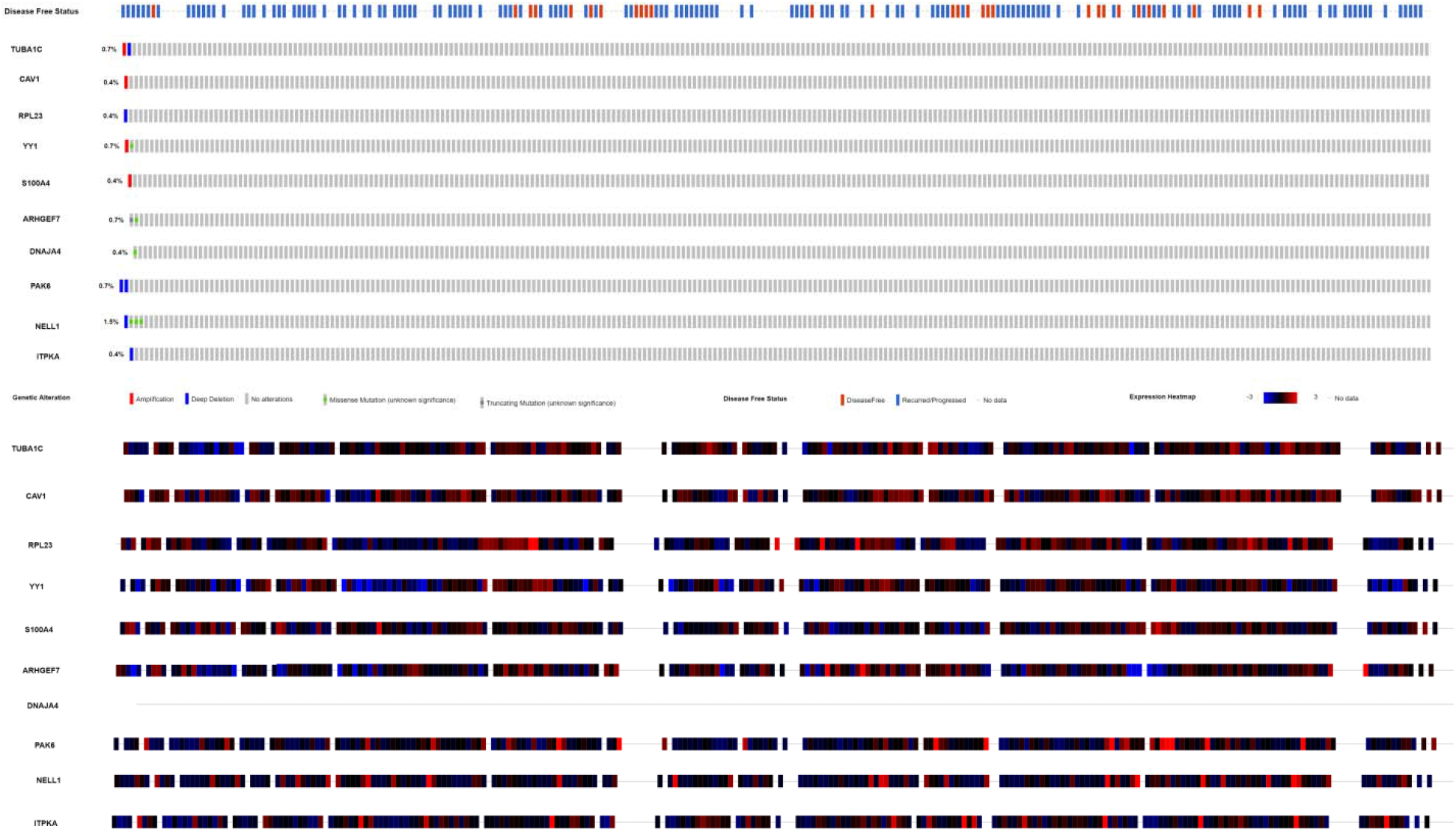
Mutation analyses of hub genes were produced using the CbioPortal online platform

**Fig. 20.**
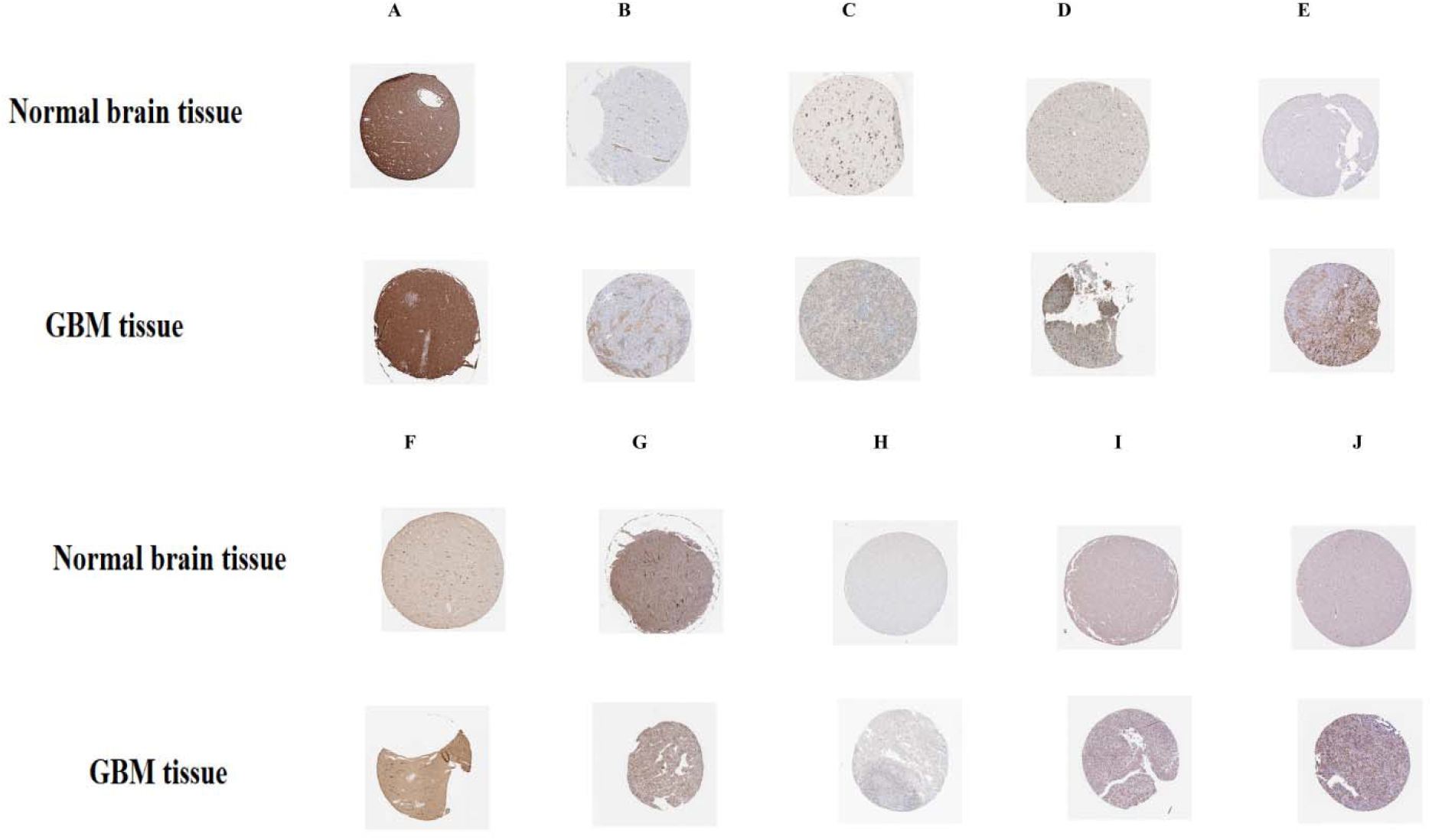
Immunohisto chemical(IHC) analyses of hub genes were produced using the human protein atlas (HPA) online platform A) TUBA1C B) CAV1 C) RPL23 D) YY1 E) S100A4 F) ARHGEF7 G) DNAJA4 H) PAK6 I) NELL1 J) ITPKA

**Fig. 21.**
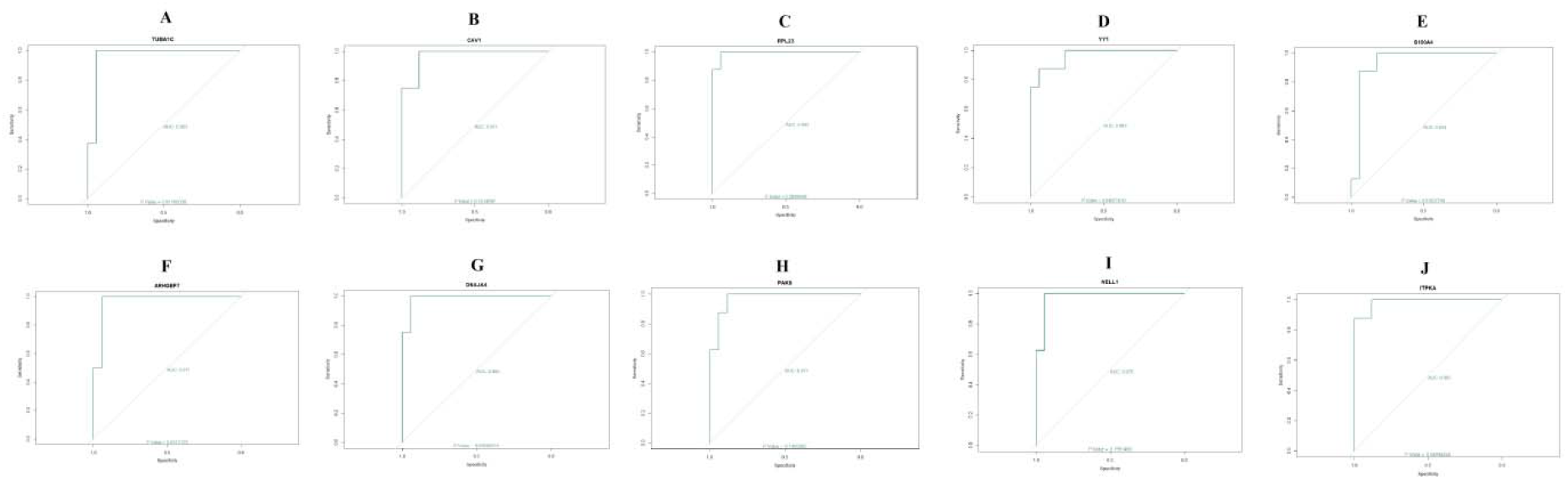
ROC curve validated the sensitivity, specificity of hub genes as a predictive biomarker for GBM prognosis A) TUBA1C B) CAV1 C) RPL23 D) YY1 E) S100A4 F) ARHGEF7 G) DNAJA4 H) PAK6 I) NELL1 J) ITPKA

**Fig. 22.**
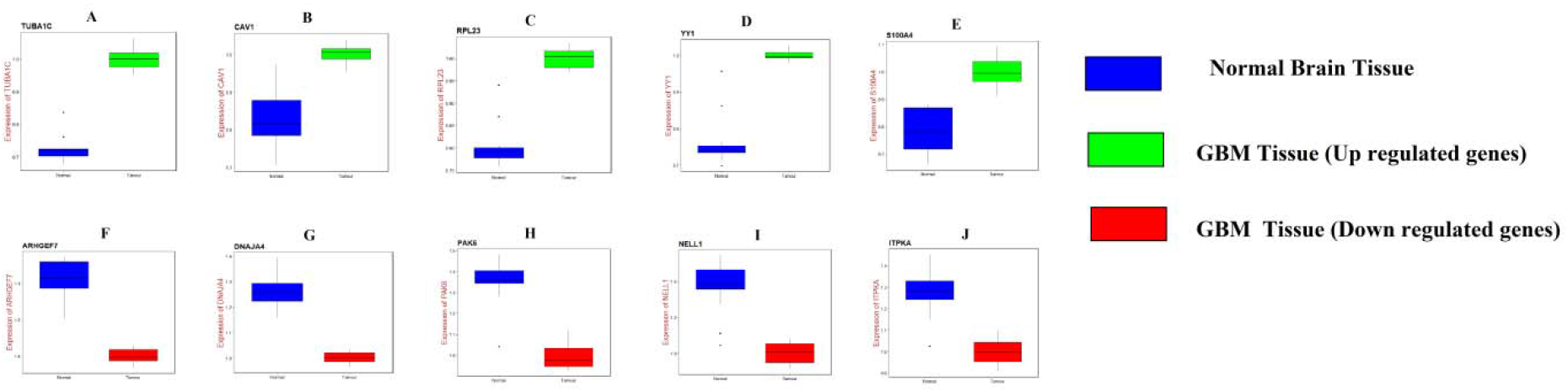
Validation of hub genes by RT-PCR. A) TUBA1C B) CAV1 C) RPL23 D) YY1 E) S100A4 F) ARHGEF7 G) DNAJA4 H) PAK6 I) NELL1 J) ITPKA

**Fig. 23.**
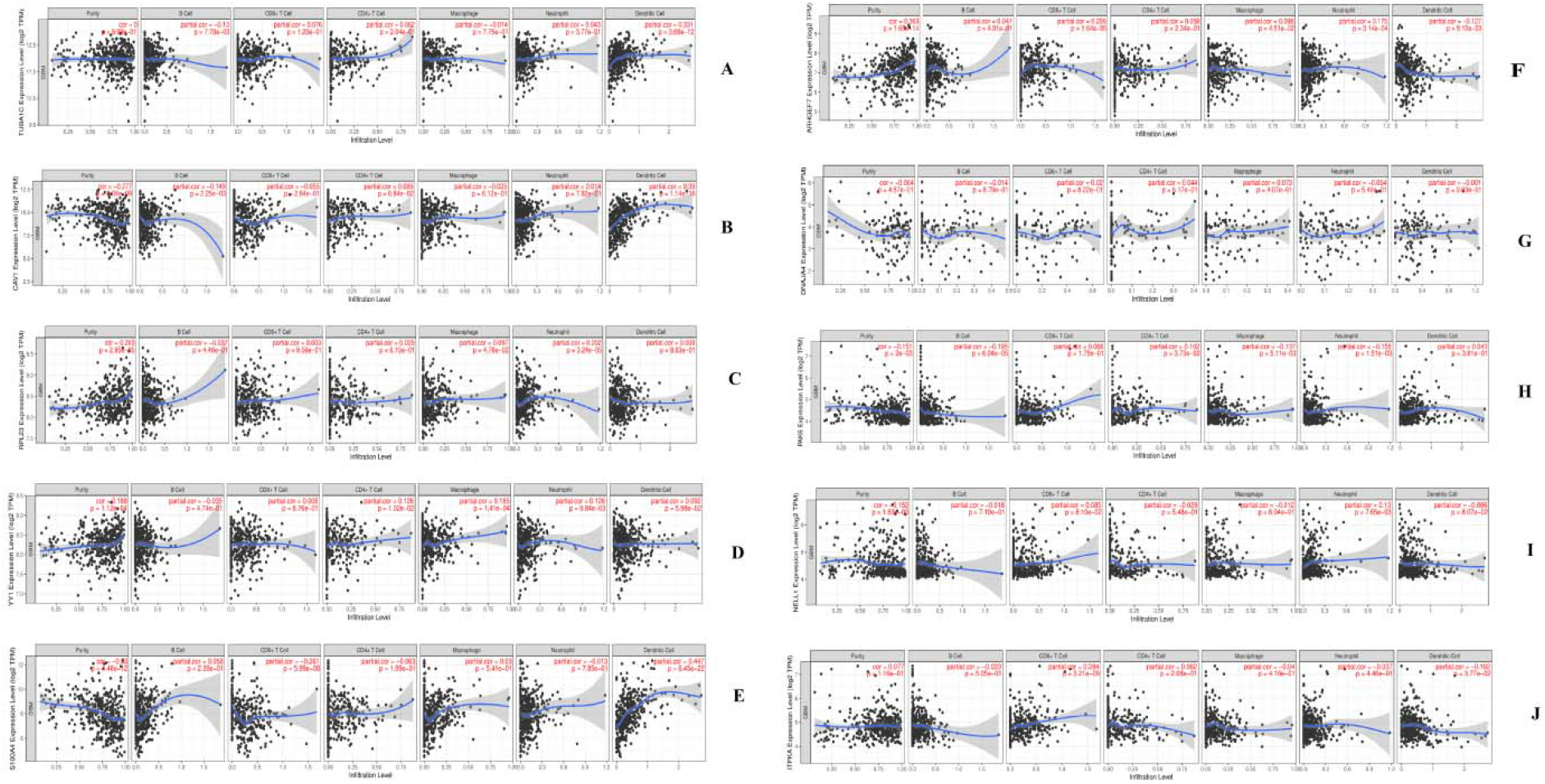
Scatter plot for immune infiltration for hub genes. A) TUBA1C B) CAV1 C) RPL23 D) YY1 E) S100A4 F) ARHGEF7 G) DNAJA4 H) PAK6 I) NELL1 J) ITPKA

**Table 9.**
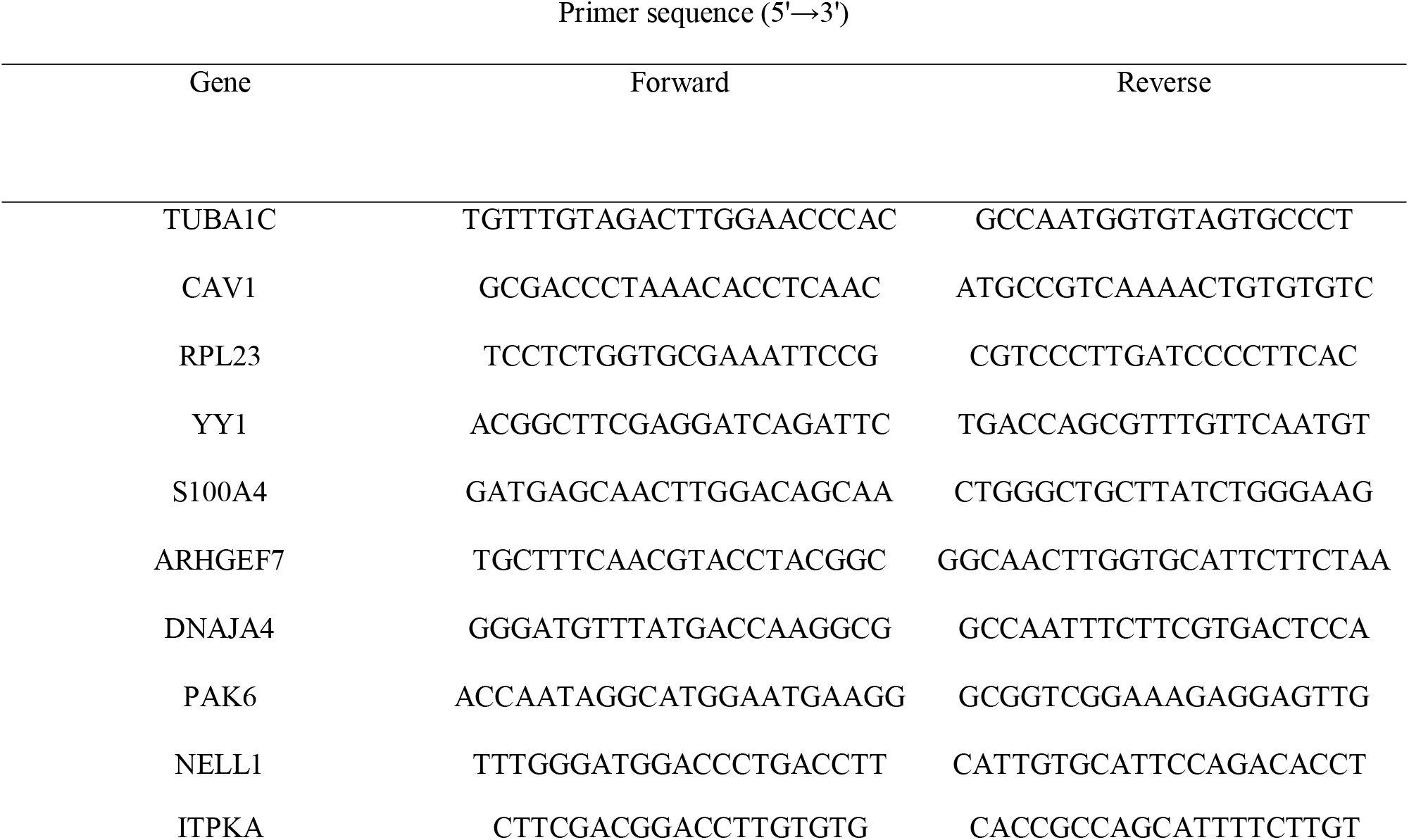
Primers used for quantitative PCR

## Discussion

The majorities of patients with GBM are diagnosed at advanced stages and have poor overall survival [57]. However, the molecular mechanisms associated in the advancement of GBM remain unclear. In the present study, to better understanding the molecular mechanisms involved in GBM progression, we characterized the expression profiles in GBM and normal brain tissues by transcriptome analysis. Using bioinformatics analysis, we obtained 947 DEGs from GSE116520 data expression profiles, including 477 up regulated genes and 470 down regulated genes. Genes such as SERPINA3 [58], VIP (vasoactive intestinal peptide) [59], ANXA2 [60] and SST (somatostatin) [61] were associated with progression of GBM. RPL39 [62] was responsible for invasion of breast cancer cells, but this gene was identified first time in GBM and may be linked with invasion of GBM cells. Genes such as TUBA1 [63], RPN2 [64], RASAL1 [65] and CCKBR (cholecystokinin B receptor) [66] were involved in proliferation of different cancer cells, but expression of these genes are not reported in GBM and may be associated with proliferation of GBM cells. High expression of KLK7 was important for pathogenesis of ovarian cancer [67], but elevated expression of this gene was identified first time in GBM and may be liable for progression of GBM.

In pathway enrichment analysis for up regulated genes was performed. Enriched genes such as SOD2 [68], RPS11 [69], RPL9 [70], MYC (MYC proto-oncogene, bHLH transcription factor) [71], SEC61G [72], BIRC5 [73], NEK2 [74], CDK2 [75], AURKB (aurora kinase B) [76], RPS3 [77], MGP (matrix Gla protein) [78], AEBP1 [79], CTHRC1 [80], COL1A1 [81], COL3A1 [82], TNC (tenascin C) [83], POSTN (periostin) [84], IGFBP2 [85], IGFBP3 [86], IGFBP4 [87], SRPX2 [88], LAMB1 [89], ESM1 [90], TGFBI (transforming growth factor beta induced) [91], ITGA5 [92], RAP1B [93], CAV1 [94], HMOX1 [95] and LOX (lysyl oxidase) [96] were linked with progression of GBM. GPX7 was important for advancement of gastric cancer [97], but this gene was identified first time in GBM and may be liable for progression of GBM. High expression of enriched genes such as RPL29 [98], RPLP1 [99], RPS2 [100], RPS3A [101], RPS13 [102], RPS15A [103], RPL7A [104], CENPA (centromere protein A) [105], CENPF (centromere protein F) [106], EIF4E [107], MXRA5 [108] and LUM (lumican) [109] were responsible for development of different cancer types, but over expression of these genes were identified first time in GBM and may be associated with development of GBM. Enriched genes such as RPS12 [110], RPL6 [111], LAMA4 [112], CCNB1 [113], CCNB2 [114], CDK1 [115] and EIF3M [116] were responsible for proliferation of different cancer cell types, but these genes were identified first time in GBM and may be associated with proliferation of GBM cells. Enriched polymorphic genes such as RPL14 [117] and LAMC1 [118] were liable for advancement of different cancer types, but these polymorphic genes were identified first time in GBM and may be important for development of GBM. Enriched genes associated such as RPL15 [119], EEF1A1 [120], SRPX (sushi repeat containing protein X-linked) [121], COL1A2 [122], COL4A1 [123], COL5A1 [124], COL5A2 [125], COL6A3 [122] and COL8A1 [126] were involved in invasion of different cancer cell types, but these genes were identified first time in GBM and may be culpable for invasion of GBM cells. EMILIN2 was associated with angiogenesis in gastric cancer [127], but this gene was identified first time in GBM and may be important for angiogenesis in GBM. Our study found that GPX8, RPL23A, RPL31, RPS4X, RPS4Y1, RPS7, RPS8, RPS10, RPS18, RPS25, RPS26, RPS27, RPS27A, RPS28, RPS29, RPL23, RPL7, RPL18A, EEF1B2, AMY1A, GBE1, PYGL (glycogen phosphorylase L), PCOLCE2, TNFAIP6 and SLC7A6 are up regulated in GBM and has potential as a novel diagnostic and prognostic biomarker, and therapeutic target. Similarly, pathway enrichment analysis for down regulated genes was performed. Enriched genes such as SLC12A5 [128], SHANK2 [129], KCNJ4 [130] and CACNA1E [131] were linked with development of different cancer types, but these genes were identified first time in GBM and may be important for progression of GBM. Enriched genes such as SLC6A1 [132], GABBR1 [133] and GAD1 [134] were associated with invasion of different cancer cells, but these genes were identified first time in GBM and may be involved in invasion of GBM cells. Enriched genes such as GLS (glutamines) [135], NEFL (neurofilament light) [136], SYN1 [137], SLC17A7 [138], SYT7 [139], EPB41L1 [140] and TF (transferrin) [141] were responsible for advancement of GBM. Methylation inactivation of enriched tumor suppressor genes such as such as GNAO1 [142], KCNMA1 [143] and CAMK2B [144] were liable for progression of different cancer types, but these genes were identified first time in GBM and loss of these genes may be linked with development of GBM. Low expression of UNC13C was associated with development of oral cancer [145], but this gene was identified first time in GBM and decrease expression of this gene may be linked with progression of GBM. Polymorphic gene CHRM3 was identified with progression of bladder cancer [146], but this polymorphic gene was identified first time in GBM and may be liable for advancement of GBM. Our study found that CARNS1, ADCY1, ADCY2, GABBR2, SLC32A1, PRKCB (protein kinase C beta), GABRA2, GABRA5, GABRB1, GABRB3, GABRG2, GAD2, KCNJ6, GNG3, SNAP25, STX1A, STXBP1, SYT1, DLGAP2, TSPOAP1, CACNG3, PPFIA2, SLC1A2, SHANK3, CPLX1, KCNK4, PTPRD (protein tyrosine phosphatase receptor type D), ABCC8, SYN2, KCNAB1, KCNQ2, KCNQ3, KCNS1, DLG2, DLG4, CAMK2A, GRIN1, GRIN2C, KCNH3, ASPA (aspartoacylase), ITPKA (inositol-trisphosphate 3-kinase A), STX1B, RIMS2 and SYP (synaptophysin) are down regulated in GBM and has potential as a novel diagnostic and prognostic biomarker, and therapeutic target.

In GO enrichment analysis for up regulated genes was performed. Enriched genes such as PTTG1 [147], HMGB1 [148], HMGB2 [149], HMMR (hyaluronan mediated motility receptor) [150], CHI3L2 [151], VEGFA (vascular endothelial growth factor A) [152], VIM (vimentin) [153], IGF2BP3 [154], UHRF1 [155], SUMO2 [156], PBK (PDZ binding kinase) [157], AURKA (aurora kinase A) [158], ADAMTS9 [159], UBE2C [160], CAST (calpastatin) [161], USP8 [162], TIMP1 [163], TIMP4 [164], CD44 [165], PCNA (proliferating cell nuclear antigen) [166], CCT8 [167], CHI3L1 [168] and ANXA1 [169] were involved in progression of GBM. HNRNPC (heterogeneous nuclear ribonucleoprotein C) was associated with drug resistance in gastric cancer [170], but this gene was identified first time in GBM and may be associated with chemo resistance in GBM. Enriched genes such as HSPA1A [171] and TUBA1C [172] were linked with proliferation of liver cancer cells, but these genes were identified first time in GBM and may be involved in proliferation of GBM cells. High expression of enriched genes such as MAD2L1 [173] and CSRP2 [174] were linked with pathogenesis of different cancer types, but high expression of these genes were identified first time in GBM and may be involved in progression of GBM. CASP4 [175] was involved in advancement of esophageal cancer, but this gene was identified first time in GBM and may be associated in development of GBM. Our study found that LSM5, CPVL (carboxypeptidasevitellogenic like), PPP2CB, CYP51A1, BNIP3L, FBXO5, ZFP36L1, RNASE2, MCTS1, LARP4, PRPH (peripherin), POTEKP (POTE ankyrin domain family member K, pseudogene), TUBB6, ACTR3 and RNA28SN5 are up regulated in GBM and has potential as a novel diagnostic and prognostic biomarker, and therapeutic target. Similarly, GO enrichment analysis for down regulated genes was performed. Enriched genes such as MAG (myelin associated glycoprotein) [176], ASIC2 [177], MBP (myelin basic protein) [178], CNP (2’,3’-cyclic nucleotide 3’ phosphodiesterase) [179], CPEB3 [180], SLC8A2 [181], PRKCZ (protein kinase C zeta) [182], RELN (reelin) [183], CYP46A1 [184], SNAP91 [185], CNTN2 [186], NPY (neuropeptide Y) [187], RGS4 [188], IL1RAPL1 [189], ERBB3 [190], SH3GL2 [191], SH3GL3 [192], ARRB1 [193], DNM3 [194], SPOCK1 [195], CCK (cholecystokinin) [196] and INA (internexin neuronal intermediate filament protein alpha) [197] were identified with progression of GBM. Decrease expression of enriched genes such as such as SCN8A [198], BRSK1 [199], ANKS1B [200], CALB2 [201], GRM3 [202], BCAS1 [203] and CLCA4 [204] were responsible for advancement of different cancer types, but low expression of these genes were identified first time in GBM and may be involved in progression of GBM. Enriched genes such as CUX2 [205], NPTX1 [206], NCS1 [207], SEPTIN4 [208] and FAIM2 [209] were associated with advancement of different cancer, but these genes were identified first time in GBM and may be linked with development of GBM. MAP4 was involved in invasion of bladder cancer cells [210], but this gene was identified first time in GBM and may be responsible for invasion of GBM cells. Enriched genes such as RAB6B [211] and MAL2 [212] were linked with proliferation of different cancer cells types, but these genes were identified first time in GBM and may be liable for proliferation of GBM cells. Methylation inactivation of tumor suppressor DMTN (dematin actin binding protein) was associated with progression of colorectal cancer [213], but loss of this gene was identified first time in GBM and may be involved in advancement of GBM. Our study found that MAP1A, PDYN (prodynorphin), TMOD2, CPNE6, SCN2A, SCN2B, FGF12, PLP1, AMPH (amphiphysin), HTR2A, NSG2, NAPB (NSF attachment protein beta), CNTNAP2, CNTNAP4, CALY (calcyon neuron specific vesicular protein), ERC2, SNCA (synuclein alpha), ATP2B2, JPH4, RIMS3, CDK5R1, SV2B, SYT4, CACNA1I, BSN (bassoon presynaptic cytomatrix protein), DNM1, NRGN (neurogranin), PHF24, PCLO (piccolo presynaptic cytomatrix protein), RAPGEF4, NETO1, SYNGR1, RIMBP2, LY6H, JPH3, PDE2A, KCNIP3, SYNPR (synaptoporin), SLITRK1, HPCA (hippocalcin), CAMKV (CaM kinase like vesicle associated), KCTD16, PPP1R1B, OLFM1, SVOP (SV2 related protein), PACSIN1, PKP4, MAGEE1, SH2D5, LGI3, ATP6V1G2, KIF1A, SLC6A17, DDN (dendrin), LAMP5, SLC30A3, NEFM (neurofilament medium), SEPTIN3, ARHGAP44, KIAA1107, RGS7BP, RGS7, KCNT1, KCNK12, PEX5L, ANO3, SCN3B and ANO4 are down regulated in GBM and has potential as a novel diagnostic and prognostic biomarker, and therapeutic target.

The up regulated hub genes obtained from PPI network. Hub genes such as VCAM1 [214], HNRNPA1 [215], CEP55 [216], A2M [217] and ETS1 [218] were responsible for advancement of GBM. HSPD1 was associated with proliferation of breast cancer cells [219], but this gene was identified first time in GBM and may be linked with proliferation of GBM cells. HNRNPK (heterogeneous nuclear ribonucleoprotein K) was liable for invasion of nasopharyngeal cancer cells [220], but this gene was identified first time in GBM and may be involved in invasion of GBM cells. High expression of CDCA5 was identified with development of esophageal cancer [221], but elevated expression of this gene was identified first time in GBM and may be linked with advancement of GBM. Our study found that PTGES3 is up regulated in GBM and has potential as a novel diagnostic and prognostic biomarker, and therapeutic target. The down regulated hub genes obtained from PPI network. Methylation inactivation of EFHD1 was liable for development of colorectal cancer [222], but loss of this gene was identified first time in GBM and may be responsible for progression of GBM. Our study found that FAM153B, RAPGEF5 and ZNF536 are down regulated in GBM and has potential as a novel diagnostic and prognostic biomarker, and therapeutic target.

Significant modules were extracted from PPI network to obtain up regulated hub genes. Hub genes such as CDKN3 [223], CCNA2 [224] and CKS2 [225] were responsible for proliferation of different cancer cells types, but these genes were identified first time in GBM and may be associated with proliferation of GBM cells. Over expression of GMNN (geminin DNA replication inhibitor) was linked with progression of liver cancer [226], but high expression of this gene was identified first time in GBM and may be liable for advancement of GBM. KPNA2 was involved in progression of GBM [227]. Our study found that CNBP (CCHC-type zinc finger nucleic acid binding protein) and NSMAF (neutral sphingomyelinase activation associated factorÂ) are up regulated in GBM and has potential as a novel diagnostic and prognostic biomarker, and therapeutic target.. Similarly, significant modules were extracted from PPI network to obtain down regulated hub genes. ARHGEF7 was linked with invasion of colorectal cancer cells [228], but this gene was identified first time in GBM and may be liable for invasion of GBM cells.

Target gene - miRNA regulatory network was constructed for up and down regulated genes. Target genes such as WEE1 [229] and G3BP1 [230] were responsible for development of GBM. RAB11FIP4 was linked with invasion of colon cancer cells [231], but this gene was identified first time in GBM and may be liable for invasion of GBM cells.

Target gene - TF regulatory network was constructed for up and down regulated genes. Target genes such as ABCC3 [232] and ABCA2 [233] were involved in progression of GBM. High expression of TNFRSF12A was liable for advancement of breast cancer [234], but elevated expression of this gene was identified first time in GBM and may be involved in development of GBM. C15orf48 (NMES1) was associated with development of esophageal cancer [235], but this gene was identified first time in GBM and may be identified with growth of GBM. Up and down regulated genes such as VKORC1, MOBP (myelin associated oligodendrocyte basic protein), PLEKHG3, TTLL7 and CAPN3 were associated in target gene - TF regulatory network and were identified as novel biomarker for pathogenesis of GBM.

High expression of hub genes (TUBA1C, CAV1, S100A4, DNAJA4, PAK6, NELL1 and ITPKA) were significantly associated with poor overall survival (OS) in GBM, while low expression of hub genes (RPL23, YY1 and ARHGEF7) were significantly associated with poor over OS in GBM and were visualized using UALCAN. Genes such as S100A4 [236] and YY1 [237] were responsible for progression of GBM. PAK6 was linked with proliferation of lung cancer cells [238], but this gene was identified first time in GBM and may be involved in proliferation of GBM cells. Polymorphic gene NELL1 was liable for progression of oral cancer [239], but this polymorphic gene was identified first time in GBM and may be linked with advancement of GBM. Next, the expression analysis of these hub genes in GBM compared with the normal and was verified on the UALCAN website. It was found that TUBA1C, CAV1, RPL23, YY1 and S100A4 were highly expressed in patients with GBM compared with normal people, while ARHGEF7, DNAJA4, PAK6, NELL1 and ITPKA were low expressed in patients with GBM compared with normal people. Next, the expression analysis of these hub genes in different age groups of GBM patients and was verified on the UALCAN website. All hub genes were showed altered expressed in all age groups of GBM patients and was verified on the UALCAN website. The mutation analysis found that mutations or alterations in all hub genes and was verified on the cBioportal website. All hub genes were validated by ICH analysis and was verified on the human protein atlas. Finally, all hub genes were validated by ROC analysis using pROC package in R software, RT-PCR and immune infiltration analysis.

In conclusion, we successfully diagnosed hub genes (TUBA1C, CAV1, RPL23, YY1, S100A4, ARHGEF7, DNAJA4, PAK6, NELL1 and ITPKA) based on bioinformatic analysis and experimental validation. This study shows that TUBA1C, CAV1, RPL23, YY1, S100A4, ARHGEF7, DNAJA4, PAK6, NELL1 and ITPKA plays a major role in the progression of GBM and has broad application potential.

## Data Availability

The datasets supporting the conclusions of this article are available in the GEO (Gene Expression Omnibus) (https://www.ncbi.nlm.nih.gov/geo/) repository. [(GSE116520) (https://www.ncbi.nlm.nih.gov/geo/query/acc.cgi?acc=GSE116520)]

## Acknowledgement

I thank Ruchi Jain, Indian Institute of Science, Molecular Reproduction Development and Genetics, C V Raman Road, Bangalore, Karnataka, India, very much, the author who deposited their microarray dataset, GSE116520, into the public GEO database.

## Ethical approval

This article does not contain any studies with human participants or animals performed by any of the authors.

## Informed consent

No informed consent because this study does not contain human or animals participants.

## Consent for publication

Not applicable.

## Author Contributions

Basavaraj Vastrad : Writing original draft, and review and editing

Chanabasayya Vastrad : Investigation and resources

Iranna Kotturshetti : Supervision and resources

